# The role of language in social-emotional, educational, and vocational outcomes in autism and in individuals who have lost the diagnosis

**DOI:** 10.1101/2025.09.10.25335495

**Authors:** Caroline Larson, Elise Taverna, Anusha Mohan, Teresa Girolamo, Deborah Fein, Inge-Marie Eigsti

## Abstract

**Background:** There is striking heterogeneity in long-term outcomes associated with an autism diagnosis, and the role of language in outcomes has not been sufficiently characterized. This study characterized the roles of structural language ability and early language milestones in long-term social-emotional, educational, and vocational outcomes in individuals with autism and individuals who have lost the autism diagnosis (LAD) relative to neurotypical (NT) peers, over and above the potential confounding role of social skills.

**Methods:** Participants were individuals with autism (*n* = 39) or LAD (*n* = 32) and NT peers (*n* = 38) age 12-39 years. Participants completed standardized and survey-based measures of social-emotional functioning and educational and vocational attainment. Language measures were an experimental structural language task (grammaticality judgement) and caregiver-report of early language milestones. Linear and generalized linear models tested how groups differed in the association between language and outcomes.

**Results:** Language was associated with certain outcomes for all groups, though there were group differences in the nature of these associations. In autism relative to LAD and NT peers, structural language was differentially associated with anxiety/depression, and language milestones were differentially associated with social relationships, quality of life, educational attainment, and full-time employment status.

**Conclusions:** Findings suggest unique pathways of influence between language and outcomes in individuals with autism versus LAD and NT peers. This evidence suggests that current language and early language development must be considered in social-emotional functioning and in educational and vocational supports from childhood *through* adulthood for individuals diagnosed with autism in childhood.

---

Autism is associated with poorer long-term social-emotional, educational, and vocational outcomes than neurotypical peers (NT), yet there is striking heterogeneity in these outcomes (Pickles et al., 2020; Steinhausen et al., 2016). One possible outcome is the loss of autistic features by adolescence or adulthood, referred to “Loss of Autism Diagnosis” (LAD). Studies of LAD report that these individuals meet gold-standard diagnostic criteria for autism early in development (e.g., prior to age 5 years), but very clearly do not meet criteria later in development (Fein et al., 2013). In fact, they display cognitive, adaptive, psychiatric, and social functioning comparable to that of NT peers in adolescence and early adulthood (Orinstein, Suh et al., 2015; Orinstein, Tyson et al., 2015). LAD status is associated with more positive social-emotional, educational, and vocational outcomes than peers who continue to meet criteria for ASD across development (Anderson et al., 2014; Moulton et al., 2016; but see Bottema-Beutel et al., 2021 and Eigsti et al., 2023 for discussion of characterizing outcomes in ASD). However, the pathways by which individuals with LAD achieve these outcomes is not well understood.

Long-term outcomes for individuals who receive an autism diagnosis in early childhood reflect myriad influences: age at diagnosis, cognitive skills (e.g., verbal and nonverbal IQ), developmental change in restricted and repetitive behaviors, having early intensive behavioral intervention, and the degree to which social difficulties change over time (Anderson et al., 2014; Le Grand et al., 2021; Pickles et al., 2020; Steinhausen et al., 2016; Usta et al., 2019; Zachor & Ben-Itzchak, 2020). One important area of heterogeneity in long-term outcomes that has received somewhat less consideration is the role of language. While many autistic individuals develop fluent speech and language, with scores on standardized assessments in the average range or higher (Eigsti et al., 2011), there is a subgroup with early language delay and structural language deficits similar to those observed in developmental language disorder, a condition characterized by impairments in structural language (e.g., verb-tense marking, grammaticality judgment; Girolamo et al., 2024; Kjelgaard & Tager-Flusberg, 2001; Larson et al., 2020a; Loucas et al., 2008; Riches et al., 2010; Wittke et al., 2017; but see Williams et al., 2008). Furthermore, our group and others have documented meaningful structural language impairments in autistic individuals who are verbally fluent, with cognitive abilities in the average range (Eigsti & Bennetto, 2009; Eigsti et al., 2007; Zane & Grossman, 2024; Zane et al., 2021), leading to an argument that standardized assessments of language may underestimate language difficulties in autism (Eigsti & Schuh, 2016).

There may be unique long-term consequences of language difficulty that are not well understood in ASD or LAD, such as underachievement in the academic domain and social-emotional problems, that are associated with structural language impairment (Conti-Ramsden et al., 2019; Conti-Ramsden et al., 2013; Durkin, et al. 2012; Durkin et al., 2017; Larson, 2025; Toseeb et al., 2017). There is also evidence suggesting that LAD individuals catch up to NT peers in some domains of language, such as pragmatic language (Crutcher et al., 2023; Eigsti et al., 2025; Irvine et al., 2016). This study addressed gaps in the literature by characterizing the longitudinal impact of early language milestones and the concurrent role of structural language ability (e.g., morphosyntax; Lugnegard et al., 2011) in long-term outcomes in ASD and LAD relative to NT peers. Known associations between language and social deficits, the latter of which is a core feature of autism, represent a potential confound. To address this possibility, we characterized the role of language in long-term social-emotional, educational, and vocational outcomes in ASD and LAD relative to NT peers *over and above* the role of social skills.

## Language and Outcomes in ASD and LAD

### Early Language Development

Language skills are an important longitudinal predictor of academic, vocational, and social-emotional outcomes (Feeny et al., 2011). For instance, several studies document remarkable difficulties in these areas for individuals with language impairments (Arkikila et al., 2008; Clegg et al., 2004) and predictive associations between individual differences in language development and later outcomes (Eadie et al., 2018; Snowling et al., 2015). Heterogeneity in early language ability likely has a substantive impact on long-term outcomes in ASD, as well, such as on long-term social functioning and LAD status (Le Grand et al., 2021; Mayo et al., 2013; Sallows & Graupner, 2005; Usta et al., 2019; Zachor & Ben-Itzchak, 2020). Delayed language milestones (e.g., late onset of first words and phrases) represents a consistent early indicator of ASD, and how language develops may be one of the most important predictors of ASD developmental trajectories in childhood (Mayo et al., 2013; Mukkades et al., 2014). Indeed, having relatively better early receptive and expressive language skills is characteristic of young children with ASD who have relatively higher adaptive functioning, fewer ASD features, and higher cognitive ability (Kim et al., 2016). Longitudinal work with a non-ASD cohort (Le et al., 2021) also found that relatively higher core language abilities (Clinical Evaluation of Language Fundamentals [CELF]) at age 4 years was associated with better parent-reported health-related quality of life at age 13 years. In fact, more than half of participants with relatively low language ability had *increasingly poor* longitudinal trajectories of health-related quality of life over time. In children with ASD, Charman et al. (2005) showed significant longitudinal correlations between composite language scores at age 3 years and reciprocal social interaction and nonverbal communication, but not restricted and repetitive behavior, at age 7 years (see also Gibson et al., 2013 for evidence of concurrent correlations between receptive language and social interaction difficulties in children with ASD).

Although these studies reveal the importance of early language abilities in social skill-related outcomes in ASD, they also underscore a potential confound: meaningful *overlap* between language and social skills. Few prior studies of autism have examined relationships between early language milestones and long-term social-emotional, vocational, and educational outcomes *separate from shared variance* between language and social skills. Moreover, inclusion of the LAD group offers a unique opportunity to test these relationships in a group of individuals who present with early language delay, similar to autistic peers, but who no longer present with ASD features later in development, unlike autistic peers. This group can reveal the ways in which early developmental patterns have greater versus lesser influence on later outcomes.

### Structural Language Ability

Though somewhat less well studied than early language milestones, there is preliminary evidence of concurrent associations between structural language (e.g., morphosyntax) and outcomes (Conti-Ramsden et al., 2013; Durkin et al., 2012; Haukedal et al., 2023; Larson, 2025; Le et al., 2021). Durkin et al. (2012) examined long-term outcomes in an adolescent group with a primary childhood diagnosis of structural language impairment who met criteria for ASD on at least one assessment (e.g., Autism Diagnostic Observation Schedule ADOS]) at study entry (*n* = 26). The comparison group included adolescents with a primary childhood diagnosis of structural language impairment who did not meet criteria for ASD (i.e., developmental language disorder, *n* = 26; Durkin et al., 2012). This study demonstrated a link across participant groups between ASD features and social-emotional (e.g., friendships, anxiety, depression) and vocational (e.g., jobs with varied literacy requirements) outcomes two years later, and a link between receptive-expressive language scores and educational outcomes two years later (see also Conti-Ramsden & Durkin, 2012). There were no differences between the structural language impairment-only versus structural language impairment-plus-ASD groups in educational outcomes or emotional health. However, the structural language impairment-plus-ASD group had poorer friendships and independent daily living scores. In another ASD cohort (Lord et al., 2020), verbal IQ accounted for concurrent ASD versus NT group differences in employment status, and ASD symptomology was associated with daily living skills and friendships. This study also showed that ASD status was associated with poorer well-being and fewer positive emotions than NT peers, highlighting the possibility of discrete pathways of influence for ASD features (e.g., social skill difficulty) versus composite language skills in long-term outcomes.

Other evidence points to causal associations between more specific measures of structural language and well-being. Kamio et al. (2012) reported an association between morphosyntax level at 6 years of age and psychological health in adulthood (e.g., self-esteem, positive feelings, and memory/concentration), but not in a briefer scale of social relationships (personal relationships, social support). In a qualitative parent-report analysis, Hantman et al. (2022) found that many factors were important to the transition to adulthood in ASD, but that linguistic and cognitive abilities were key factors in degree of independence (e.g., self-advocacy). Eadie et al. (2018) demonstrated lower parent-reported quality of life in a structural language impairment group relative to a NT group, *decreasing* quality of life from 4 to 9 years of age in the language impairment group, and an association between language skills at 7 years of age and quality of life at 9 years across groups. Indeed, a growing body of work provides additional evidence that structural language impairment is associated with poor long-term social-emotional functioning (Conti-Ramsden & Durkin, 2012; Whitehouse et al., 2009), such as an increased prevalence of anxiety relative to NT peers (Gillott et al., 2001; Howlin et al., 2000; Mawhood et al., 2000; Whitehouse et al., 2009).

Recent evidence also points to a dissociation between language and ASD features across development in at least some individuals diagnosed with autism in childhood. Larson et al. (2022) described a subgroup of individuals with LAD who had structural language difficulty, even in the absence of current ASD features. This study demonstrated that nearly an identical proportion of individuals with ASD (22.9%) and LAD (22.6%) met clinical criteria for structural language impairment. This study also reported functional brain patterns unique to this structural language impairment status over and above the influence of social skills. Taken together, examining the role of structural language in outcomes for individuals with a current *and* past diagnosis of ASD may reveal distinct pathways of influence on long-term outcomes for language versus ASD features. This line of questioning will contribute to a better understanding of heterogeneity in long-term social-emotional, educational, and vocational outcomes in individuals diagnosed with ASD in childhood.

## The Current Study

Given substantive evidence of heterogeneity in language and long-term social-emotional, vocational, and educational outcomes in autism, as well as preliminary evidence that individual differences in early language development (e.g., milestones, like age of first words) and concurrent structural language skills (e.g., morphosyntax) are important factors in long-term outcomes, the current study examined the *distinct* role of language in outcomes, separate from the overlapping influence of social skills, and examined these relationships in ASD *versus* LAD and NT groups. The LAD group allows us to probe potentially divergent developmental pathways for language versus autism features (Durkin et al., 2012; Loucas et al., 2008). This pre-registered (https://osf.io/tzvpq) study asked: To what extent do early language milestones and concurrent structural language predict long-term social-emotional, educational, and vocational outcomes, *over and above* social skills in autistic, LAD, and NT individuals, and how do these associations differ among groups?

We predicted that relatively earlier (on-time) language milestones and better structural language would be associated with relatively better social-emotional, educational, and vocational outcomes for each group, with no *a priori* hypotheses that this relationship would differ among groups (Durkin et al., 2012; Gibson et al., 2013; Lord et al., 2020; Simonoff et al. 2020). We also predicted that better social skills would attenuate this relationship across groups, but to a greater degree in autism and LAD, because of their current or history of social skills difficulty, and to a greater degree for social-emotional and vocational outcomes than educational outcomes (Durkin et al., 2012).

## Methods

This study was approved by the Institutional Review Board at the University of Connecticut and represents one study from a larger project (ASD Long-term Outcomes Study; ALTOS). Participants were individuals with ASD (*n* = 39) or LAD (*n* = 32) and NT peers (*n* = 38) age 12-39 years (groups did not differ on age; Table 1) and their caregivers. All participants had no history of intellectual disability, major psychiatric or neurological diagnoses, or uncorrected hearing or vision impairments. Additional NT criteria were: not meeting criteria for ASD per the ADOS; age-appropriate functional skills per parent/caregiver report; and no first-degree family member with an ASD diagnosis. Additional LAD eligibility criteria were: not currently meeting criteria for ASD per the ADOS; inclusion in regular education classroom; a documented diagnosis of ASD prior to five years of age; no words by 18 months or no phrases by 24 months of age (i.e., early language delay; see Fein et al., 2013 for additional information on LAD group eligibility). Current ASD diagnosis was confirmed with the ADOS-2 (Lord et al., 2000) and the Autism Diagnostic Interview-Revised (ADI-R; Lord et al., 1994) per the Diagnostic and Statistical Manual, 5^th^ Edition (DSM-5; American Psychiatric Association, 2013). There were two cohorts: (1) adults ages 18 years and older, and (2) youth ages 12-18 years. Due to the COVID pandemic, data were collected remotely using Qualtrics surveys and videoconferencing, as well as in person (with the youth cohort primarily in-person). Nonverbal ability was measured using the Penn Computerized Neurocognitive Battery Matrix Reasoning efficiency scores (Moore et al., 2014); groups did not differ significantly on this measure (we also report Penn verbal analogies efficiency scores; Table 1). See Table 1 for participant characteristics and Supplemental Materials 1, Table 1 for socioeconomic status characteristics.

**Table 1.** Participant characteristics and descriptive results.

| Participant Characteristics | NT<br>(*n* = 38) | LAD<br>(*n* = 32) | ASD<br>(*n* = 39) | *p*-value |
| --- | --- | --- | --- | --- |
| Age | 21.78<br>(5.99) | 22.86<br>(3.86) | 21.13<br>(5.60) | NT-ASD *p* = .63<br>NT-LAD *p* = .37<br>ASD-LAD *p* = .13 |
| Youth:Adult | 12:26 | 5:27 | 18:21 |  |
| Gender identity<br>(F:M:Non-binary/Other) | 19:17:1 | 7:24:1 | 9:27:3 | NT-ASD *p* < .05*<br>NT-LAD *p* < .05*<br>ASD-LAD *p* = .73 |
| Race/Ethnicity |  |  |  |  |
| *Asian or Pacific Islander* | 1 | 2 | 3 |  |
| *American Indian/Native* | 1 | 0 | 1 |  |
| *Hispanic* | 4 | 2 | 3 |  |
| *Multiracial* | 2 | 1 | 1 |  |
| *White* | 34 | 29 | 34 |  |
| ADOS-2 Communication | 0.59<br>(0.96) | 0.78<br>(0.83) | 3.32<br>(1.78) | NT-ASD *p* < .001***<br>NT-LAD *p* = .39<br>ASD-LAD *p* < .001*** |
| ADOS-2 Social<br>Interaction | 0.68<br>(0.97) | 1.19<br>(1.23) | 6.30<br>(2.80) | NT-ASD *p* < .001***<br>NT-LAD *p* = .06<br>ASD-LAD *p* < .001*** |
| ADOS-2 Stereotyped<br>Behaviors and Restricted<br>Interest | 0.11<br>(0.31) | 0.25<br>(0.57) | 1.05<br>(1.03) | NT-ASD *p* < .001***<br>NT-LAD *p* = .22<br>ASD-LAD *p* < .001*** |
| VABS-3 Socialization | 105.5<br>(9.50) | 102.7<br>(11.43) | 83.71<br>(18.41) | NT-ASD *p* < .001***<br>NT-LAD *p* = .35<br>ASD-LAD *p* < .001*** |
| Penn Nonverbal<br>Efficiency | 8.56<br>(1.38) | 8.46<br>(1.63) | 8.06<br>(1.80) | NT-ASD *p* = .22<br>NT-LAD *p* = .78<br>ASD-LAD *p* = .39 |
| Penn Verbal<br>Efficiency | 7.40<br>(1.97) | 7.50<br>(1.81) | 6.65<br>(2.54) | NT-ASD *p* = .20<br>NT-LAD *p* = .83<br>ASD-LAD *p* = .15 |

*Note.* ADOS = Autism Diagnostic Observation Schedule, scaled scores; VABS-3 = Vineland Adaptive Behavior Scales, Third Edition; Penn Nonverbal = Penn Matrix Reasoning, efficiency scores; Penn Verbal = Penn Verbal Analogies, efficiency scores; \**p* < .05; \*\**p* < .01; \*\*\**p* < .001

### Language Measures

#### Language Milestones

We collected caregiver-reported age of first words and age of first phrases in months using Qualtrics surveys and the Autism Diagnostic Interview-Revised.

#### Grammaticality Judgement

This experimental task was administered via Qualtrics survey. There were 23 grammatical and 23 ungrammatical sentences, fully randomized. Ungrammatical sentences contained errors of morphosyntax per General American English, including word order, omission, substitutions, and tense marking (see Supplemental Materials 1, p. 1-2 for stimuli). Stimuli were presented auditorily from recordings of a native English speaker. Participants made a two alternative forced choice judgement of correct (grammatical) or incorrect (ungrammatical). Participants completed two training trials with feedback to verify task comprehension before doing the task. We examined performance according to signal detection theory, using A’ (A prime) to measure participants’ ability to detect grammatical sentences separate from response bias. We also report descriptive percent accuracy for interpretability. See Table 2 and Supplemental Materials 1, Table 2 for descriptive performance on language measures.

**Table 2.** Descriptive data for language and outcomes measures.

| Measure | NT<br>(*n* = 38) | LAD<br>(*n* = 32) | ASD<br>(*n* = 39) |  |
| --- | --- | --- | --- | --- |
| Age of first words | 12.94<br>(4.34) | 21.92<br>(10.55) | 20.81<br>(11.01) | NT-ASD $p < .01^{**}$<br>NT-LAD $p < .001^{***}$<br>ASD-LAD $p = .71$ |
| Age of first phrases | 19.30<br>(6.82) | 29.12<br>(15.18) | 34.12<br>(12.41) | NT-ASD $p < .001^{***}$<br>NT-LAD $p < .01^{**}$<br>ASD-LAD $p = .21$ |
| <sup>†</sup>Grammaticality Judgement A` | 0.88<br>(0.07) | 0.89<br>(0.07) | 0.82<br>(0.12) | NT-ASD $p < .05^{*}$<br>NT-LAD $p = .64$<br>ASD-LAD $p < .01^{**}$ |
| Quality of Life | 47.62<br>(7.51)<br>*n* = 24 | 48.44<br>(8.76)<br>25 | 42.06<br>(10.02)<br>16 | NT-ASD $p = .07$<br>NT-LAD $p = .73$<br>ASD-LAD $p < .05^{*}$ |
| Life Satisfaction | 24.71<br>(5.38)<br>*n* = 24 | 24.84<br>(5.42)<br>25 | 20.56<br>(6.69)<br>16 | NT-ASD $p < .05^{*}$<br>NT-LAD $p = .93$<br>ASD-LAD $p < .05^{*}$ |
| Social Support | 50.00<br>(8.42)<br>*n* = 37 | 51.17<br>(6.09)<br>29 | 47.30<br>(8.23)<br>33 | NT-ASD $p = .18$<br>NT-LAD $p = .51$<br>ASD-LAD $p < .05^{*}$ |
| ASEBA Total Problems | 44.96<br>(9.42)<br>*n* = 27 | 44.30<br>(11.04)<br>27 | 58.00<br>(12.08)<br>29 | NT-ASD $p < .001^{***}$<br>NT-LAD $p = .81$<br>ASD-LAD $p < .001^{***}$ |
| ASEBA Anxiety/Depression | 53.57<br>(5.88) | 53.85<br>(5.48) | 64.07<br>(11.60) | NT-ASD $p < .001^{***}$<br>NT-LAD $p = .86$ |
|  | *n* = 28 | 27 | 28 | ASD-LAD *p* < .001*** |
| ASEBA Friends/CBCL Social | 52.21<br>(8.78) | 53.42<br>(6.82) | 37.24<br>(11.27) | NT-ASD *p* < .001***<br>NT-LAD *p* = .57 |
|  | *n* = 28 | 26 | 29 | ASD-LAD *p* < .001*** |
| Full-time vs. not full-time school or work status | 26:6 | 26:2 | 23:8 | NT-ASD *p* = .71<br>NT-LAD *p* = .35<br>ASD-LAD *p* = .12 |
| Education Completed in years | 12.11 | 12.21 | 10.91 | NT-ASD *p* = .13 |
| > 22 years old | (2.00) | (0.70) | (2.02) | NT-LAD *p* = .84 |
|  | *n* = 18 | 14 | 11 | ASD-LAD *p* = .06 |
| Hours Working per week at job | 38.00 | 40.00 | 33.67 | NT-ASD *p* = .22 |
| > 18 years old and not in college | (3.10) | (0.00) | (7.21) | NT-LAD *p* = .17 |
|  | *n* = 6 | 10 | 6 | ASD-LAD *p* = .09 |

*Note.* Sample sizes reported for cohort-specific measures and for measures with missing data, see Supplementary Materials 1 for more information on sample sizes for outcome measures; A` = a prime; Age of first words/phrases = in months, from caregiver report; ASEBA = Achenbach System of Empirically Based Assessment, Child Behavior Checklist, *t*-scores ( $\leq 18$ years) and Adult Behavior Checklist, *t*-scores ( $> 18$ years); <sup>†</sup>See Supplementary Materials 1, Table 2 for GJ percent accuracy; \**p* < .05; \*\**p* < .01; \*\*\**p* < .001

#### Standardized Assessment

We administered the CELF-5 (Wiig et al., 2013) to our in-person youth cohort, and we report Core Language standard scores and Recalling Sentences scaled scores (i.e., a clinical marker of structural language impairment) for participant characteristics information.

### Social Skills Standardized Assessment

We administered the Vineland Adaptive Behavior Scales, Third Edition (VABS-3; Sparrow et al., 2016) to participant caregivers. VABS-3 measures adaptive behavior, and we used Socialization scale T-scores as our measure of concurrent social skills. This scale includes measures of functioning in social situations, including interpersonal relationships and coping skills.

### Social-Emotional Outcomes

#### Surveys – Adult Participants

Quality of Life was measured using the ASD-Quality of Life Scale (McConachie et al., 2007) involving 9 questions using a 5-point Likert scale. Higher ratings suggest better Quality of life (items 6-8 are reverse scored). We used the Satisfaction with Life Scale (Deiner et al., 1985) to ask about broad dimensions of happiness; this scale encompasses seven questions about life satisfaction using a 7-point Likert scale. Higher ratings suggest more life satisfaction. We administered the Multidimensional Scale of Perceived Social Support (Zimet et al., 1988) involving 12 questions using a 5-point Likert scale, yielding an average score. Higher scores indicate higher levels of perceived social support. Each of these measures was self-report.

#### Standardized Assessments

The Achenbach System of Empirically Based Assessment (ASEBA) self-report Adult Behavior Checklist (ABCL; ages 19 and older) and the caregiver-report Child Behavioral Checklist (CBCL; ages 12-18 years) from the Achenbach System of Empirically Based Assessment (Achenbach & Rescorla, 2004) provided measures of Social Relationships (e.g., friendships and close relationships), Anxiety/Depression, and Total Problems, which we collapsed across adolescents and young adults. These scales yield T-scores for self-or caregiver-report. See Table 2 and Supplemental Materials 1, Table 2 for descriptive performance on social-emotional measures.

### Education and Vocation Outcomes

#### Surveys

We administered 19 exploratory items, including multiple choice and text entry, with responses from all participants and caregivers of adolescent participants. We examined outcomes by age, such as “How much education have you completed so far?” and “How many hours/week do you work?” (see Supplemental Materials 1 for complete list of questions) for participants 22 years of age or older. We also examined full-time status either working or in college for participants 18 years of age or older. These questions yielded yes/no and numerical responses (e.g., hours/week spent working; salary). Additional text entry responses were used for descriptive information to clarify statistical relationships. See Supplemental Materials 1, p. 3 for questionnaire, Table 2 for descriptive data, and Supplemental Materials 1, Table 4 for additional descriptive data on vocation and education measures.

### Data Analysis

We log transformed Likert scale values for quality of life, life satisfaction, and social support survey scales, and we log transformed hours working per week and Grammaticality Judgement task A’ scores. Linear models were used to test research questions regarding continuous outcome variables and generalized linear models were used to examine binary outcome variables. Models tested the association between language and outcome measures and tested whether this association differed by group (group by language interaction terms) *separately* for each language variable: grammaticality judgement À, age of first words, and age of first phrases (i.e., three separate models). Covariates included age, due to the wide age range of the sample (12-39 years), and VABS-3 Socialization *T*-scores, to account for the possibility that concurrent social skills mediate the relationship between language and outcomes. In our pre-registration, we planned to covary VABS-3 Daily Living Skills *T*-scores; however, given the closer association between Socialization and language (i.e., a greater, more relevant potential confound), we opted to not conduct these additional analyses covarying Daily Living Skills. We conducted planned follow-up within-group analyses (ASD, LAD, NT) to characterize between– and within-group associations.

## Results

See Table 2 for descriptive data, Table 3 for a summary of significant effects related to language variables, and Supplemental Materials 2 for complete model output.

**Table 3.**
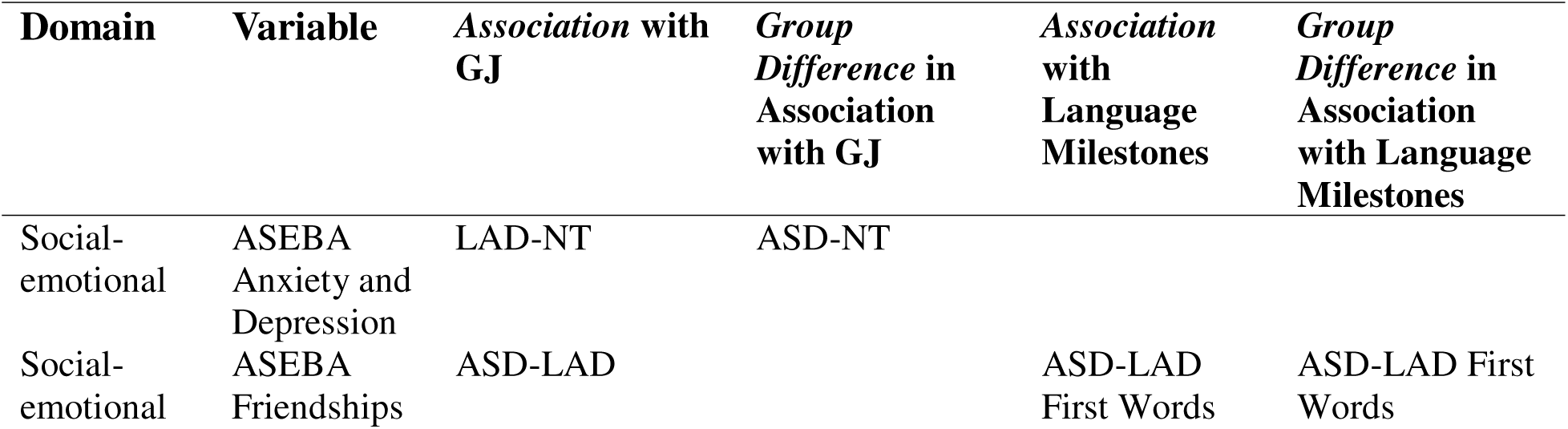
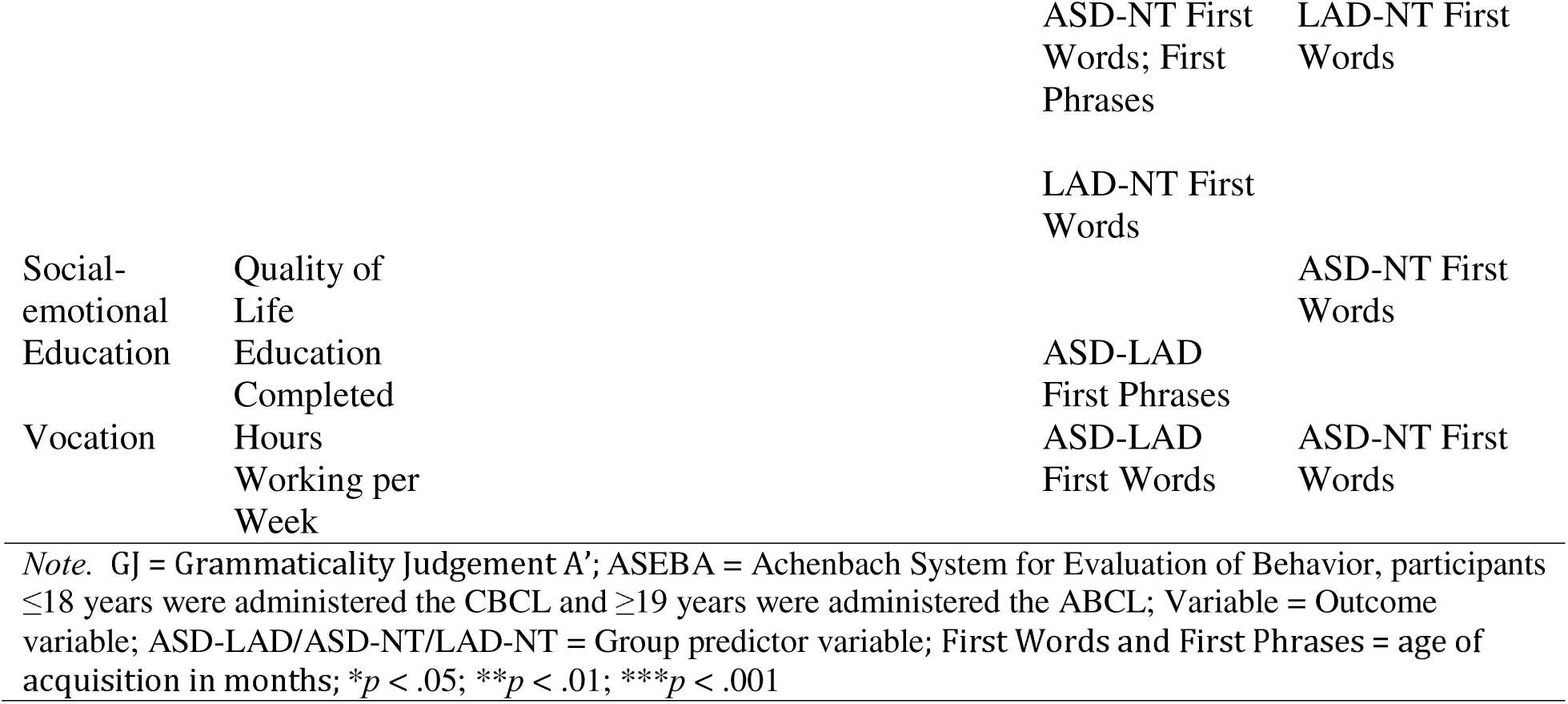
Summary of significant results for language variables.

### First-order Results: Simple Group Comparisons

Descriptively, autism and LAD groups both had significantly later caregiver-reported onset of first words and first phrases relative to the NT group, with no autism-LAD group differences. Autism, LAD, and NT group comparisons were broadly consistent for all socio-emotional outcome measures; the autism group showed more current struggles and impairments (e.g., significantly higher endorsement of anxiety and depression, significantly lower self-reported life satisfaction and quality of life). The autism group had significantly lower accuracy on grammaticality judgment, whereas the LAD and NT groups did not differ, suggesting that the autistic participants had poorer morphosyntactic processing. In contrast, there were no group differences in occupational or academic outcomes, suggesting no overall group differences in this important sociodemographic domain in a verbally fluent group of individuals with age-appropriate cognitive skills. Given the wide individual variability and the fact that adolescent participants had not yet completed their education, this result likely underestimates the effects of autism on academic and occupational success.

### Language and Long-term Outcomes: Social-emotional Outcomes

#### ASEBA Anxiety/Depression

##### ASD-LAD models

There were no significant effects (*p*’s < .05), indicating no group differences in anxiety/depression symptoms when accounting for language and social skills and no group differences in the association between language and anxiety/depression symptoms.

##### ASD-NT models

There were significant effects of socialization (*b* = –0.197; SE = 0.090; *t* = –2.180; *p* < .05) and the interaction between group and À (*b* = 62.326; SE = 29.161; *t* = 2.137; *p* < .05). Follow-up within-group analyses indicated that relatively better grammaticality judgement performance was associated with *fewer* anxiety/depression symptoms for the ASD group (*b* = –26.087) and with *more* anxiety/depression symptoms for the NT group (*b* = 33.226; Figure 1). The effect of socialization was significant in all models (*p*’s < .05), indicating that relatively better socialization scores were associated with fewer anxiety and depression symptoms across groups. There were no other significant effects (*p*’s > .06), indicating no group differences in anxiety/depression symptoms when accounting for language and social skills.

**Figure 1.**
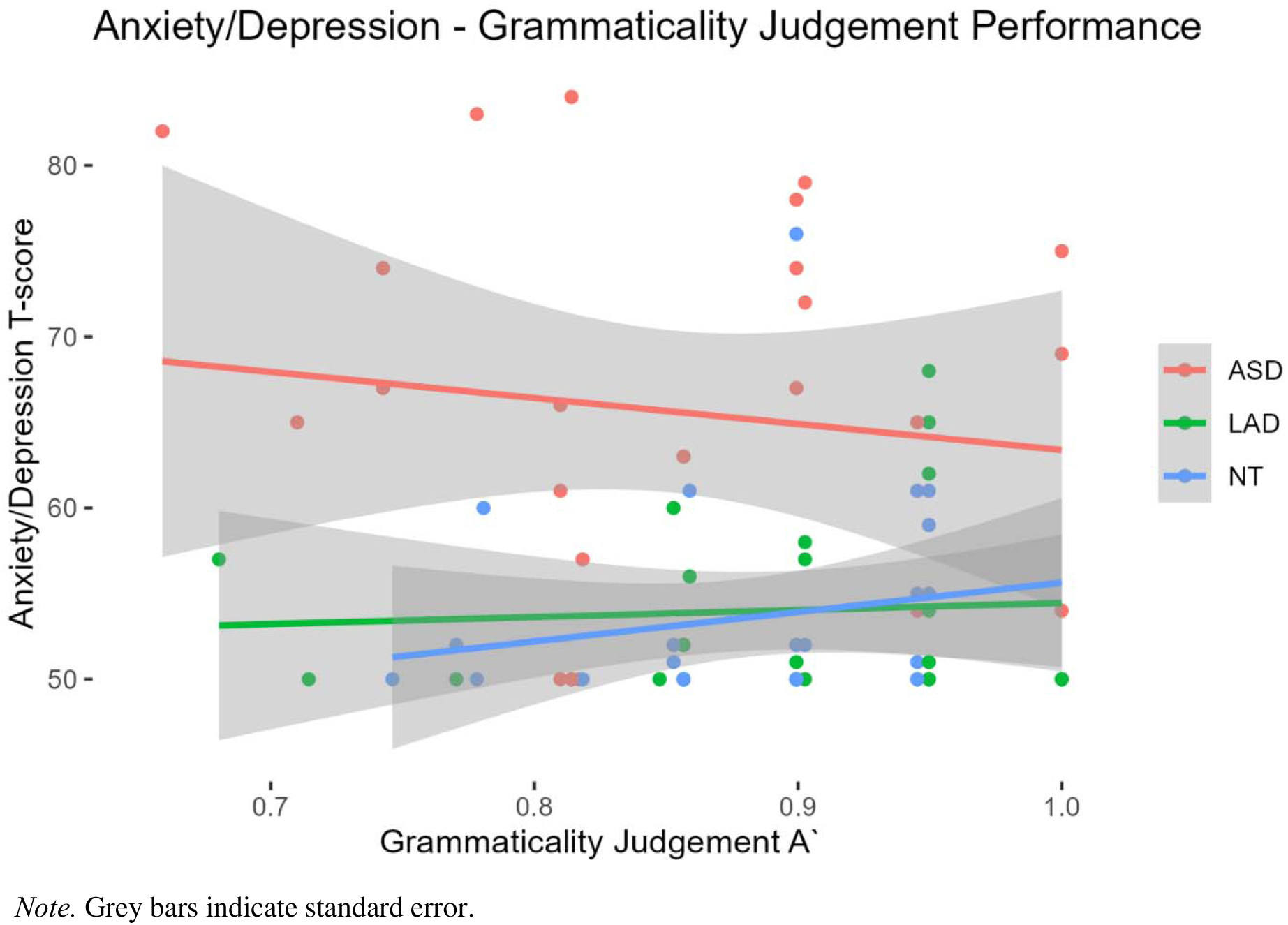
The association between anxiety/depression symptoms and grammaticality judgement À performance for each group.

##### LAD-NT models

There were significant effects of À (*b* = 26.008; SE = 10.844; *t* = 2.398; *p* < .05), indicating that relatively better grammaticality judgement performance was associated with more anxiety and depression symptoms across groups (Figure 1), and socialization (*b* = –0.191; SE = 0.076; *t* = –2.499; *p* < .05), indicating that relatively better socialization scores were associated with fewer anxiety and depression symptoms across groups. The effect of group was significant (*b* = 9.612; SE = 4.467; *t* = 2.152; *p* < .05) in the first words and first phrases models (*b* = 8.344; SE = 4.075; *t* = 2.047; *p* < .05), indicating that the NT group had relatively more anxiety and depression symptoms than the LAD group when accounting for age of first words/phrases and social skills. There were no other significant effects (*p*’s > .06).

### ASEBA Total Problems

#### ASD-LAD models

The effect of socialization was significant across models (*p*’s < .01), indicating that relatively better socialization scores were associated with fewer total problems across groups. There were no other significant effects (*p*’s > .11), indicating no group differences in total problems when accounting for language and social skills and no group differences in the association between language and total problems.

#### ASD-NT models

The effect of socialization was significant across models (*p*’s < .01), indicating that relatively better socialization scores were associated with fewer total problems across groups. The effect of age was significant in the À and age of first phrases models (*p*’s < .05), indicating that relatively younger participants had fewer total problems across groups when accounting for À or age of first phrases and social skills. There were no other significant effects (*p*’s > .17), indicating no group differences in total problems when accounting for language and social skills and no group differences in the association between language and total problems.

#### LAD-NT models

There were significant effects of socialization (*p*’s < .01), indicating that relatively better socialization scores were associated with fewer total problems across groups, and age (*p*’s < .05), indicating that relatively younger participants had fewer total problems. There were no other significant effects (*p*’s > .06), indicating no group differences in total problems when accounting for language and social skills and no group differences in the association between language and total problems.

### ASEBA Social Relationships

#### ASD-LAD models

There was a significant effect of the interaction between group and À (*b* = –68.743; SE = 31.787; *t* = –2.163; *p* < .05). Follow-up within-group analyses indicated that relatively better grammaticality judgement performance was associated with better social relationships for the ASD group (*b* = 41.884) and with worse social relationships for the LAD group (*b* = –38.457; Figure 2). There were also significant effects of age of first words (*b* = – 0.406; SE = 0.148; *t* = –2.752; *p* < .01) and the interaction between group and age of first words (*b* = 0.595; SE = 0.284; *t* = 2.099; *p* < .05). Follow-up within-group analyses indicated that relatively younger age of first words was associated with better social relationships for the ASD (*b* = –0.698) and LAD group (*b* = –0.115; Figure 3), though the effect was significantly larger for the ASD group. There were no other significant effects (*p*’s > .07), indicating no group differences in social relationships when accounting for language and social skills.

**Figure 2.**
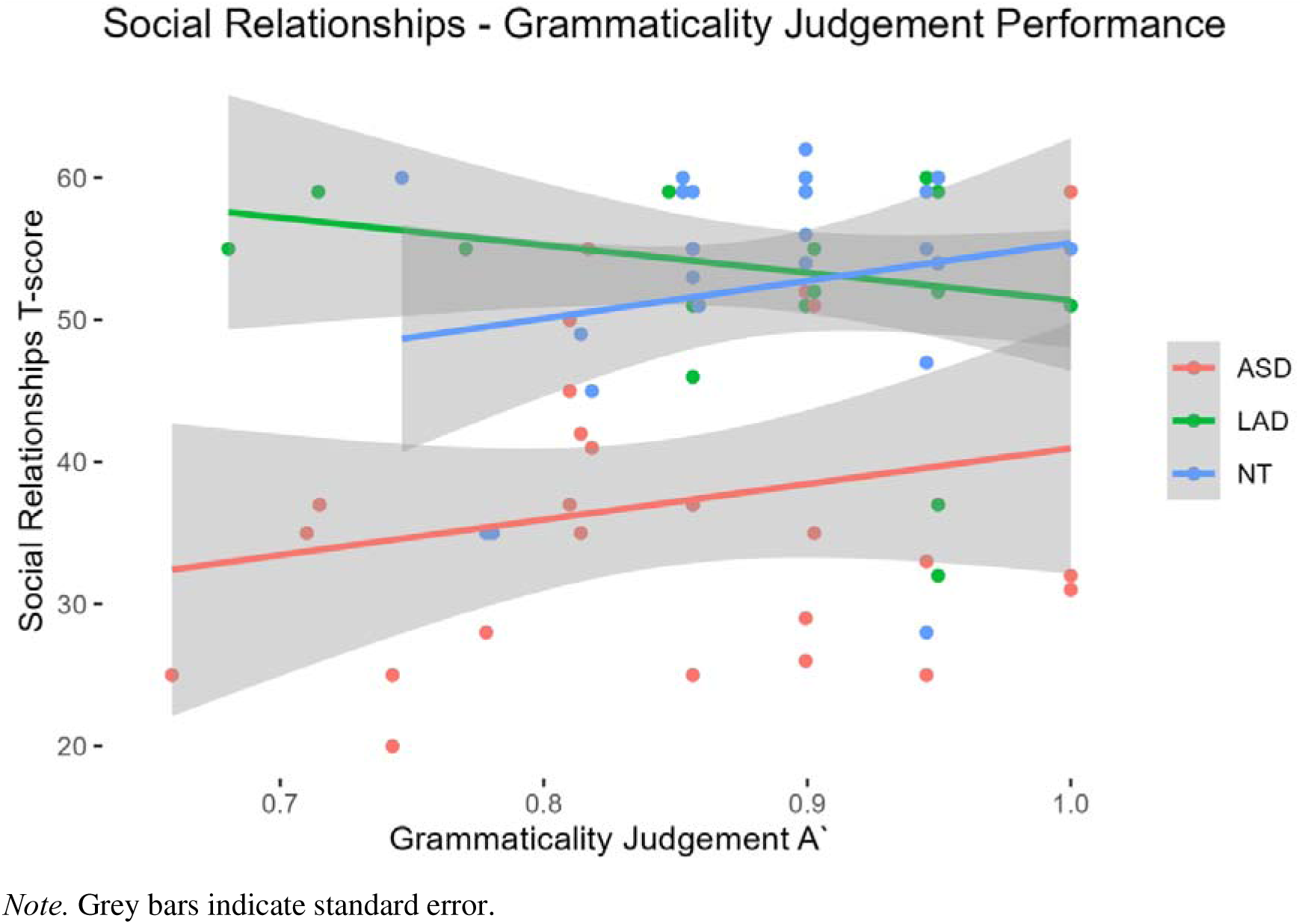
The association between friendships and grammaticality judgement À performance for each group.

**Figure 3.**
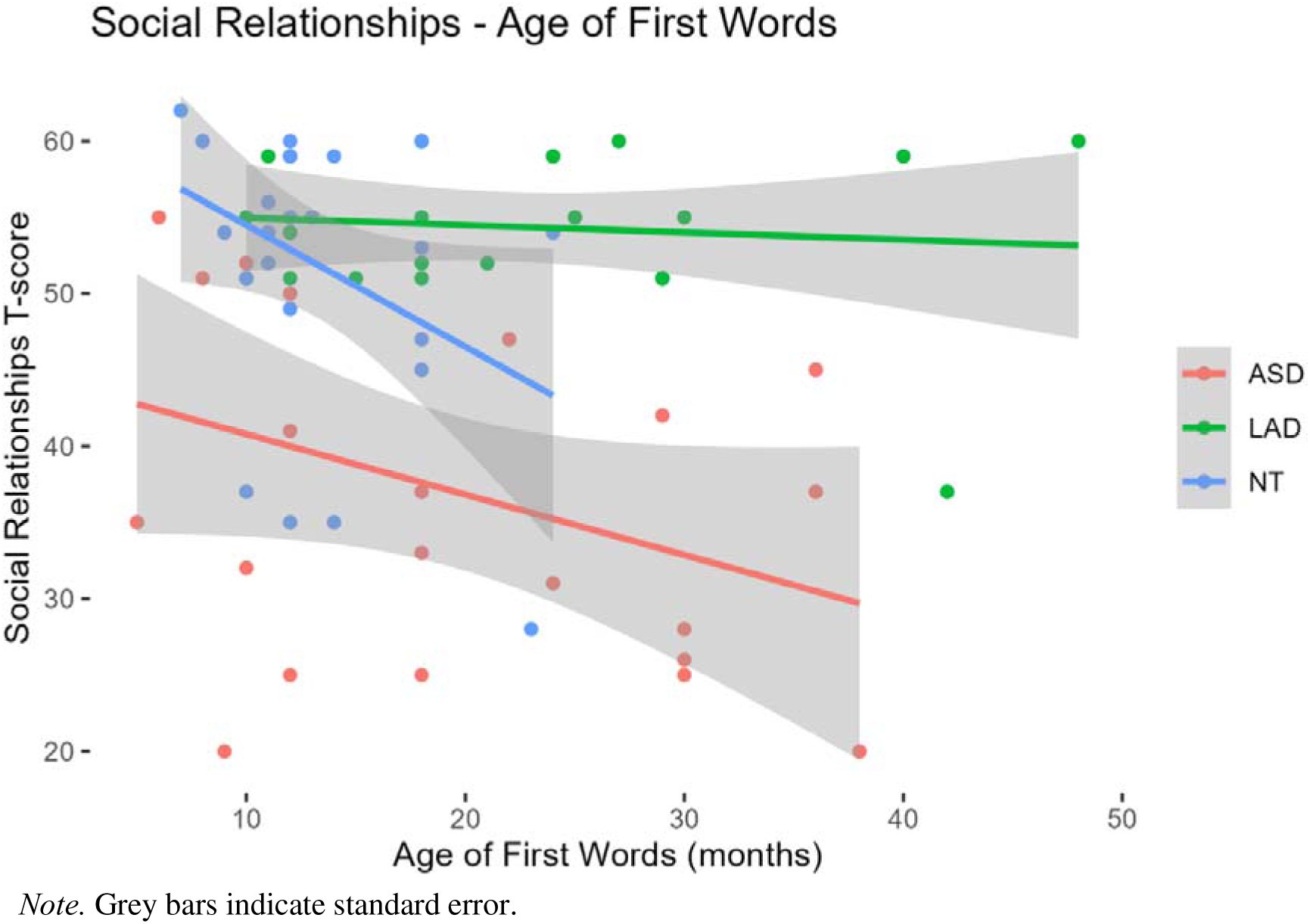
The association between social relationships and age of first words for each group.

#### ASD-NT models

There was a significant effect of age of first words (*b* = –0.772; SE = 0.266; *t* = –2.906; *p* < .01) and age of first phrases (*b* = –0.416; SE = 0.200; *t* = –2.084; *p* < .05; Figure 3), indicating that relatively younger age of first words and phrases was associated with better social relationships across groups. There were no other significant effects (*p*’s > .13), indicating no group differences in social relationships when accounting for language and social skills and no group differences in the association between language and social relationships.

#### LAD-NT models

There were significant effects of age of first words (*b* = –0.505; SE = 0.188; *t* = –2.678; *p* < .05), age (*b* = 0.427; SE = 0.194; *t* = 2.200; *p* < .05), and the interaction between group and age of first words (*b* = –0.800; SE = 0.375; *t* = –2.133; *p* < .05). Follow-up within-group analyses indicated that relatively younger age of first words was associated with better social relationships for the LAD (*b* = –0.115) and NT group (*b* = –0.922; Figure 3), though this effect was significantly larger for the NT group. There were no other significant effects (*p*’s > .09), indicating no group differences in social relationships when accounting for language and social skills.

### Quality of Life, Life Satisfaction, and Social Support Surveys

#### ASD-LAD models

There were no significant effects for Quality of Life (*p*’s > .07), Life Satisfaction (*p*’s > .06), or Social Support (*p*’s > .25), indicating no group differences in survey measures when accounting for language and social skills and no group differences in the association between language and survey measures.

#### ASD-NT models

There were significant effects of group (*b* = 0.590; SE = 0.202; *t* = 2.920; *p* < .01), socialization (*b* = 0.011; SE = 0.004; *t* = 2.749; *p* < .05), age (*b* = 0.017; SE = 0.008; *t* = 2.190; *p* < .05), and the interaction between group and age of first words (*b* = –0.033; SE = 0.014; *t* = –2.271; *p* < .05) for Quality of Life. These results indicate that the ASD group reported lower Quality of Life than NT peers, relatively better social skills were associated with better quality of life across groups, and relatively older participants reported better quality of life across groups. Follow-up within-group analyses indicated that relatively older age of first words was associated with better Quality of Life in the ASD group (*b* = 0.009) and with relatively worse Quality of Life in the NT group (*b* = –0.006; see Supplemental Materials 1, Figure 1). The effect of age was significant across Quality of Life models (*p*’s < .05). There were significant effects of group (*b* = 0.725; SE = 0.292; *t* = 2.480; *p* < .05), age (*b* = 0.024; SE = 0.011; *t* = 2.096; *p* < .05), and socialization (*b* = 0.021; SE = 0.006; *t* = 3.447; *p* < .01) in the first words model. The effect of socialization was significant across Life Satisfaction models (*p*’s < .05), and there were no other significant effects (*p*’s > .05). Beyond the interaction effects, significant results indicate that the ASD group reported lower Quality of Life than NT peers, relatively better social skills were associated with better quality of life across groups, and relatively older participants reported better quality of life across groups.

#### LAD-NT models

There were no significant effects for Quality of Life (*p*’s > .12), Life Satisfaction (*p*’s > .06), or Social Support (*p*’s > .27), indicating no group differences in survey measures when accounting for language and social skills and no group differences in the association between language and survey measures.

### Language and Long-term Outcomes: Educational and Vocational Outcomes

Given the small sample sizes for education and vocation variables (Supplemental Materials 1, Table 4), we simplified statistical models by removing the age covariate, given that the samples were subsetted by age as indicated by the outcome variable (see below) prior to running statistical models, and by analyzing within-group patterns related to language variables with no covariates (i.e., simple correlations). Results should be interpreted cautiously given the small samples and analytical approach.

### Full Time School or Work Status, Individuals ***≥***18 years

#### ASD-LAD models

There were no significant effects (*p*’s > .12), indicating no group differences in full-time status when accounting for language and social skills and no group differences in the association between language and full-time status.

#### ASD-NT models

There were no significant effects (*p*’s > .05), indicating no group differences in full-time status when accounting for language and social skills and no group differences in the association between language and full-time status.

#### LAD-NT models

There were no significant effects (*p*’s > .12), indicating no group differences in full-time status when accounting for language and social skills and no group differences in the association between language and full-time status.

#### Within-group models

There were no significant effects (*p*’s > .10), indicating no significant associations between language and full-time status.

### Education Completed, Individuals ***≥***22 years

#### ASD-LAD models

There was a significant effect of age of first phrases (*b* = 0.090; SE = 0.025; *t* = 3.619; *p* < .01), indicating that relatively older age of first phrases was associated with more education completed across groups. There were no other significant effects (*p*’s > .10), indicating no group differences in education completed when accounting for language and social skills and no group differences in the association between language and education completed.

#### ASD-NT models

There were no significant effects (*p*’s > .13), indicating no group differences in education completed when accounting for language and social skills and no group differences in the association between language and education completed.

#### LAD-NT models

There were no significant effects (*p*’s > .17), indicating no group differences in education completed when accounting for language and social skills and no group differences in the association between language and education completed.

#### Within-group models

There was a significant effect of age of first phrases (*b* = 0.060; SE = 0.016; *t* = 3.686; *p* < .01) for the LAD group, indicating that relatively older age of first phrases was associated with more education completed. There were no other significant effects of language variables (*p*’s > .05), indicating no significant associations between language and education completed for the ASD and NT groups.

### Hours Working per Week, Individuals ***≥***22 years

#### ASD-LAD models

There was a significant effect of group (*b* = 0.514; SE = 0.082; *t* = 6.286; *p* < .01) and socialization (*b* = –0.017; SE = 0.002; *t* = –8.362; *p* < .001) in the À model. There were significant effects of group (*b* = 0.894; SE = 0.016; *t* = 56.453; *p* < .001), age of first words (*b* = 0.016; SE = 0.000; *t* = 37.558; *p* < .001), socialization (*b* = 0.002; SE = 0.001; *t* = 3.783; *p* < .001), and the interaction between group and age of first words (*b* = –0.033; SE = 0.001; *t* = –37.109; *p* < .001). Follow-up within-group analyses (covarying socialization) indicated that relatively older age of first words was associated with more hours working per week for the ASD (*b* = 0.033) and LAD group (*b* = 0.000), though this effect was significantly larger for the ASD group. There was also a significant effect of socialization (*b* = –0.018; SE = 0.005; *t* = –3.729; *p* < .01) in the first phrases model. Beyond the interaction effects, significant results indicate that the LAD group had more working hours per week than the ASD group and relatively better social skills were associated with more working hours per week across groups. There were no other significant effects (*p*’s > .07).

#### ASD-NT models

There was a significant effect of group (*b* = 0.865; SE = 0.165; *t* = 5.229; *p* < .01) and the interaction between group and age of first words (*b* = –0.034; SE = 0.009; *t* = –3.724; *p* < .05). Follow-up within-group analyses (covarying socialization) indicated that relatively older age of first words was associated with more working hours per week for the ASD group (*b* = 0.033) and with fewer working hours per week for the NT group (*b* = –0.015). There were significant effects of socialization in the À and age of first phrases models (*p*’s < .01), and no other significant effects (*p*’s > .07). Beyond the interaction effects, significant results indicate that the NT group had more working hours per week than the ASD group and relatively better social skills were associated with more working hours per week.

#### LAD-NT models

There were no significant effects (*p*’s > .06), indicating no group differences in hours working per week when accounting for language and social skills and no group differences in the association between language and hours working per week.

#### Within-group models

There was a significant effect of age of first words (*b* = 0.029; SE = 0.005; *t* = 5.886; *p* < .01) for the ASD group, indicating that relatively older age of first words was associated with more hours working per week. There were no other significant effects of language variables (*p*’s > .24), indicating no significant associations between language and education completed for the LAD and NT groups.

## Discussion

This study examined the role of early language milestones and concurrent structural language (grammaticality judgement) in long-term social-emotional, educational, and vocational outcomes in adolescents and young adults with ASD or LAD relative to NT peers. We controlled for the likely confounding role of social skills based on known associations between language and social skills, as well as known difficulties in the area of social skills in autism. In simple group comparisons, the ASD group had poorer social-emotional outcomes than the LAD and NT groups, yet these differences were no longer evident when accounting for language and social skills (except for self-reported quality of life). In fact, LAD-NT group differences emerged in one social-emotional outcome when accounting for early language milestones; the LAD group had relatively fewer anxiety/depression symptoms than the NT group after controlling for early language development. There were no simple group differences in educational and vocational outcomes, though sample sizes were small for these measures and the LAD and NT groups appeared to work more hours per week than the ASD group when accounting for language and social skills.

With regard to the role of language in outcomes, we hypothesized that earlier language acquisition milestones and better current structural language would be associated with relatively better outcomes across groups, and we did not predict any *a priori* group differences in these associations. These predictions were partially borne out in the data. There were associations between language milestones and social-emotional outcomes, including social relationships and quality of life, an educational outcome (education completed), and a vocational outcome (hours working per week). There were also associations between current structural language and social-emotional outcomes, including anxiety/depression symptoms for the LAD and NT groups and social relationships for the ASD and LAD groups, but not between current structural language and educational or vocational outcomes. Some of these associations differed between groups. Relatively younger age of first words was associated with better social relationships to a greater degree for the ASD and NT groups than the LAD group. Relatively younger age of first words was associated with worse quality of life for the ASD group and with better quality of life for the NT group; this pattern could reflect non-speaking children being referred for clinical services at an earlier age. Self-reported quality of life was poorer overall in the ASD group even after controlling language and social skills, suggesting notable challenges for autistic individuals on this measure of social-emotional outcomes. Relatively younger age of first words was (counter-intuitively) associated with fewer hours working per week for the ASD group and with more hours working per week for the NT group. Sample sizes for hours working per week, however, were small, and this result should be interpreted cautiously. Relatively better concurrent structural language performance was associated with fewer anxiety/depression symptoms for the ASD group and (somewhat surprisingly) with more anxiety/depression symptoms for the LAD and NT groups. These symptoms were relatively similar in the LAD and NT groups overall and, importantly, did not appear to reflect clinical-level symptomology, suggesting that the variability accounted for by structural language was within the range of generally well-adapted psychiatric functioning.

Taken together, current language ability appears to be associated with social-emotional outcomes, but not educational and vocational outcomes, in this sample of adolescents and young adults. In contrast, early language milestones appear to play a role across social-emotional, educational, and vocational outcomes. Findings suggest unique pathways of influence between language and outcomes in individuals with autism and a history of autism. However, the effect of early language development on long-term educational (education completed) and vocational (hours working per week) outcomes observed in the current study should be interpreted as preliminary given the limited sample size contributing data to these measures and the retrospectively recalled nature of these milestones.

### Language and Social-Emotional Outcomes

#### Language Acquisition Milestones

The current study demonstrated associations between developmental language milestones and social-emotional outcomes, including social relationships and self-reported quality of life. Relatively earlier acquisition of first words (on-time language development) was associated with better social relationships across groups, though this association was stronger in the ASD and NT groups than the LAD group. Relatively earlier acquisition of first words was also associated with worse quality of life for the ASD group, but with better quality of life for the NT group. These findings suggest that the influence of early language development is stronger for autistic and NT individuals than for LAD individuals. These findings align with the developmental trajectory of autistic features in LAD individuals. In LAD, clinical-threshold autistic features are present early in development and diminish across development, and individuals with LAD generally have more positive outcomes (e.g., fewer total problems, better self-reported quality of life and life satisfaction; Supplemental Materials 1, Table 3) than individuals who present with clinical-threshold autistic features across development (e.g., Eigsti et al., 2022; Fein et al., 2013). Thus, the influence of early autistic features *and* language development milestones explain social-emotional outcomes in LAD to a lesser degree than in autistic and NT peers.

The role of developmental milestones in autistic social-emotional outcomes converges on prior evidence indicating that early language development is a key predictor of how autism progresses, even for an older age group (Charman et al., 2005; Fenson et al., 1993; Gibson et al., 2013; Howlin et al., 2000; Mayo et al., 2013; Mukkades et al., 2014). Prior literature suggests that milder presentation of autism features in early childhood, such as less severe developmental delay, is associated with better mental health and well-being outcomes in later childhood and adolescence (Anderson et al., 2014; Usta et al., 2019; Zachor & Ben-Itzchak, 2020). Adding to this literature, we have shown that early language milestones *continue to be important* for social relationships, over and above the influence of social skills, in adolescence and adulthood for individuals with autism, and these milestones share a *similar relationship* with social relationships for NT individuals. However, relatively earlier acquisition of language milestones (on-time language development) was associated with poorer self-reported quality of life for autistic individuals, but with better quality of life for NT individuals. Notably, autistic individuals reported poorer quality of life over and above the influence of language and social skills, suggesting that the association between language milestones and quality of life may not reflect *meaningful differences* in long-term quality of life for autistic adolescents and adults. This unexpected finding does, however, suggest different pathways of influence between early language development and this measure of well-being. It is possible that, similar to the association between concurrent structural language and anxiety/depression symptoms in LAD, autistic individuals who experienced less severe early language delay had less access to intensive services or were placed in more neurotypical contexts with less support. This self-report measure of quality of life may capture the effects of these factors, such as those related to feelings of belonging or happiness in one’s personal or professional life, that are not as well-captured in caregiver/teacher report measures or in measures of symptomology which are more commonly used in the current evidence base. Though speculative, this interpretation may guide future research that examines the role of early language delay on self-reported long-term outcomes for autistic individuals while considering their service and setting history. Collectively, early language development appears to be an important pathway to some aspects of social-emotional functioning in autistic and NT adolescents and adults, beyond the contributions of social functioning.

#### Structural Language Skills

Current structural language performance (grammaticality judgement) was an important factor in some social-emotional outcomes across participants, over and above current social skills. However, the precise role current structural language played in these social-emotional outcomes differed remarkably in individuals with a current autism diagnosis relative to individuals with LAD and NT peers. Individuals with autism who had relatively better structural language had fewer anxiety and depression symptoms and better quality of social relationships, whereas LAD and NT individuals who had relatively better structural language had *more* anxiety and depression symptoms. These symptoms appeared to be within the non-clinical range for the LAD and NT groups, however, suggesting that individual differences in structural language in the current sample were associated with relatively good psychiatric functioning. Given that structural language performance was relatively high in our NT and LAD groups, it would be useful for future studies to examine how structural language is associated with psychiatric functioning in ways that the current study was not able to capture, such as for individuals with structural language impairment. Additionally, LAD individuals with relatively better structural language also had *worse* quality of social relationships.

For the ASD group, these findings align with literature reporting links between language and social-emotional outcomes in autistic individuals and individuals with language impairment. Durkin et al. (2012) reported no group differences in social-emotional outcomes, such as anxiety and depression, for individuals with language impairment who were not autistic relative to individuals with language impairment and ASD (Durkin et al., 2012; see also Howlin et al., 2000). The lack of group difference suggests the possibility of a specific association between language skills and social-emotional outcomes, independent from ASD features. Patterns reported in Durkin et al. (2012) also converge on findings for the LAD and NT groups in the current study given that there were associations between current language ability and social-emotional outcomes for participants who did not have current ASD features (see also Conti-Ramsden et al., 2013; Haukedal et al., 2023; Larson, 2025). This interpretation is further supported by literature demonstrating poorer social-emotional outcomes for individuals with language impairment without ASD (e.g., increased prevalence of anxiety; Durkin et al., 2012; Conti-Ramsden & Durkin, 2012; Howlin et al., 2000; Mawhood et al., 2000; Whitehouse et al., 2009). In contrast to the current findings, Durkin et al. (2012) found that autism features were associated with friendships, whereas language skills were not associated with friendships when controlling for autism features across language impairment groups. It is possible that the current study detected more subtle associations with social relationships, as well as unexpected patterns that Durkin et al. (2012) did not capture when collapsing across diagnostic groups.

Specifically, an unexpected pattern emerged for LAD individuals; better current language was associated with *more* anxiety/depression symptoms, within the non-clinical range, and *poorer* social relationships, though in the context of generally more positive social relationships than autistic peers. These associations differed significantly from autistic peers but were similar to NT peers for anxiety/depression. Overall, anxiety/depression and social relationship scores do not differ between the LAD and NT group, suggesting that these scores do not reflect clinical-threshold challenges regardless of language skills. It is possible that any elevated scores in these groups reflect challenges related to the COVID-19 pandemic, given that these data were largely collected during and shortly after the pandemic. Associations between language and these social-emotional outcomes observed in the current study mirror preliminary evidence from a developmental language disorder group. Relatively better structural language (grammaticality judgement) was associated with lower ratings of happiness and the ability to ‘be yourself,’ as well as with experiencing more environmental barriers, in a small developmental language disorder sample (Larson, 2025).

Though speculative, it is possible that language, in the form of verbal mediation (i.e., self-talk), results in an inner-monologue that leads to self-imposed pressure or heightened expectations (e.g., verbal mediation is associated with slower initiation of goal-directed activities in individuals with language impairment and individuals with ASD; Larson et al., 2019; Larson et al., 2020a). LAD individuals may experience this effect due in part to having been diagnosed with autism and overcoming functional and societal barriers associated with their early autistic features or diagnostic status. Although they no longer meet criteria for autism, they may continue to experience some barriers (e.g., subtle adaptive or social challenges) that result in anxiety and depression symptoms, particularly affecting those with relatively strong language skills who may be *more able* or have a *greater desire* to participate in neurotypical environments (e.g., Larson, 2025). For instance, LAD individuals may place pressure on themselves (potentially through a language-based inner-monologue) to perform well on standardized testing or within social contexts (e.g., Buckley et al., 2021). Similarly, the emerging literature on camouflaging, the use of strategies to mask autistic traits in order to be socially accepted, suggests that this behavior is associated with anxiety and depression in both autistic and non-autistic populations (Bernardin et al., 2021; Bargiela et al., 2016; Hull et al., 2017). Camouflaging (i.e., monitoring others’ reactions and adjusting social behavior in response) may be a verbally mediated process that contributes to the association between language and anxiety/depression symptoms and social relationships for the LAD group; though, emerging evidence suggests that LAD individuals do not engage in significantly more camouflaging than NT peers (Dieckhaus, 2023). These interpretations are speculative given the novelty of this work, but are based on our clinical observations, and on literature indicating anxiety in autistic individuals who are highly independent and who are high performers (e.g., at work, during music auditions, and during social interactions; Buckley et al., 2021), some of whom may have LAD status. For autistic individuals, relatively strong structural language skills appeared to be a protective factor in internalizing symptoms and disorders and social relationships. In contrast, mental health and relationships for autistic individuals who *struggle with language* may be of particular concern; they may experience both social *and* language challenges as barriers to their well-being. It is critical to also bear in mind that these patterns were present over and above the influence of individual differences in social skills, suggesting a specific pathway of influence related to concurrent structural language skills.

### Language and Educational and Vocational Outcomes

The current study demonstrated several preliminary associations between language and educational and vocational outcomes, though the sample sizes were small for this set of outcome measures. Developmental language milestones were associated with education completed by 22 years of age for the ASD and LAD groups and with hours working per week for individuals older than 22 years across groups. Note that we selected this age cutoff to reflect anticipated working status after completing potential college coursework, and the results and sample size did not change when we lowered the age cutoff to 18 years.

Relatively older age of first phrases was associated with more education completed for the ASD and LAD groups, and this pattern appeared to be driven by the LAD group. This finding suggests that LAD individuals with relatively later developmental language milestones completed more education than LAD individuals with relatively earlier milestones. Relatively older age of first words was also associated with more working hours per week for the ASD and LAD groups, but with fewer working hours per week for the NT group. The former pattern appeared to be driven by the ASD group and suggests that autistic individuals with relatively later developmental language milestones were employed at full time status to a greater degree than autistic individuals with relatively earlier milestones.

Our findings do not clearly align with prior research on educational and vocational outcomes. Previous studies suggest that individuals who experience atypical early language development have poorer educational and vocational outcomes than NT peers (Howlin et al., 2000; Mawhood et al., 2000; Whitehouse et al., 2009). In the current study, however, we observed consistency in the associations between language milestones and these education outcomes for the LAD group and vocation outcomes for the ASD group; these patterns also align with social-emotional findings that relatively better current language or earlier language acquisition was associated with poorer outcomes. Given the consistency among patterns in the current study, we may interpret these effects as reflecting contextual factors, such as being placed into more neurotypical contexts, or intervention factors, such as availability of fewer support services for those with less versus more severe language delay.

Language-mediated camouflaging may be an additional factor that is necessary to function in neurotypical contexts, especially in the absence of support services. Indeed, qualitative data on supports at school and work (Supplemental Materials 1, Table 5) suggest that some autistic and LAD individuals receive accommodations, like extra time on tests and note-takers, though these supports may not sufficiently reduce barriers to participating in neurotypical settings. Only seven autistic individuals and three LAD individuals contributed data to this question, so it is difficult to draw conclusions regarding barriers and supports. However, relatively few autistic and LAD individuals self-disclosed their education and vocation information overall (Table 2). Collectively, autistic and LAD individuals with relatively mild early language delay may be at risk for unexpected barriers to educational and vocational achievement that reflect additional, potentially contextual– and support service-related, factors. These interpretations should be examined in future studies.

### Limitations

The primary limitation of this work is the limited and unbalanced sample size. Given that we had two cohorts of participants and much of our educational and vocational data were collected remotely using Qualtrics surveys, we had unbalanced sample sizes for the standardized social-emotional data relative to the survey data. It is possible that some missingness and elevated challenges in outcomes are related to data being partly collected during the COVID-19 pandemic, as well as to individual factors, such as group differences in self-disclosure. Future work would benefit from collecting more balanced samples of outcomes data, such as hours working per week and quality of life self-report surveys. Future work would also benefit from analyzing the precise occupations and supports at school/work for autistic and LAD individuals with a larger sample to better understand language demands and needs in the workplace and other neurotypical contexts, as well as the role of sex and gender identity in long-term outcomes given the disparity in gender identity among groups in the current study.

Another limitation of this work is the lack of standardized language measures in primary analyses. Though we had standardized language data for some of our participants, the sample size of the adolescent cohort was too small to use these data in analyses. One consequence of this limitation was being unable examine the role of possible structural language impairment in autistic or LAD individuals, which has been shown in prior work to be present in both groups (Larson et al., 2022) and relevant to outcomes (Durkin et al., 2012). Future work should examine associations between language and outcomes for individuals with versus without language impairment as associations between language and various measures of functioning may differ depending on language impairment status (e.g., Larson et al., 2019; Larson et al., 2020b).

## Conclusions

This study showed that early language milestone acquisition and concurrent structural language skills were associated with some social-emotional, educational, and vocational outcomes in adolescents and young adults with autism, LAD, and NT status. These associations were evident even after accounting for the potential confound of close relationships between language and social functioning, suggesting a distinct influence of language. Yet, language appeared to *operate differently* as a factor in long-term outcomes in autism, suggesting differences in concurrent and developmental associations for individuals who retain the autism diagnosis relative to LAD individuals who no longer meet criteria for autism and relative to NT peers.

Language milestones and concurrent language played a protective role in several social-emotional outcomes for autistic individuals, including anxiety/depression symptomology and social relationships. These findings align with prior work suggesting that early factors contribute to the developmental trajectory of autism and that language impacts long-term outcomes in autistic and individuals. In LAD and NT adolescents and young adults, relatively better current language was associated with worse anxiety/depression symptomology. Additional unexpected patterns emerged for the ASD and LAD groups that differed from those observed in NT peers.

Younger age of first words was associated with worse quality of life for the ASD group and fewer hours working per week for the ASD and LAD groups. Younger age of first phrases was also associated with less education completed for the LAD group. These unexpected patterns may suggest that individuals with autism and a history of autism experience challenges as a function of their language-based inner-monologue or camouflaging/masking and participation challenges in neurotypical settings. Though these interpretations are speculative, they align with literature reporting increased anxiety in highly independent and high performing autistic individuals and anxiety associated with camouflaging in autistic *and* non-autistic individuals.

Collectively, this evidence suggests that current language and history of language development must be considered in social-emotional functioning and as part of educational and vocational supports from childhood *through* adulthood for individuals diagnosed with autism in childhood.

## Data Availability

All data produced in the present study are available upon reasonable request to the authors

## Acknowledgements

We gratefully acknowledge funding from the National Institutes of Health R01MH112687-01A1 and T32DC017703. The authors would like to thank the study participants and their families who made this research possible, and research assistants Karina Patel and Marissa Birmingham for their contributions.

## Ethics

All procedures were in accordance with the ethical standards of the institutional review committee and with the 1964 Helsinki declaration and later amendments. The study was approved by the Institutional Review Board at the University of Connecticut. Informed consent was obtained from all participants.

## Pre-registration

https://osf.io/tzvpq

## Data Accessibility

The data that support the findings of this study are available from the National Institute of Mental Health Data Archive (NDA) and OSF (https://osf.io/vxukc).

## Authors’ Contributions

(CRediT) Statement: **Caroline Larson** – Conceptualization, Methodology, Validation, Formal analysis, Visualization, Writing – original draft. **Elise Taverna** – Conceptualization, Investigation, Data curation, Writing – review/editing. **Anusha Mohan** – Conceptualization, Investigation, Data curation, Writing – review/editing. **Teresa Girolamo** – Conceptualization, Writing – review/editing. **Deb Fein** – Funding acquisition, Methodology, Resources, and Writing – review/editing. **Inge-Marie Eigsti** – Funding acquisition, Methodology, Validation, Investigation, Resources, Data curation, Project administration, Supervision, Visualization, and Writing – review/editing.

