## Supplementary material for "The role of language in social-emotional, educational, and vocational outcomes in autism and in individuals who have lost the diagnosis": Language LTO_Supplementary Materials 1_9-2-25 (from another version of this preprint: version 1, 2025-09-12)

**Supplemental Materials 1**

**Grammaticality Judgement Stimuli**

Grammatical # GJ-1. Right this minute the old ship is sinking in the harbor.

Grammatical # GJ-2. The boy lives with his father every summer.

Grammatical # GJ-3. Yesterday the photographer shot a deer.

Grammatical # GJ-4. By the time Jennifer arrives, I will have finished dressing.

Ungrammatical # GJ-5. By the time Mr. Martin finishes college, he will have take 32 classes.

Ungrammatical # GJ-6. The girl cooks eggs for her family last night.

Ungrammatical # GJ-7. Right now Judy shops for a new party dress.

Ungrammatical # GJ-8. Yesterday the man is standing in the rain for his bus.

Grammatical # GJ-9. Every night after the family eats dinner, Sammy's dad does the dishes.

Grammatical # GJ-10. The red-haired older woman likes to listen to the news on her iPad.

Ungrammatical # GJ-11. On Sunday afternoons in the winter, Ashley exercise at the gym with her friends.

Ungrammatical # GJ-12. Because he gets really hungry after school, Franklin usually eat a snack.

Grammatical # GJ-13. John's dog always waits for him at the corner.

Grammatical # GJ-14. Every Friday our neighbor washes her car.

Ungrammatical # GJ-15. Every day Terri talk with her mom on the phone.

Ungrammatical # GJ-16. Mrs. Sampson clean her house every Wednesday.

Grammatical # GJ-17. Mrs. Newport will be leaving the party early.

Grammatical # GJ-18. The cat is hiding underneath the couch.

Grammatical # GJ-19. The kids would have eaten by now.

Grammatical # GJ-20. Sue normally would have been saying her prayers by now.

Ungrammatical # GJ-21. Leonard should has written a letter to his mother.

Ungrammatical # GJ-22. Arnold may have be sleeping when we called.

Ungrammatical # GJ-23. The baby bird has fall from the oak tree.

Ungrammatical # GJ-24. The Johnsons may is moving to Chicago this fall.

Grammatical # GJ-25. The horse jumped over the fence yesterday.

Grammatical # GJ-26. Kevin called up Amy for a date.

Grammatical # GJ-27. The man climbed up the ladder carefully.	###

Ungrammatical # GJ-28. Her cousin usually cooked on the grill.

Ungrammatical # GJ-29. The striped cat gracefully jumped the counter off.

Ungrammatical # GJ-30. Tom called the hall down to his mother.

Ungrammatical # GJ-31. They stood the line very patiently.

Ungrammatical # GJ-32. The man looked the new cars at yesterday.

Grammatical # GJ-33. A bat flew into our attic last night.

Grammatical # GJ-34. John sang for the church choir yesterday.

Grammatical # GJ-35. Yesterday she tried a new recipe.

Grammatical # GJ-36. Mr. Murphy hid his money under his mattress yesterday.

Grammatical # GJ-37. Last night the baby fell out of her crib.

Grammatical # GJ-38. Sandy filled the jar with cookies last night.

Grammatical # GJ-39. A policeman gave Tom a ticket for speeding yesterday.

Ungrammatical # GJ-40. Last night the old woman satted on her porch.

Ungrammatical # GJ-41. Where did Arnie hunted last year?

Ungrammatical # GJ-42. Yesterday the man make chocolate cake.

Ungrammatical # GJ-43. Last week the baby throwed a cat into the bathtub.

Ungrammatical # GJ-44. Yesterday the teacher sented Tommy to the principal.

Ungrammatical # GJ-45. Did Bobby stayed at home last night?

Ungrammatical # GJ-46. Last night the old lady dream in her sleep.

**Vocation** **and** **Education Measures**

We categorized participants based on current activities using the following questions on work versus school classification variables:

What best describes your daily activities (multiple choice)? full-time postsecondary degree-seeking program without supports (e.g. college), part-time degree-seeking education/employment without supports (10 hours/week or less), employed in the community without supports, full-time employment in the community with supports (11 hours/week or more), part-time employment in the community with supports (10 hours/week or less), sheltered or supported vocational setting with community employment; sheltered or supported vocational setting with volunteering, sheltered or supported vocational setting, sheltered or supported vocational setting, part-time (10 hours/week or less), volunteering or non-degree-seeking education (e.g. single classes) only, no vocational/educational activities, still in high school.

Then we analyzed responses to the following questions:

| Question | Response Type |
| --- | --- |
| How much education have you completed so far? | multiple choice |
| Are you currently attending classes at a university? | yes/no |
| Do you receive any supports/accommodations? | yes/no and text entry |
| How many classes do you take each semester? | text entry |
| Do you have a job? | Yes/no |
| What is your job title and responsibilities? | text entry |
| How many hours/week do you work? | multiple choice: 1-10; 11-20; 21-34; 35 or more |
| How long have you worked at this job? | enter # of years, months or weeks |
| Is there support at this job (e.g. a job coach)? | yes/no and text entry |
| Approximate yearly, monthly, or weekly salary | text entry |
| Do you participate in a sheltered/supported vocational program? | yes/no |

Yes/no (i.e., 0,1) responses and numerical responses (e.g., hours/week spent working; salary) will be statistically analyzed. Text entry responses will be used for descriptive information to further clarify statistical relationships observed for the friendship/social support scale (e.g., job title and responsibilities).

**Sociodemographic Questionnaire**

From the MacArthur Network on SES and Health (Adler, 2007)

**Question 1**:

What is the highest grade (or year) of regular school you have completed? (Check one.)

| Elementary School | High School | College | Graduate School |
| --- | --- | --- | --- |
| 01____ | 09____ | 13_____ | 17_____ |
| 02____ | 10____ | 14_____ | 18_____ |
| 03____ | 11____ | 15_____ | 19_____ |
| 04____ | 12____ | 16_____ | 20+____ |
| 05____ |  |  |  |
| 06____ |  |  |  |
| 07____ |  |  |  |
| 08____ |  |  |  |

**Data coding:** Continuous depending on responses; categorical if ≤ 3 unique responses (e.g., 16, 18, 20).

**Question 2**:

Which of these categories best describes your total combined family income for the past 12 months?

This should include income (before taxes) from all sources, wages, rent from properties, social security, disability and/or veteran's benefits, unemployment benefits, workman's compensation, help from relatives (including child payments and alimony), and so on.

_____Less than $5,000
_____$5,000 through $11,999
_____$12,000 through $15,999
_____$16,000 through $24,999
_____$25,000 through $34,999
_____$35,000 through $49,999
_____$50,000 through $74,999
_____$75,000 through $99,999
_____$100,000 and greater
_____Don't know

| **Table 1.** *Parent socioeconomic characteristics.* | | | | |
| --- | --- | --- | --- | --- |
| Socioeconomic Variable | NT (*n* = 38) | LAD (*n* = 32) | ASD (*n* = 39) |  |
| Parent Highest | 16.64 | 18.07 | 17.19 | NT-ASD *p* = .35 |
| Education | (2.50) | (1.96) | (2.22) | NT-LAD *p* < .05** |
|  |  |  |  | ASD-LAD *p* = .11 |
| Family Income | 89,896 | 97,222 | 89,689 | NT-ASD *p* = .97 |
|  | (24,171) | (10,591) | (21,711) | NT-LAD *p* = .15 |
|  |  |  |  | ASD-LAD *p* = .10 |
| *Note*. Parent report was predominantly biological mothers if parents self-identified; **p* < .05; ***p* < .01; ****p* < .001 | | | | |

| **Table 2.**  *Additional descriptive data for all language measures.* | | | | |
| --- | --- | --- | --- | --- |
| Language Variable | NT (*n* = 38) | LAD (*n* = 32) | ASD (*n* = 39) |  |
| GJ % accuracy | 94.75 | 94.83 | 89.88 | NT-ASD *p* < .05* |
|  | (3.26) | (3.92) | (10.57) | NT-LAD *p* = .93 |
|  |  |  |  | ASD-LAD *p* < .0*5 |
| Age of first words |  |  |  |  |
| *n* > 18 months vs. *n* ≤ 18 months | 2:31 | 12:13 | 13:13 |  |
| Age of first phrases |  |  |  |  |
| *n* > 30 months vs. *n* ≤ 30 months | 2:31 | 8:27 | 12:14 |  |
| CELF-5 Core Language | 115.6 | 106.8 | 103.9 | NT-ASD *p* < .05* |
| Standard Scores | (11.31) | (11.58) | (12.32) | NT-LAD *p* = .19 |
| *n* = | 11 | 5 | 15 | ASD-LAD *p* = .64 |
| CELF-5 Recalling Sentences | 12.55 | 10.60 | 10.93 | NT-ASD *p* = .19 |
| Scaled Scores | (2.70) | (3.97) | (3.41) | NT-LAD *p* = .36 |
| *n* = |  |  | 15 | ASD-LAD *p* = .87 |
| *Note*. GJ % accuracy = Grammaticality Judgement percent accuracy, *r* = .32*** with A`; Age of first words/phrases = in months, from parent report; CELF-5 = Clinical Evaluation of Language Fundamentals, 5^th^ Edition; **p* < .05; ***p* < .01; ****p* < .001 | | | | |

| **Table 3.**  *Additional descriptive data on educational and vocational outcomes.* | | | | | | | |
| --- | --- | --- | --- | --- | --- | --- | --- |
| Education/Vocation Variable | | NT (*n* = 38) | | LAD (*n* = 32) | ASD (*n* = 39) |  | |
| High school vs. not in high school | | 7:25 | | 2:26 | 10:21 | NT-ASD *p* = .52 | |
|  | |  | |  |  | NT-LAD *p* = .22 | |
|  | |  | |  |  | ASD-LAD *p* < .05* | |
| Currently in classes vs. not | |  | |  |  |  | |
| currently in classes at university | |  | |  |  | NT-ASD *p* = .23;   *p* = .20 | |
| 18-22 years old | | 4:1 | | 10:1 | 3:4 | NT-LAD *p* = .64;  *p* = .35 | |
| > 22 years | | 9:9 | | 5:10 | 3:8 | ASD-LAD *p*  .06  *p* = .75 | |
| Number of classes per semester | | 4.29 | | 4.00 | 3.83 | NT-ASD *p* = .42 | |
| (for participants in college) | | (0.95) | | (1.48) | (0.98) | NT-LAD *p* = .63 | |
| *n* = | | 7 | | 11 | 6 | ASD-LAD *p* = .79 | |
| Has a job vs. does not have a job | |  | |  |  |  | |
| > 22 years old and not in college | | 6:3 | | 10:0 | 6:2 | NT-ASD *p* = 1.00 | |
| > 18 years old and not in college | | 6:4 | | 10:1 | 6:6 | NT-LAD *p* = .17 | |
|  | |  | |  |  | ASD-LAD *p* = .35 | |
| Hours Working per week at job | | 38.00 | | 40.00 | 33.67 | NT-ASD *p* = .22 | |
| > 18 years old and not in college | | (3.10) | | (0.00) | (7.21) | NT-LAD *p* = .17 | |
| *n* = | | 6 | | 10 | 6 | ASD-LAD *p* = .09 | |
| Receiving supports vs. not | |  | |  |  | NT-ASD *p* < .05* | |
| receiving | | 2:11 | | 2:13 | 5:2 | NT-LAD *p* = 1.00 | |
| (at school or work) | |  | |  |  | ASD-LAD *p* < .05* | |
| *Note.* Hours working per week at job: results identical for > 18 years and > 22 years; **p* < .05; ***p* < .01; ****p* < .001 | | | | | | | |
| **Table 4.**  *Qualitative data on supports.* | | | | | | | |
|  | ASD | | LAD | | | | NT |
| Supports at school | 5 Extra time, quiet room, note-taker, weekly tutoring, disability service meetings 1 Reduced courseload | | 1 Extra time on tests,  1 Accommodations for housing | | | | 1 Academic (unspecified) |
| Supports at work | 1 Supervisors/coaches available | | 1 Follow directions/  ask for help | | | | 1 Training classes 1 Mentor |

**Figure 1.**

*The association between quality of life and age of first words for each group.*

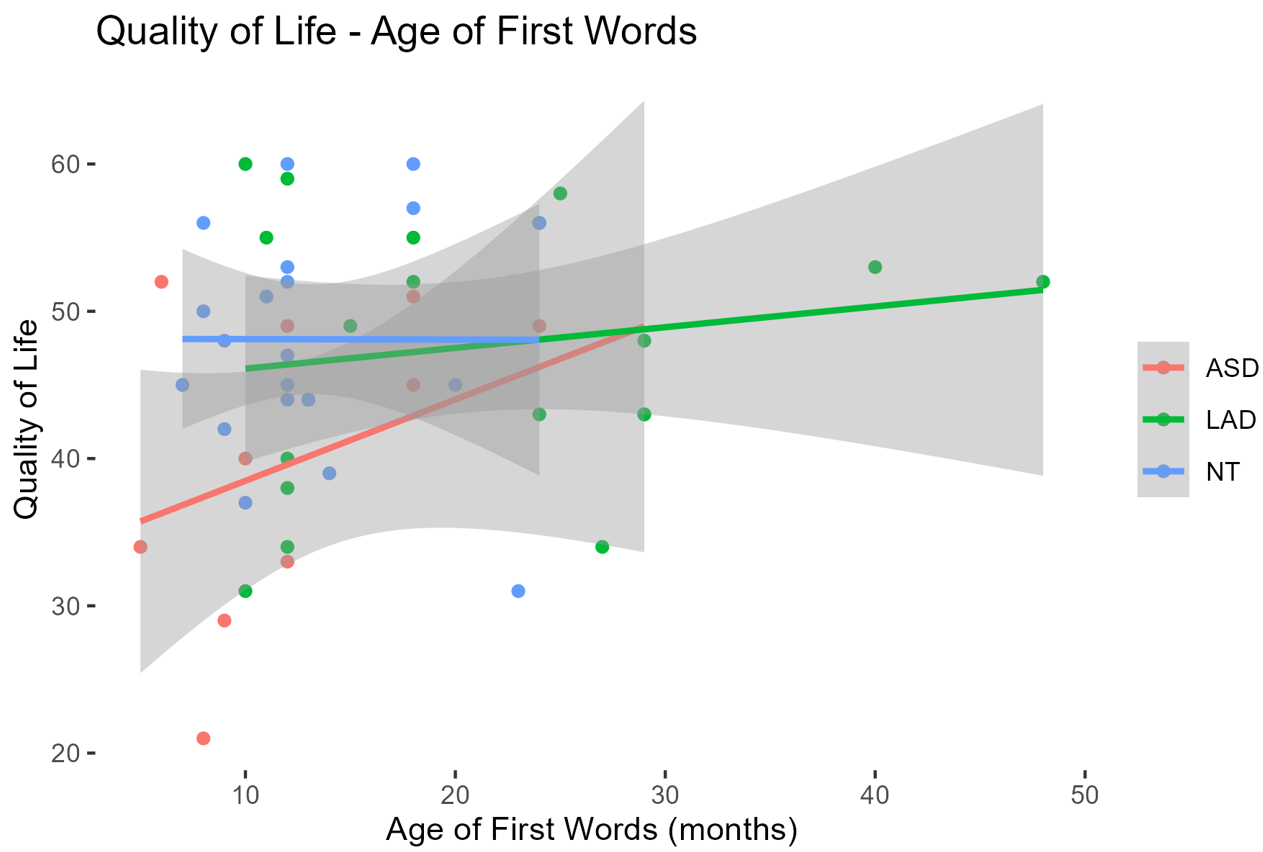

*Note.* Grey bars indicate standard error.
