## Supplementary material for "The role of language in social-emotional, educational, and vocational outcomes in autism and in individuals who have lost the diagnosis": Language LTO_Supplementary Materials 2_9-2-25 (from another version of this preprint: version 1, 2025-09-12)

**Supplemental Materials 2**

```
> #### Surveys
> #QoLLifeSatSocSupp.log
> ### ASD LAD ###
> mSurvey_ASDLAD.GJ = lm(QoLLifeSatSocSupp.log ~
+                           Group.NTref*aprime.log
+                  #+ age
+                       + vabs_soc_ss
+                         , data = dASDLAD.older18)
> summary(mSurvey_ASDLAD.GJ)

Call:
lm(formula = QoLLifeSatSocSupp.log ~ Group.NTref * aprime.log +
    vabs_soc_ss, data = dASDLAD.older18)

Residuals:
     Min       1Q   Median       3Q      Max
-0.34659 -0.11645  0.03970  0.08941  0.25976

Coefficients:
                        Estimate Std. Error t value Pr(>|t|)
(Intercept)             4.224747   0.329352  12.827 1.09e-11 ***
Group.NTref             0.135963   0.171253   0.794    0.436
aprime.log             -0.433855   0.456465  -0.950    0.352
vabs_soc_ss             0.004307   0.003289   1.309    0.204
Group.NTref:aprime.log -0.483766   0.954952  -0.507    0.617
---
Signif. codes:  0 ‘***’ 0.001 ‘**’ 0.01 ‘*’ 0.05 ‘.’ 0.1 ‘ ’ 1

Residual standard error: 0.1811 on 22 degrees of freedom
  (28 observations deleted due to missingness)
Multiple R-squared:  0.3207,	Adjusted R-squared:  0.1972
F-statistic: 2.597 on 4 and 22 DF,  p-value: 0.06432

>
> mSurvey_ASDLAD.Words = lm(QoLLifeSatSocSupp.log ~
+                           Group.NTref*FirstWords.months
+                  #+ age
+                       + vabs_soc_ss
+                         , data = dASDLAD.older18)
> summary(mSurvey_ASDLAD.Words)

Call:
lm(formula = QoLLifeSatSocSupp.log ~ Group.NTref * FirstWords.months +
    vabs_soc_ss, data = dASDLAD.older18)

Residuals:
     Min       1Q   Median       3Q      Max
-0.32569 -0.08547 -0.00326  0.12806  0.36735

Coefficients:
                               Estimate Std. Error t value Pr(>|t|)
(Intercept)                    4.203944   0.448718   9.369 3.99e-08 ***
Group.NTref                    0.290059   0.209922   1.382    0.185
FirstWords.months              0.006669   0.006899   0.967    0.347
vabs_soc_ss                    0.003883   0.004125   0.941    0.360
Group.NTref:FirstWords.months -0.008860   0.013716  -0.646    0.527
---
Signif. codes:  0 ‘***’ 0.001 ‘**’ 0.01 ‘*’ 0.05 ‘.’ 0.1 ‘ ’ 1

Residual standard error: 0.1944 on 17 degrees of freedom
  (33 observations deleted due to missingness)
Multiple R-squared:  0.2932,	Adjusted R-squared:  0.1268
F-statistic: 1.763 on 4 and 17 DF,  p-value: 0.1829

>
> mSurvey_ASDLAD.Phrases = lm(QoLLifeSatSocSupp.log ~
+                           Group.NTref*FirstPhrases.months
+                  #+ age
+                       + vabs_soc_ss
+                         , data = dASDLAD.older18)
> summary(mSurvey_ASDLAD.Phrases)

Call:
lm(formula = QoLLifeSatSocSupp.log ~ Group.NTref * FirstPhrases.months +
    vabs_soc_ss, data = dASDLAD.older18)

Residuals:
     Min       1Q   Median       3Q      Max
-0.34335 -0.08685  0.00699  0.11825  0.30209

Coefficients:
                                Estimate Std. Error t value Pr(>|t|)
(Intercept)                     4.311556   0.368904  11.687 7.71e-10 ***
Group.NTref                     0.114105   0.235034   0.485    0.633
FirstPhrases.months             0.002078   0.003459   0.601    0.556
vabs_soc_ss                     0.003147   0.003428   0.918    0.371
Group.NTref:FirstPhrases.months 0.003199   0.006945   0.461    0.651
---
Signif. codes:  0 ‘***’ 0.001 ‘**’ 0.01 ‘*’ 0.05 ‘.’ 0.1 ‘ ’ 1

Residual standard error: 0.1866 on 18 degrees of freedom
  (32 observations deleted due to missingness)
Multiple R-squared:  0.3741,	Adjusted R-squared:  0.235
F-statistic: 2.689 on 4 and 18 DF,  p-value: 0.06441

>
>
>
>
> ### ASD NT ###
> mSurvey_ASDNT.GJ = lm(QoLLifeSatSocSupp.log ~
+                           Group.LADref*aprime.log
+                 #+ age
+                       + vabs_soc_ss
+                         , data = dASDNT.older18)
> summary(mSurvey_ASDNT.GJ)

Call:
lm(formula = QoLLifeSatSocSupp.log ~ Group.LADref * aprime.log +
    vabs_soc_ss, data = dASDNT.older18)

Residuals:
     Min       1Q   Median       3Q      Max
-0.45061 -0.10279  0.05645  0.13469  0.23583

Coefficients:
                        Estimate Std. Error t value Pr(>|t|)
(Intercept)             3.957773   0.373839  10.587    1e-10 ***
Group.LADref            0.151629   0.177696   0.853   0.4016
aprime.log              0.058508   0.459872   0.127   0.8998
vabs_soc_ss             0.007492   0.003837   1.952   0.0622 .
Group.LADref:aprime.log 0.198679   0.904008   0.220   0.8278
---
Signif. codes:  0 ‘***’ 0.001 ‘**’ 0.01 ‘*’ 0.05 ‘.’ 0.1 ‘ ’ 1

Residual standard error: 0.1938 on 25 degrees of freedom
  (22 observations deleted due to missingness)
Multiple R-squared:  0.294,	Adjusted R-squared:  0.181
F-statistic: 2.603 on 4 and 25 DF,  p-value: 0.06025

>
> mSurvey_ASDNT.Words = lm(QoLLifeSatSocSupp.log ~
+                           Group.LADref*FirstWords.months
+                 #+ age
+                       + vabs_soc_ss
+                         , data = dASDNT.older18)
> summary(mSurvey_ASDNT.Words)

Call:
lm(formula = QoLLifeSatSocSupp.log ~ Group.LADref * FirstWords.months +
    vabs_soc_ss, data = dASDNT.older18)

Residuals:
    Min      1Q  Median      3Q     Max
-0.3806 -0.0790 -0.0045  0.1381  0.3191

Coefficients:
                                Estimate Std. Error t value Pr(>|t|)
(Intercept)                     3.646037   0.472895   7.710 1.48e-07 ***
Group.LADref                    0.443610   0.213192   2.081   0.0499 *
FirstWords.months               0.003903   0.007600   0.514   0.6130
vabs_soc_ss                     0.009900   0.004395   2.253   0.0351 *
Group.LADref:FirstWords.months -0.021405   0.015242  -1.404   0.1748
---
Signif. codes:  0 ‘***’ 0.001 ‘**’ 0.01 ‘*’ 0.05 ‘.’ 0.1 ‘ ’ 1

Residual standard error: 0.1874 on 21 degrees of freedom
  (26 observations deleted due to missingness)
Multiple R-squared:  0.3857,	Adjusted R-squared:  0.2686
F-statistic: 3.296 on 4 and 21 DF,  p-value: 0.03031

>
> mSurvey_ASDNT.Phrases = lm(QoLLifeSatSocSupp.log ~
+                           Group.LADref*FirstPhrases.months
+                 #+ age
+                       + vabs_soc_ss
+                         , data = dASDNT.older18)
> summary(mSurvey_ASDNT.Phrases)

Call:
lm(formula = QoLLifeSatSocSupp.log ~ Group.LADref * FirstPhrases.months +
    vabs_soc_ss, data = dASDNT.older18)

Residuals:
     Min       1Q   Median       3Q      Max
-0.43674 -0.11871  0.05183  0.12935  0.27961

Coefficients:
                                   Estimate Std. Error t value Pr(>|t|)
(Intercept)                       3.9895459  0.4111278   9.704 8.81e-10 ***
Group.LADref                      0.2160642  0.2510017   0.861   0.3979
FirstPhrases.months               0.0001518  0.0043055   0.035   0.9722
vabs_soc_ss                       0.0068621  0.0037752   1.818   0.0816 .
Group.LADref:FirstPhrases.months -0.0021580  0.0085363  -0.253   0.8026
---
Signif. codes:  0 ‘***’ 0.001 ‘**’ 0.01 ‘*’ 0.05 ‘.’ 0.1 ‘ ’ 1

Residual standard error: 0.1933 on 24 degrees of freedom
  (23 observations deleted due to missingness)
Multiple R-squared:   0.32,	Adjusted R-squared:  0.2067
F-statistic: 2.824 on 4 and 24 DF,  p-value: 0.04729

>
>
>
>
>
> ### LAD NT ###
> mSurvey_LADNT.GJ = lm(QoLLifeSatSocSupp.log ~
+                           Group.ASDref*aprime.log
+                 #+ age
+                       + vabs_soc_ss
+                         , data = dLADNT.older18)
> summary(mSurvey_LADNT.GJ)

Call:
lm(formula = QoLLifeSatSocSupp.log ~ Group.ASDref * aprime.log +
    vabs_soc_ss, data = dLADNT.older18)

Residuals:
     Min       1Q   Median       3Q      Max
-0.46350 -0.06390  0.04269  0.09133  0.19089

Coefficients:
                         Estimate Std. Error t value Pr(>|t|)
(Intercept)              4.397920   0.315361  13.946 4.38e-16 ***
Group.ASDref             0.068370   0.098067   0.697    0.490
aprime.log              -0.266132   0.373859  -0.712    0.481
vabs_soc_ss              0.003651   0.002938   1.243    0.222
Group.ASDref:aprime.log  0.769114   0.756723   1.016    0.316
---
Signif. codes:  0 ‘***’ 0.001 ‘**’ 0.01 ‘*’ 0.05 ‘.’ 0.1 ‘ ’ 1

Residual standard error: 0.1584 on 36 degrees of freedom
  (16 observations deleted due to missingness)
Multiple R-squared:  0.06307,	Adjusted R-squared:  -0.04103
F-statistic: 0.6059 on 4 and 36 DF,  p-value: 0.6609

>
> mSurvey_LADNT.Words = lm(QoLLifeSatSocSupp.log ~
+                           Group.ASDref*FirstWords.months
+                 #+ age
+                       + vabs_soc_ss
+                         , data = dLADNT.older18)
> summary(mSurvey_LADNT.Words)

Call:
lm(formula = QoLLifeSatSocSupp.log ~ Group.ASDref * FirstWords.months +
    vabs_soc_ss, data = dLADNT.older18)

Residuals:
     Min       1Q   Median       3Q      Max
-0.40468 -0.06519  0.00874  0.11327  0.21374

Coefficients:
                                Estimate Std. Error t value Pr(>|t|)
(Intercept)                     4.450225   0.332810  13.372 2.06e-14 ***
Group.ASDref                    0.145243   0.139154   1.044    0.305
FirstWords.months              -0.002200   0.004305  -0.511    0.613
vabs_soc_ss                     0.003613   0.003096   1.167    0.252
Group.ASDref:FirstWords.months -0.008847   0.008614  -1.027    0.312
---
Signif. codes:  0 ‘***’ 0.001 ‘**’ 0.01 ‘*’ 0.05 ‘.’ 0.1 ‘ ’ 1

Residual standard error: 0.1622 on 31 degrees of freedom
  (21 observations deleted due to missingness)
Multiple R-squared:  0.07194,	Adjusted R-squared:  -0.04781
F-statistic: 0.6008 on 4 and 31 DF,  p-value: 0.6649

>
> mSurvey_LADNT.Phrases = lm(QoLLifeSatSocSupp.log ~
+                           Group.ASDref*FirstPhrases.months
+                 #+ age
+                       + vabs_soc_ss
+                         , data = dLADNT.older18)
> summary(mSurvey_LADNT.Phrases)

Call:
lm(formula = QoLLifeSatSocSupp.log ~ Group.ASDref * FirstPhrases.months +
    vabs_soc_ss, data = dLADNT.older18)

Residuals:
     Min       1Q   Median       3Q      Max
-0.45082 -0.08263  0.01791  0.11748  0.22846

Coefficients:
                                  Estimate Std. Error t value Pr(>|t|)
(Intercept)                       4.483334   0.328860  13.633 4.13e-15 ***
Group.ASDref                      0.128633   0.121269   1.061    0.297
FirstPhrases.months               0.001151   0.002558   0.450    0.656
vabs_soc_ss                       0.002661   0.003068   0.867    0.392
Group.ASDref:FirstPhrases.months -0.005089   0.005132  -0.992    0.329
---
Signif. codes:  0 ‘***’ 0.001 ‘**’ 0.01 ‘*’ 0.05 ‘.’ 0.1 ‘ ’ 1

Residual standard error: 0.1611 on 33 degrees of freedom
  (19 observations deleted due to missingness)
Multiple R-squared:  0.09605,	Adjusted R-squared:  -0.01352
F-statistic: 0.8766 on 4 and 33 DF,  p-value: 0.4884

>


> ###Follow up
> #aprime.log
> mSurvey_ASD.GJ = lm(QoLLifeSatSocSupp.log ~ aprime.log
+                 #+ age
+                       + vabs_soc_ss
+                         , data = dASD.older18)
> summary(mSurvey_ASD.GJ)

Call:
lm(formula = QoLLifeSatSocSupp.log ~ aprime.log + vabs_soc_ss,
    data = dASD.older18)

Residuals:
       8       12       17       19       20       21       23       24
-0.14265  0.19843  0.23562  0.09568 -0.30481 -0.34031  0.18420  0.07384

Coefficients:
             Estimate Std. Error t value Pr(>|t|)
(Intercept)  3.833464   0.674185   5.686  0.00234 **
aprime.log  -0.014160   1.020096  -0.014  0.98946
vabs_soc_ss  0.008054   0.007460   1.080  0.32964
---
Signif. codes:  0 ‘***’ 0.001 ‘**’ 0.01 ‘*’ 0.05 ‘.’ 0.1 ‘ ’ 1

Residual standard error: 0.2729 on 5 degrees of freedom
  (17 observations deleted due to missingness)
Multiple R-squared:  0.211,	Adjusted R-squared:  -0.1046
F-statistic: 0.6685 on 2 and 5 DF,  p-value: 0.553

>
> mSurvey_NT.GJ = lm(QoLLifeSatSocSupp.log ~ aprime.log
+                 #+ age
+                       + vabs_soc_ss
+                         , data = dNT.older18)
> summary(mSurvey_NT.GJ)

Call:
lm(formula = QoLLifeSatSocSupp.log ~ aprime.log + vabs_soc_ss,
    data = dNT.older18)

Residuals:
     Min       1Q   Median       3Q      Max
-0.45269 -0.06203  0.05668  0.11705  0.17929

Coefficients:
            Estimate Std. Error t value Pr(>|t|)
(Intercept) 4.097946   0.519491   7.888 2.06e-07 ***
aprime.log  0.151481   0.518033   0.292    0.773
vabs_soc_ss 0.006871   0.004955   1.387    0.182
---
Signif. codes:  0 ‘***’ 0.001 ‘**’ 0.01 ‘*’ 0.05 ‘.’ 0.1 ‘ ’ 1

Residual standard error: 0.1726 on 19 degrees of freedom
  (5 observations deleted due to missingness)
Multiple R-squared:  0.09297,	Adjusted R-squared:  -0.002509
F-statistic: 0.9737 on 2 and 19 DF,  p-value: 0.3957

>
> mSurvey_LAD.GJ = lm(QoLLifeSatSocSupp.log ~ aprime.log
+                 #+ age
+                       + vabs_soc_ss
+                         , data = dLAD.older18)
> summary(mSurvey_LAD.GJ)

Call:
lm(formula = QoLLifeSatSocSupp.log ~ aprime.log + vabs_soc_ss,
    data = dLAD.older18)

Residuals:
     Min       1Q   Median       3Q      Max
-0.32434 -0.06119  0.04628  0.07793  0.20387

Coefficients:
             Estimate Std. Error t value Pr(>|t|)
(Intercept)  4.613745   0.374851  12.308 1.42e-09 ***
aprime.log  -0.562494   0.524591  -1.072    0.300
vabs_soc_ss  0.001344   0.003412   0.394    0.699
---
Signif. codes:  0 ‘***’ 0.001 ‘**’ 0.01 ‘*’ 0.05 ‘.’ 0.1 ‘ ’ 1

Residual standard error: 0.1404 on 16 degrees of freedom
  (11 observations deleted due to missingness)
Multiple R-squared:  0.06798,	Adjusted R-squared:  -0.04852
F-statistic: 0.5835 on 2 and 16 DF,  p-value: 0.5694

>
> #Words
> mSurvey_ASD.Words = lm(QoLLifeSatSocSupp.log ~ FirstWords.months
+                 #+ age
+                       + vabs_soc_ss
+                         , data = dASD.older18)
> summary(mSurvey_ASD.Words)

Call:
lm(formula = QoLLifeSatSocSupp.log ~ FirstWords.months + vabs_soc_ss,
    data = dASD.older18)

Residuals:
        8        12        17        19        20        21
-0.063999  0.295033  0.213546 -0.007325 -0.096208 -0.341047

Coefficients:
                  Estimate Std. Error t value Pr(>|t|)
(Intercept)        3.10760    1.25613   2.474   0.0898 .
FirstWords.months  0.01635    0.02037   0.803   0.4809
vabs_soc_ss        0.01290    0.01162   1.110   0.3479
---
Signif. codes:  0 ‘***’ 0.001 ‘**’ 0.01 ‘*’ 0.05 ‘.’ 0.1 ‘ ’ 1

Residual standard error: 0.2957 on 3 degrees of freedom
  (19 observations deleted due to missingness)
Multiple R-squared:  0.3249,	Adjusted R-squared:  -0.1251
F-statistic: 0.722 on 2 and 3 DF,  p-value: 0.5546

>
> mSurvey_NT.Words = lm(QoLLifeSatSocSupp.log ~ FirstWords.months
+                 #+ age
+                       + vabs_soc_ss
+                         , data = dNT.older18)
> summary(mSurvey_NT.Words)

Call:
lm(formula = QoLLifeSatSocSupp.log ~ FirstWords.months + vabs_soc_ss,
    data = dNT.older18)

Residuals:
     Min       1Q   Median       3Q      Max
-0.38699 -0.03917  0.01580  0.11485  0.20303

Coefficients:
                   Estimate Std. Error t value Pr(>|t|)
(Intercept)        4.040855   0.515796   7.834 4.85e-07 ***
FirstWords.months -0.006753   0.007745  -0.872    0.395
vabs_soc_ss        0.008239   0.004840   1.702    0.107
---
Signif. codes:  0 ‘***’ 0.001 ‘**’ 0.01 ‘*’ 0.05 ‘.’ 0.1 ‘ ’ 1

Residual standard error: 0.1656 on 17 degrees of freedom
  (7 observations deleted due to missingness)
Multiple R-squared:  0.1751,	Adjusted R-squared:  0.07801
F-statistic: 1.804 on 2 and 17 DF,  p-value: 0.1948

>
> mSurvey_LAD.Words = lm(QoLLifeSatSocSupp.log ~ FirstWords.months
+                 #+ age
+                       + vabs_soc_ss
+                         , data = dNT.older18)
> summary(mSurvey_LAD.Words)

Call:
lm(formula = QoLLifeSatSocSupp.log ~ FirstWords.months + vabs_soc_ss,
    data = dNT.older18)

Residuals:
     Min       1Q   Median       3Q      Max
-0.38699 -0.03917  0.01580  0.11485  0.20303

Coefficients:
                   Estimate Std. Error t value Pr(>|t|)
(Intercept)        4.040855   0.515796   7.834 4.85e-07 ***
FirstWords.months -0.006753   0.007745  -0.872    0.395
vabs_soc_ss        0.008239   0.004840   1.702    0.107
---
Signif. codes:  0 ‘***’ 0.001 ‘**’ 0.01 ‘*’ 0.05 ‘.’ 0.1 ‘ ’ 1

Residual standard error: 0.1656 on 17 degrees of freedom
  (7 observations deleted due to missingness)
Multiple R-squared:  0.1751,	Adjusted R-squared:  0.07801
F-statistic: 1.804 on 2 and 17 DF,  p-value: 0.1948

>
> #Phrases
> mSurvey_ASD.Phrases = lm(QoLLifeSatSocSupp.log ~ FirstPhrases.months
+                 #+ age
+                       + vabs_soc_ss
+                         , data = dASD.older18)
> summary(mSurvey_ASD.Phrases)

Call:
lm(formula = QoLLifeSatSocSupp.log ~ FirstPhrases.months + vabs_soc_ss,
    data = dASD.older18)

Residuals:
       8       12       17       19       20       21       24
-0.12441  0.23583  0.28018  0.12137 -0.28242 -0.28685  0.05631

Coefficients:
                    Estimate Std. Error t value Pr(>|t|)
(Intercept)         3.860735   0.835674   4.620  0.00988 **
FirstPhrases.months 0.001273   0.009868   0.129  0.90360
vabs_soc_ss         0.007069   0.007635   0.926  0.40692
---
Signif. codes:  0 ‘***’ 0.001 ‘**’ 0.01 ‘*’ 0.05 ‘.’ 0.1 ‘ ’ 1

Residual standard error: 0.287 on 4 degrees of freedom
  (18 observations deleted due to missingness)
Multiple R-squared:  0.1765,	Adjusted R-squared:  -0.2352
F-statistic: 0.4288 on 2 and 4 DF,  p-value: 0.6781

>
> mSurvey_NT.Phrases = lm(QoLLifeSatSocSupp.log ~ FirstPhrases.months
+                 #+ age
+                       + vabs_soc_ss
+                         , data = dNT.older18)
> summary(mSurvey_NT.Phrases)

Call:
lm(formula = QoLLifeSatSocSupp.log ~ FirstPhrases.months + vabs_soc_ss,
    data = dNT.older18)

Residuals:
     Min       1Q   Median       3Q      Max
-0.43755 -0.05009  0.04774  0.11942  0.18926

Coefficients:
                     Estimate Std. Error t value Pr(>|t|)
(Intercept)          4.123485   0.541306   7.618 3.44e-07 ***
FirstPhrases.months -0.000954   0.004899  -0.195    0.848
vabs_soc_ss          0.006620   0.004969   1.332    0.199
---
Signif. codes:  0 ‘***’ 0.001 ‘**’ 0.01 ‘*’ 0.05 ‘.’ 0.1 ‘ ’ 1

Residual standard error: 0.1728 on 19 degrees of freedom
  (5 observations deleted due to missingness)
Multiple R-squared:  0.0907,	Adjusted R-squared:  -0.005015
F-statistic: 0.9476 on 2 and 19 DF,  p-value: 0.4052

>
> mSurvey_LAD.Phrases = lm(QoLLifeSatSocSupp.log ~ FirstPhrases.months
+                 #+ age
+                       + vabs_soc_ss
+                         , data = dLAD.older18)
> summary(mSurvey_LAD.Phrases)

Call:
lm(formula = QoLLifeSatSocSupp.log ~ FirstPhrases.months + vabs_soc_ss,
    data = dLAD.older18)

Residuals:
     Min       1Q   Median       3Q      Max
-0.24985 -0.07266  0.01029  0.05463  0.25262

Coefficients:
                     Estimate Std. Error t value Pr(>|t|)
(Intercept)          4.739241   0.375504  12.621 1.14e-08 ***
FirstPhrases.months  0.003815   0.002050   1.861   0.0855 .
vabs_soc_ss         -0.000429   0.003562  -0.120   0.9060
---
Signif. codes:  0 ‘***’ 0.001 ‘**’ 0.01 ‘*’ 0.05 ‘.’ 0.1 ‘ ’ 1

Residual standard error: 0.1402 on 13 degrees of freedom
  (14 observations deleted due to missingness)
Multiple R-squared:  0.2104,	Adjusted R-squared:  0.08891
F-statistic: 1.732 on 2 and 13 DF,  p-value: 0.2154


> ###############
> ##### QoL #####
> ###############
>
> ### ASD LAD ###
> mQoL_ASDLAD.GJ = lm(qol.pos.log ~
+                           Group.NTref*aprime.log
+                  #+ age
+                       + vabs_soc_ss
+                         , data = dASDLAD.older18)
> summary(mQoL_ASDLAD.GJ)

Call:
lm(formula = qol.pos.log ~ Group.NTref * aprime.log + vabs_soc_ss,
    data = dASDLAD.older18)

Residuals:
     Min       1Q   Median       3Q      Max
-0.55261 -0.13681  0.08444  0.16995  0.27672

Coefficients:
                        Estimate Std. Error t value Pr(>|t|)
(Intercept)             3.261691   0.436301   7.476 1.78e-07 ***
Group.NTref             0.033309   0.226863   0.147    0.885
aprime.log             -0.323651   0.604690  -0.535    0.598
vabs_soc_ss             0.004664   0.004357   1.071    0.296
Group.NTref:aprime.log -1.058278   1.265049  -0.837    0.412
---
Signif. codes:  0 ‘***’ 0.001 ‘**’ 0.01 ‘*’ 0.05 ‘.’ 0.1 ‘ ’ 1

Residual standard error: 0.2399 on 22 degrees of freedom
  (28 observations deleted due to missingness)
Multiple R-squared:  0.2087,	Adjusted R-squared:  0.06484
F-statistic: 1.451 on 4 and 22 DF,  p-value: 0.251

>
> mQoL_ASDLAD.Words = lm(qol.pos.log ~
+                           Group.NTref*FirstWords.months
+                  #+ age
+                       + vabs_soc_ss
+                         , data = dASDLAD.older18)
> summary(mQoL_ASDLAD.Words)

Call:
lm(formula = qol.pos.log ~ Group.NTref * FirstWords.months +
    vabs_soc_ss, data = dASDLAD.older18)

Residuals:
     Min       1Q   Median       3Q      Max
-0.36438 -0.15523  0.01645  0.14130  0.46856

Coefficients:
                               Estimate Std. Error t value Pr(>|t|)
(Intercept)                    2.883981   0.558928   5.160 7.85e-05 ***
Group.NTref                    0.527993   0.261481   2.019   0.0595 .
FirstWords.months              0.014403   0.008593   1.676   0.1120
vabs_soc_ss                    0.006354   0.005138   1.236   0.2331
Group.NTref:FirstWords.months -0.025798   0.017085  -1.510   0.1494
---
Signif. codes:  0 ‘***’ 0.001 ‘**’ 0.01 ‘*’ 0.05 ‘.’ 0.1 ‘ ’ 1

Residual standard error: 0.2422 on 17 degrees of freedom
  (33 observations deleted due to missingness)
Multiple R-squared:  0.3335,	Adjusted R-squared:  0.1766
F-statistic: 2.126 on 4 and 17 DF,  p-value: 0.122

>
> mQoL_ASDLAD.Phrases = lm(qol.pos.log ~
+                           Group.NTref*FirstPhrases.months
+                  #+ age
+                       + vabs_soc_ss
+                         , data = dASDLAD.older18)
> summary(mQoL_ASDLAD.Phrases)

Call:
lm(formula = qol.pos.log ~ Group.NTref * FirstPhrases.months +
    vabs_soc_ss, data = dASDLAD.older18)

Residuals:
     Min       1Q   Median       3Q      Max
-0.52493 -0.13814  0.00208  0.19920  0.30088

Coefficients:
                                Estimate Std. Error t value Pr(>|t|)
(Intercept)                     3.419389   0.489893   6.980 1.61e-06 ***
Group.NTref                     0.111519   0.312118   0.357    0.725
FirstPhrases.months             0.003028   0.004594   0.659    0.518
vabs_soc_ss                     0.002201   0.004552   0.483    0.635
Group.NTref:FirstPhrases.months 0.003575   0.009223   0.388    0.703
---
Signif. codes:  0 ‘***’ 0.001 ‘**’ 0.01 ‘*’ 0.05 ‘.’ 0.1 ‘ ’ 1

Residual standard error: 0.2478 on 18 degrees of freedom
  (32 observations deleted due to missingness)
Multiple R-squared:  0.2629,	Adjusted R-squared:  0.09908
F-statistic: 1.605 on 4 and 18 DF,  p-value: 0.2162

>
>
>
>
> ### ASD NT ###
> mQoL_ASDNT.GJ = lm(qol.pos.log ~
+                           Group.LADref*aprime.log
+                 #+ age
+                       + vabs_soc_ss
+                         , data = dASDNT.older18)
> summary(mQoL_ASDNT.GJ)

Call:
lm(formula = qol.pos.log ~ Group.LADref * aprime.log + vabs_soc_ss,
    data = dASDNT.older18)

Residuals:
     Min       1Q   Median       3Q      Max
-0.53016 -0.08200  0.01078  0.17808  0.27579

Coefficients:
                        Estimate Std. Error t value Pr(>|t|)
(Intercept)             3.133664   0.427106   7.337  1.1e-07 ***
Group.LADref            0.124843   0.203015   0.615    0.544
aprime.log              0.339891   0.525397   0.647    0.524
vabs_soc_ss             0.006678   0.004384   1.523    0.140
Group.LADref:aprime.log 0.077636   1.032817   0.075    0.941
---
Signif. codes:  0 ‘***’ 0.001 ‘**’ 0.01 ‘*’ 0.05 ‘.’ 0.1 ‘ ’ 1

Residual standard error: 0.2215 on 25 degrees of freedom
  (22 observations deleted due to missingness)
Multiple R-squared:  0.2271,	Adjusted R-squared:  0.1034
F-statistic: 1.837 on 4 and 25 DF,  p-value: 0.1533

>
> mQoL_ASDNT.Words = lm(qol.pos.log ~
+                           Group.LADref*FirstWords.months
+                 #+ age
+                       + vabs_soc_ss
+                         , data = dASDNT.older18)
> summary(mQoL_ASDNT.Words)

Call:
lm(formula = qol.pos.log ~ Group.LADref * FirstWords.months +
    vabs_soc_ss, data = dASDNT.older18)

Residuals:
     Min       1Q   Median       3Q      Max
-0.36088 -0.12970  0.00904  0.12333  0.42482

Coefficients:
                                Estimate Std. Error t value Pr(>|t|)
(Intercept)                     2.350877   0.486535   4.832 8.92e-05 ***
Group.LADref                    0.612517   0.219340   2.793   0.0109 *
FirstWords.months               0.014047   0.007820   1.796   0.0868 .
vabs_soc_ss                     0.011808   0.004522   2.611   0.0163 *
Group.LADref:FirstWords.months -0.032868   0.015682  -2.096   0.0484 *
---
Signif. codes:  0 ‘***’ 0.001 ‘**’ 0.01 ‘*’ 0.05 ‘.’ 0.1 ‘ ’ 1

Residual standard error: 0.1928 on 21 degrees of freedom
  (26 observations deleted due to missingness)
Multiple R-squared:  0.4787,	Adjusted R-squared:  0.3794
F-statistic:  4.82 on 4 and 21 DF,  p-value: 0.006452

>
> mQoL_ASDNT.Phrases = lm(qol.pos.log ~
+                           Group.LADref*FirstPhrases.months
+                 #+ age
+                       + vabs_soc_ss
+                         , data = dASDNT.older18)
> summary(mQoL_ASDNT.Phrases)

Call:
lm(formula = qol.pos.log ~ Group.LADref * FirstPhrases.months +
    vabs_soc_ss, data = dASDNT.older18)

Residuals:
     Min       1Q   Median       3Q      Max
-0.47365 -0.09144  0.02329  0.15733  0.31099

Coefficients:
                                   Estimate Std. Error t value Pr(>|t|)
(Intercept)                       3.1390660  0.4714169   6.659  6.9e-07 ***
Group.LADref                      0.2137351  0.2878094   0.743    0.465
FirstPhrases.months               0.0016754  0.0049369   0.339    0.737
vabs_soc_ss                       0.0055018  0.0043288   1.271    0.216
Group.LADref:FirstPhrases.months -0.0004682  0.0097881  -0.048    0.962
---
Signif. codes:  0 ‘***’ 0.001 ‘**’ 0.01 ‘*’ 0.05 ‘.’ 0.1 ‘ ’ 1

Residual standard error: 0.2217 on 24 degrees of freedom
  (23 observations deleted due to missingness)
Multiple R-squared:  0.2477,	Adjusted R-squared:  0.1223
F-statistic: 1.975 on 4 and 24 DF,  p-value: 0.1306

>
>
>
>
>
> ### LAD NT ###
> mQoL_LADNT.GJ = lm(qol.pos.log ~
+                           Group.ASDref*aprime.log
+                 #+ age
+                       + vabs_soc_ss
+                         , data = dLADNT.older18)
> summary(mQoL_LADNT.GJ)

Call:
lm(formula = qol.pos.log ~ Group.ASDref * aprime.log + vabs_soc_ss,
    data = dLADNT.older18)

Residuals:
     Min       1Q   Median       3Q      Max
-0.44130 -0.07671  0.02868  0.11354  0.22929

Coefficients:
                         Estimate Std. Error t value Pr(>|t|)
(Intercept)              3.277115   0.353835   9.262 4.63e-11 ***
Group.ASDref             0.129544   0.110031   1.177    0.247
aprime.log              -0.255894   0.419469  -0.610    0.546
vabs_soc_ss              0.005274   0.003296   1.600    0.118
Group.ASDref:aprime.log  1.240359   0.849041   1.461    0.153
---
Signif. codes:  0 ‘***’ 0.001 ‘**’ 0.01 ‘*’ 0.05 ‘.’ 0.1 ‘ ’ 1

Residual standard error: 0.1777 on 36 degrees of freedom
  (16 observations deleted due to missingness)
Multiple R-squared:  0.1001,	Adjusted R-squared:  0.0001017
F-statistic: 1.001 on 4 and 36 DF,  p-value: 0.4197

>
> mQoL_LADNT.Words =  lm(qol.pos.log ~
+                           Group.ASDref*FirstWords.months
+                 #+ age
+                       + vabs_soc_ss
+                         , data = dLADNT.older18)
> summary(mQoL_LADNT.Words)

Call:
lm(formula = qol.pos.log ~ Group.ASDref * FirstWords.months +
    vabs_soc_ss, data = dLADNT.older18)

Residuals:
     Min       1Q   Median       3Q      Max
-0.38898 -0.08318 -0.00498  0.14526  0.25439

Coefficients:
                                 Estimate Std. Error t value Pr(>|t|)
(Intercept)                     3.3856699  0.3832048   8.835 5.66e-10 ***
Group.ASDref                    0.0739282  0.1602245   0.461    0.648
FirstWords.months              -0.0003923  0.0049570  -0.079    0.937
vabs_soc_ss                     0.0044605  0.0035653   1.251    0.220
Group.ASDref:FirstWords.months -0.0035791  0.0099180  -0.361    0.721
---
Signif. codes:  0 ‘***’ 0.001 ‘**’ 0.01 ‘*’ 0.05 ‘.’ 0.1 ‘ ’ 1

Residual standard error: 0.1868 on 31 degrees of freedom
  (21 observations deleted due to missingness)
Multiple R-squared:  0.05333,	Adjusted R-squared:  -0.06883
F-statistic: 0.4366 on 4 and 31 DF,  p-value: 0.7812

>
> mQoL_LADNT.Phrases = lm(qol.pos.log ~
+                           Group.ASDref*FirstPhrases.months
+                 #+ age
+                       + vabs_soc_ss
+                         , data = dLADNT.older18)
> summary(mQoL_LADNT.Phrases)

Call:
lm(formula = qol.pos.log ~ Group.ASDref * FirstPhrases.months +
    vabs_soc_ss, data = dLADNT.older18)

Residuals:
     Min       1Q   Median       3Q      Max
-0.41015 -0.07646 -0.00342  0.15109  0.27783

Coefficients:
                                  Estimate Std. Error t value Pr(>|t|)
(Intercept)                       3.393604   0.367658   9.230 1.16e-10 ***
Group.ASDref                      0.119670   0.135576   0.883    0.384
FirstPhrases.months               0.002995   0.002860   1.047    0.303
vabs_soc_ss                       0.003565   0.003429   1.039    0.306
Group.ASDref:FirstPhrases.months -0.003537   0.005737  -0.616    0.542
---
Signif. codes:  0 ‘***’ 0.001 ‘**’ 0.01 ‘*’ 0.05 ‘.’ 0.1 ‘ ’ 1

Residual standard error: 0.1802 on 33 degrees of freedom
  (19 observations deleted due to missingness)
Multiple R-squared:  0.1223,	Adjusted R-squared:  0.01597
F-statistic:  1.15 on 4 and 33 DF,  p-value: 0.3505

>


> ###Follow up
>
> #aprime.log
> mQoL_ASD.GJ = lm(qol.pos.log ~ aprime.log
+                 #+ age
+                       + vabs_soc_ss
+                         , data = dASD.older18)
> summary(mQoL_ASD.GJ)

Call:
lm(formula = qol.pos.log ~ aprime.log + vabs_soc_ss, data = dASD.older18)

Residuals:
      8      12      17      19      20      21      23      24
-0.3062  0.2043  0.1545  0.2223 -0.5386 -0.2355  0.2761  0.2230

Coefficients:
            Estimate Std. Error t value Pr(>|t|)
(Intercept) 3.136444   0.911122   3.442   0.0184 *
aprime.log  0.265212   1.378599   0.192   0.8550
vabs_soc_ss 0.005923   0.010082   0.587   0.5824
---
Signif. codes:  0 ‘***’ 0.001 ‘**’ 0.01 ‘*’ 0.05 ‘.’ 0.1 ‘ ’ 1

Residual standard error: 0.3689 on 5 degrees of freedom
  (17 observations deleted due to missingness)
Multiple R-squared:  0.0646,	Adjusted R-squared:  -0.3096
F-statistic: 0.1726 on 2 and 5 DF,  p-value: 0.8463

>
> mQoL_NT.GJ = data = lm(qol.pos.log ~ aprime.log
+                 #+ age
+                       + vabs_soc_ss
+                         , dNT.older18)
> summary(mQoL_NT.GJ)

Call:
lm(formula = qol.pos.log ~ aprime.log + vabs_soc_ss, data = dNT.older18)

Residuals:
     Min       1Q   Median       3Q      Max
-0.41249 -0.06869  0.00101  0.12148  0.22054

Coefficients:
            Estimate Std. Error t value Pr(>|t|)
(Intercept) 3.109553   0.509400   6.104  7.2e-06 ***
aprime.log  0.387269   0.507970   0.762    0.455
vabs_soc_ss 0.007512   0.004859   1.546    0.139
---
Signif. codes:  0 ‘***’ 0.001 ‘**’ 0.01 ‘*’ 0.05 ‘.’ 0.1 ‘ ’ 1

Residual standard error: 0.1692 on 19 degrees of freedom
  (5 observations deleted due to missingness)
Multiple R-squared:  0.1271,	Adjusted R-squared:  0.03522
F-statistic: 1.383 on 2 and 19 DF,  p-value: 0.2749

>
> mQoL_LAD.GJ = lm(qol.pos.log ~ aprime.log
+                 #+ age
+                       + vabs_soc_ss
+                         , data = dLAD.older18)
> summary(mQoL_LAD.GJ)

Call:
lm(formula = qol.pos.log ~ aprime.log + vabs_soc_ss, data = dLAD.older18)

Residuals:
     Min       1Q   Median       3Q      Max
-0.44520 -0.08552  0.08274  0.11091  0.23831

Coefficients:
             Estimate Std. Error t value Pr(>|t|)
(Intercept)  3.386170   0.509258   6.649 5.59e-06 ***
aprime.log  -0.814753   0.712688  -1.143     0.27
vabs_soc_ss  0.003669   0.004635   0.792     0.44
---
Signif. codes:  0 ‘***’ 0.001 ‘**’ 0.01 ‘*’ 0.05 ‘.’ 0.1 ‘ ’ 1

Residual standard error: 0.1907 on 16 degrees of freedom
  (11 observations deleted due to missingness)
Multiple R-squared:  0.08994,	Adjusted R-squared:  -0.02382
F-statistic: 0.7906 on 2 and 16 DF,  p-value: 0.4705

>
> #Words
> mQoL_ASD.Words = lm(qol.pos.log ~ FirstWords.months
+                 #+ age
+                       + vabs_soc_ss
+                         , data = dASD.older18)
> summary(mQoL_ASD.Words)

Call:
lm(formula = qol.pos.log ~ FirstWords.months + vabs_soc_ss, data = dASD.older18)

Residuals:
       8       12       17       19       20       21
-0.20312  0.37500  0.10955  0.06219 -0.05926 -0.28435

Coefficients:
                  Estimate Std. Error t value Pr(>|t|)
(Intercept)        1.38919    1.30256   1.067    0.364
FirstWords.months  0.03410    0.02113   1.614    0.205
vabs_soc_ss        0.01802    0.01205   1.496    0.232

Residual standard error: 0.3067 on 3 degrees of freedom
  (19 observations deleted due to missingness)
Multiple R-squared:  0.5482,	Adjusted R-squared:  0.2469
F-statistic:  1.82 on 2 and 3 DF,  p-value: 0.3037

>
> mQoL_NT.Words = lm(qol.pos.log ~ FirstWords.months
+                 #+ age
+                       + vabs_soc_ss
+                         , data = dNT.older18)
> summary(mQoL_NT.Words)

Call:
lm(formula = qol.pos.log ~ FirstWords.months + vabs_soc_ss, data = dNT.older18)

Residuals:
     Min       1Q   Median       3Q      Max
-0.37403 -0.10476 -0.00275  0.11779  0.22465

Coefficients:
                   Estimate Std. Error t value Pr(>|t|)
(Intercept)        3.015269   0.512261   5.886  1.8e-05 ***
FirstWords.months -0.002291   0.007692  -0.298   0.7694
vabs_soc_ss        0.008371   0.004807   1.742   0.0997 .
---
Signif. codes:  0 ‘***’ 0.001 ‘**’ 0.01 ‘*’ 0.05 ‘.’ 0.1 ‘ ’ 1

Residual standard error: 0.1645 on 17 degrees of freedom
  (7 observations deleted due to missingness)
Multiple R-squared:  0.1544,	Adjusted R-squared:  0.05493
F-statistic: 1.552 on 2 and 17 DF,  p-value: 0.2403

>
> mQoL_LAD.Words = lm(qol.pos.log ~ FirstWords.months
+                 #+ age
+                       + vabs_soc_ss
+                         , data = dNT.older18)
> summary(mQoL_LAD.Words)

Call:
lm(formula = qol.pos.log ~ FirstWords.months + vabs_soc_ss, data = dNT.older18)

Residuals:
     Min       1Q   Median       3Q      Max
-0.37403 -0.10476 -0.00275  0.11779  0.22465

Coefficients:
                   Estimate Std. Error t value Pr(>|t|)
(Intercept)        3.015269   0.512261   5.886  1.8e-05 ***
FirstWords.months -0.002291   0.007692  -0.298   0.7694
vabs_soc_ss        0.008371   0.004807   1.742   0.0997 .
---
Signif. codes:  0 ‘***’ 0.001 ‘**’ 0.01 ‘*’ 0.05 ‘.’ 0.1 ‘ ’ 1

Residual standard error: 0.1645 on 17 degrees of freedom
  (7 observations deleted due to missingness)
Multiple R-squared:  0.1544,	Adjusted R-squared:  0.05493
F-statistic: 1.552 on 2 and 17 DF,  p-value: 0.2403

>
> #Phrases
> mQoL_ASD.Phrases = lm(qol.pos.log ~ FirstPhrases.months
+                 #+ age
+                       + vabs_soc_ss
+                         , data = dASD.older18)
> summary(mQoL_ASD.Phrases)

Call:
lm(formula = qol.pos.log ~ FirstPhrases.months + vabs_soc_ss,
    data = dASD.older18)

Residuals:
      8      12      17      19      20      21      24
-0.3078  0.2716  0.2186  0.3036 -0.4980 -0.1640  0.1761

Coefficients:
                    Estimate Std. Error t value Pr(>|t|)
(Intercept)         3.189799   1.141084   2.795    0.049 *
FirstPhrases.months 0.001592   0.013474   0.118    0.912
vabs_soc_ss         0.003932   0.010425   0.377    0.725
---
Signif. codes:  0 ‘***’ 0.001 ‘**’ 0.01 ‘*’ 0.05 ‘.’ 0.1 ‘ ’ 1

Residual standard error: 0.3919 on 4 degrees of freedom
  (18 observations deleted due to missingness)
Multiple R-squared:  0.03517,	Adjusted R-squared:  -0.4472
F-statistic: 0.07291 on 2 and 4 DF,  p-value: 0.9309

>
> mQoL_NT.Phrases = lm(qol.pos.log ~ FirstPhrases.months
+                 #+ age
+                       + vabs_soc_ss
+                         , data = dNT.older18)
> summary(mQoL_NT.Phrases)

Call:
lm(formula = qol.pos.log ~ FirstPhrases.months + vabs_soc_ss,
    data = dNT.older18)

Residuals:
     Min       1Q   Median       3Q      Max
-0.39752 -0.07319  0.02571  0.13268  0.23034

Coefficients:
                    Estimate Std. Error t value Pr(>|t|)
(Intercept)         3.049442   0.536559   5.683 1.77e-05 ***
FirstPhrases.months 0.001645   0.004856   0.339    0.739
vabs_soc_ss         0.007336   0.004926   1.489    0.153
---
Signif. codes:  0 ‘***’ 0.001 ‘**’ 0.01 ‘*’ 0.05 ‘.’ 0.1 ‘ ’ 1

Residual standard error: 0.1713 on 19 degrees of freedom
  (5 observations deleted due to missingness)
Multiple R-squared:  0.1058,	Adjusted R-squared:  0.01167
F-statistic: 1.124 on 2 and 19 DF,  p-value: 0.3457

>
> mQoL_LAD.Phrases = lm(qol.pos.log ~ FirstPhrases.months
+                 #+ age
+                       + vabs_soc_ss
+                         , data = dLAD.older18)
> summary(mQoL_LAD.Phrases)

Call:
lm(formula = qol.pos.log ~ FirstPhrases.months + vabs_soc_ss,
    data = dLAD.older18)

Residuals:
     Min       1Q   Median       3Q      Max
-0.31574 -0.11548 -0.01572  0.15831  0.30084

Coefficients:
                    Estimate Std. Error t value Pr(>|t|)
(Intercept)         3.638765   0.516003   7.052 8.65e-06 ***
FirstPhrases.months 0.004877   0.002817   1.731    0.107
vabs_soc_ss         0.000622   0.004895   0.127    0.901
---
Signif. codes:  0 ‘***’ 0.001 ‘**’ 0.01 ‘*’ 0.05 ‘.’ 0.1 ‘ ’ 1

Residual standard error: 0.1927 on 13 degrees of freedom
  (14 observations deleted due to missingness)
Multiple R-squared:  0.1904,	Adjusted R-squared:  0.06582
F-statistic: 1.528 on 2 and 13 DF,  p-value: 0.2534


> #############################
> ##### Life satisfaction #####
> #############################
>
> #lifesat.log
>
> ### ASD LAD ###
> mLifeSat_ASDLAD.GJ = lm(lifesat.log ~
+                           Group.NTref*aprime.log
+                  #+ age
+                       + vabs_soc_ss
+                         , data = dASDLAD.older18)
> summary(mLifeSat_ASDLAD.GJ)

Call:
lm(formula = lifesat.log ~ Group.NTref * aprime.log + vabs_soc_ss,
    data = dASDLAD.older18)

Residuals:
     Min       1Q   Median       3Q      Max
-0.77014 -0.11690 -0.01132  0.21681  0.43829

Coefficients:
                        Estimate Std. Error t value Pr(>|t|)
(Intercept)             1.906269   0.542384   3.515  0.00195 **
Group.NTref             0.313343   0.282023   1.111  0.27854
aprime.log             -1.455508   0.751716  -1.936  0.06579 .
vabs_soc_ss             0.008825   0.005417   1.629  0.11751
Group.NTref:aprime.log -0.834831   1.572634  -0.531  0.60084
---
Signif. codes:  0 ‘***’ 0.001 ‘**’ 0.01 ‘*’ 0.05 ‘.’ 0.1 ‘ ’ 1

Residual standard error: 0.2982 on 22 degrees of freedom
  (28 observations deleted due to missingness)
Multiple R-squared:  0.4326,	Adjusted R-squared:  0.3295
F-statistic: 4.194 on 4 and 22 DF,  p-value: 0.0113

>
> mLifeSat_ASDLAD.Words = lm(lifesat.log ~
+                           Group.NTref*FirstWords.months
+                  #+ age
+                       + vabs_soc_ss
+                         , data = dASDLAD.older18)
> summary(mLifeSat_ASDLAD.Words)

Call:
lm(formula = lifesat.log ~ Group.NTref * FirstWords.months +
    vabs_soc_ss, data = dASDLAD.older18)

Residuals:
     Min       1Q   Median       3Q      Max
-0.75566 -0.13087  0.06009  0.20127  0.44864

Coefficients:
                               Estimate Std. Error t value Pr(>|t|)
(Intercept)                    1.949165   0.803613   2.426   0.0267 *
Group.NTref                    0.472363   0.375951   1.256   0.2259
FirstWords.months              0.011080   0.012355   0.897   0.3823
vabs_soc_ss                    0.008629   0.007388   1.168   0.2589
Group.NTref:FirstWords.months -0.013876   0.024565  -0.565   0.5795
---
Signif. codes:  0 ‘***’ 0.001 ‘**’ 0.01 ‘*’ 0.05 ‘.’ 0.1 ‘ ’ 1

Residual standard error: 0.3482 on 17 degrees of freedom
  (33 observations deleted due to missingness)
Multiple R-squared:  0.2952,	Adjusted R-squared:  0.1294
F-statistic:  1.78 on 4 and 17 DF,  p-value: 0.1793

>
> mLifeSat_ASDLAD.Phrases = lm(lifesat.log ~
+                           Group.NTref*FirstPhrases.months
+                  #+ age
+                       + vabs_soc_ss
+                         , data = dASDLAD.older18)
> summary(mLifeSat_ASDLAD.Phrases)

Call:
lm(formula = lifesat.log ~ Group.NTref * FirstPhrases.months +
    vabs_soc_ss, data = dASDLAD.older18)

Residuals:
     Min       1Q   Median       3Q      Max
-0.80832 -0.14336  0.02323  0.24440  0.47237

Coefficients:
                                Estimate Std. Error t value Pr(>|t|)
(Intercept)                     1.998654   0.662243   3.018  0.00739 **
Group.NTref                     0.243575   0.421925   0.577  0.57089
FirstPhrases.months             0.003799   0.006210   0.612  0.54831
vabs_soc_ss                     0.008424   0.006154   1.369  0.18789
Group.NTref:FirstPhrases.months 0.003489   0.012467   0.280  0.78278
---
Signif. codes:  0 ‘***’ 0.001 ‘**’ 0.01 ‘*’ 0.05 ‘.’ 0.1 ‘ ’ 1

Residual standard error: 0.335 on 18 degrees of freedom
  (32 observations deleted due to missingness)
Multiple R-squared:  0.3853,	Adjusted R-squared:  0.2486
F-statistic:  2.82 on 4 and 18 DF,  p-value: 0.05601

>
>
>
>
>
> ### ASD NT ###
> mLifeSat_ASDNT.GJ = lm(lifesat.log ~
+                           Group.LADref*aprime.log
+                 #+ age
+                       + vabs_soc_ss
+                         , data = dASDNT.older18)
> summary(mLifeSat_ASDNT.GJ)

Call:
lm(formula = lifesat.log ~ Group.LADref * aprime.log + vabs_soc_ss,
    data = dASDNT.older18)

Residuals:
     Min       1Q   Median       3Q      Max
-0.73732 -0.04813  0.06424  0.17179  0.43566

Coefficients:
                         Estimate Std. Error t value Pr(>|t|)
(Intercept)              1.331843   0.558731   2.384  0.02505 *
Group.LADref             0.401686   0.265580   1.512  0.14295
aprime.log              -0.246753   0.687312  -0.359  0.72260
vabs_soc_ss              0.015993   0.005735   2.789  0.00997 **
Group.LADref:aprime.log  0.902238   1.351107   0.668  0.51039
---
Signif. codes:  0 ‘***’ 0.001 ‘**’ 0.01 ‘*’ 0.05 ‘.’ 0.1 ‘ ’ 1

Residual standard error: 0.2897 on 25 degrees of freedom
  (22 observations deleted due to missingness)
Multiple R-squared:  0.4627,	Adjusted R-squared:  0.3767
F-statistic: 5.382 on 4 and 25 DF,  p-value: 0.002878

>
> mLifeSat_ASDNT.Words = lm(lifesat.log ~
+                           Group.LADref*FirstWords.months
+                 #+ age
+                       + vabs_soc_ss
+                         , data = dASDNT.older18)
> summary(mLifeSat_ASDNT.Words)

Call:
lm(formula = lifesat.log ~ Group.LADref * FirstWords.months +
    vabs_soc_ss, data = dASDNT.older18)

Residuals:
     Min       1Q   Median       3Q      Max
-0.79918 -0.08071  0.02645  0.14488  0.34731

Coefficients:
                                Estimate Std. Error t value Pr(>|t|)
(Intercept)                     0.758232   0.698078   1.086  0.28971
Group.LADref                    0.756262   0.314709   2.403  0.02558 *
FirstWords.months               0.007193   0.011219   0.641  0.52836
vabs_soc_ss                     0.021265   0.006488   3.278  0.00359 **
Group.LADref:FirstWords.months -0.036377   0.022500  -1.617  0.12085
---
Signif. codes:  0 ‘***’ 0.001 ‘**’ 0.01 ‘*’ 0.05 ‘.’ 0.1 ‘ ’ 1

Residual standard error: 0.2766 on 21 degrees of freedom
  (26 observations deleted due to missingness)
Multiple R-squared:  0.5204,	Adjusted R-squared:  0.4291
F-statistic: 5.697 on 4 and 21 DF,  p-value: 0.002882

>
> mLifeSat_ASDNT.Phrases = lm(lifesat.log ~
+                           Group.NTref*FirstPhrases.months
+                  #+ age
+                       + vabs_soc_ss
+                         , data = dASDNT.older18)
> summary(mLifeSat_ASDNT.Phrases)

Call:
lm(formula = lifesat.log ~ Group.NTref * FirstPhrases.months +
    vabs_soc_ss, data = dASDNT.older18)

Residuals:
     Min       1Q   Median       3Q      Max
-0.74000 -0.02161  0.02852  0.13700  0.49363

Coefficients:
                                 Estimate Std. Error t value Pr(>|t|)
(Intercept)                      1.623047   0.630347   2.575  0.01662 *
Group.NTref                      1.044066   0.755044   1.383  0.17946
FirstPhrases.months             -0.007059   0.008215  -0.859  0.39868
vabs_soc_ss                      0.016152   0.005678   2.845  0.00895 **
Group.NTref:FirstPhrases.months -0.021359   0.025678  -0.832  0.41373
---
Signif. codes:  0 ‘***’ 0.001 ‘**’ 0.01 ‘*’ 0.05 ‘.’ 0.1 ‘ ’ 1

Residual standard error: 0.2908 on 24 degrees of freedom
  (23 observations deleted due to missingness)
Multiple R-squared:  0.4803,	Adjusted R-squared:  0.3937
F-statistic: 5.546 on 4 and 24 DF,  p-value: 0.002623

>
>
>
>
> ### LAD NT  ###
> mLifeSat_LADNT.GJ = lm(lifesat.log ~
+                           Group.ASDref*aprime.log
+                 #+ age
+                       + vabs_soc_ss
+                         , data = dLADNT.older18)
> summary(mLifeSat_LADNT.GJ)

Call:
lm(formula = lifesat.log ~ Group.ASDref * aprime.log + vabs_soc_ss,
    data = dLADNT.older18)

Residuals:
     Min       1Q   Median       3Q      Max
-0.64122 -0.13481  0.06048  0.17850  0.26888

Coefficients:
                         Estimate Std. Error t value Pr(>|t|)
(Intercept)              2.838368   0.477525   5.944 8.26e-07 ***
Group.ASDref             0.184904   0.148495   1.245    0.221
aprime.log              -0.782967   0.566102  -1.383    0.175
vabs_soc_ss              0.002521   0.004449   0.567    0.574
Group.ASDref:aprime.log  1.698049   1.145839   1.482    0.147
---
Signif. codes:  0 ‘***’ 0.001 ‘**’ 0.01 ‘*’ 0.05 ‘.’ 0.1 ‘ ’ 1

Residual standard error: 0.2398 on 36 degrees of freedom
  (16 observations deleted due to missingness)
Multiple R-squared:  0.08785,	Adjusted R-squared:  -0.0135
F-statistic: 0.8668 on 4 and 36 DF,  p-value: 0.4932

>
> mLifeSat_LADNT.Words = lm(lifesat.log ~
+                           Group.ASDref*FirstWords.months
+                 #+ age
+                       + vabs_soc_ss
+                         , data = dLADNT.older18)
> summary(mLifeSat_LADNT.Words)

Call:
lm(formula = lifesat.log ~ Group.ASDref * FirstWords.months +
    vabs_soc_ss, data = dLADNT.older18)

Residuals:
     Min       1Q   Median       3Q      Max
-0.66275 -0.09278  0.03768  0.17927  0.32198

Coefficients:
                                Estimate Std. Error t value Pr(>|t|)
(Intercept)                     2.921478   0.520964   5.608 3.76e-06 ***
Group.ASDref                    0.256851   0.217824   1.179    0.247
FirstWords.months              -0.003331   0.006739  -0.494    0.625
vabs_soc_ss                     0.002898   0.004847   0.598    0.554
Group.ASDref:FirstWords.months -0.014301   0.013483  -1.061    0.297
---
Signif. codes:  0 ‘***’ 0.001 ‘**’ 0.01 ‘*’ 0.05 ‘.’ 0.1 ‘ ’ 1

Residual standard error: 0.2539 on 31 degrees of freedom
  (21 observations deleted due to missingness)
Multiple R-squared:  0.05212,	Adjusted R-squared:  -0.07019
F-statistic: 0.4261 on 4 and 31 DF,  p-value: 0.7886

>
> mLifeSat_LADNT.Phrases = lm(lifesat.log ~
+                           Group.ASDref*FirstPhrases.months
+                 #+ age
+                       + vabs_soc_ss
+                         , data = dLADNT.older18)
> summary(mLifeSat_LADNT.Phrases)

Call:
lm(formula = lifesat.log ~ Group.ASDref * FirstPhrases.months +
    vabs_soc_ss, data = dLADNT.older18)

Residuals:
     Min       1Q   Median       3Q      Max
-0.63252 -0.15068  0.04155  0.13869  0.36635

Coefficients:
                                   Estimate Std. Error t value Pr(>|t|)
(Intercept)                       3.1190637  0.4981208   6.262 4.48e-07 ***
Group.ASDref                      0.3579295  0.1836847   1.949   0.0599 .
FirstPhrases.months              -0.0014722  0.0038745  -0.380   0.7064
vabs_soc_ss                       0.0005152  0.0046464   0.111   0.9124
Group.ASDref:FirstPhrases.months -0.0146409  0.0077728  -1.884   0.0685 .
---
Signif. codes:  0 ‘***’ 0.001 ‘**’ 0.01 ‘*’ 0.05 ‘.’ 0.1 ‘ ’ 1

Residual standard error: 0.2441 on 33 degrees of freedom
  (19 observations deleted due to missingness)
Multiple R-squared:  0.1197,	Adjusted R-squared:  0.01299
F-statistic: 1.122 on 4 and 33 DF,  p-value: 0.363


> ###Follow up
> #aprime.log
> mLifeSat_ASD.GJ = lm(lifesat.log ~ aprime.log
+                 #+ age
+                       + vabs_soc_ss
+                         , data = dASD.older18)
> summary(mLifeSat_ASD.GJ)

Call:
lm(formula = lifesat.log ~ aprime.log + vabs_soc_ss, data = dASD.older18)

Residuals:
       8       12       17       19       20       21       23       24
-0.08090  0.11411  0.43327  0.17803 -0.61767 -0.29339  0.07076  0.19578

Coefficients:
            Estimate Std. Error t value Pr(>|t|)
(Intercept)  0.56963    0.95658   0.595   0.5774
aprime.log  -0.38913    1.44738  -0.269   0.7988
vabs_soc_ss  0.02250    0.01058   2.126   0.0869 .
---
Signif. codes:  0 ‘***’ 0.001 ‘**’ 0.01 ‘*’ 0.05 ‘.’ 0.1 ‘ ’ 1

Residual standard error: 0.3873 on 5 degrees of freedom
  (17 observations deleted due to missingness)
Multiple R-squared:  0.5314,	Adjusted R-squared:  0.344
F-statistic: 2.835 on 2 and 5 DF,  p-value: 0.1503

>
> mLifeSat_NT.GJ = lm(lifesat.log ~ aprime.log
+                 #+ age
+                       + vabs_soc_ss
+                         , data = dNT.older18)
> summary(mLifeSat_NT.GJ)

Call:
lm(formula = lifesat.log ~ aprime.log + vabs_soc_ss, data = dNT.older18)

Residuals:
     Min       1Q   Median       3Q      Max
-0.68612 -0.04600  0.05644  0.16539  0.25224

Coefficients:
            Estimate Std. Error t value Pr(>|t|)
(Intercept) 2.277700   0.765586   2.975  0.00778 **
aprime.log  0.130667   0.763438   0.171  0.86591
vabs_soc_ss 0.008815   0.007303   1.207  0.24225
---
Signif. codes:  0 ‘***’ 0.001 ‘**’ 0.01 ‘*’ 0.05 ‘.’ 0.1 ‘ ’ 1

Residual standard error: 0.2544 on 19 degrees of freedom
  (5 observations deleted due to missingness)
Multiple R-squared:  0.07134,	Adjusted R-squared:  -0.02641
F-statistic: 0.7298 on 2 and 19 DF,  p-value: 0.495

>
> mLifeSat_LAD.GJ = lm(lifesat.log ~ aprime.log
+                 #+ age
+                       + vabs_soc_ss
+                         , data = dLAD.older18)
> summary(mLifeSat_LAD.GJ)

Call:
lm(formula = lifesat.log ~ aprime.log + vabs_soc_ss, data = dLAD.older18)

Residuals:
     Min       1Q   Median       3Q      Max
-0.52627 -0.07758  0.01043  0.16321  0.28005

Coefficients:
             Estimate Std. Error t value Pr(>|t|)
(Intercept)  3.234565   0.581372   5.564 4.28e-05 ***
aprime.log  -1.459611   0.813609  -1.794   0.0917 .
vabs_soc_ss -0.001989   0.005292  -0.376   0.7120
---
Signif. codes:  0 ‘***’ 0.001 ‘**’ 0.01 ‘*’ 0.05 ‘.’ 0.1 ‘ ’ 1

Residual standard error: 0.2177 on 16 degrees of freedom
  (11 observations deleted due to missingness)
Multiple R-squared:  0.1975,	Adjusted R-squared:  0.09722
F-statistic: 1.969 on 2 and 16 DF,  p-value: 0.172

>
> #Words
> mLifeSat_ASD.Words = lm(lifesat.log ~ FirstWords.months
+                 #+ age
+                       + vabs_soc_ss
+                         , data = dASD.older18)
> summary(mLifeSat_ASD.Words)

Call:
lm(formula = lifesat.log ~ FirstWords.months + vabs_soc_ss, data = dASD.older18)

Residuals:
       8       12       17       19       20       21
 0.05937  0.20903  0.35922 -0.10479 -0.13360 -0.38922

Coefficients:
                  Estimate Std. Error t value Pr(>|t|)
(Intercept)       -1.43904    1.46440  -0.983   0.3982
FirstWords.months  0.03543    0.02375   1.492   0.2326
vabs_soc_ss        0.03851    0.01355   2.843   0.0655 .
---
Signif. codes:  0 ‘***’ 0.001 ‘**’ 0.01 ‘*’ 0.05 ‘.’ 0.1 ‘ ’ 1

Residual standard error: 0.3448 on 3 degrees of freedom
  (19 observations deleted due to missingness)
Multiple R-squared:  0.7372,	Adjusted R-squared:  0.5621
F-statistic: 4.209 on 2 and 3 DF,  p-value: 0.1347

>
> mLifeSat_NT.Words = lm(lifesat.log ~ FirstWords.months
+                 #+ age
+                       + vabs_soc_ss
+                         , data = dNT.older18)
> summary(mLifeSat_NT.Words)

Call:
lm(formula = lifesat.log ~ FirstWords.months + vabs_soc_ss, data = dNT.older18)

Residuals:
     Min       1Q   Median       3Q      Max
-0.72831 -0.03317  0.02045  0.10220  0.28683

Coefficients:
                   Estimate Std. Error t value Pr(>|t|)
(Intercept)        2.130361   0.736915   2.891   0.0102 *
FirstWords.months -0.010728   0.011066  -0.969   0.3459
vabs_soc_ss        0.011724   0.006915   1.696   0.1082
---
Signif. codes:  0 ‘***’ 0.001 ‘**’ 0.01 ‘*’ 0.05 ‘.’ 0.1 ‘ ’ 1

Residual standard error: 0.2366 on 17 degrees of freedom
  (7 observations deleted due to missingness)
Multiple R-squared:  0.1811,	Adjusted R-squared:  0.08471
F-statistic: 1.879 on 2 and 17 DF,  p-value: 0.1831

>
> mLifeSat_LAD.Words = lm(lifesat.log ~ FirstWords.months
+                 #+ age
+                       + vabs_soc_ss
+                         , data = dLAD.older18)
> summary(mLifeSat_LAD.Words)

Call:
lm(formula = lifesat.log ~ FirstWords.months + vabs_soc_ss, data = dLAD.older18)

Residuals:
     Min       1Q   Median       3Q      Max
-0.61399 -0.07921  0.03621  0.17036  0.35066

Coefficients:
                   Estimate Std. Error t value Pr(>|t|)
(Intercept)        3.489167   0.713411   4.891 0.000295 ***
FirstWords.months  0.003450   0.006571   0.525 0.608394
vabs_soc_ss       -0.003667   0.006589  -0.557 0.587309
---
Signif. codes:  0 ‘***’ 0.001 ‘**’ 0.01 ‘*’ 0.05 ‘.’ 0.1 ‘ ’ 1

Residual standard error: 0.2614 on 13 degrees of freedom
  (14 observations deleted due to missingness)
Multiple R-squared:  0.04555,	Adjusted R-squared:  -0.1013
F-statistic: 0.3102 on 2 and 13 DF,  p-value: 0.7386

>
> #Phrases
> mLifeSat_ASD.Phrases = lm(lifesat.log ~ FirstPhrases.months
+                 #+ age
+                       + vabs_soc_ss
+                         , data = dASD.older18)
> summary(mLifeSat_ASD.Phrases)

Call:
lm(formula = lifesat.log ~ FirstPhrases.months + vabs_soc_ss,
    data = dASD.older18)

Residuals:
       8       12       17       19       20       21       24
-0.10557  0.08983  0.51334  0.15042 -0.57697 -0.23497  0.16392

Coefficients:
                    Estimate Std. Error t value Pr(>|t|)
(Intercept)         0.381799   1.235522   0.309    0.773
FirstPhrases.months 0.005071   0.014589   0.348    0.746
vabs_soc_ss         0.023315   0.011288   2.065    0.108

Residual standard error: 0.4244 on 4 degrees of freedom
  (18 observations deleted due to missingness)
Multiple R-squared:  0.5161,	Adjusted R-squared:  0.2742
F-statistic: 2.133 on 2 and 4 DF,  p-value: 0.2341

>
> mLifeSat_NT.Phrases = lm(lifesat.log ~ FirstPhrases.months
+                 #+ age
+                       + vabs_soc_ss
+                         , data = dNT.older18)
> summary(mLifeSat_NT.Phrases)

Call:
lm(formula = lifesat.log ~ FirstPhrases.months + vabs_soc_ss,
    data = dNT.older18)

Residuals:
     Min       1Q   Median       3Q      Max
-0.68246 -0.04925  0.03653  0.13243  0.33368

Coefficients:
                     Estimate Std. Error t value Pr(>|t|)
(Intercept)          2.519742   0.771219   3.267  0.00405 **
FirstPhrases.months -0.007987   0.006980  -1.144  0.26668
vabs_soc_ss          0.007781   0.007080   1.099  0.28549
---
Signif. codes:  0 ‘***’ 0.001 ‘**’ 0.01 ‘*’ 0.05 ‘.’ 0.1 ‘ ’ 1

Residual standard error: 0.2462 on 19 degrees of freedom
  (5 observations deleted due to missingness)
Multiple R-squared:  0.1299,	Adjusted R-squared:  0.03829
F-statistic: 1.418 on 2 and 19 DF,  p-value: 0.2667

>
> mLifeSat_LAD.Phrases = lm(lifesat.log ~ FirstPhrases.months
+                 #+ age
+                       + vabs_soc_ss
+                         , data = dLAD.older18)
> summary(mLifeSat_LAD.Phrases)

Call:
lm(formula = lifesat.log ~ FirstPhrases.months + vabs_soc_ss,
    data = dLAD.older18)

Residuals:
     Min       1Q   Median       3Q      Max
-0.50499 -0.10272  0.01766  0.06671  0.39061

Coefficients:
                     Estimate Std. Error t value Pr(>|t|)
(Intercept)          3.527665   0.621501   5.676 7.59e-05 ***
FirstPhrases.months  0.006066   0.003392   1.788   0.0971 .
vabs_soc_ss         -0.005154   0.005896  -0.874   0.3979
---
Signif. codes:  0 ‘***’ 0.001 ‘**’ 0.01 ‘*’ 0.05 ‘.’ 0.1 ‘ ’ 1

Residual standard error: 0.2321 on 13 degrees of freedom
  (14 observations deleted due to missingness)
Multiple R-squared:  0.2248,	Adjusted R-squared:  0.1055
F-statistic: 1.885 on 2 and 13 DF,  p-value: 0.1911


> ##########################
> ##### Social support #####
> ##########################
>
> ### ASD LAD ###
> mSupp_ASDLAD.GJ = lm(support.log ~
+                           Group.NTref*aprime.log
+                  #+ age
+                       + vabs_soc_ss
+                         , data = dASDLAD.older18)
> summary(mSupp_ASDLAD.GJ)

Call:
lm(formula = support.log ~ Group.NTref * aprime.log + vabs_soc_ss,
    data = dASDLAD.older18)

Residuals:
     Min       1Q   Median       3Q      Max
-0.41998 -0.08557 -0.00319  0.10101  0.27626

Coefficients:
                       Estimate Std. Error t value Pr(>|t|)
(Intercept)            3.590782   0.257924  13.922 1.46e-13 ***
Group.NTref            0.153226   0.128058   1.197    0.242
aprime.log             0.002952   0.348561   0.008    0.993
vabs_soc_ss            0.002685   0.002502   1.073    0.293
Group.NTref:aprime.log 0.116176   0.698674   0.166    0.869
---
Signif. codes:  0 ‘***’ 0.001 ‘**’ 0.01 ‘*’ 0.05 ‘.’ 0.1 ‘ ’ 1

Residual standard error: 0.1563 on 26 degrees of freedom
  (24 observations deleted due to missingness)
Multiple R-squared:  0.2623,	Adjusted R-squared:  0.1488
F-statistic: 2.311 on 4 and 26 DF,  p-value: 0.08449

>
> mSupp_ASDLAD.Words = lm(support.log ~
+                           Group.NTref*FirstWords.months
+                  #+ age
+                       + vabs_soc_ss
+                         , data = dASDLAD.older18)
> summary(mSupp_ASDLAD.Words)

Call:
lm(formula = support.log ~ Group.NTref * FirstWords.months +
    vabs_soc_ss, data = dASDLAD.older18)

Residuals:
     Min       1Q   Median       3Q      Max
-0.38853 -0.06171  0.00252  0.04530  0.28274

Coefficients:
                                Estimate Std. Error t value Pr(>|t|)
(Intercept)                    3.7542389  0.3593976  10.446 2.59e-09 ***
Group.NTref                    0.0423657  0.1489560   0.284    0.779
FirstWords.months             -0.0005349  0.0043621  -0.123    0.904
vabs_soc_ss                    0.0009129  0.0032436   0.281    0.781
Group.NTref:FirstWords.months  0.0062222  0.0085833   0.725    0.477
---
Signif. codes:  0 ‘***’ 0.001 ‘**’ 0.01 ‘*’ 0.05 ‘.’ 0.1 ‘ ’ 1

Residual standard error: 0.157 on 19 degrees of freedom
  (31 observations deleted due to missingness)
Multiple R-squared:  0.2492,	Adjusted R-squared:  0.09111
F-statistic: 1.576 on 4 and 19 DF,  p-value: 0.2212

>
> mSupp_ASDLAD.Phrases = lm(support.log ~
+                           Group.NTref*FirstPhrases.months
+                  #+ age
+                       + vabs_soc_ss
+                         , data = dASDLAD.older18)
> summary(mSupp_ASDLAD.Phrases)

Call:
lm(formula = support.log ~ Group.NTref * FirstPhrases.months +
    vabs_soc_ss, data = dASDLAD.older18)

Residuals:
     Min       1Q   Median       3Q      Max
-0.38933 -0.07282 -0.00356  0.06685  0.30725

Coefficients:
                                 Estimate Std. Error t value Pr(>|t|)
(Intercept)                     3.5513470  0.2942075  12.071 1.23e-10 ***
Group.NTref                     0.0954405  0.1989719   0.480    0.637
FirstPhrases.months             0.0008669  0.0029142   0.297    0.769
vabs_soc_ss                     0.0026706  0.0027152   0.984    0.337
Group.NTref:FirstPhrases.months 0.0023062  0.0058538   0.394    0.698
---
Signif. codes:  0 ‘***’ 0.001 ‘**’ 0.01 ‘*’ 0.05 ‘.’ 0.1 ‘ ’ 1

Residual standard error: 0.158 on 20 degrees of freedom
  (30 observations deleted due to missingness)
Multiple R-squared:  0.3482,	Adjusted R-squared:  0.2179
F-statistic: 2.672 on 4 and 20 DF,  p-value: 0.06199

>
>
>
>
>
> ### ASD NT ###
> mSupp_ASDNT.GJ = lm(support.log ~
+                           Group.LADref*aprime.log
+                 #+ age
+                       + vabs_soc_ss
+                         , data = dASDNT.older18)
> summary(mSupp_ASDNT.GJ)

Call:
lm(formula = support.log ~ Group.LADref * aprime.log + vabs_soc_ss,
    data = dASDNT.older18)

Residuals:
     Min       1Q   Median       3Q      Max
-0.48784 -0.07227  0.02676  0.14473  0.24757

Coefficients:
                         Estimate Std. Error t value Pr(>|t|)
(Intercept)              3.371613   0.347287   9.708 1.29e-10 ***
Group.LADref             0.082863   0.161896   0.512    0.613
aprime.log              -0.055748   0.399138  -0.140    0.890
vabs_soc_ss              0.004679   0.003437   1.361    0.184
Group.LADref:aprime.log -0.010624   0.797854  -0.013    0.989
---
Signif. codes:  0 ‘***’ 0.001 ‘**’ 0.01 ‘*’ 0.05 ‘.’ 0.1 ‘ ’ 1

Residual standard error: 0.2026 on 29 degrees of freedom
  (18 observations deleted due to missingness)
Multiple R-squared:  0.1553,	Adjusted R-squared:  0.03873
F-statistic: 1.332 on 4 and 29 DF,  p-value: 0.2816

>
> mSupp_ASDNT.Words = lm(support.log ~
+                           Group.LADref*FirstWords.months
+                 #+ age
+                       + vabs_soc_ss
+                         , data = dASDNT.older18)
> summary(mSupp_ASDNT.Words)

Call:
lm(formula = support.log ~ Group.LADref * FirstWords.months +
    vabs_soc_ss, data = dASDNT.older18)

Residuals:
     Min       1Q   Median       3Q      Max
-0.44322 -0.06106  0.02402  0.13430  0.26363

Coefficients:
                                Estimate Std. Error t value Pr(>|t|)
(Intercept)                     3.469373   0.536450   6.467 1.66e-06 ***
Group.LADref                    0.215915   0.222809   0.969    0.343
FirstWords.months              -0.004027   0.007459  -0.540    0.595
vabs_soc_ss                     0.004265   0.004883   0.874    0.392
Group.LADref:FirstWords.months -0.008095   0.015016  -0.539    0.595
---
Signif. codes:  0 ‘***’ 0.001 ‘**’ 0.01 ‘*’ 0.05 ‘.’ 0.1 ‘ ’ 1

Residual standard error: 0.215 on 22 degrees of freedom
  (25 observations deleted due to missingness)
Multiple R-squared:  0.1622,	Adjusted R-squared:  0.009853
F-statistic: 1.065 on 4 and 22 DF,  p-value: 0.3975

>
> mSupp_ASDNT.Phrases = lm(support.log ~
+                           Group.LADref*FirstPhrases.months
+                 #+ age
+                       + vabs_soc_ss
+                         , data = dASDNT.older18)
> summary(mSupp_ASDNT.Phrases)

Call:
lm(formula = support.log ~ Group.LADref * FirstPhrases.months +
    vabs_soc_ss, data = dASDNT.older18)

Residuals:
     Min       1Q   Median       3Q      Max
-0.49194 -0.06388  0.02726  0.14350  0.27818

Coefficients:
                                  Estimate Std. Error t value Pr(>|t|)
(Intercept)                      3.3597401  0.4176374   8.045 2.12e-08 ***
Group.LADref                     0.1069586  0.2726355   0.392    0.698
FirstPhrases.months              0.0003657  0.0046770   0.078    0.938
vabs_soc_ss                      0.0046382  0.0038083   1.218    0.235
Group.LADref:FirstPhrases.months 0.0001852  0.0092558   0.020    0.984
---
Signif. codes:  0 ‘***’ 0.001 ‘**’ 0.01 ‘*’ 0.05 ‘.’ 0.1 ‘ ’ 1

Residual standard error: 0.2101 on 25 degrees of freedom
  (22 observations deleted due to missingness)
Multiple R-squared:  0.1783,	Adjusted R-squared:  0.04681
F-statistic: 1.356 on 4 and 25 DF,  p-value: 0.2773

>
>
>
>
> ### LAD NT ###
> mSupp_LADNT.GJ = lm(support.log ~
+                           Group.ASDref*aprime.log
+                 #+ age
+                       + vabs_soc_ss
+                         , data = dLADNT.older18)
> summary(mSupp_LADNT.GJ)

Call:
lm(formula = support.log ~ Group.ASDref * aprime.log + vabs_soc_ss,
    data = dLADNT.older18)

Residuals:
     Min       1Q   Median       3Q      Max
-0.49617 -0.05972  0.04249  0.12375  0.18288

Coefficients:
                         Estimate Std. Error t value Pr(>|t|)
(Intercept)              3.656270   0.317719  11.508 5.99e-14 ***
Group.ASDref            -0.046188   0.095561  -0.483    0.632
aprime.log              -0.004340   0.373553  -0.012    0.991
vabs_soc_ss              0.002575   0.002954   0.872    0.389
Group.ASDref:aprime.log -0.138894   0.752276  -0.185    0.855
---
Signif. codes:  0 ‘***’ 0.001 ‘**’ 0.01 ‘*’ 0.05 ‘.’ 0.1 ‘ ’ 1

Residual standard error: 0.1608 on 38 degrees of freedom
  (14 observations deleted due to missingness)
Multiple R-squared:  0.03062,	Adjusted R-squared:  -0.07143
F-statistic:   0.3 on 4 and 38 DF,  p-value: 0.8761

>
> mSupp_LADNT.Words = lm(support.log ~
+                           Group.ASDref*FirstWords.months
+                 #+ age
+                       + vabs_soc_ss
+                         , data = dLADNT.older18)
> summary(mSupp_LADNT.Words)

Call:
lm(formula = support.log ~ Group.ASDref * FirstWords.months +
    vabs_soc_ss, data = dLADNT.older18)

Residuals:
     Min       1Q   Median       3Q      Max
-0.43699 -0.05086  0.00328  0.10738  0.20042

Coefficients:
                                Estimate Std. Error t value Pr(>|t|)
(Intercept)                     3.642016   0.332688  10.947 2.37e-12 ***
Group.ASDref                    0.154751   0.136092   1.137    0.264
FirstWords.months              -0.002677   0.004197  -0.638    0.528
vabs_soc_ss                     0.002902   0.003099   0.936    0.356
Group.ASDref:FirstWords.months -0.010718   0.008397  -1.276    0.211
---
Signif. codes:  0 ‘***’ 0.001 ‘**’ 0.01 ‘*’ 0.05 ‘.’ 0.1 ‘ ’ 1

Residual standard error: 0.1624 on 32 degrees of freedom
  (20 observations deleted due to missingness)
Multiple R-squared:  0.07241,	Adjusted R-squared:  -0.04354
F-statistic: 0.6245 on 4 and 32 DF,  p-value: 0.6484

>
> mSupp_LADNT.Phrases = lm(support.log ~
+                           Group.ASDref*FirstPhrases.months
+                 #+ age
+                       + vabs_soc_ss
+                         , data = dLADNT.older18)
> summary(mSupp_LADNT.Phrases)

Call:
lm(formula = support.log ~ Group.ASDref * FirstPhrases.months +
    vabs_soc_ss, data = dLADNT.older18)

Residuals:
     Min       1Q   Median       3Q      Max
-0.49883 -0.05100  0.01795  0.11967  0.18807

Coefficients:
                                  Estimate Std. Error t value Pr(>|t|)
(Intercept)                       3.620965   0.336447  10.762 1.72e-12 ***
Group.ASDref                      0.025231   0.124047   0.203    0.840
FirstPhrases.months               0.001127   0.002608   0.432    0.668
vabs_soc_ss                       0.002581   0.003138   0.823    0.417
Group.ASDref:FirstPhrases.months -0.001793   0.005231  -0.343    0.734
---
Signif. codes:  0 ‘***’ 0.001 ‘**’ 0.01 ‘*’ 0.05 ‘.’ 0.1 ‘ ’ 1

Residual standard error: 0.1649 on 34 degrees of freedom
  (18 observations deleted due to missingness)
Multiple R-squared:  0.04976,	Adjusted R-squared:  -0.06204
F-statistic: 0.4451 on 4 and 34 DF,  p-value: 0.7752


> ###Follow up
>
> #aprime.log
> mSupp_ASD.GJ = lm(support.log ~ aprime.log
+                 #+ age
+                       + vabs_soc_ss
+                         , data = dASD.older18)
> summary(mSupp_ASD.GJ)

Call:
lm(formula = support.log ~ aprime.log + vabs_soc_ss, data = dASD.older18)

Residuals:
     Min       1Q   Median       3Q      Max
-0.44825 -0.09637 -0.02126  0.17417  0.24803

Coefficients:
             Estimate Std. Error t value Pr(>|t|)
(Intercept)  3.362494   0.475532   7.071 0.000105 ***
aprime.log  -0.051262   0.604246  -0.085 0.934477
vabs_soc_ss  0.004329   0.004889   0.885 0.401776
---
Signif. codes:  0 ‘***’ 0.001 ‘**’ 0.01 ‘*’ 0.05 ‘.’ 0.1 ‘ ’ 1

Residual standard error: 0.2297 on 8 degrees of freedom
  (14 observations deleted due to missingness)
Multiple R-squared:  0.09032,	Adjusted R-squared:  -0.1371
F-statistic: 0.3971 on 2 and 8 DF,  p-value: 0.6848

>
> mSupp_NT.GJ = lm(support.log ~ aprime.log
+                 #+ age
+                       + vabs_soc_ss
+                         , data = dNT.older18)
> summary(mSupp_NT.GJ)

Call:
lm(formula = support.log ~ aprime.log + vabs_soc_ss, data = dNT.older18)

Residuals:
     Min       1Q   Median       3Q      Max
-0.48542 -0.05227  0.06254  0.13630  0.20617

Coefficients:
             Estimate Std. Error t value Pr(>|t|)
(Intercept)  3.349248   0.581240   5.762 1.22e-05 ***
aprime.log  -0.057372   0.575022  -0.100    0.922
vabs_soc_ss  0.005289   0.005502   0.961    0.348
---
Signif. codes:  0 ‘***’ 0.001 ‘**’ 0.01 ‘*’ 0.05 ‘.’ 0.1 ‘ ’ 1

Residual standard error: 0.1959 on 20 degrees of freedom
  (4 observations deleted due to missingness)
Multiple R-squared:  0.04525,	Adjusted R-squared:  -0.05022
F-statistic: 0.474 on 2 and 20 DF,  p-value: 0.6293

>
> mSupp_LAD.GJ = lm(support.log ~ aprime.log
+                 #+ age
+                       + vabs_soc_ss
+                         , data = dLAD.older18)
> summary(mSupp_LAD.GJ)

Call:
lm(formula = support.log ~ aprime.log + vabs_soc_ss, data = dLAD.older18)

Residuals:
      Min        1Q    Median        3Q       Max
-0.193924 -0.065114 -0.006081  0.072444  0.151604

Coefficients:
             Estimate Std. Error t value Pr(>|t|)
(Intercept) 3.8982127  0.2879431  13.538 1.56e-10 ***
aprime.log  0.1394678  0.3975950   0.351    0.730
vabs_soc_ss 0.0005497  0.0026299   0.209    0.837
---
Signif. codes:  0 ‘***’ 0.001 ‘**’ 0.01 ‘*’ 0.05 ‘.’ 0.1 ‘ ’ 1

Residual standard error: 0.1084 on 17 degrees of freedom
  (10 observations deleted due to missingness)
Multiple R-squared:  0.01249,	Adjusted R-squared:  -0.1037
F-statistic: 0.1075 on 2 and 17 DF,  p-value: 0.8987

>
>
> #Words
> mSupp_ASD.Words = lm(support.log ~ FirstWords.months
+                 #+ age
+                       + vabs_soc_ss
+                         , data = dASD.older18)
> summary(mSupp_ASD.Words)

Call:
lm(formula = support.log ~ FirstWords.months + vabs_soc_ss, data = dASD.older18)

Residuals:
        4         8        12        17        19        20        21
 0.005155  0.009456  0.278882  0.270195 -0.042484 -0.124375 -0.396830

Coefficients:
                   Estimate Std. Error t value Pr(>|t|)
(Intercept)        3.658035   1.163737   3.143   0.0347 *
FirstWords.months -0.002906   0.017124  -0.170   0.8735
vabs_soc_ss        0.001590   0.010300   0.154   0.8848
---
Signif. codes:  0 ‘***’ 0.001 ‘**’ 0.01 ‘*’ 0.05 ‘.’ 0.1 ‘ ’ 1

Residual standard error: 0.2853 on 4 degrees of freedom
  (18 observations deleted due to missingness)
Multiple R-squared:  0.03696,	Adjusted R-squared:  -0.4446
F-statistic: 0.07675 on 2 and 4 DF,  p-value: 0.9275

>
> mSupp_NT.Words = lm(support.log ~ FirstWords.months
+                 #+ age
+                       + vabs_soc_ss
+                         , data = dNT.older18)
> summary(mSupp_NT.Words)

Call:
lm(formula = support.log ~ FirstWords.months + vabs_soc_ss, data = dNT.older18)

Residuals:
     Min       1Q   Median       3Q      Max
-0.45123 -0.05534  0.04987  0.14516  0.19655

Coefficients:
                   Estimate Std. Error t value Pr(>|t|)
(Intercept)        3.394591   0.623819   5.442  4.4e-05 ***
FirstWords.months -0.008123   0.009367  -0.867    0.398
vabs_soc_ss        0.006019   0.005853   1.028    0.318
---
Signif. codes:  0 ‘***’ 0.001 ‘**’ 0.01 ‘*’ 0.05 ‘.’ 0.1 ‘ ’ 1

Residual standard error: 0.2003 on 17 degrees of freedom
  (7 observations deleted due to missingness)
Multiple R-squared:  0.09473,	Adjusted R-squared:  -0.01178
F-statistic: 0.8894 on 2 and 17 DF,  p-value: 0.4292

>
> mSupp_LAD.Words = lm(support.log ~ FirstWords.months
+                 #+ age
+                       + vabs_soc_ss
+                         , data = dLAD.older18)
> summary(mSupp_LAD.Words)

Call:
lm(formula = support.log ~ FirstWords.months + vabs_soc_ss, data = dLAD.older18)

Residuals:
     Min       1Q   Median       3Q      Max
-0.15942 -0.05948  0.00563  0.02884  0.18795

Coefficients:
                  Estimate Std. Error t value Pr(>|t|)
(Intercept)       3.810392   0.273975  13.908 1.38e-09 ***
FirstWords.months 0.002559   0.002221   1.152    0.269
vabs_soc_ss       0.000583   0.002537   0.230    0.822
---
Signif. codes:  0 ‘***’ 0.001 ‘**’ 0.01 ‘*’ 0.05 ‘.’ 0.1 ‘ ’ 1

Residual standard error: 0.1007 on 14 degrees of freedom
  (13 observations deleted due to missingness)
Multiple R-squared:  0.08812,	Adjusted R-squared:  -0.04215
F-statistic: 0.6764 on 2 and 14 DF,  p-value: 0.5243

>
> #Phrases
> mSupp_ASD.Phrases = lm(support.log ~ FirstPhrases.months
+                 #+ age
+                       + vabs_soc_ss
+                         , data = dASD.older18)
> summary(mSupp_ASD.Phrases)

Call:
lm(formula = support.log ~ FirstPhrases.months + vabs_soc_ss,
    data = dASD.older18)

Residuals:
        4         8        12        17        19        20        21        24
 0.038362  0.009565  0.273389  0.277865 -0.043096 -0.049899 -0.417395 -0.088792

Coefficients:
                    Estimate Std. Error t value Pr(>|t|)
(Intercept)         3.333141   0.681364   4.892  0.00451 **
FirstPhrases.months 0.000197   0.008961   0.022  0.98332
vabs_soc_ss         0.004370   0.006087   0.718  0.50494
---
Signif. codes:  0 ‘***’ 0.001 ‘**’ 0.01 ‘*’ 0.05 ‘.’ 0.1 ‘ ’ 1

Residual standard error: 0.2608 on 5 degrees of freedom
  (17 observations deleted due to missingness)
Multiple R-squared:  0.09577,	Adjusted R-squared:  -0.2659
F-statistic: 0.2648 on 2 and 5 DF,  p-value: 0.7775

>
> mSupp_NT.Phrases = lm(support.log ~ FirstPhrases.months
+                 #+ age
+                       + vabs_soc_ss
+                         , data = dNT.older18)
> summary(mSupp_NT.Phrases)

Call:
lm(formula = support.log ~ FirstPhrases.months + vabs_soc_ss,
    data = dNT.older18)

Residuals:
     Min       1Q   Median       3Q      Max
-0.49058 -0.05969  0.05052  0.14207  0.20645

Coefficients:
                     Estimate Std. Error t value Pr(>|t|)
(Intercept)         3.3696680  0.6279151   5.366 3.53e-05 ***
FirstPhrases.months 0.0005034  0.0056826   0.089    0.930
vabs_soc_ss         0.0050448  0.0057642   0.875    0.392
---
Signif. codes:  0 ‘***’ 0.001 ‘**’ 0.01 ‘*’ 0.05 ‘.’ 0.1 ‘ ’ 1

Residual standard error: 0.2005 on 19 degrees of freedom
  (5 observations deleted due to missingness)
Multiple R-squared:  0.03876,	Adjusted R-squared:  -0.06243
F-statistic: 0.383 on 2 and 19 DF,  p-value: 0.6869

>
> mSupp_LAD.Phrases = lm(support.log ~ FirstPhrases.months
+                 #+ age
+                       + vabs_soc_ss
+                         , data = dLAD.older18)
> summary(mSupp_LAD.Phrases)

Call:
lm(formula = support.log ~ FirstPhrases.months + vabs_soc_ss,
    data = dLAD.older18)

Residuals:
      Min        1Q    Median        3Q       Max
-0.156735 -0.053136  0.004896  0.043379  0.183651

Coefficients:
                     Estimate Std. Error t value Pr(>|t|)
(Intercept)         3.8074457  0.2750001  13.845 1.46e-09 ***
FirstPhrases.months 0.0020909  0.0014754   1.417    0.178
vabs_soc_ss         0.0006605  0.0026084   0.253    0.804
---
Signif. codes:  0 ‘***’ 0.001 ‘**’ 0.01 ‘*’ 0.05 ‘.’ 0.1 ‘ ’ 1

Residual standard error: 0.1027 on 14 degrees of freedom
  (13 observations deleted due to missingness)
Multiple R-squared:  0.1318,	Adjusted R-squared:  0.007785
F-statistic: 1.063 on 2 and 14 DF,  p-value: 0.3718


> ###################
> ##### Friends #####
> ###################
> #combined friends and social
>
> ### ASD LAD ###
> mFriend.AB_ASDLAD.GJ = lm(friends ~
+                           Group.NTref*aprime.log
+                  #+ age
+                       + vabs_soc_ss
+                         , data = dASDLAD)
> summary(mFriend.AB_ASDLAD.GJ)

Call:
lm(formula = friends ~ Group.NTref * aprime.log + vabs_soc_ss,
    data = dASDLAD)

Residuals:
     Min       1Q   Median       3Q      Max
-17.4525  -4.1744   0.1857   6.4812  16.9376

Coefficients:
                       Estimate Std. Error t value Pr(>|t|)
(Intercept)             35.1036    10.6622   3.292  0.00223 **
Group.NTref              1.0372     6.1262   0.169  0.86651
aprime.log               8.5916    15.4889   0.555  0.58253
vabs_soc_ss              0.1394     0.1066   1.308  0.19917
Group.NTref:aprime.log -66.9677    31.5135  -2.125  0.04052 *
---
Signif. codes:  0 ‘***’ 0.001 ‘**’ 0.01 ‘*’ 0.05 ‘.’ 0.1 ‘ ’ 1

Residual standard error: 9.306 on 36 degrees of freedom
  (30 observations deleted due to missingness)
Multiple R-squared:  0.4627,	Adjusted R-squared:  0.403
F-statistic: 7.752 on 4 and 36 DF,  p-value: 0.0001298

>
> mFriend.AB_ASDLAD.Words = lm(friends ~
+                           Group.NTref*FirstWords.months
+                  #+ age
+                       + vabs_soc_ss
+                         , data = dASDLAD)
> summary(mFriend.AB_ASDLAD.Words)

Call:
lm(formula = friends ~ Group.NTref * FirstWords.months + vabs_soc_ss,
    data = dASDLAD)

Residuals:
     Min       1Q   Median       3Q      Max
-26.5377  -3.8599  -0.6976   5.3181  17.8144

Coefficients:
                              Estimate Std. Error t value Pr(>|t|)
(Intercept)                   52.95676   10.68910   4.954 2.45e-05 ***
Group.NTref                    3.40782    6.29649   0.541  0.59222
FirstWords.months             -0.40128    0.14458  -2.776  0.00926 **
vabs_soc_ss                    0.01522    0.09952   0.153  0.87941
Group.NTref:FirstWords.months  0.59709    0.27928   2.138  0.04051 *
---
Signif. codes:  0 ‘***’ 0.001 ‘**’ 0.01 ‘*’ 0.05 ‘.’ 0.1 ‘ ’ 1

Residual standard error: 8.376 on 31 degrees of freedom
  (35 observations deleted due to missingness)
Multiple R-squared:  0.5753,	Adjusted R-squared:  0.5205
F-statistic:  10.5 on 4 and 31 DF,  p-value: 1.706e-05

>
> mFriend.AB_ASDLAD.Phrases = lm(friends ~
+                           Group.NTref*FirstPhrases.months
+                  #+ age
+                       + vabs_soc_ss
+                         , data = dASDLAD)
> summary(mFriend.AB_ASDLAD.Phrases)

Call:
lm(formula = friends ~ Group.NTref * FirstPhrases.months + vabs_soc_ss,
    data = dASDLAD)

Residuals:
     Min       1Q   Median       3Q      Max
-16.8647  -7.6100   0.6704   5.3433  16.4505

Coefficients:
                                Estimate Std. Error t value Pr(>|t|)
(Intercept)                      43.1477    10.5828   4.077 0.000282 ***
Group.NTref                      -2.1795     8.6152  -0.253 0.801899
FirstPhrases.months              -0.2112     0.1309  -1.613 0.116467
vabs_soc_ss                       0.1010     0.1001   1.009 0.320673
Group.NTref:FirstPhrases.months   0.4843     0.2617   1.851 0.073416 .
---
Signif. codes:  0 ‘***’ 0.001 ‘**’ 0.01 ‘*’ 0.05 ‘.’ 0.1 ‘ ’ 1

Residual standard error: 9.05 on 32 degrees of freedom
  (34 observations deleted due to missingness)
Multiple R-squared:  0.4852,	Adjusted R-squared:  0.4209
F-statistic: 7.541 on 4 and 32 DF,  p-value: 0.0002131

>
>
>
>
> ### ASD NT ###
> mFriend.AB_ASDNT.GJ = lm(friends ~
+                           Group.LADref*aprime.log
+                 #+ age
+                       + vabs_soc_ss
+                         , data = dASDNT)
> summary(mFriend.AB_ASDNT.GJ)

Call:
lm(formula = friends ~ Group.LADref * aprime.log + vabs_soc_ss,
    data = dASDNT)

Residuals:
     Min       1Q   Median       3Q      Max
-25.3498  -5.4847   0.9872   6.8459  16.9797

Coefficients:
                        Estimate Std. Error t value Pr(>|t|)
(Intercept)              40.2984    10.7875   3.736 0.000571 ***
Group.LADref              6.4971     6.6670   0.975 0.335516
aprime.log               32.0884    17.2543   1.860 0.070109 .
vabs_soc_ss               0.1095     0.1098   0.997 0.324482
Group.LADref:aprime.log -19.3954    34.5467  -0.561 0.577563
---
Signif. codes:  0 ‘***’ 0.001 ‘**’ 0.01 ‘*’ 0.05 ‘.’ 0.1 ‘ ’ 1

Residual standard error: 10.06 on 41 degrees of freedom
  (31 observations deleted due to missingness)
Multiple R-squared:  0.3891,	Adjusted R-squared:  0.3295
F-statistic: 6.528 on 4 and 41 DF,  p-value: 0.0003681

>
> mFriend.AB_ASDNT.Words = lm(friends ~
+                           Group.LADref*FirstWords.months
+                 #+ age
+                       + vabs_soc_ss
+                         , data = dASDNT)
> summary(mFriend.AB_ASDNT.Words)

Call:
lm(formula = friends ~ Group.LADref * FirstWords.months + vabs_soc_ss,
    data = dASDNT)

Residuals:
     Min       1Q   Median       3Q      Max
-27.0590  -4.7470   0.9386   5.9832  17.7092

Coefficients:
                               Estimate Std. Error t value Pr(>|t|)
(Intercept)                     51.5560    12.5464   4.109 0.000227 ***
Group.LADref                     8.9937     8.4024   1.070 0.291779
FirstWords.months               -0.7254     0.2638  -2.750 0.009378 **
vabs_soc_ss                      0.0578     0.1169   0.494 0.624250
Group.LADref:FirstWords.months  -0.1149     0.5293  -0.217 0.829397
---
Signif. codes:  0 ‘***’ 0.001 ‘**’ 0.01 ‘*’ 0.05 ‘.’ 0.1 ‘ ’ 1

Residual standard error: 9.86 on 35 degrees of freedom
  (37 observations deleted due to missingness)
Multiple R-squared:  0.4472,	Adjusted R-squared:  0.3841
F-statistic:  7.08 on 4 and 35 DF,  p-value: 0.0002755

>
> mFriend.AB_ASDNT.Phrases = lm(friends ~
+                           Group.LADref*FirstPhrases.months
+                 #+ age
+                       + vabs_soc_ss
+                         , data = dASDNT)
> summary(mFriend.AB_ASDNT.Phrases)

Call:
lm(formula = friends ~ Group.LADref * FirstPhrases.months + vabs_soc_ss,
    data = dASDNT)

Residuals:
    Min      1Q  Median      3Q     Max
-21.117  -8.451   1.282   5.931  16.188

Coefficients:
                                 Estimate Std. Error t value Pr(>|t|)
(Intercept)                      42.53356   12.17663   3.493  0.00126 **
Group.LADref                      4.02881   10.49220   0.384  0.70319
FirstPhrases.months              -0.41732    0.19840  -2.103  0.04229 *
vabs_soc_ss                       0.14211    0.11297   1.258  0.21628
Group.LADref:FirstPhrases.months  0.05193    0.39676   0.131  0.89657
---
Signif. codes:  0 ‘***’ 0.001 ‘**’ 0.01 ‘*’ 0.05 ‘.’ 0.1 ‘ ’ 1

Residual standard error: 10.22 on 37 degrees of freedom
  (35 observations deleted due to missingness)
Multiple R-squared:  0.3927,	Adjusted R-squared:  0.3271
F-statistic: 5.982 on 4 and 37 DF,  p-value: 0.0008128

>
>
>
>
> ### LAD NT ###
> mFriend.AB_LADNT.GJ = lm(friends ~
+                           Group.ASDref*aprime.log
+                 #+ age
+                       + vabs_soc_ss
+                         , data = dLADNT)
> summary(mFriend.AB_LADNT.GJ)

Call:
lm(formula = friends ~ Group.ASDref * aprime.log + vabs_soc_ss,
    data = dLADNT)

Residuals:
     Min       1Q   Median       3Q      Max
-24.9395  -2.4434   0.6224   6.0033  10.3747

Coefficients:
                        Estimate Std. Error t value Pr(>|t|)
(Intercept)              30.8034    13.1712   2.339   0.0242 *
Group.ASDref              5.1474     4.6381   1.110   0.2734
aprime.log               -4.0470    16.1119  -0.251   0.8029
vabs_soc_ss               0.2059     0.1197   1.720   0.0927 .
Group.ASDref:aprime.log  51.7960    31.8431   1.627   0.1113
---
Signif. codes:  0 ‘***’ 0.001 ‘**’ 0.01 ‘*’ 0.05 ‘.’ 0.1 ‘ ’ 1

Residual standard error: 8.169 on 42 degrees of freedom
  (23 observations deleted due to missingness)
Multiple R-squared:  0.09827,	Adjusted R-squared:  0.01239
F-statistic: 1.144 on 4 and 42 DF,  p-value: 0.349

>
> mFriend.AB_LADNT.Words = lm(friends ~
+                           Group.ASDref*FirstWords.months
+                 #+ age
+                       + vabs_soc_ss
+                         , data = dLADNT)
> summary(mFriend.AB_LADNT.Words)

Call:
lm(formula = friends ~ Group.ASDref * FirstWords.months + vabs_soc_ss,
    data = dLADNT)

Residuals:
     Min       1Q   Median       3Q      Max
-17.7020  -2.5264   0.8874   5.0977  11.4786

Coefficients:
                               Estimate Std. Error t value Pr(>|t|)
(Intercept)                     49.2244    11.4450   4.301  0.00011 ***
Group.ASDref                     6.0640     6.2618   0.968  0.33880
FirstWords.months               -0.4361     0.1948  -2.239  0.03095 *
vabs_soc_ss                      0.0944     0.1045   0.903  0.37194
Group.ASDref:FirstWords.months  -0.6966     0.3900  -1.786  0.08185 .
---
Signif. codes:  0 ‘***’ 0.001 ‘**’ 0.01 ‘*’ 0.05 ‘.’ 0.1 ‘ ’ 1

Residual standard error: 7.554 on 39 degrees of freedom
  (26 observations deleted due to missingness)
Multiple R-squared:  0.1551,	Adjusted R-squared:  0.0684
F-statistic: 1.789 on 4 and 39 DF,  p-value: 0.1506

>
> mFriend.AB_LADNT.Phrases = lm(friends ~
+                           Group.ASDref*FirstPhrases.months
+                 #+ age
+                       + vabs_soc_ss
+                         , data = dLADNT)
> summary(mFriend.AB_LADNT.Phrases)

Call:
lm(formula = friends ~ Group.ASDref * FirstPhrases.months + vabs_soc_ss,
    data = dLADNT)

Residuals:
    Min      1Q  Median      3Q     Max
-21.309  -2.553   1.149   5.320  10.648

Coefficients:
                                 Estimate Std. Error t value Pr(>|t|)
(Intercept)                      46.88344   11.39538   4.114 0.000188 ***
Group.ASDref                      6.80926    5.92432   1.149 0.257228
FirstPhrases.months              -0.18030    0.12650  -1.425 0.161810
vabs_soc_ss                       0.08722    0.10603   0.823 0.415656
Group.ASDref:FirstPhrases.months -0.42241    0.25299  -1.670 0.102796
---
Signif. codes:  0 ‘***’ 0.001 ‘**’ 0.01 ‘*’ 0.05 ‘.’ 0.1 ‘ ’ 1

Residual standard error: 7.719 on 40 degrees of freedom
  (25 observations deleted due to missingness)
Multiple R-squared:  0.09357,	Adjusted R-squared:  0.002923
F-statistic: 1.032 on 4 and 40 DF,  p-value: 0.4025


> ###Follow up
>
> #aprime.log
> mFriend.AB_ASD.GJ = lm(friends ~ aprime.log
+                 #+ age
+                       + vabs_soc_ss
+                         , data = dASD)
> summary(mFriend.AB_ASD.GJ)

Call:
lm(formula = friends ~ aprime.log + vabs_soc_ss, data = dASD)

Residuals:
     Min       1Q   Median       3Q      Max
-15.7653  -7.9147  -0.0057   9.5987  17.0072

Coefficients:
            Estimate Std. Error t value Pr(>|t|)
(Intercept) 38.66118   13.47655   2.869   0.0106 *
aprime.log  41.59695   23.06047   1.804   0.0890 .
vabs_soc_ss  0.09005    0.15136   0.595   0.5598
---
Signif. codes:  0 ‘***’ 0.001 ‘**’ 0.01 ‘*’ 0.05 ‘.’ 0.1 ‘ ’ 1

Residual standard error: 11.4 on 17 degrees of freedom
  (19 observations deleted due to missingness)
Multiple R-squared:  0.1701,	Adjusted R-squared:  0.0725
F-statistic: 1.743 on 2 and 17 DF,  p-value: 0.2049

>
> mFriend.AB_NT.GJ = lm(friends ~ aprime.log
+                 #+ age
+                       + vabs_soc_ss
+                         , data = dNT)
> summary(mFriend.AB_NT.GJ)

Call:
lm(formula = friends ~ aprime.log + vabs_soc_ss, data = dNT)

Residuals:
    Min      1Q  Median      3Q     Max
-25.176  -2.075   1.614   5.343  10.711

Coefficients:
            Estimate Std. Error t value Pr(>|t|)
(Intercept)  39.2300    18.9950   2.065   0.0503 .
aprime.log   22.1616    25.4338   0.871   0.3926
vabs_soc_ss   0.1504     0.1761   0.854   0.4016
---
Signif. codes:  0 ‘***’ 0.001 ‘**’ 0.01 ‘*’ 0.05 ‘.’ 0.1 ‘ ’ 1

Residual standard error: 9.158 on 23 degrees of freedom
  (12 observations deleted due to missingness)
Multiple R-squared:  0.06312,	Adjusted R-squared:  -0.01835
F-statistic: 0.7747 on 2 and 23 DF,  p-value: 0.4725

>
> mFriend.AB_LAD.GJ = lm(friends ~ aprime.log
+                 #+ age
+                       + vabs_soc_ss
+                         , data = dLAD)
> summary(mFriend.AB_LAD.GJ)

Call:
lm(formula = friends ~ aprime.log + vabs_soc_ss, data = dLAD)

Residuals:
     Min       1Q   Median       3Q      Max
-14.9816  -2.4972   0.5485   5.4182   8.8524

Coefficients:
            Estimate Std. Error t value Pr(>|t|)
(Intercept)  19.6916    17.5060   1.125   0.2754
aprime.log  -35.7808    21.1017  -1.696   0.1072
vabs_soc_ss   0.2827     0.1558   1.814   0.0864 .
---
Signif. codes:  0 ‘***’ 0.001 ‘**’ 0.01 ‘*’ 0.05 ‘.’ 0.1 ‘ ’ 1

Residual standard error: 6.888 on 18 degrees of freedom
  (11 observations deleted due to missingness)
Multiple R-squared:  0.1803,	Adjusted R-squared:  0.08927
F-statistic:  1.98 on 2 and 18 DF,  p-value: 0.167

>
> #Words
> mFriend.AB_ASD.Words = lm(friends ~ FirstWords.months
+                 #+ age
+                       + vabs_soc_ss
+                         , data = dASD)
> summary(mFriend.AB_ASD.Words)

Call:
lm(formula = friends ~ FirstWords.months + vabs_soc_ss, data = dASD)

Residuals:
    Min      1Q  Median      3Q     Max
-26.386  -5.484  -3.080   6.476  17.845

Coefficients:
                   Estimate Std. Error t value Pr(>|t|)
(Intercept)       52.472176  17.753109   2.956   0.0111 *
FirstWords.months -0.709103   0.305360  -2.322   0.0371 *
vabs_soc_ss        0.002845   0.169041   0.017   0.9868
---
Signif. codes:  0 ‘***’ 0.001 ‘**’ 0.01 ‘*’ 0.05 ‘.’ 0.1 ‘ ’ 1

Residual standard error: 11.32 on 13 degrees of freedom
  (23 observations deleted due to missingness)
Multiple R-squared:  0.3351,	Adjusted R-squared:  0.2328
F-statistic: 3.276 on 2 and 13 DF,  p-value: 0.07044

>
> mFriend.AB_NT.Words = lm(friends ~ FirstWords.months
+                 #+ age
+                       + vabs_soc_ss
+                         , data = dNT)
> summary(mFriend.AB_NT.Words)

Call:
lm(formula = friends ~ FirstWords.months + vabs_soc_ss, data = dNT)

Residuals:
    Min      1Q  Median      3Q     Max
-17.980  -1.774   2.639   5.442  11.163

Coefficients:
                  Estimate Std. Error t value Pr(>|t|)
(Intercept)        46.3130    19.1462   2.419   0.0247 *
FirstWords.months  -0.7868     0.4212  -1.868   0.0758 .
vabs_soc_ss         0.1517     0.1757   0.864   0.3976
---
Signif. codes:  0 ‘***’ 0.001 ‘**’ 0.01 ‘*’ 0.05 ‘.’ 0.1 ‘ ’ 1

Residual standard error: 8.999 on 21 degrees of freedom
  (14 observations deleted due to missingness)
Multiple R-squared:  0.166,	Adjusted R-squared:  0.08653
F-statistic: 2.089 on 2 and 21 DF,  p-value: 0.1487

>
> mFriend.AB_LAD.Words = lm(friends ~ FirstWords.months
+                 #+ age
+                       + vabs_soc_ss
+                         , data = dLAD)
> summary(mFriend.AB_LAD.Words)

Call:
lm(formula = friends ~ FirstWords.months + vabs_soc_ss, data = dLAD)

Residuals:
     Min       1Q   Median       3Q      Max
-15.0354  -2.7363  -0.2104   3.6918   8.3373

Coefficients:
                  Estimate Std. Error t value Pr(>|t|)
(Intercept)       52.37744   11.79312   4.441 0.000358 ***
FirstWords.months -0.09870    0.11953  -0.826 0.420392
vabs_soc_ss        0.03657    0.10715   0.341 0.737056
---
Signif. codes:  0 ‘***’ 0.001 ‘**’ 0.01 ‘*’ 0.05 ‘.’ 0.1 ‘ ’ 1

Residual standard error: 5.464 on 17 degrees of freedom
  (12 observations deleted due to missingness)
Multiple R-squared:  0.05136,	Adjusted R-squared:  -0.06025
F-statistic: 0.4602 on 2 and 17 DF,  p-value: 0.6388

>
> #Phrases
> mFriend.AB_ASD.Phrases = lm(friends ~ FirstPhrases.months
+                 #+ age
+                       + vabs_soc_ss
+                         , data = dASD)
> summary(mFriend.AB_ASD.Phrases)

Call:
lm(formula = friends ~ FirstPhrases.months + vabs_soc_ss, data = dASD)

Residuals:
     Min       1Q   Median       3Q      Max
-13.9183  -9.8710   0.8971   9.7325  16.2138

Coefficients:
                    Estimate Std. Error t value Pr(>|t|)
(Intercept)          40.8856    17.7238   2.307   0.0369 *
FirstPhrases.months  -0.4443     0.3057  -1.453   0.1682
vabs_soc_ss           0.1381     0.1628   0.848   0.4106
---
Signif. codes:  0 ‘***’ 0.001 ‘**’ 0.01 ‘*’ 0.05 ‘.’ 0.1 ‘ ’ 1

Residual standard error: 12.1 on 14 degrees of freedom
  (22 observations deleted due to missingness)
Multiple R-squared:  0.1864,	Adjusted R-squared:  0.07015
F-statistic: 1.604 on 2 and 14 DF,  p-value: 0.236

>
> mFriend.AB_NT.Phrases = lm(friends ~ FirstPhrases.months
+                 #+ age
+                       + vabs_soc_ss
+                         , data = dNT)
> summary(mFriend.AB_NT.Phrases)

Call:
lm(formula = friends ~ FirstPhrases.months + vabs_soc_ss, data = dNT)

Residuals:
    Min      1Q  Median      3Q     Max
-21.087  -3.349   1.818   5.518  11.123

Coefficients:
                    Estimate Std. Error t value Pr(>|t|)
(Intercept)          43.6671    19.2225   2.272   0.0332 *
FirstPhrases.months  -0.3913     0.2683  -1.459   0.1588
vabs_soc_ss           0.1505     0.1762   0.855   0.4020
---
Signif. codes:  0 ‘***’ 0.001 ‘**’ 0.01 ‘*’ 0.05 ‘.’ 0.1 ‘ ’ 1

Residual standard error: 9.079 on 22 degrees of freedom
  (13 observations deleted due to missingness)
Multiple R-squared:  0.1151,	Adjusted R-squared:  0.0347
F-statistic: 1.431 on 2 and 22 DF,  p-value: 0.2604

>
> mFriend.AB_LAD.Phrases = lm(friends ~ FirstPhrases.months
+                 #+ age
+                       + vabs_soc_ss
+                         , data = dLAD)
> summary(mFriend.AB_LAD.Phrases)

Call:
lm(formula = friends ~ FirstPhrases.months + vabs_soc_ss, data = dLAD)

Residuals:
    Min      1Q  Median      3Q     Max
-16.766  -2.214   1.093   2.821   6.825

Coefficients:
                    Estimate Std. Error t value Pr(>|t|)
(Intercept)         50.03161   11.68972   4.280 0.000506 ***
FirstPhrases.months  0.03051    0.08057   0.379 0.709653
vabs_soc_ss          0.02358    0.11049   0.213 0.833506
---
Signif. codes:  0 ‘***’ 0.001 ‘**’ 0.01 ‘*’ 0.05 ‘.’ 0.1 ‘ ’ 1

Residual standard error: 5.68 on 17 degrees of freedom
  (12 observations deleted due to missingness)
Multiple R-squared:  0.01091,	Adjusted R-squared:  -0.1055
F-statistic: 0.09378 on 2 and 17 DF,  p-value: 0.9109


> ### CELF ###
>
> #Core Language "cls_std_scr",
> mFriend_ASD.CL = data = lm(cbcl_social_t ~ cls_std_scr
+                       #+ age
+                       + vabs_soc_ss
+                         , dASD.inperson)
> summary(mFriend_ASD.CL)

Call:
lm(formula = cbcl_social_t ~ cls_std_scr + vabs_soc_ss, data = dASD.inperson)

Residuals:
    Min      1Q  Median      3Q     Max
-13.061  -8.678  -3.051   7.672  21.981

Coefficients:
            Estimate Std. Error t value Pr(>|t|)
(Intercept) 29.89561   34.66633   0.862    0.417
cls_std_scr  0.11252    0.30144   0.373    0.720
vabs_soc_ss -0.05527    0.26070  -0.212    0.838

Residual standard error: 13.01 on 7 degrees of freedom
  (8 observations deleted due to missingness)
Multiple R-squared:  0.02247,	Adjusted R-squared:  -0.2568
F-statistic: 0.08046 on 2 and 7 DF,  p-value: 0.9235

>
> mFriend_LAD.CL = data = lm(cbcl_social_t ~ cls_std_scr
+                       #+ age
+                       + vabs_soc_ss
+                         , dLAD.inperson)
> summary(mFriend_LAD.CL)

Call:
lm(formula = cbcl_social_t ~ cls_std_scr + vabs_soc_ss, data = dLAD.inperson)

Residuals:
ALL 3 residuals are 0: no residual degrees of freedom!

Coefficients:
             Estimate Std. Error t value Pr(>|t|)
(Intercept) -124.6443        NaN     NaN      NaN
cls_std_scr    2.1804        NaN     NaN      NaN
vabs_soc_ss   -0.4794        NaN     NaN      NaN

Residual standard error: NaN on 0 degrees of freedom
  (2 observations deleted due to missingness)
Multiple R-squared:      1,	Adjusted R-squared:    NaN
F-statistic:   NaN on 2 and 0 DF,  p-value: NA

>
> mFriend_NT.CL = data = lm(cbcl_social_t ~ cls_std_scr
+                       #+ age
+                       + vabs_soc_ss
+                         , dNT.inperson)
> summary(mFriend_NT.CL)

Call:
lm(formula = cbcl_social_t ~ cls_std_scr + vabs_soc_ss, data = dNT.inperson)

Residuals:
      1       2       3       5       6       7       9      10
 7.7581 -9.6882  5.7673 -6.9298  0.7642 -5.1400 12.7314 -5.2630

Coefficients:
             Estimate Std. Error t value Pr(>|t|)
(Intercept) -112.9152   101.8217  -1.109    0.318
cls_std_scr    0.7409     0.5008   1.479    0.199
vabs_soc_ss    0.6962     0.4563   1.526    0.188

Residual standard error: 9.509 on 5 degrees of freedom
  (4 observations deleted due to missingness)
Multiple R-squared:  0.3292,	Adjusted R-squared:  0.06087
F-statistic: 1.227 on 2 and 5 DF,  p-value: 0.3685

>
> #Recalling Sentences "rs_ss"
> mFriend_ASD.RS = lm(cbcl_social_t ~ rs_ss
+                       #+ age
+                       + vabs_soc_ss
+                         , data = dASD.inperson)
> summary(mFriend_ASD.RS)

Call:
lm(formula = cbcl_social_t ~ rs_ss + vabs_soc_ss, data = dASD.inperson)

Residuals:
    Min      1Q  Median      3Q     Max
-12.448  -7.390  -3.446   7.769  23.058

Coefficients:
            Estimate Std. Error t value Pr(>|t|)
(Intercept)  41.5264    19.8761   2.089   0.0751 .
rs_ss         1.0589     1.5962   0.663   0.5283
vabs_soc_ss  -0.2126     0.3642  -0.584   0.5777
---
Signif. codes:  0 ‘***’ 0.001 ‘**’ 0.01 ‘*’ 0.05 ‘.’ 0.1 ‘ ’ 1

Residual standard error: 12.75 on 7 degrees of freedom
  (8 observations deleted due to missingness)
Multiple R-squared:  0.06199,	Adjusted R-squared:  -0.206
F-statistic: 0.2313 on 2 and 7 DF,  p-value: 0.7993

>
> mFriend_LAD.RS = lm(cbcl_social_t ~ rs_ss
+                       #+ age
+                       + vabs_soc_ss
+                         , data = dLAD.inperson)
> summary(mFriend_LAD.RS)

Call:
lm(formula = cbcl_social_t ~ rs_ss + vabs_soc_ss, data = dLAD.inperson)

Residuals:
ALL 3 residuals are 0: no residual degrees of freedom!

Coefficients:
            Estimate Std. Error t value Pr(>|t|)
(Intercept)  51.7581        NaN     NaN      NaN
rs_ss         3.4113        NaN     NaN      NaN
vabs_soc_ss  -0.3387        NaN     NaN      NaN

Residual standard error: NaN on 0 degrees of freedom
  (2 observations deleted due to missingness)
Multiple R-squared:      1,	Adjusted R-squared:    NaN
F-statistic:   NaN on 2 and 0 DF,  p-value: NA

>
> mFriend_NT.RS = lm(cbcl_social_t ~ rs_ss
+                       #+ age
+                       + vabs_soc_ss
+                         , data = dNT.inperson)
> summary(mFriend_NT.RS)

Call:
lm(formula = cbcl_social_t ~ rs_ss + vabs_soc_ss, data = dNT.inperson)

Residuals:
      1       2       3       5       6       7       9      10
  8.561  -2.394   1.165 -13.729  -2.946  -1.018  13.935  -3.574

Coefficients:
            Estimate Std. Error t value Pr(>|t|)
(Intercept) -19.4227    48.2782  -0.402    0.704
rs_ss         2.2945     1.7626   1.302    0.250
vabs_soc_ss   0.3501     0.3059   1.145    0.304

Residual standard error: 9.854 on 5 degrees of freedom
  (4 observations deleted due to missingness)
Multiple R-squared:  0.2797,	Adjusted R-squared:  -0.008485
F-statistic: 0.9706 on 2 and 5 DF,  p-value: 0.4404


> ######################
> ##### Total Prob #####
> ######################
> #combined
> ### ASD LAD ###
> mProb.AB_ASDLAD.GJ = lm(TotalProb ~
+                           Group.NTref*aprime.log
+                  #+ age
+                       + vabs_soc_ss
+                         , data = dASDLAD)
> summary(mProb.AB_ASDLAD.GJ)

Call:
lm(formula = TotalProb ~ Group.NTref * aprime.log + vabs_soc_ss,
    data = dASDLAD)

Residuals:
     Min       1Q   Median       3Q      Max
-22.4753  -8.0655   0.2016   5.7779  20.8954

Coefficients:
                       Estimate Std. Error t value Pr(>|t|)
(Intercept)             95.7475    12.3441   7.757 3.43e-09 ***
Group.NTref             -1.3270     7.0803  -0.187  0.85238
aprime.log               1.0885    17.9912   0.061  0.95209
vabs_soc_ss             -0.4833     0.1233  -3.920  0.00038 ***
Group.NTref:aprime.log  25.6169    36.6030   0.700  0.48852
---
Signif. codes:  0 ‘***’ 0.001 ‘**’ 0.01 ‘*’ 0.05 ‘.’ 0.1 ‘ ’ 1

Residual standard error: 10.78 on 36 degrees of freedom
  (30 observations deleted due to missingness)
Multiple R-squared:  0.4807,	Adjusted R-squared:  0.423
F-statistic: 8.332 on 4 and 36 DF,  p-value: 7.276e-05

>
> mProb.AB_ASDLAD.Words = lm(TotalProb ~
+                           Group.NTref*FirstWords.months
+                  #+ age
+                       + vabs_soc_ss
+                         , data = dASDLAD)
> summary(mProb.AB_ASDLAD.Words)

Call:
lm(formula = TotalProb ~ Group.NTref * FirstWords.months + vabs_soc_ss,
    data = dASDLAD)

Residuals:
    Min      1Q  Median      3Q     Max
-18.579  -6.767  -1.630   7.166  20.976

Coefficients:
                              Estimate Std. Error t value Pr(>|t|)
(Intercept)                   90.45033   13.78662   6.561 2.94e-07 ***
Group.NTref                   -6.10997    8.27492  -0.738  0.46603
FirstWords.months              0.10490    0.18796   0.558  0.58092
vabs_soc_ss                   -0.44974    0.12758  -3.525  0.00138 **
Group.NTref:FirstWords.months -0.04779    0.36146  -0.132  0.89569
---
Signif. codes:  0 ‘***’ 0.001 ‘**’ 0.01 ‘*’ 0.05 ‘.’ 0.1 ‘ ’ 1

Residual standard error: 10.72 on 30 degrees of freedom
  (36 observations deleted due to missingness)
Multiple R-squared:  0.5349,	Adjusted R-squared:  0.4729
F-statistic: 8.625 on 4 and 30 DF,  p-value: 9.308e-05

>
> mProb.AB_ASDLAD.Phrases = lm(TotalProb ~
+                           Group.NTref*FirstPhrases.months
+                  #+ age
+                       + vabs_soc_ss
+                         , data = dASDLAD)
> summary(mProb.AB_ASDLAD.Phrases)

Call:
lm(formula = TotalProb ~ Group.NTref * FirstPhrases.months +
    vabs_soc_ss, data = dASDLAD)

Residuals:
     Min       1Q   Median       3Q      Max
-19.7556  -6.3901  -0.8667   6.3696  19.1034

Coefficients:
                                 Estimate Std. Error t value Pr(>|t|)
(Intercept)                      87.12395   12.73786   6.840 1.15e-07 ***
Group.NTref                     -13.47941   10.49387  -1.285  0.20848
FirstPhrases.months               0.09778    0.15833   0.618  0.54137
vabs_soc_ss                      -0.42537    0.11992  -3.547  0.00126 **
Group.NTref:FirstPhrases.months   0.26878    0.31624   0.850  0.40188
---
Signif. codes:  0 ‘***’ 0.001 ‘**’ 0.01 ‘*’ 0.05 ‘.’ 0.1 ‘ ’ 1

Residual standard error: 10.83 on 31 degrees of freedom
  (35 observations deleted due to missingness)
Multiple R-squared:  0.488,	Adjusted R-squared:  0.4219
F-statistic: 7.386 on 4 and 31 DF,  p-value: 0.0002671

>
>
>
>
> ### ASD NT ###
> mProb.AB_ASDNT.GJ = lm(TotalProb ~
+                           Group.LADref*aprime.log
+                 #+ age
+                       + vabs_soc_ss
+                         , data = dASDNT)
> summary(mProb.AB_ASDNT.GJ)

Call:
lm(formula = TotalProb ~ Group.LADref * aprime.log + vabs_soc_ss,
    data = dASDNT)

Residuals:
     Min       1Q   Median       3Q      Max
-21.8286  -5.7485   0.1479   7.7079  17.2007

Coefficients:
                        Estimate Std. Error t value Pr(>|t|)
(Intercept)              93.1321    11.3491   8.206 4.16e-10 ***
Group.LADref              0.2822     6.8166   0.041 0.967189
aprime.log               -0.2325    17.8280  -0.013 0.989660
vabs_soc_ss              -0.4419     0.1163  -3.800 0.000484 ***
Group.LADref:aprime.log  22.1718    35.6064   0.623 0.537022
---
Signif. codes:  0 ‘***’ 0.001 ‘**’ 0.01 ‘*’ 0.05 ‘.’ 0.1 ‘ ’ 1

Residual standard error: 10.26 on 40 degrees of freedom
  (32 observations deleted due to missingness)
Multiple R-squared:  0.4432,	Adjusted R-squared:  0.3876
F-statistic: 7.961 on 4 and 40 DF,  p-value: 8.08e-05

>
> mProb.AB_ASDNT.Words = lm(TotalProb ~
+                           Group.LADref*FirstWords.months
+                 #+ age
+                       + vabs_soc_ss
+                         , data = dASDNT)
> summary(mProb.AB_ASDNT.Words)

Call:
lm(formula = TotalProb ~ Group.LADref * FirstWords.months + vabs_soc_ss,
    data = dASDNT)

Residuals:
     Min       1Q   Median       3Q      Max
-18.8424  -5.7458  -0.6258   7.9786  17.6045

Coefficients:
                               Estimate Std. Error t value Pr(>|t|)
(Intercept)                    93.29861   13.10105   7.121 3.14e-08 ***
Group.LADref                    2.11823    8.56246   0.247  0.80610
FirstWords.months              -0.06924    0.27110  -0.255  0.79994
vabs_soc_ss                    -0.43689    0.12399  -3.524  0.00124 **
Group.LADref:FirstWords.months -0.41534    0.55526  -0.748  0.45960
---
Signif. codes:  0 ‘***’ 0.001 ‘**’ 0.01 ‘*’ 0.05 ‘.’ 0.1 ‘ ’ 1

Residual standard error: 9.997 on 34 degrees of freedom
  (38 observations deleted due to missingness)
Multiple R-squared:  0.5023,	Adjusted R-squared:  0.4437
F-statistic: 8.578 on 4 and 34 DF,  p-value: 6.73e-05

>
> mProb.AB_ASDNT.Phrases = lm(TotalProb ~
+                           Group.LADref*FirstPhrases.months
+                 #+ age
+                       + vabs_soc_ss
+                         , data = dASDNT)
> summary(mProb.AB_ASDNT.Phrases)

Call:
lm(formula = TotalProb ~ Group.LADref * FirstPhrases.months +
    vabs_soc_ss, data = dASDNT)

Residuals:
    Min      1Q  Median      3Q     Max
-19.710  -6.239  -1.065   7.697  17.066

Coefficients:
                                 Estimate Std. Error t value Pr(>|t|)
(Intercept)                       93.1911    13.0239   7.155 2.06e-08 ***
Group.LADref                      -0.5079    10.8757  -0.047  0.96301
FirstPhrases.months               -0.1061     0.2048  -0.518  0.60760
vabs_soc_ss                       -0.4207     0.1208  -3.484  0.00132 **
Group.LADref:FirstPhrases.months  -0.1412     0.4090  -0.345  0.73192
---
Signif. codes:  0 ‘***’ 0.001 ‘**’ 0.01 ‘*’ 0.05 ‘.’ 0.1 ‘ ’ 1

Residual standard error: 10.53 on 36 degrees of freedom
  (36 observations deleted due to missingness)
Multiple R-squared:  0.4208,	Adjusted R-squared:  0.3564
F-statistic: 6.538 on 4 and 36 DF,  p-value: 0.0004619

>
>
>
>
>
> ### LAD NT ###
> mProb.AB_LADNT.GJ = lm(TotalProb ~
+                           Group.ASDref*aprime.log
+                 #+ age
+                       + vabs_soc_ss
+                         , data = dLADNT)
> summary(mProb.AB_LADNT.GJ)

Call:
lm(formula = TotalProb ~ Group.ASDref * aprime.log + vabs_soc_ss,
    data = dLADNT)

Residuals:
     Min       1Q   Median       3Q      Max
-15.3945  -8.3395   0.3989   6.8262  21.1078

Coefficients:
                        Estimate Std. Error t value Pr(>|t|)
(Intercept)              89.6154    16.4658   5.443 2.68e-06 ***
Group.ASDref              2.9343     5.4642   0.537  0.59417
aprime.log               10.0669    18.9820   0.530  0.59874
vabs_soc_ss              -0.4210     0.1503  -2.802  0.00773 **
Group.ASDref:aprime.log   1.8546    38.5715   0.048  0.96188
---
Signif. codes:  0 ‘***’ 0.001 ‘**’ 0.01 ‘*’ 0.05 ‘.’ 0.1 ‘ ’ 1

Residual standard error: 9.599 on 41 degrees of freedom
  (24 observations deleted due to missingness)
Multiple R-squared:  0.183,	Adjusted R-squared:  0.1032
F-statistic: 2.295 on 4 and 41 DF,  p-value: 0.07548

>
> mProb.AB_LADNT.Words = lm(TotalProb ~
+                           Group.ASDref*FirstWords.months
+                 #+ age
+                       + vabs_soc_ss
+                         , data = dLADNT)
> summary(mProb.AB_LADNT.Words)

Call:
lm(formula = TotalProb ~ Group.ASDref * FirstWords.months + vabs_soc_ss,
    data = dLADNT)

Residuals:
     Min       1Q   Median       3Q      Max
-14.9707  -7.4852  -0.4294   6.2225  20.6496

Coefficients:
                               Estimate Std. Error t value Pr(>|t|)
(Intercept)                    85.11069   15.67682   5.429 3.72e-06 ***
Group.ASDref                    8.66999    8.38678   1.034   0.3080
FirstWords.months              -0.09879    0.25935  -0.381   0.7054
vabs_soc_ss                    -0.38838    0.14454  -2.687   0.0107 *
Group.ASDref:FirstWords.months -0.38643    0.52416  -0.737   0.4656
---
Signif. codes:  0 ‘***’ 0.001 ‘**’ 0.01 ‘*’ 0.05 ‘.’ 0.1 ‘ ’ 1

Residual standard error: 9.791 on 37 degrees of freedom
  (28 observations deleted due to missingness)
Multiple R-squared:  0.197,	Adjusted R-squared:  0.1102
F-statistic:  2.27 on 4 and 37 DF,  p-value: 0.08014

>
> mProb.AB_LADNT.Phrases = lm(TotalProb ~
+                           Group.ASDref*FirstPhrases.months
+                 #+ age
+                       + vabs_soc_ss
+                         , data = dLADNT)
> summary(mProb.AB_LADNT.Phrases)

Call:
lm(formula = TotalProb ~ Group.ASDref * FirstPhrases.months +
    vabs_soc_ss, data = dLADNT)

Residuals:
    Min      1Q  Median      3Q     Max
-15.634  -6.178   0.090   6.173  19.037

Coefficients:
                                 Estimate Std. Error t value Pr(>|t|)
(Intercept)                      81.46030   14.85848   5.482 2.92e-06 ***
Group.ASDref                     12.91010    7.44644   1.734  0.09107 .
FirstPhrases.months               0.02898    0.15576   0.186  0.85342
vabs_soc_ss                      -0.37342    0.13725  -2.721  0.00977 **
Group.ASDref:FirstPhrases.months -0.40838    0.31146  -1.311  0.19766
---
Signif. codes:  0 ‘***’ 0.001 ‘**’ 0.01 ‘*’ 0.05 ‘.’ 0.1 ‘ ’ 1

Residual standard error: 9.421 on 38 degrees of freedom
  (27 observations deleted due to missingness)
Multiple R-squared:  0.2244,	Adjusted R-squared:  0.1427
F-statistic: 2.748 on 4 and 38 DF,  p-value: 0.04213


> ###Follow up
> #aprime.log
> mProb.AB_ASD.GJ = lm(TotalProb ~ aprime.log
+                 #+ age
+                       + vabs_soc_ss
+                         , data = dASD)
> summary(mProb.AB_ASD.GJ)

Call:
lm(formula = TotalProb ~ aprime.log + vabs_soc_ss, data = dASD)

Residuals:
    Min      1Q  Median      3Q     Max
-22.381  -5.703   1.280   6.885  17.073

Coefficients:
            Estimate Std. Error t value Pr(>|t|)
(Intercept)  95.9141    13.7684   6.966 2.28e-06 ***
aprime.log  -11.6615    23.5599  -0.495   0.6270
vabs_soc_ss  -0.4773     0.1546  -3.086   0.0067 **
---
Signif. codes:  0 ‘***’ 0.001 ‘**’ 0.01 ‘*’ 0.05 ‘.’ 0.1 ‘ ’ 1

Residual standard error: 11.65 on 17 degrees of freedom
  (19 observations deleted due to missingness)
Multiple R-squared:  0.3613,	Adjusted R-squared:  0.2861
F-statistic: 4.807 on 2 and 17 DF,  p-value: 0.02215

>
> mProb.AB_NT.GJ = lm(TotalProb ~ aprime.log
+                 #+ age
+                       + vabs_soc_ss
+                         , data = dNT)
> summary(mProb.AB_NT.GJ)

Call:
lm(formula = TotalProb ~ aprime.log + vabs_soc_ss, data = dNT)

Residuals:
     Min       1Q   Median       3Q      Max
-14.7885  -5.3749   0.1877   8.0114  12.9427

Coefficients:
            Estimate Std. Error t value Pr(>|t|)
(Intercept)  83.3213    21.4533   3.884   0.0008 ***
aprime.log   11.4930    26.1227   0.440   0.6643
vabs_soc_ss  -0.3470     0.2011  -1.726   0.0984 .
---
Signif. codes:  0 ‘***’ 0.001 ‘**’ 0.01 ‘*’ 0.05 ‘.’ 0.1 ‘ ’ 1

Residual standard error: 9.247 on 22 degrees of freedom
  (13 observations deleted due to missingness)
Multiple R-squared:  0.129,	Adjusted R-squared:  0.04983
F-statistic: 1.629 on 2 and 22 DF,  p-value: 0.2188

>
> mProb.AB_LAD.GJ = lm(TotalProb ~ aprime.log
+                 #+ age
+                       + vabs_soc_ss
+                         , data = dLAD)
> summary(mProb.AB_LAD.GJ)

Call:
lm(formula = TotalProb ~ aprime.log + vabs_soc_ss, data = dLAD)

Residuals:
     Min       1Q   Median       3Q      Max
-14.3873  -8.1600   0.1624   4.1108  20.8361

Coefficients:
            Estimate Std. Error t value Pr(>|t|)
(Intercept)  97.0173    25.8803   3.749  0.00147 **
aprime.log   15.2231    31.3750   0.485  0.63339
vabs_soc_ss  -0.5006     0.2300  -2.176  0.04310 *
---
Signif. codes:  0 ‘***’ 0.001 ‘**’ 0.01 ‘*’ 0.05 ‘.’ 0.1 ‘ ’ 1

Residual standard error: 10.2 on 18 degrees of freedom
  (11 observations deleted due to missingness)
Multiple R-squared:  0.2347,	Adjusted R-squared:  0.1497
F-statistic:  2.76 on 2 and 18 DF,  p-value: 0.09005

>
> #Words
> mProb.AB_ASD.Words = lm(TotalProb ~ FirstWords.months
+                 #+ age
+                       + vabs_soc_ss
+                         , data = dASD)
> summary(mProb.AB_ASD.Words)

Call:
lm(formula = TotalProb ~ FirstWords.months + vabs_soc_ss, data = dASD)

Residuals:
    Min      1Q  Median      3Q     Max
-18.127  -6.329  -3.528   9.430  17.168

Coefficients:
                  Estimate Std. Error t value Pr(>|t|)
(Intercept)        95.6805    17.6039   5.435 0.000114 ***
FirstWords.months   0.1123     0.3028   0.371 0.716814
vabs_soc_ss        -0.4718     0.1676  -2.815 0.014609 *
---
Signif. codes:  0 ‘***’ 0.001 ‘**’ 0.01 ‘*’ 0.05 ‘.’ 0.1 ‘ ’ 1

Residual standard error: 11.22 on 13 degrees of freedom
  (23 observations deleted due to missingness)
Multiple R-squared:  0.4533,	Adjusted R-squared:  0.3692
F-statistic:  5.39 on 2 and 13 DF,  p-value: 0.01973

>
> mProb.AB_NT.Words = lm(TotalProb ~ FirstWords.months
+                 #+ age
+                       + vabs_soc_ss
+                         , data = dNT)
> summary(mProb.AB_NT.Words)

Call:
lm(formula = TotalProb ~ FirstWords.months + vabs_soc_ss, data = dNT)

Residuals:
     Min       1Q   Median       3Q      Max
-14.5600  -5.3664  -0.5707   6.8811  13.7122

Coefficients:
                  Estimate Std. Error t value Pr(>|t|)
(Intercept)        86.4953    21.9267   3.945 0.000801 ***
FirstWords.months  -0.3011     0.4532  -0.664 0.514074
vabs_soc_ss        -0.3592     0.2078  -1.729 0.099302 .
---
Signif. codes:  0 ‘***’ 0.001 ‘**’ 0.01 ‘*’ 0.05 ‘.’ 0.1 ‘ ’ 1

Residual standard error: 9.334 on 20 degrees of freedom
  (15 observations deleted due to missingness)
Multiple R-squared:  0.1609,	Adjusted R-squared:  0.07698
F-statistic: 1.917 on 2 and 20 DF,  p-value: 0.1731

>
> mProb.AB_LAD.Words = lm(TotalProb ~ FirstWords.months
+                 #+ age
+                       + vabs_soc_ss
+                         , data = dLAD)
> summary(mProb.AB_LAD.Words)

Call:
lm(formula = TotalProb ~ FirstWords.months + vabs_soc_ss, data = dLAD)

Residuals:
    Min      1Q  Median      3Q     Max
-13.168  -7.359  -0.347   3.110  20.771

Coefficients:
                  Estimate Std. Error t value Pr(>|t|)
(Intercept)       83.24150   23.27885   3.576  0.00252 **
FirstWords.months  0.08943    0.24053   0.372  0.71493
vabs_soc_ss       -0.41124    0.20925  -1.965  0.06699 .
---
Signif. codes:  0 ‘***’ 0.001 ‘**’ 0.01 ‘*’ 0.05 ‘.’ 0.1 ‘ ’ 1

Residual standard error: 10.61 on 16 degrees of freedom
  (13 observations deleted due to missingness)
Multiple R-squared:  0.2172,	Adjusted R-squared:  0.1193
F-statistic:  2.22 on 2 and 16 DF,  p-value: 0.141

>
> #Phrases
> mProb.AB_ASD.Phrases = lm(TotalProb ~ FirstPhrases.months
+                 #+ age
+                       + vabs_soc_ss
+                         , data = dASD)
> summary(mProb.AB_ASD.Phrases)

Call:
lm(formula = TotalProb ~ FirstPhrases.months + vabs_soc_ss, data = dASD)

Residuals:
    Min      1Q  Median      3Q     Max
-19.940  -6.155  -3.712  10.566  16.957

Coefficients:
                    Estimate Std. Error t value Pr(>|t|)
(Intercept)         95.58066   18.02964   5.301 0.000112 ***
FirstPhrases.months -0.04125    0.31099  -0.133 0.896355
vabs_soc_ss         -0.44435    0.16557  -2.684 0.017815 *
---
Signif. codes:  0 ‘***’ 0.001 ‘**’ 0.01 ‘*’ 0.05 ‘.’ 0.1 ‘ ’ 1

Residual standard error: 12.31 on 14 degrees of freedom
  (22 observations deleted due to missingness)
Multiple R-squared:  0.3412,	Adjusted R-squared:  0.2471
F-statistic: 3.625 on 2 and 14 DF,  p-value: 0.05387

>
> mProb.AB_NT.Phrases = lm(TotalProb ~ FirstPhrases.months
+                 #+ age
+                       + vabs_soc_ss
+                         , data = dNT)
> summary(mProb.AB_NT.Phrases)

Call:
lm(formula = TotalProb ~ FirstPhrases.months + vabs_soc_ss, data = dNT)

Residuals:
     Min       1Q   Median       3Q      Max
-15.7057  -6.1559  -0.4702   7.9144  11.9790

Coefficients:
                    Estimate Std. Error t value Pr(>|t|)
(Intercept)          86.2828    22.6453   3.810  0.00102 **
FirstPhrases.months  -0.1747     0.2785  -0.627  0.53712
vabs_soc_ss          -0.3580     0.2065  -1.734  0.09760 .
---
Signif. codes:  0 ‘***’ 0.001 ‘**’ 0.01 ‘*’ 0.05 ‘.’ 0.1 ‘ ’ 1

Residual standard error: 9.415 on 21 degrees of freedom
  (14 observations deleted due to missingness)
Multiple R-squared:  0.1376,	Adjusted R-squared:  0.05551
F-statistic: 1.676 on 2 and 21 DF,  p-value: 0.2112

>
> mProb.AB_LAD.Phrases = lm(TotalProb ~ FirstPhrases.months
+                 #+ age
+                       + vabs_soc_ss
+                         , data = dLAD)
> summary(mProb.AB_LAD.Phrases)

Call:
lm(formula = TotalProb ~ FirstPhrases.months + vabs_soc_ss, data = dLAD)

Residuals:
     Min       1Q   Median       3Q      Max
-13.4412  -6.1825  -0.2734   1.8357  19.0526

Coefficients:
                    Estimate Std. Error t value Pr(>|t|)
(Intercept)          76.2629    20.2038   3.775  0.00166 **
FirstPhrases.months   0.2329     0.1436   1.622  0.12444
vabs_soc_ss          -0.3856     0.1893  -2.036  0.05863 .
---
Signif. codes:  0 ‘***’ 0.001 ‘**’ 0.01 ‘*’ 0.05 ‘.’ 0.1 ‘ ’ 1

Residual standard error: 9.716 on 16 degrees of freedom
  (13 observations deleted due to missingness)
Multiple R-squared:  0.3027,	Adjusted R-squared:  0.2156
F-statistic: 3.474 on 2 and 16 DF,  p-value: 0.05587


> ### CELF ###
>
> #Core Language "cls_std_scr",
> mProb_ASD.CL = lm(cbcl_totalprob_t ~ cls_std_scr
+                       #+ age
+                       + vabs_soc_ss
+                         , data = dASD.inperson)
> summary(mProb_ASD.CL)

Call:
lm(formula = cbcl_totalprob_t ~ cls_std_scr + vabs_soc_ss, data = dASD.inperson)

Residuals:
   Min     1Q Median     3Q    Max
-8.359 -4.042 -1.420  4.913 12.592

Coefficients:
            Estimate Std. Error t value Pr(>|t|)
(Intercept) 114.4522    20.2380   5.655  0.00077 ***
cls_std_scr  -0.1977     0.1760  -1.124  0.29826
vabs_soc_ss  -0.4196     0.1522  -2.757  0.02822 *
---
Signif. codes:  0 ‘***’ 0.001 ‘**’ 0.01 ‘*’ 0.05 ‘.’ 0.1 ‘ ’ 1

Residual standard error: 7.597 on 7 degrees of freedom
  (8 observations deleted due to missingness)
Multiple R-squared:  0.5959,	Adjusted R-squared:  0.4804
F-statistic: 5.161 on 2 and 7 DF,  p-value: 0.04195

>
> mProb_LAD.CL = lm(cbcl_totalprob_t ~ cls_std_scr
+                       #+ age
+                       + vabs_soc_ss
+                         , data = dLAD.inperson)
> summary(mProb_LAD.CL)

Call:
lm(formula = cbcl_totalprob_t ~ cls_std_scr + vabs_soc_ss, data = dLAD.inperson)

Residuals:
ALL 3 residuals are 0: no residual degrees of freedom!

Coefficients:
            Estimate Std. Error t value Pr(>|t|)
(Intercept) 142.5206        NaN     NaN      NaN
cls_std_scr  -1.0258        NaN     NaN      NaN
vabs_soc_ss   0.2113        NaN     NaN      NaN

Residual standard error: NaN on 0 degrees of freedom
  (2 observations deleted due to missingness)
Multiple R-squared:      1,	Adjusted R-squared:    NaN
F-statistic:   NaN on 2 and 0 DF,  p-value: NA

>
> mProb_NT.CL = lm(cbcl_totalprob_t ~ cls_std_scr
+                       #+ age
+                       + vabs_soc_ss
+                         , data = dNT.inperson)
> summary(mProb_NT.CL)

Call:
lm(formula = cbcl_totalprob_t ~ cls_std_scr + vabs_soc_ss, data = dNT.inperson)

Residuals:
     1      2      3      5      7      9     10
 7.995 -1.209 -6.328  1.394 -2.832  4.781 -3.801

Coefficients:
             Estimate Std. Error t value Pr(>|t|)
(Intercept) 101.38621   66.61476   1.522    0.203
cls_std_scr   0.06499    0.32854   0.198    0.853
vabs_soc_ss  -0.53689    0.31705  -1.693    0.166

Residual standard error: 6.179 on 4 degrees of freedom
  (5 observations deleted due to missingness)
Multiple R-squared:  0.6482,	Adjusted R-squared:  0.4722
F-statistic: 3.684 on 2 and 4 DF,  p-value: 0.1238

>
> #Recalling Sentences "rs_ss"
> mProb_ASD.RS = lm(cbcl_totalprob_t ~ rs_ss
+                       #+ age
+                       + vabs_soc_ss
+                         , data = dASD.inperson)
> summary(mProb_ASD.RS)

Call:
lm(formula = cbcl_totalprob_t ~ rs_ss + vabs_soc_ss, data = dASD.inperson)

Residuals:
   Min     1Q Median     3Q    Max
-8.130 -3.069 -2.013  3.418 13.415

Coefficients:
            Estimate Std. Error t value Pr(>|t|)
(Intercept)  94.7443    11.6303   8.146 8.12e-05 ***
rs_ss        -1.1706     0.9340  -1.253    0.250
vabs_soc_ss  -0.2571     0.2131  -1.206    0.267
---
Signif. codes:  0 ‘***’ 0.001 ‘**’ 0.01 ‘*’ 0.05 ‘.’ 0.1 ‘ ’ 1

Residual standard error: 7.459 on 7 degrees of freedom
  (8 observations deleted due to missingness)
Multiple R-squared:  0.6104,	Adjusted R-squared:  0.4991
F-statistic: 5.484 on 2 and 7 DF,  p-value: 0.0369

>
> mProb_LAD.RS = lm(cbcl_totalprob_t ~ rs_ss
+                       #+ age
+                       + vabs_soc_ss
+                         , data = dLAD.inperson)
> summary(mProb_LAD.RS)

Call:
lm(formula = cbcl_totalprob_t ~ rs_ss + vabs_soc_ss, data = dLAD.inperson)

Residuals:
ALL 3 residuals are 0: no residual degrees of freedom!

Coefficients:
            Estimate Std. Error t value Pr(>|t|)
(Intercept)  59.5323        NaN     NaN      NaN
rs_ss        -1.6048        NaN     NaN      NaN
vabs_soc_ss   0.1452        NaN     NaN      NaN

Residual standard error: NaN on 0 degrees of freedom
  (2 observations deleted due to missingness)
Multiple R-squared:      1,	Adjusted R-squared:    NaN
F-statistic:   NaN on 2 and 0 DF,  p-value: NA

>
> mProb_NT.RS = lm(cbcl_totalprob_t ~ rs_ss
+                       #+ age
+                       + vabs_soc_ss
+                         , data = dNT.inperson)
> summary(mProb_NT.RS)

Call:
lm(formula = cbcl_totalprob_t ~ rs_ss + vabs_soc_ss, data = dNT.inperson)

Residuals:
      1       2       3       5       7       9      10
 7.8443 -1.0038 -6.7259  0.8879 -2.0686  5.0444 -3.9782

Coefficients:
             Estimate Std. Error t value Pr(>|t|)
(Intercept) 112.85852   30.52800   3.697   0.0209 *
rs_ss         0.05983    1.35401   0.044   0.9669
vabs_soc_ss  -0.58159    0.21843  -2.663   0.0562 .
---
Signif. codes:  0 ‘***’ 0.001 ‘**’ 0.01 ‘*’ 0.05 ‘.’ 0.1 ‘ ’ 1

Residual standard error: 6.207 on 4 degrees of freedom
  (5 observations deleted due to missingness)
Multiple R-squared:  0.6449,	Adjusted R-squared:  0.4673
F-statistic: 3.632 on 2 and 4 DF,  p-value: 0.1261


> ###################
> ##### AnxDep ######
> ###################
> #combined
>
> ### ASD LAD ###
> mAnxDep.AB_ASDLAD.GJ = lm(AnxDep ~
+                           Group.NTref*aprime.log
+                  #+ age
+                       + vabs_soc_ss
+                         , data = dASDLAD)
> summary(mAnxDep.AB_ASDLAD.GJ)

Call:
lm(formula = AnxDep ~ Group.NTref * aprime.log + vabs_soc_ss,
    data = dASDLAD)

Residuals:
     Min       1Q   Median       3Q      Max
-18.8913  -3.8456  -0.9767   3.2065  17.4488

Coefficients:
                       Estimate Std. Error t value Pr(>|t|)
(Intercept)            80.22124    9.72107   8.252    1e-09 ***
Group.NTref            -0.41614    5.40644  -0.077   0.9391
aprime.log             -2.05454   14.16534  -0.145   0.8855
vabs_soc_ss            -0.23843    0.09657  -2.469   0.0186 *
Group.NTref:aprime.log 43.66871   28.18153   1.550   0.1302
---
Signif. codes:  0 ‘***’ 0.001 ‘**’ 0.01 ‘*’ 0.05 ‘.’ 0.1 ‘ ’ 1

Residual standard error: 8.228 on 35 degrees of freedom
  (31 observations deleted due to missingness)
Multiple R-squared:  0.4424,	Adjusted R-squared:  0.3787
F-statistic: 6.942 on 4 and 35 DF,  p-value: 0.0003179

>
> mAnxDep.AB_ASDLAD.Words = lm(AnxDep ~
+                           Group.NTref*FirstWords.months
+                  #+ age
+                       + vabs_soc_ss
+                         , data = dASDLAD)
> summary(mAnxDep.AB_ASDLAD.Words)

Call:
lm(formula = AnxDep ~ Group.NTref * FirstWords.months + vabs_soc_ss,
    data = dASDLAD)

Residuals:
     Min       1Q   Median       3Q      Max
-18.8138  -4.0721  -0.4901   3.1467  16.5316

Coefficients:
                               Estimate Std. Error t value Pr(>|t|)
(Intercept)                   72.813687  10.265375   7.093  6.9e-08 ***
Group.NTref                   -7.767308   6.161419  -1.261   0.2172
FirstWords.months              0.156650   0.139956   1.119   0.2719
vabs_soc_ss                   -0.193429   0.094997  -2.036   0.0506 .
Group.NTref:FirstWords.months  0.001951   0.269140   0.007   0.9943
---
Signif. codes:  0 ‘***’ 0.001 ‘**’ 0.01 ‘*’ 0.05 ‘.’ 0.1 ‘ ’ 1

Residual standard error: 7.979 on 30 degrees of freedom
  (36 observations deleted due to missingness)
Multiple R-squared:  0.4516,	Adjusted R-squared:  0.3785
F-statistic: 6.176 on 4 and 30 DF,  p-value: 0.0009489

>
> mAnxDep.AB_ASDLAD.Phrases = lm(AnxDep ~
+                           Group.NTref*FirstPhrases.months
+                  #+ age
+                       + vabs_soc_ss
+                         , data = dASDLAD)
> summary(mAnxDep.AB_ASDLAD.Phrases)

Call:
lm(formula = AnxDep ~ Group.NTref * FirstPhrases.months + vabs_soc_ss,
    data = dASDLAD)

Residuals:
    Min      1Q  Median      3Q     Max
-16.096  -3.966  -1.227   3.138  17.218

Coefficients:
                                 Estimate Std. Error t value Pr(>|t|)
(Intercept)                     74.682354  10.014746   7.457 2.12e-08 ***
Group.NTref                     -6.311623   8.250480  -0.765   0.4501
FirstPhrases.months              0.035063   0.124485   0.282   0.7801
vabs_soc_ss                     -0.195512   0.094286  -2.074   0.0465 *
Group.NTref:FirstPhrases.months -0.002705   0.248635  -0.011   0.9914
---
Signif. codes:  0 ‘***’ 0.001 ‘**’ 0.01 ‘*’ 0.05 ‘.’ 0.1 ‘ ’ 1

Residual standard error: 8.516 on 31 degrees of freedom
  (35 observations deleted due to missingness)
Multiple R-squared:  0.3645,	Adjusted R-squared:  0.2825
F-statistic: 4.445 on 4 and 31 DF,  p-value: 0.005902

>
>
>
>
>
> ### ASD NT ###
> mAnxDep.AB_ASDNT.GJ = lm(AnxDep ~
+                           Group.LADref*aprime.log
+                 #+ age
+                       + vabs_soc_ss
+                         , data = dASDNT)
> summary(mAnxDep.AB_ASDNT.GJ)

Call:
lm(formula = AnxDep ~ Group.LADref * aprime.log + vabs_soc_ss,
    data = dASDNT)

Residuals:
     Min       1Q   Median       3Q      Max
-18.6707  -4.8407  -0.4955   3.9366  20.5967

Coefficients:
                        Estimate Std. Error t value Pr(>|t|)
(Intercept)             79.75038    9.04803   8.814 6.45e-11 ***
Group.LADref             1.22458    5.43692   0.225   0.8229
aprime.log              -0.50668   14.32048  -0.035   0.9720
vabs_soc_ss             -0.22305    0.09167  -2.433   0.0195 *
Group.LADref:aprime.log 46.85541   28.63123   1.637   0.1096
---
Signif. codes:  0 ‘***’ 0.001 ‘**’ 0.01 ‘*’ 0.05 ‘.’ 0.1 ‘ ’ 1

Residual standard error: 8.198 on 40 degrees of freedom
  (32 observations deleted due to missingness)
Multiple R-squared:  0.4167,	Adjusted R-squared:  0.3584
F-statistic: 7.145 on 4 and 40 DF,  p-value: 0.0001938

>
> mAnxDep.AB_ASDNT.Words = lm(AnxDep ~
+                           Group.LADref*FirstWords.months
+                 #+ age
+                       + vabs_soc_ss
+                         , data = dASDNT)
> summary(mAnxDep.AB_ASDNT.Words)

Call:
lm(formula = AnxDep ~ Group.LADref * FirstWords.months + vabs_soc_ss,
    data = dASDNT)

Residuals:
    Min      1Q  Median      3Q     Max
-19.114  -4.894  -1.098   3.491  19.009

Coefficients:
                               Estimate Std. Error t value Pr(>|t|)
(Intercept)                    81.16510   10.43936   7.775 3.93e-09 ***
Group.LADref                    2.37984    6.99131   0.340   0.7356
FirstWords.months              -0.11085    0.21951  -0.505   0.6168
vabs_soc_ss                    -0.22671    0.09731  -2.330   0.0257 *
Group.LADref:FirstWords.months -0.48318    0.44039  -1.097   0.2801
---
Signif. codes:  0 ‘***’ 0.001 ‘**’ 0.01 ‘*’ 0.05 ‘.’ 0.1 ‘ ’ 1

Residual standard error: 8.204 on 35 degrees of freedom
  (37 observations deleted due to missingness)
Multiple R-squared:  0.3802,	Adjusted R-squared:  0.3094
F-statistic: 5.368 on 4 and 35 DF,  p-value: 0.00177

>
> mAnxDep.AB_ASDNT.Phrases = lm(AnxDep ~
+                           Group.LADref*FirstPhrases.months
+                 #+ age
+                       + vabs_soc_ss
+                         , data = dASDNT)
> summary(mAnxDep.AB_ASDNT.Phrases)

Call:
lm(formula = AnxDep ~ Group.LADref * FirstPhrases.months + vabs_soc_ss,
    data = dASDNT)

Residuals:
    Min      1Q  Median      3Q     Max
-16.314  -4.639  -1.001   3.025  19.194

Coefficients:
                                 Estimate Std. Error t value Pr(>|t|)
(Intercept)                      81.04870   10.02107   8.088 1.06e-09 ***
Group.LADref                      2.35971    8.63483   0.273   0.7862
FirstPhrases.months              -0.12657    0.16328  -0.775   0.4432
vabs_soc_ss                      -0.21795    0.09297  -2.344   0.0245 *
Group.LADref:FirstPhrases.months -0.31498    0.32653  -0.965   0.3410
---
Signif. codes:  0 ‘***’ 0.001 ‘**’ 0.01 ‘*’ 0.05 ‘.’ 0.1 ‘ ’ 1

Residual standard error: 8.409 on 37 degrees of freedom
  (35 observations deleted due to missingness)
Multiple R-squared:  0.3279,	Adjusted R-squared:  0.2553
F-statistic: 4.513 on 4 and 37 DF,  p-value: 0.004534

>
>
>
>
>
> ### LAD NT ###
> mAnxDep.AB_LADNT.GJ = lm(AnxDep ~
+                           Group.ASDref*aprime.log
+                 #+ age
+                       + vabs_soc_ss
+                         , data = dLADNT)
> summary(mAnxDep.AB_LADNT.GJ)

Call:
lm(formula = AnxDep ~ Group.ASDref * aprime.log + vabs_soc_ss,
    data = dLADNT)

Residuals:
    Min      1Q  Median      3Q     Max
-8.2462 -2.8738 -0.8438  1.7546 20.7011

Coefficients:
                        Estimate Std. Error t value Pr(>|t|)
(Intercept)             76.48905    8.63959   8.853 3.73e-11 ***
Group.ASDref             2.21377    3.03278   0.730   0.4695
aprime.log              19.68875   10.59998   1.857   0.0703 .
vabs_soc_ss             -0.19684    0.07844  -2.509   0.0160 *
Group.ASDref:aprime.log  6.17093   20.95008   0.295   0.7698
---
Signif. codes:  0 ‘***’ 0.001 ‘**’ 0.01 ‘*’ 0.05 ‘.’ 0.1 ‘ ’ 1

Residual standard error: 5.361 on 42 degrees of freedom
  (23 observations deleted due to missingness)
Multiple R-squared:  0.1693,	Adjusted R-squared:  0.09016
F-statistic:  2.14 on 4 and 42 DF,  p-value: 0.09269

>
> mAnxDep.AB_LADNT.Words = lm(AnxDep ~
+                           Group.ASDref*FirstWords.months
+                 #+ age
+                       + vabs_soc_ss
+                         , data = dLADNT)
> summary(mAnxDep.AB_LADNT.Words)

Call:
lm(formula = AnxDep ~ Group.ASDref * FirstWords.months + vabs_soc_ss,
    data = dLADNT)

Residuals:
    Min      1Q  Median      3Q     Max
-6.2014 -2.8357 -0.9443  1.3128 19.3369

Coefficients:
                               Estimate Std. Error t value Pr(>|t|)
(Intercept)                    65.99362    8.02939   8.219 5.89e-10 ***
Group.ASDref                   10.28285    4.45214   2.310   0.0264 *
FirstWords.months              -0.09150    0.13637  -0.671   0.5063
vabs_soc_ss                    -0.11854    0.07293  -1.625   0.1123
Group.ASDref:FirstWords.months -0.53100    0.27309  -1.944   0.0593 .
---
Signif. codes:  0 ‘***’ 0.001 ‘**’ 0.01 ‘*’ 0.05 ‘.’ 0.1 ‘ ’ 1

Residual standard error: 5.257 on 38 degrees of freedom
  (27 observations deleted due to missingness)
Multiple R-squared:  0.1869,	Adjusted R-squared:  0.1013
F-statistic: 2.184 on 4 and 38 DF,  p-value: 0.08928

>
> mAnxDep.AB_LADNT.Phrases = lm(AnxDep ~
+                           Group.ASDref*FirstPhrases.months
+                 #+ age
+                       + vabs_soc_ss
+                         , data = dLADNT)
> summary(mAnxDep.AB_LADNT.Phrases)

Call:
lm(formula = AnxDep ~ Group.ASDref * FirstPhrases.months + vabs_soc_ss,
    data = dLADNT)

Residuals:
    Min      1Q  Median      3Q     Max
-7.5036 -2.1138 -1.2007  0.6769 19.4881

Coefficients:
                                 Estimate Std. Error t value Pr(>|t|)
(Intercept)                      69.38002    7.78326   8.914 5.98e-11 ***
Group.ASDref                      8.29160    4.12853   2.008   0.0516 .
FirstPhrases.months              -0.12448    0.08665  -1.436   0.1588
vabs_soc_ss                      -0.13473    0.07212  -1.868   0.0693 .
Group.ASDref:FirstPhrases.months -0.31870    0.17330  -1.839   0.0735 .
---
Signif. codes:  0 ‘***’ 0.001 ‘**’ 0.01 ‘*’ 0.05 ‘.’ 0.1 ‘ ’ 1

Residual standard error: 5.246 on 39 degrees of freedom
  (26 observations deleted due to missingness)
Multiple R-squared:  0.1702,	Adjusted R-squared:  0.08505
F-statistic: 1.999 on 4 and 39 DF,  p-value: 0.1137


> ###Follow up
> #aprime.log
> mAnxDep.AB_ASD.GJ = lm(AnxDep ~ aprime.log
+                 #+ age
+                       + vabs_soc_ss
+                         , data = dASD)
> summary(mAnxDep.AB_ASD.GJ)

Call:
lm(formula = AnxDep ~ aprime.log + vabs_soc_ss, data = dASD)

Residuals:
    Min      1Q  Median      3Q     Max
-18.988  -8.224   1.084   8.659  17.518

Coefficients:
            Estimate Std. Error t value Pr(>|t|)
(Intercept)  80.9944    13.5570   5.974 1.94e-05 ***
aprime.log  -23.8690    23.2394  -1.027    0.320
vabs_soc_ss  -0.2452     0.1506  -1.627    0.123
---
Signif. codes:  0 ‘***’ 0.001 ‘**’ 0.01 ‘*’ 0.05 ‘.’ 0.1 ‘ ’ 1

Residual standard error: 10.96 on 16 degrees of freedom
  (20 observations deleted due to missingness)
Multiple R-squared:  0.1906,	Adjusted R-squared:  0.08947
F-statistic: 1.884 on 2 and 16 DF,  p-value: 0.1841

>
> mAnxDep.AB_NT.GJ = lm(AnxDep ~ aprime.log
+                 #+ age
+                       + vabs_soc_ss
+                         , data = dNT)
> summary(mAnxDep.AB_NT.GJ)

Call:
lm(formula = AnxDep ~ aprime.log + vabs_soc_ss, data = dNT)

Residuals:
    Min      1Q  Median      3Q     Max
-6.6849 -3.3495 -0.8324  1.6093 20.7689

Coefficients:
            Estimate Std. Error t value Pr(>|t|)
(Intercept)  75.7968    11.9143   6.362 1.72e-06 ***
aprime.log   22.6787    15.9530   1.422    0.169
vabs_soc_ss  -0.1798     0.1104  -1.628    0.117
---
Signif. codes:  0 ‘***’ 0.001 ‘**’ 0.01 ‘*’ 0.05 ‘.’ 0.1 ‘ ’ 1

Residual standard error: 5.744 on 23 degrees of freedom
  (12 observations deleted due to missingness)
Multiple R-squared:  0.1636,	Adjusted R-squared:  0.09089
F-statistic:  2.25 on 2 and 23 DF,  p-value: 0.1281

>
> mAnxDep.AB_LAD.GJ = lm(AnxDep ~ aprime.log
+                 #+ age
+                       + vabs_soc_ss
+                         , data = dLAD)
> summary(mAnxDep.AB_LAD.GJ)

Call:
lm(formula = AnxDep ~ aprime.log + vabs_soc_ss, data = dLAD)

Residuals:
   Min     1Q Median     3Q    Max
-8.845 -2.471 -1.575  1.630 13.867

Coefficients:
            Estimate Std. Error t value Pr(>|t|)
(Intercept)  77.9948    12.6384   6.171 7.95e-06 ***
aprime.log   18.3953    15.3217   1.201   0.2455
vabs_soc_ss  -0.2203     0.1123  -1.961   0.0655 .
---
Signif. codes:  0 ‘***’ 0.001 ‘**’ 0.01 ‘*’ 0.05 ‘.’ 0.1 ‘ ’ 1

Residual standard error: 4.981 on 18 degrees of freedom
  (11 observations deleted due to missingness)
Multiple R-squared:  0.1766,	Adjusted R-squared:  0.08512
F-statistic:  1.93 on 2 and 18 DF,  p-value: 0.174

>
> #Words
> mAnxDep.AB_ASD.Words = lm(AnxDep ~ FirstWords.months
+                 #+ age
+                       + vabs_soc_ss
+                         , data = dASD)
> summary(mAnxDep.AB_ASD.Words)

Call:
lm(formula = AnxDep ~ FirstWords.months + vabs_soc_ss, data = dASD)

Residuals:
    Min      1Q  Median      3Q     Max
-19.444  -6.868  -3.527   7.769  15.659

Coefficients:
                  Estimate Std. Error t value Pr(>|t|)
(Intercept)        83.5691    17.2662   4.840 0.000323 ***
FirstWords.months   0.1034     0.2970   0.348 0.733270
vabs_soc_ss        -0.2632     0.1644  -1.601 0.133414
---
Signif. codes:  0 ‘***’ 0.001 ‘**’ 0.01 ‘*’ 0.05 ‘.’ 0.1 ‘ ’ 1

Residual standard error: 11.01 on 13 degrees of freedom
  (23 observations deleted due to missingness)
Multiple R-squared:  0.2262,	Adjusted R-squared:  0.1071
F-statistic:   1.9 on 2 and 13 DF,  p-value: 0.1888

>
> mAnxDep.AB_NT.Words = lm(AnxDep ~ FirstWords.months
+                 #+ age
+                       + vabs_soc_ss
+                         , data = dNT)
> summary(mAnxDep.AB_NT.Words)

Call:
lm(formula = AnxDep ~ FirstWords.months + vabs_soc_ss, data = dNT)

Residuals:
   Min     1Q Median     3Q    Max
-7.215 -3.547 -1.634  2.777 19.198

Coefficients:
                  Estimate Std. Error t value Pr(>|t|)
(Intercept)        75.8884    12.8352   5.913 7.21e-06 ***
FirstWords.months  -0.3551     0.2824  -1.257    0.222
vabs_soc_ss        -0.1644     0.1178  -1.396    0.177
---
Signif. codes:  0 ‘***’ 0.001 ‘**’ 0.01 ‘*’ 0.05 ‘.’ 0.1 ‘ ’ 1

Residual standard error: 6.033 on 21 degrees of freedom
  (14 observations deleted due to missingness)
Multiple R-squared:  0.1461,	Adjusted R-squared:  0.06473
F-statistic: 1.796 on 2 and 21 DF,  p-value: 0.1905

>
> mAnxDep.AB_LAD.Words = lm(AnxDep ~ FirstWords.months
+                 #+ age
+                       + vabs_soc_ss
+                         , data = dLAD)
> summary(mAnxDep.AB_LAD.Words)

Call:
lm(formula = AnxDep ~ FirstWords.months + vabs_soc_ss, data = dLAD)

Residuals:
    Min      1Q  Median      3Q     Max
-6.7549 -1.9327 -0.3380  0.9641 11.9198

Coefficients:
                  Estimate Std. Error t value Pr(>|t|)
(Intercept)       55.80755    9.09015   6.139 1.43e-05 ***
FirstWords.months  0.18423    0.09392   1.961   0.0675 .
vabs_soc_ss       -0.07178    0.08171  -0.878   0.3927
---
Signif. codes:  0 ‘***’ 0.001 ‘**’ 0.01 ‘*’ 0.05 ‘.’ 0.1 ‘ ’ 1

Residual standard error: 4.143 on 16 degrees of freedom
  (13 observations deleted due to missingness)
Multiple R-squared:  0.2549,	Adjusted R-squared:  0.1617
F-statistic: 2.736 on 2 and 16 DF,  p-value: 0.09502

>
> #Phrases
> mAnxDep.AB_ASD.Phrases = lm(AnxDep ~ FirstPhrases.months
+                 #+ age
+                       + vabs_soc_ss
+                         , data = dASD)
> summary(mAnxDep.AB_ASD.Phrases)

Call:
lm(formula = AnxDep ~ FirstPhrases.months + vabs_soc_ss, data = dASD)

Residuals:
    Min      1Q  Median      3Q     Max
-16.541  -6.628  -4.966   8.902  16.571

Coefficients:
                    Estimate Std. Error t value Pr(>|t|)
(Intercept)         81.97458   16.96082   4.833 0.000266 ***
FirstPhrases.months  0.02523    0.29255   0.086 0.932504
vabs_soc_ss         -0.24123    0.15576  -1.549 0.143748
---
Signif. codes:  0 ‘***’ 0.001 ‘**’ 0.01 ‘*’ 0.05 ‘.’ 0.1 ‘ ’ 1

Residual standard error: 11.58 on 14 degrees of freedom
  (22 observations deleted due to missingness)
Multiple R-squared:  0.1506,	Adjusted R-squared:  0.0293
F-statistic: 1.242 on 2 and 14 DF,  p-value: 0.3189

>
> mAnxDep.AB_NT.Phrases = lm(AnxDep ~ FirstPhrases.months
+                 #+ age
+                       + vabs_soc_ss
+                         , data = dNT)
> summary(mAnxDep.AB_NT.Phrases)

Call:
lm(formula = AnxDep ~ FirstPhrases.months + vabs_soc_ss, data = dNT)

Residuals:
    Min      1Q  Median      3Q     Max
-8.3227 -2.6895 -0.9609  1.0857 19.3649

Coefficients:
                    Estimate Std. Error t value Pr(>|t|)
(Intercept)          77.1672    12.2010   6.325  2.3e-06 ***
FirstPhrases.months  -0.2839     0.1703  -1.668    0.110
vabs_soc_ss          -0.1696     0.1118  -1.516    0.144
---
Signif. codes:  0 ‘***’ 0.001 ‘**’ 0.01 ‘*’ 0.05 ‘.’ 0.1 ‘ ’ 1

Residual standard error: 5.763 on 22 degrees of freedom
  (13 observations deleted due to missingness)
Multiple R-squared:  0.1873,	Adjusted R-squared:  0.1134
F-statistic: 2.535 on 2 and 22 DF,  p-value: 0.1021

>
> mAnxDep.AB_LAD.Phrases = lm(AnxDep ~ FirstPhrases.months
+                 #+ age
+                       + vabs_soc_ss
+                         , data = dLAD)
> summary(mAnxDep.AB_LAD.Phrases)

Call:
lm(formula = AnxDep ~ FirstPhrases.months + vabs_soc_ss, data = dLAD)

Residuals:
   Min     1Q Median     3Q    Max
-5.080 -1.835 -1.391  0.457 15.268

Coefficients:
                    Estimate Std. Error t value Pr(>|t|)
(Intercept)         61.59779    9.52987   6.464 7.82e-06 ***
FirstPhrases.months  0.03555    0.06776   0.525    0.607
vabs_soc_ss         -0.09961    0.08931  -1.115    0.281
---
Signif. codes:  0 ‘***’ 0.001 ‘**’ 0.01 ‘*’ 0.05 ‘.’ 0.1 ‘ ’ 1

Residual standard error: 4.583 on 16 degrees of freedom
  (13 observations deleted due to missingness)
Multiple R-squared:  0.0883,	Adjusted R-squared:  -0.02567
F-statistic: 0.7748 on 2 and 16 DF,  p-value: 0.4773


> ### CELF ###
> #Core Language "cls_std_scr",
> mProb_ASD.CL = lm(cbcl_anxdep_t ~ cls_std_scr
+                       #+ age
+                       + vabs_soc_ss
+                         , data = dASD.inperson)
Error in lm.fit(x, y, offset = offset, singular.ok = singular.ok, ...) :
  NA/NaN/Inf in 'y'
> summary(mProb_ASD.CL)

Call:
lm(formula = cbcl_totalprob_t ~ cls_std_scr + vabs_soc_ss, data = dASD.inperson)

Residuals:
   Min     1Q Median     3Q    Max
-8.359 -4.042 -1.420  4.913 12.592

Coefficients:
            Estimate Std. Error t value Pr(>|t|)
(Intercept) 114.4522    20.2380   5.655  0.00077 ***
cls_std_scr  -0.1977     0.1760  -1.124  0.29826
vabs_soc_ss  -0.4196     0.1522  -2.757  0.02822 *
---
Signif. codes:  0 ‘***’ 0.001 ‘**’ 0.01 ‘*’ 0.05 ‘.’ 0.1 ‘ ’ 1

Residual standard error: 7.597 on 7 degrees of freedom
  (8 observations deleted due to missingness)
Multiple R-squared:  0.5959,	Adjusted R-squared:  0.4804
F-statistic: 5.161 on 2 and 7 DF,  p-value: 0.04195

>
> mProb_LAD.CL = lm(cbcl_anxdep_t ~ cls_std_scr
+                       #+ age
+                       + vabs_soc_ss
+                         , data = dLAD.inperson)
> summary(mProb_LAD.CL)

Call:
lm(formula = cbcl_anxdep_t ~ cls_std_scr + vabs_soc_ss, data = dLAD.inperson)

Residuals:
ALL 3 residuals are 0: no residual degrees of freedom!

Coefficients:
            Estimate Std. Error t value Pr(>|t|)
(Intercept) 198.0155        NaN     NaN      NaN
cls_std_scr  -1.6443        NaN     NaN      NaN
vabs_soc_ss   0.2835        NaN     NaN      NaN

Residual standard error: NaN on 0 degrees of freedom
  (2 observations deleted due to missingness)
Multiple R-squared:      1,	Adjusted R-squared:    NaN
F-statistic:   NaN on 2 and 0 DF,  p-value: NA

>
> mProb_NT.CL = lm(cbcl_anxdep_t ~ cls_std_scr
+                       #+ age
+                       + vabs_soc_ss
+                         , data = dNT.inperson)
> summary(mProb_NT.CL)

Call:
lm(formula = cbcl_anxdep_t ~ cls_std_scr + vabs_soc_ss, data = dNT.inperson)

Residuals:
       1        2        3        5        6        7        9       10
 -0.5717  -2.4730  -0.8894  -1.3962 -10.0952  -5.9834  18.8999   2.5091

Coefficients:
             Estimate Std. Error t value Pr(>|t|)
(Intercept)  29.67398  108.18478   0.274    0.795
cls_std_scr   0.23051    0.53207   0.433    0.683
vabs_soc_ss  -0.01145    0.48481  -0.024    0.982

Residual standard error: 10.1 on 5 degrees of freedom
  (4 observations deleted due to missingness)
Multiple R-squared:  0.126,	Adjusted R-squared:  -0.2236
F-statistic: 0.3605 on 2 and 5 DF,  p-value: 0.7141

>
> #Recalling Sentences "rs_ss"
> mProb_ASD.RS = lm(cbcl_anxdep_t ~ rs_ss
+                       #+ age
+                       + vabs_soc_ss
+                         , data = dASD.inperson)
Error in lm.fit(x, y, offset = offset, singular.ok = singular.ok, ...) :
  NA/NaN/Inf in 'y'
> summary(mProb_ASD.RS)

Call:
lm(formula = cbcl_totalprob_t ~ rs_ss + vabs_soc_ss, data = dASD.inperson)

Residuals:
   Min     1Q Median     3Q    Max
-8.130 -3.069 -2.013  3.418 13.415

Coefficients:
            Estimate Std. Error t value Pr(>|t|)
(Intercept)  94.7443    11.6303   8.146 8.12e-05 ***
rs_ss        -1.1706     0.9340  -1.253    0.250
vabs_soc_ss  -0.2571     0.2131  -1.206    0.267
---
Signif. codes:  0 ‘***’ 0.001 ‘**’ 0.01 ‘*’ 0.05 ‘.’ 0.1 ‘ ’ 1

Residual standard error: 7.459 on 7 degrees of freedom
  (8 observations deleted due to missingness)
Multiple R-squared:  0.6104,	Adjusted R-squared:  0.4991
F-statistic: 5.484 on 2 and 7 DF,  p-value: 0.0369

>
> mProb_LAD.RS =  lm(cbcl_anxdep_t ~ rs_ss
+                       #+ age
+                       + vabs_soc_ss
+                         , data = dLAD.inperson)
> summary(mProb_LAD.RS)

Call:
lm(formula = cbcl_anxdep_t ~ rs_ss + vabs_soc_ss, data = dLAD.inperson)

Residuals:
ALL 3 residuals are 0: no residual degrees of freedom!

Coefficients:
            Estimate Std. Error t value Pr(>|t|)
(Intercept)  64.9839        NaN     NaN      NaN
rs_ss        -2.5726        NaN     NaN      NaN
vabs_soc_ss   0.1774        NaN     NaN      NaN

Residual standard error: NaN on 0 degrees of freedom
  (2 observations deleted due to missingness)
Multiple R-squared:      1,	Adjusted R-squared:    NaN
F-statistic:   NaN on 2 and 0 DF,  p-value: NA

>
> mProb_NT.RS = lm(cbcl_anxdep_t ~ rs_ss
+                       #+ age
+                       + vabs_soc_ss
+                         , data = dNT.inperson)
> summary(mProb_NT.RS)

Call:
lm(formula = cbcl_anxdep_t ~ rs_ss + vabs_soc_ss, data = dNT.inperson)

Residuals:
     1      2      3      5      6      7      9     10
-1.501 -2.504 -2.305 -3.038 -9.593 -2.575 20.147  1.370

Coefficients:
            Estimate Std. Error t value Pr(>|t|)
(Intercept) 75.87389   50.41825   1.505    0.193
rs_ss       -0.03884    1.84078  -0.021    0.984
vabs_soc_ss -0.19284    0.31942  -0.604    0.572

Residual standard error: 10.29 on 5 degrees of freedom
  (4 observations deleted due to missingness)
Multiple R-squared:  0.09331,	Adjusted R-squared:  -0.2694
F-statistic: 0.2573 on 2 and 5 DF,  p-value: 0.7828


> ### RQ 2 B: Educational Outcome ~ Group*Core Language (youth cohort)/Grammaticality Judgement (A’; all participants)/age of first words/phrases
> ###############################
> ##### Full time anything ######
> ###############################
> ### ASD LAD ###
> mFullTime_ASDLAD.GJ = glm(NotvsFullTime ~
+                           Group.NTref*aprime.log
+                  #+ age
+                       + vabs_soc_ss
+                         , family = binomial, data = dASDLAD.older18)
> summary(mFullTime_ASDLAD.GJ)

Call:
glm(formula = NotvsFullTime ~ Group.NTref * aprime.log + vabs_soc_ss,
    family = binomial, data = dASDLAD.older18)

Deviance Residuals:
    Min       1Q   Median       3Q      Max
-2.1947   0.4154   0.4553   0.4969   1.2411

Coefficients:
                         Estimate Std. Error z value Pr(>|z|)
(Intercept)              2.644737   4.474522   0.591    0.554
Group.NTref             -1.215862   2.724364  -0.446    0.655
aprime.log               7.219802   7.158991   1.008    0.313
vabs_soc_ss              0.003379   0.043030   0.079    0.937
Group.NTref:aprime.log -10.781046  14.435813  -0.747    0.455

(Dispersion parameter for binomial family taken to be 1)

    Null deviance: 30.462  on 30  degrees of freedom
Residual deviance: 23.955  on 26  degrees of freedom
  (24 observations deleted due to missingness)
AIC: 33.955

Number of Fisher Scoring iterations: 5

>
> mFullTime_ASDLAD.Words = glm(NotvsFullTime ~
+                           Group.NTref*FirstWords.months
+                  #+ age
+                       + vabs_soc_ss
+                         , family = binomial, data = dASDLAD.older18)
> summary(mFullTime_ASDLAD.Words)

Call:
glm(formula = NotvsFullTime ~ Group.NTref * FirstWords.months +
    vabs_soc_ss, family = binomial, data = dASDLAD.older18)

Deviance Residuals:
    Min       1Q   Median       3Q      Max
-2.3874   0.3467   0.4100   0.6020   1.1325

Coefficients:
                              Estimate Std. Error z value Pr(>|z|)
(Intercept)                   -5.87343    4.50539  -1.304    0.192
Group.NTref                    0.95490    2.47461   0.386    0.700
FirstWords.months              0.04929    0.06374   0.773    0.439
vabs_soc_ss                    0.06770    0.04120   1.643    0.100
Group.NTref:FirstWords.months -0.04381    0.12172  -0.360    0.719

(Dispersion parameter for binomial family taken to be 1)

    Null deviance: 26.276  on 27  degrees of freedom
Residual deviance: 21.793  on 23  degrees of freedom
  (27 observations deleted due to missingness)
AIC: 31.793

Number of Fisher Scoring iterations: 5

>
> mFullTime_ASDLAD.Phrases = glm(NotvsFullTime ~
+                           Group.NTref*FirstPhrases.months
+                  #+ age
+                       + vabs_soc_ss
+                        , family = binomial, data = dASDLAD.older18)
> summary(mFullTime_ASDLAD.Phrases)

Call:
glm(formula = NotvsFullTime ~ Group.NTref * FirstPhrases.months +
    vabs_soc_ss, family = binomial, data = dASDLAD.older18)

Deviance Residuals:
    Min       1Q   Median       3Q      Max
-2.8233   0.1843   0.2865   0.5644   1.1233

Coefficients:
                                Estimate Std. Error z value Pr(>|z|)
(Intercept)                     -5.02432    4.84724  -1.037    0.300
Group.NTref                      5.49498    3.58262   1.534    0.125
FirstPhrases.months              0.01114    0.05313   0.210    0.834
vabs_soc_ss                      0.06901    0.04475   1.542    0.123
Group.NTref:FirstPhrases.months -0.17197    0.11702  -1.470    0.142

(Dispersion parameter for binomial family taken to be 1)

    Null deviance: 26.662  on 28  degrees of freedom
Residual deviance: 19.452  on 24  degrees of freedom
  (26 observations deleted due to missingness)
AIC: 29.452

Number of Fisher Scoring iterations: 6

>
>
>
>
> ### ASD NT ###
> mFullTime_ASDNT.GJ = glm(NotvsFullTime ~
+                           Group.LADref*aprime.log
+                 #+ age
+                       + vabs_soc_ss
+                         , family = binomial, data = dASDNT.older18)
> summary(mFullTime_ASDNT.GJ)

Call:
glm(formula = NotvsFullTime ~ Group.LADref * aprime.log + vabs_soc_ss,
    family = binomial, data = dASDNT.older18)

Deviance Residuals:
    Min       1Q   Median       3Q      Max
-2.1128   0.3132   0.5781   0.6741   1.2052

Coefficients:
                         Estimate Std. Error z value Pr(>|z|)
(Intercept)               1.44055    4.22434   0.341    0.733
Group.LADref             -1.88250    2.55633  -0.736    0.461
aprime.log                7.67666    5.65806   1.357    0.175
vabs_soc_ss               0.01310    0.04218   0.311    0.756
Group.LADref:aprime.log -10.24760   11.23050  -0.912    0.362

(Dispersion parameter for binomial family taken to be 1)

    Null deviance: 37.100  on 33  degrees of freedom
Residual deviance: 32.039  on 29  degrees of freedom
  (18 observations deleted due to missingness)
AIC: 42.039

Number of Fisher Scoring iterations: 4

>
> mFullTime_ASDNT.Words = glm(NotvsFullTime ~
+                           Group.LADref*FirstWords.months
+                 #+ age
+                       + vabs_soc_ss
+                         , family = binomial, data = dASDNT.older18)
> summary(mFullTime_ASDNT.Words)

Call:
glm(formula = NotvsFullTime ~ Group.LADref * FirstWords.months +
    vabs_soc_ss, family = binomial, data = dASDNT.older18)

Deviance Residuals:
    Min       1Q   Median       3Q      Max
-2.3802   0.2223   0.3465   0.6141   1.1764

Coefficients:
                               Estimate Std. Error z value Pr(>|z|)
(Intercept)                    -7.76380    5.40547  -1.436   0.1509
Group.LADref                    3.96321    2.81004   1.410   0.1584
FirstWords.months              -0.03887    0.07410  -0.525   0.5999
vabs_soc_ss                     0.09962    0.05295   1.882   0.0599 .
Group.LADref:FirstWords.months -0.29420    0.17222  -1.708   0.0876 .
---
Signif. codes:  0 ‘***’ 0.001 ‘**’ 0.01 ‘*’ 0.05 ‘.’ 0.1 ‘ ’ 1

(Dispersion parameter for binomial family taken to be 1)

    Null deviance: 30.024  on 29  degrees of freedom
Residual deviance: 22.939  on 25  degrees of freedom
  (22 observations deleted due to missingness)
AIC: 32.939

Number of Fisher Scoring iterations: 5

>
> mFullTime_ASDNT.Phrases = glm(NotvsFullTime ~
+                           Group.LADref*FirstPhrases.months
+                 #+ age
+                       + vabs_soc_ss
+                         , family = binomial, data = dASDNT.older18)
> summary(mFullTime_ASDNT.Phrases)

Call:
glm(formula = NotvsFullTime ~ Group.LADref * FirstPhrases.months +
    vabs_soc_ss, family = binomial, data = dASDNT.older18)

Deviance Residuals:
    Min       1Q   Median       3Q      Max
-2.0880   0.3171   0.5179   0.6727   1.0807

Coefficients:
                                 Estimate Std. Error z value Pr(>|z|)
(Intercept)                      -5.21416    4.69692  -1.110    0.267
Group.LADref                      2.90856    3.16751   0.918    0.358
FirstPhrases.months               0.02036    0.05962   0.342    0.733
vabs_soc_ss                       0.05930    0.04097   1.447    0.148
Group.LADref:FirstPhrases.months -0.13205    0.11949  -1.105    0.269

(Dispersion parameter for binomial family taken to be 1)

    Null deviance: 34.106  on 32  degrees of freedom
Residual deviance: 30.539  on 28  degrees of freedom
  (19 observations deleted due to missingness)
AIC: 40.539

Number of Fisher Scoring iterations: 5

>
>
>
>
> ### LAD NT ###
> mFullTime_LADNT.GJ = glm(NotvsFullTime ~
+                           Group.ASDref*aprime.log
+                 #+ age
+                       + vabs_soc_ss
+                         , family = binomial, data = dLADNT.older18)
> summary(mFullTime_LADNT.GJ)

Call:
glm(formula = NotvsFullTime ~ Group.ASDref * aprime.log + vabs_soc_ss,
    family = binomial, data = dLADNT.older18)

Deviance Residuals:
    Min       1Q   Median       3Q      Max
-2.0940   0.3456   0.4866   0.6609   0.7942

Coefficients:
                        Estimate Std. Error z value Pr(>|z|)
(Intercept)             11.25297   10.05959   1.119    0.263
Group.ASDref            -0.77147    1.76984  -0.436    0.663
aprime.log               2.60050    6.54217   0.397    0.691
vabs_soc_ss             -0.08530    0.09289  -0.918    0.358
Group.ASDref:aprime.log -1.83807   13.20262  -0.139    0.889

(Dispersion parameter for binomial family taken to be 1)

    Null deviance: 34.754  on 42  degrees of freedom
Residual deviance: 32.862  on 38  degrees of freedom
  (14 observations deleted due to missingness)
AIC: 42.862

Number of Fisher Scoring iterations: 6

>
> mFullTime_LADNT.Words = glm(NotvsFullTime ~
+                           Group.ASDref*FirstWords.months
+                 #+ age
+                       + vabs_soc_ss
+                         , family = binomial, data = dLADNT.older18)
> summary(mFullTime_LADNT.Words)

Call:
glm(formula = NotvsFullTime ~ Group.ASDref * FirstWords.months +
    vabs_soc_ss, family = binomial, data = dLADNT.older18)

Deviance Residuals:
    Min       1Q   Median       3Q      Max
-2.3257   0.2652   0.4142   0.5618   0.9319

Coefficients:
                               Estimate Std. Error z value Pr(>|z|)
(Intercept)                    16.46483   16.04986   1.026    0.305
Group.ASDref                    2.53572    2.73508   0.927    0.354
FirstWords.months              -0.09105    0.08079  -1.127    0.260
vabs_soc_ss                    -0.12411    0.14338  -0.866    0.387
Group.ASDref:FirstWords.months -0.20354    0.15698  -1.297    0.195

(Dispersion parameter for binomial family taken to be 1)

    Null deviance: 29.593  on 37  degrees of freedom
Residual deviance: 26.427  on 33  degrees of freedom
  (19 observations deleted due to missingness)
AIC: 36.427

Number of Fisher Scoring iterations: 6

>
> mFullTime_LADNT.Phrases = glm(NotvsFullTime ~
+                           Group.ASDref*FirstPhrases.months
+                 #+ age
+                       + vabs_soc_ss
+                         , family = binomial, data = dLADNT.older18)
> summary(mFullTime_LADNT.Phrases)

Call:
glm(formula = NotvsFullTime ~ Group.ASDref * FirstPhrases.months +
    vabs_soc_ss, family = binomial, data = dLADNT.older18)

Deviance Residuals:
    Min       1Q   Median       3Q      Max
-2.3590   0.1687   0.4320   0.5889   1.0610

Coefficients:
                                 Estimate Std. Error z value Pr(>|z|)
(Intercept)                      17.75337   14.69181   1.208    0.227
Group.ASDref                     -1.06698    2.50987  -0.425    0.671
FirstPhrases.months              -0.07016    0.04557  -1.540    0.124
vabs_soc_ss                      -0.13214    0.13230  -0.999    0.318
Group.ASDref:FirstPhrases.months -0.02553    0.09283  -0.275    0.783

(Dispersion parameter for binomial family taken to be 1)

    Null deviance: 33.817  on 39  degrees of freedom
Residual deviance: 28.461  on 35  degrees of freedom
  (17 observations deleted due to missingness)
AIC: 38.461

Number of Fisher Scoring iterations: 6


> ###Follow up
>
> #aprime.log
> mFullTime_ASD.GJ = glm(NotvsFullTime ~ aprime.log
+                 #+ age
+                       + vabs_soc_ss
+                         , family = binomial, data = dASD.older18)
> summary(mFullTime_ASD.GJ)

Call:
glm(formula = NotvsFullTime ~ aprime.log + vabs_soc_ss, family = binomial,
    data = dASD.older18)

Deviance Residuals:
    Min       1Q   Median       3Q      Max
-2.0230  -0.6538   0.4255   0.7446   1.1586

Coefficients:
            Estimate Std. Error z value Pr(>|z|)
(Intercept)  0.19588    5.12828   0.038    0.970
aprime.log  13.87515    9.41758   1.473    0.141
vabs_soc_ss  0.03917    0.05728   0.684    0.494

(Dispersion parameter for binomial family taken to be 1)

    Null deviance: 14.421  on 10  degrees of freedom
Residual deviance: 10.471  on  8  degrees of freedom
  (14 observations deleted due to missingness)
AIC: 16.471

Number of Fisher Scoring iterations: 5

>
> mFullTime_NT.GJ = glm(NotvsFullTime ~ aprime.log
+                 #+ age
+                       + vabs_soc_ss
+                         , family = binomial, data = dNT.older18)
> summary(mFullTime_NT.GJ)

Call:
glm(formula = NotvsFullTime ~ aprime.log + vabs_soc_ss, family = binomial,
    data = dNT.older18)

Deviance Residuals:
    Min       1Q   Median       3Q      Max
-1.9913   0.4953   0.5905   0.6519   0.7741

Coefficients:
            Estimate Std. Error z value Pr(>|z|)
(Intercept)  6.23360    9.67393   0.644    0.519
aprime.log   2.03647    7.27247   0.280    0.779
vabs_soc_ss -0.04169    0.09099  -0.458    0.647

(Dispersion parameter for binomial family taken to be 1)

    Null deviance: 21.254  on 22  degrees of freedom
Residual deviance: 20.906  on 20  degrees of freedom
  (4 observations deleted due to missingness)
AIC: 26.906

Number of Fisher Scoring iterations: 4

>
> mFullTime_LAD.GJ = glm(NotvsFullTime ~ aprime.log
+                 #+ age
+                       + vabs_soc_ss
+                         , family = binomial, data = dLAD.older18)
> summary(mFullTime_LAD.GJ)

Call:
glm(formula = NotvsFullTime ~ aprime.log + vabs_soc_ss, family = binomial,
    data = dLAD.older18)

Deviance Residuals:
     Min        1Q    Median        3Q       Max
-2.05445   0.05107   0.32052   0.53189   0.80706

Coefficients:
            Estimate Std. Error z value Pr(>|z|)
(Intercept)  55.9295    58.7996   0.951    0.342
aprime.log    7.2369    11.6191   0.623    0.533
vabs_soc_ss  -0.4870     0.5302  -0.919    0.358

(Dispersion parameter for binomial family taken to be 1)

    Null deviance: 13.003  on 19  degrees of freedom
Residual deviance: 10.736  on 17  degrees of freedom
  (10 observations deleted due to missingness)
AIC: 16.736

Number of Fisher Scoring iterations: 8

>
> #Words
> mFullTime_ASD.Words = glm(NotvsFullTime ~ FirstWords.months
+                 #+ age
+                       + vabs_soc_ss
+                         , family = binomial, data = dASD.older18)
> summary(mFullTime_ASD.Words)

Call:
glm(formula = NotvsFullTime ~ FirstWords.months + vabs_soc_ss,
    family = binomial, data = dASD.older18)

Deviance Residuals:
       Min          1Q      Median          3Q         Max
-3.607e-05  -1.100e-08   2.100e-08   2.100e-08   3.336e-05

Coefficients:
                    Estimate Std. Error z value Pr(>|z|)
(Intercept)         -2660.07 1970091.59  -0.001    0.999
FirstWords.months      33.88   25245.57   0.001    0.999
vabs_soc_ss            22.85   16836.08   0.001    0.999

(Dispersion parameter for binomial family taken to be 1)

    Null deviance: 1.2217e+01  on 9  degrees of freedom
Residual deviance: 3.2074e-09  on 7  degrees of freedom
  (15 observations deleted due to missingness)
AIC: 6

Number of Fisher Scoring iterations: 25

>
> mFullTime_NT.Words = glm(NotvsFullTime ~ FirstWords.months
+                 #+ age
+                       + vabs_soc_ss
+                         , family = binomial, data = dNT.older18)
> summary(mFullTime_NT.Words)

Call:
glm(formula = NotvsFullTime ~ FirstWords.months + vabs_soc_ss,
    family = binomial, data = dNT.older18)

Deviance Residuals:
    Min       1Q   Median       3Q      Max
-2.2025   0.3262   0.3966   0.5314   0.9417

Coefficients:
                  Estimate Std. Error z value Pr(>|z|)
(Intercept)        10.9523    15.9665   0.686    0.493
FirstWords.months  -0.1706     0.1333  -1.280    0.201
vabs_soc_ss        -0.0638     0.1426  -0.447    0.655

(Dispersion parameter for binomial family taken to be 1)

    Null deviance: 16.908  on 19  degrees of freedom
Residual deviance: 14.978  on 17  degrees of freedom
  (7 observations deleted due to missingness)
AIC: 20.978

Number of Fisher Scoring iterations: 6

>
> mFullTime_LAD.Words = glm(NotvsFullTime ~ FirstWords.months
+                 #+ age
+                       + vabs_soc_ss
+                         , family = binomial, data = dLAD.older18)
> summary(mFullTime_LAD.Words)

Call:
glm(formula = NotvsFullTime ~ FirstWords.months + vabs_soc_ss,
    family = binomial, data = dLAD.older18)

Deviance Residuals:
    Min       1Q   Median       3Q      Max
-1.9089   0.1226   0.3760   0.5951   0.8143

Coefficients:
                   Estimate Std. Error z value Pr(>|z|)
(Intercept)       38.592110  45.672519   0.845    0.398
FirstWords.months  0.002593   0.074096   0.035    0.972
vabs_soc_ss       -0.336514   0.412807  -0.815    0.415

(Dispersion parameter for binomial family taken to be 1)

    Null deviance: 12.558  on 17  degrees of freedom
Residual deviance: 10.921  on 15  degrees of freedom
  (12 observations deleted due to missingness)
AIC: 16.921

Number of Fisher Scoring iterations: 7

>
> #Phrases
> mFullTime_ASD.Phrases = glm(NotvsFullTime ~ FirstPhrases.months
+                 #+ age
+                       + vabs_soc_ss
+                         , family = binomial, data = dASD.older18)
> summary(mFullTime_ASD.Phrases)

Call:
glm(formula = NotvsFullTime ~ FirstPhrases.months + vabs_soc_ss,
    family = binomial, data = dASD.older18)

Deviance Residuals:
    Min       1Q   Median       3Q      Max
-1.5044  -0.1780   0.1002   0.3385   1.4814

Coefficients:
                    Estimate Std. Error z value Pr(>|z|)
(Intercept)         -21.8788    16.4719  -1.328    0.184
FirstPhrases.months   0.2479     0.2148   1.154    0.248
vabs_soc_ss           0.1929     0.1404   1.374    0.169

(Dispersion parameter for binomial family taken to be 1)

    Null deviance: 12.8910  on 10  degrees of freedom
Residual deviance:  6.5977  on  8  degrees of freedom
  (14 observations deleted due to missingness)
AIC: 12.598

Number of Fisher Scoring iterations: 7

>
> mFullTime_NT.Phrases = glm(NotvsFullTime ~ FirstPhrases.months
+                 #+ age
+                       + vabs_soc_ss
+                         , family = binomial, data = dNT.older18)
> summary(mFullTime_NT.Phrases)

Call:
glm(formula = NotvsFullTime ~ FirstPhrases.months + vabs_soc_ss,
    family = binomial, data = dNT.older18)

Deviance Residuals:
    Min       1Q   Median       3Q      Max
-2.1753   0.3187   0.5782   0.6685   0.9736

Coefficients:
                    Estimate Std. Error z value Pr(>|z|)
(Intercept)         12.51832   14.52924   0.862    0.389
FirstPhrases.months -0.07079    0.07999  -0.885    0.376
vabs_soc_ss         -0.09059    0.12809  -0.707    0.479

(Dispersion parameter for binomial family taken to be 1)

    Null deviance: 20.862  on 21  degrees of freedom
Residual deviance: 19.654  on 19  degrees of freedom
  (5 observations deleted due to missingness)
AIC: 25.654

Number of Fisher Scoring iterations: 5

>
> mFullTime_LAD.Phrases = glm(NotvsFullTime ~ FirstPhrases.months
+                 #+ age
+                       + vabs_soc_ss
+                         , family = binomial, data = dLAD.older18)
> summary(mFullTime_LAD.Phrases)

Call:
glm(formula = NotvsFullTime ~ FirstPhrases.months + vabs_soc_ss,
    family = binomial, data = dLAD.older18)

Deviance Residuals:
     Min        1Q    Median        3Q       Max
-2.27126   0.02903   0.21933   0.44688   0.88726

Coefficients:
                    Estimate Std. Error z value Pr(>|z|)
(Intercept)         61.10264   66.31096   0.921    0.357
FirstPhrases.months -0.05904    0.04187  -1.410    0.158
vabs_soc_ss         -0.52309    0.60000  -0.872    0.383

(Dispersion parameter for binomial family taken to be 1)

    Null deviance: 12.5580  on 17  degrees of freedom
Residual deviance:  8.0764  on 15  degrees of freedom
  (12 observations deleted due to missingness)
AIC: 14.076

Number of Fisher Scoring iterations: 8


> ##################################################
> ##### Education completed 22 years and older #####
> ##################################################
> ### ASD LAD ###
> mEdComp_ASDLAD.GJ = lm(Edcompl.clean ~
+                           Group.NTref*aprime.log
+                  #+ age
+                       + vabs_soc_ss
+                         , data = dASDLAD.older22)
> summary(mEdComp_ASDLAD.GJ)

Call:
lm(formula = Edcompl.clean ~ Group.NTref * aprime.log + vabs_soc_ss,
    data = dASDLAD.older22)

Residuals:
    Min      1Q  Median      3Q     Max
-3.0290 -0.3982 -0.1665  0.7914  1.6001

Coefficients:
                        Estimate Std. Error t value Pr(>|t|)
(Intercept)            1.191e+01  3.258e+00   3.657  0.00377 **
Group.NTref            1.257e+00  1.466e+00   0.858  0.40930
aprime.log             2.358e+00  4.764e+00   0.495  0.63042
vabs_soc_ss            1.129e-04  3.221e-02   0.004  0.99727
Group.NTref:aprime.log 1.269e+00  9.678e+00   0.131  0.89804
---
Signif. codes:  0 ‘***’ 0.001 ‘**’ 0.01 ‘*’ 0.05 ‘.’ 0.1 ‘ ’ 1

Residual standard error: 1.271 on 11 degrees of freedom
  (17 observations deleted due to missingness)
Multiple R-squared:  0.2516,	Adjusted R-squared:  -0.02057
F-statistic: 0.9244 on 4 and 11 DF,  p-value: 0.4842

>
> mEdComp_ASDLAD.Words = lm(Edcompl.clean ~
+                           Group.NTref*FirstWords.months
+                  #+ age
+                       + vabs_soc_ss
+                         , data = dASDLAD.older22)
> summary(mEdComp_ASDLAD.Words)

Call:
lm(formula = Edcompl.clean ~ Group.NTref * FirstWords.months +
    vabs_soc_ss, data = dASDLAD.older22)

Residuals:
     Min       1Q   Median       3Q      Max
-3.07355 -0.32611  0.01582  0.70715  1.20374

Coefficients:
                               Estimate Std. Error t value Pr(>|t|)
(Intercept)                   12.067571   3.319235   3.636  0.00457 **
Group.NTref                    0.808797   1.550505   0.522  0.61328
FirstWords.months              0.023298   0.038569   0.604  0.55926
vabs_soc_ss                   -0.008522   0.028737  -0.297  0.77288
Group.NTref:FirstWords.months  0.023106   0.074582   0.310  0.76307
---
Signif. codes:  0 ‘***’ 0.001 ‘**’ 0.01 ‘*’ 0.05 ‘.’ 0.1 ‘ ’ 1

Residual standard error: 1.223 on 10 degrees of freedom
  (18 observations deleted due to missingness)
Multiple R-squared:  0.3119,	Adjusted R-squared:  0.03665
F-statistic: 1.133 on 4 and 10 DF,  p-value: 0.3948

>
> mEdComp_ASDLAD.Phrases = lm(Edcompl.clean ~
+                           Group.NTref*FirstPhrases.months
+                  #+ age
+                       + vabs_soc_ss
+                         , data = dASDLAD.older22)
> summary(mEdComp_ASDLAD.Phrases)

Call:
lm(formula = Edcompl.clean ~ Group.NTref * FirstPhrases.months +
    vabs_soc_ss, data = dASDLAD.older22)

Residuals:
     Min       1Q   Median       3Q      Max
-1.78518  0.01388  0.08595  0.40848  1.10303

Coefficients:
                                 Estimate Std. Error t value Pr(>|t|)
(Intercept)                      8.247710   2.046826   4.030  0.00167 **
Group.NTref                      2.558456   1.447428   1.768  0.10252
FirstPhrases.months              0.090218   0.024931   3.619  0.00352 **
vabs_soc_ss                      0.008007   0.016860   0.475  0.64338
Group.NTref:FirstPhrases.months -0.048902   0.047969  -1.019  0.32810
---
Signif. codes:  0 ‘***’ 0.001 ‘**’ 0.01 ‘*’ 0.05 ‘.’ 0.1 ‘ ’ 1

Residual standard error: 0.8511 on 12 degrees of freedom
  (16 observations deleted due to missingness)
Multiple R-squared:  0.6132,	Adjusted R-squared:  0.4842
F-statistic: 4.755 on 4 and 12 DF,  p-value: 0.01568

>
>
>
> ### ASD NT ###
> mEdComp_ASDNT = lm(Edcompl.clean ~
+                           Group.LADref
+                 #+ age
+                       + vabs_soc_ss
+                         , data = dASDNT.older22)
> summary(mEdComp_ASDNT)

Call:
lm(formula = Edcompl.clean ~ Group.LADref + vabs_soc_ss, data = dASDNT.older22)

Residuals:
    Min      1Q  Median      3Q     Max
-7.1312 -0.1141 -0.0623  0.9097  2.9033

Coefficients:
              Estimate Std. Error t value Pr(>|t|)
(Intercept)  12.511029   2.857394   4.378 0.000219 ***
Group.LADref  1.017072   0.958132   1.062 0.299475
vabs_soc_ss  -0.008625   0.029384  -0.294 0.771764
---
Signif. codes:  0 ‘***’ 0.001 ‘**’ 0.01 ‘*’ 0.05 ‘.’ 0.1 ‘ ’ 1

Residual standard error: 1.876 on 23 degrees of freedom
  (9 observations deleted due to missingness)
Multiple R-squared:  0.05165,	Adjusted R-squared:  -0.03081
F-statistic: 0.6263 on 2 and 23 DF,  p-value: 0.5434

>
> mEdComp_ASDNT.GJ = lm(Edcompl.clean ~
+                           Group.LADref*aprime.log
+                 #+ age
+                       + vabs_soc_ss
+                         , data = dASDNT.older22)
> summary(mEdComp_ASDNT.GJ)

Call:
lm(formula = Edcompl.clean ~ Group.LADref * aprime.log + vabs_soc_ss,
    data = dASDNT.older22)

Residuals:
    Min      1Q  Median      3Q     Max
-6.9984 -0.1355  0.1062  0.8487  2.9617

Coefficients:
                         Estimate Std. Error t value Pr(>|t|)
(Intercept)             10.799225   4.693424   2.301   0.0336 *
Group.LADref             0.525654   2.063496   0.255   0.8018
aprime.log               0.273204   5.387478   0.051   0.9601
vabs_soc_ss              0.008425   0.047484   0.177   0.8612
Group.LADref:aprime.log -3.203236  10.641893  -0.301   0.7669
---
Signif. codes:  0 ‘***’ 0.001 ‘**’ 0.01 ‘*’ 0.05 ‘.’ 0.1 ‘ ’ 1

Residual standard error: 2.098 on 18 degrees of freedom
  (12 observations deleted due to missingness)
Multiple R-squared:  0.06365,	Adjusted R-squared:  -0.1444
F-statistic: 0.3059 on 4 and 18 DF,  p-value: 0.8702

>
> mEdComp_ASDNT.Words = lm(Edcompl.clean ~
+                           Group.LADref*FirstWords.months
+                 #+ age
+                       + vabs_soc_ss
+                         , data = dASDNT.older22)
> summary(mEdComp_ASDNT.Words)

Call:
lm(formula = Edcompl.clean ~ Group.LADref * FirstWords.months +
    vabs_soc_ss, data = dASDNT.older22)

Residuals:
    Min      1Q  Median      3Q     Max
-7.0110 -0.1446  0.2557  0.9872  2.5339

Coefficients:
                                Estimate Std. Error t value Pr(>|t|)
(Intercept)                    1.085e+01  5.034e+00   2.156   0.0457 *
Group.LADref                   4.467e-01  2.685e+00   0.166   0.8698
FirstWords.months              4.990e-02  7.534e-02   0.662   0.5167
vabs_soc_ss                    2.588e-04  4.378e-02   0.006   0.9954
Group.LADref:FirstWords.months 5.157e-02  1.518e-01   0.340   0.7383
---
Signif. codes:  0 ‘***’ 0.001 ‘**’ 0.01 ‘*’ 0.05 ‘.’ 0.1 ‘ ’ 1

Residual standard error: 2.139 on 17 degrees of freedom
  (13 observations deleted due to missingness)
Multiple R-squared:  0.08046,	Adjusted R-squared:  -0.1359
F-statistic: 0.3719 on 4 and 17 DF,  p-value: 0.8254

>
> mEdComp_ASDNT.Phrases = lm(Edcompl.clean ~
+                           Group.LADref*FirstPhrases.months
+                 #+ age
+                       + vabs_soc_ss
+                         , data = dASDNT.older22)
> summary(mEdComp_ASDNT.Phrases)

Call:
lm(formula = Edcompl.clean ~ Group.LADref * FirstPhrases.months +
    vabs_soc_ss, data = dASDNT.older22)

Residuals:
    Min      1Q  Median      3Q     Max
-6.8401 -0.4470  0.2273  0.9980  2.8766

Coefficients:
                                  Estimate Std. Error t value Pr(>|t|)
(Intercept)                       8.561865   3.989170   2.146   0.0443 *
Group.LADref                      3.418135   2.920506   1.170   0.2556
FirstPhrases.months               0.082562   0.053455   1.545   0.1381
vabs_soc_ss                       0.008993   0.033709   0.267   0.7924
Group.LADref:FirstPhrases.months -0.066221   0.109194  -0.606   0.5510
---
Signif. codes:  0 ‘***’ 0.001 ‘**’ 0.01 ‘*’ 0.05 ‘.’ 0.1 ‘ ’ 1

Residual standard error: 1.9 on 20 degrees of freedom
  (10 observations deleted due to missingness)
Multiple R-squared:  0.1473,	Adjusted R-squared:  -0.02324
F-statistic: 0.8637 on 4 and 20 DF,  p-value: 0.5026

>
>
>
>
> ### LAD NT ###
> mEdComp_LADNT.GJ = lm(Edcompl.clean ~
+                           Group.ASDref*aprime.log
+                 #+ age
+                       + vabs_soc_ss
+                         , data = dLADNT.older22)
> summary(mEdComp_LADNT.GJ)

Call:
lm(formula = Edcompl.clean ~ Group.ASDref * aprime.log + vabs_soc_ss,
    data = dLADNT.older22)

Residuals:
    Min      1Q  Median      3Q     Max
-6.9807 -0.3710 -0.0292  0.7295  2.9174

Coefficients:
                        Estimate Std. Error t value Pr(>|t|)
(Intercept)              9.68271    4.63835   2.088   0.0476 *
Group.ASDref            -0.51035    1.39429  -0.366   0.7176
aprime.log               0.50862    6.07225   0.084   0.9339
vabs_soc_ss              0.02426    0.04346   0.558   0.5819
Group.ASDref:aprime.log -3.12609   12.34797  -0.253   0.8023
---
Signif. codes:  0 ‘***’ 0.001 ‘**’ 0.01 ‘*’ 0.05 ‘.’ 0.1 ‘ ’ 1

Residual standard error: 1.739 on 24 degrees of freedom
  (7 observations deleted due to missingness)
Multiple R-squared:  0.02139,	Adjusted R-squared:  -0.1417
F-statistic: 0.1311 on 4 and 24 DF,  p-value: 0.9695

>
> mEdComp_LADNT.Words = lm(Edcompl.clean ~
+                           Group.ASDref*FirstWords.months
+                 #+ age
+                       + vabs_soc_ss
+                         , data = dLADNT.older22)
> summary(mEdComp_LADNT.Words)

Call:
lm(formula = Edcompl.clean ~ Group.ASDref * FirstWords.months +
    vabs_soc_ss, data = dLADNT.older22)

Residuals:
    Min      1Q  Median      3Q     Max
-6.9564 -0.2251  0.0535  0.8170  2.4662

Coefficients:
                               Estimate Std. Error t value Pr(>|t|)
(Intercept)                     7.90764    5.25570   1.505    0.148
Group.ASDref                   -0.36396    1.83249  -0.199    0.845
FirstWords.months               0.05645    0.05128   1.101    0.284
vabs_soc_ss                     0.03238    0.04895   0.661    0.516
Group.ASDref:FirstWords.months  0.03639    0.10268   0.354    0.727

Residual standard error: 1.821 on 20 degrees of freedom
  (11 observations deleted due to missingness)
Multiple R-squared:  0.07841,	Adjusted R-squared:  -0.1059
F-statistic: 0.4254 on 4 and 20 DF,  p-value: 0.7885

>
> mEdComp_LADNT.Phrases = lm(Edcompl.clean ~
+                           Group.ASDref*FirstPhrases.months
+                 #+ age
+                       + vabs_soc_ss
+                         , data = dLADNT.older22)
> summary(mEdComp_LADNT.Phrases)

Call:
lm(formula = Edcompl.clean ~ Group.ASDref * FirstPhrases.months +
    vabs_soc_ss, data = dLADNT.older22)

Residuals:
    Min      1Q  Median      3Q     Max
-6.8010 -0.2354  0.1310  0.7083  2.8401

Coefficients:
                                 Estimate Std. Error t value Pr(>|t|)
(Intercept)                       7.51577    4.93013   1.524    0.141
Group.ASDref                      0.99527    2.07724   0.479    0.636
FirstPhrases.months               0.05827    0.04154   1.403    0.174
vabs_soc_ss                       0.03082    0.04520   0.682    0.502
Group.ASDref:FirstPhrases.months -0.02230    0.08393  -0.266    0.793

Residual standard error: 1.699 on 23 degrees of freedom
  (8 observations deleted due to missingness)
Multiple R-squared:  0.09551,	Adjusted R-squared:  -0.06179
F-statistic: 0.6072 on 4 and 23 DF,  p-value: 0.6615

>


> ###Follow up
> #aprime.log
> mEdComp_ASD.GJ = lm(Edcompl.clean ~ aprime.log
+                 #+ age
+                       + vabs_soc_ss
+                        , data = dASD.older22)
> summary(mEdComp_ASD.GJ)

Call:
lm(formula = Edcompl.clean ~ aprime.log + vabs_soc_ss, data = dASD.older22)

Residuals:
      6      10      12      14      15
 1.1701 -3.0160  0.7040  0.3718  0.7701

Coefficients:
            Estimate Std. Error t value Pr(>|t|)
(Intercept) 12.34274    7.32771   1.684    0.234
aprime.log   1.50861    9.94556   0.152    0.893
vabs_soc_ss -0.01163    0.07825  -0.149    0.895

Residual standard error: 2.418 on 2 degrees of freedom
  (11 observations deleted due to missingness)
Multiple R-squared:  0.02566,	Adjusted R-squared:  -0.9487
F-statistic: 0.02633 on 2 and 2 DF,  p-value: 0.9743

>
> mEdComp_NT.GJ = lm(Edcompl.clean ~ aprime.log
+                 #+ age
+                       + vabs_soc_ss
+                         , data = dNT.older22)
> summary(mEdComp_NT.GJ)

Call:
lm(formula = Edcompl.clean ~ aprime.log + vabs_soc_ss, data = dNT.older22)

Residuals:
    Min      1Q  Median      3Q     Max
-6.9769 -0.2506  0.1255  0.7607  2.9080

Coefficients:
            Estimate Std. Error t value Pr(>|t|)
(Intercept)  9.08029    6.95838   1.305    0.212
aprime.log  -0.99621    6.48976  -0.154    0.880
vabs_soc_ss  0.02762    0.06681   0.413    0.685

Residual standard error: 2.11 on 15 degrees of freedom
  (1 observation deleted due to missingness)
Multiple R-squared:  0.01473,	Adjusted R-squared:  -0.1166
F-statistic: 0.1121 on 2 and 15 DF,  p-value: 0.8947

>
> mEdComp_LAD.GJ = lm(Edcompl.clean ~ aprime.log
+                 #+ age
+                       + vabs_soc_ss
+                         , data = dLAD.older22)
> summary(mEdComp_LAD.GJ)

Call:
lm(formula = Edcompl.clean ~ aprime.log + vabs_soc_ss, data = dLAD.older22)

Residuals:
    Min      1Q  Median      3Q     Max
-1.1849 -0.4184 -0.1839  0.4300  1.5236

Coefficients:
            Estimate Std. Error t value Pr(>|t|)
(Intercept)  10.5377     3.7626   2.801   0.0232 *
aprime.log    2.2837     5.4756   0.417   0.6876
vabs_soc_ss   0.0187     0.0345   0.542   0.6026
---
Signif. codes:  0 ‘***’ 0.001 ‘**’ 0.01 ‘*’ 0.05 ‘.’ 0.1 ‘ ’ 1

Residual standard error: 0.8473 on 8 degrees of freedom
  (6 observations deleted due to missingness)
Multiple R-squared:  0.07102,	Adjusted R-squared:  -0.1612
F-statistic: 0.3058 on 2 and 8 DF,  p-value: 0.7448

>
> #Words
> mEdComp_ASD.Words = lm(Edcompl.clean ~ FirstWords.months
+                 #+ age
+                       + vabs_soc_ss
+                         , data = dASD.older22)
> summary(mEdComp_ASD.Words)

Call:
lm(formula = Edcompl.clean ~ FirstWords.months + vabs_soc_ss,
    data = dASD.older22)

Residuals:
      2       5       6      10      11      12
 0.3699  0.3108  1.2130 -3.0528  0.2599  0.8991

Coefficients:
                   Estimate Std. Error t value Pr(>|t|)
(Intercept)       13.155361   6.566521   2.003    0.139
FirstWords.months -0.006112   0.114163  -0.054    0.961
vabs_soc_ss       -0.021197   0.053099  -0.399    0.716

Residual standard error: 1.992 on 3 degrees of freedom
  (10 observations deleted due to missingness)
Multiple R-squared:  0.0727,	Adjusted R-squared:  -0.5455
F-statistic: 0.1176 on 2 and 3 DF,  p-value: 0.893

>
> mEdComp_NT.Words = lm(Edcompl.clean ~ FirstWords.months
+                 #+ age
+                       + vabs_soc_ss
+                         , data = dNT.older22)
> summary(mEdComp_NT.Words)

Call:
lm(formula = Edcompl.clean ~ FirstWords.months + vabs_soc_ss,
    data = dNT.older22)

Residuals:
    Min      1Q  Median      3Q     Max
-6.9586 -0.2880  0.4381  0.9687  2.4690

Coefficients:
                  Estimate Std. Error t value Pr(>|t|)
(Intercept)        7.86479    7.56799   1.039    0.318
FirstWords.months  0.07469    0.10654   0.701    0.496
vabs_soc_ss        0.03104    0.07103   0.437    0.669

Residual standard error: 2.224 on 13 degrees of freedom
  (3 observations deleted due to missingness)
Multiple R-squared:  0.05081,	Adjusted R-squared:  -0.09521
F-statistic: 0.348 on 2 and 13 DF,  p-value: 0.7125

>
> mEdComp_LAD.Words = lm(Edcompl.clean ~ FirstWords.months
+                 #+ age
+                       + vabs_soc_ss
+                         , data = dLAD.older22)
> summary(mEdComp_LAD.Words)

Call:
lm(formula = Edcompl.clean ~ FirstWords.months + vabs_soc_ss,
    data = dLAD.older22)

Residuals:
     Min       1Q   Median       3Q      Max
-0.70075 -0.23838 -0.16132  0.00571  1.06111

Coefficients:
                  Estimate Std. Error t value Pr(>|t|)
(Intercept)        7.74281    3.13784   2.468   0.0486 *
FirstWords.months  0.03853    0.01742   2.212   0.0689 .
vabs_soc_ss        0.03561    0.02902   1.227   0.2657
---
Signif. codes:  0 ‘***’ 0.001 ‘**’ 0.01 ‘*’ 0.05 ‘.’ 0.1 ‘ ’ 1

Residual standard error: 0.5833 on 6 degrees of freedom
  (8 observations deleted due to missingness)
Multiple R-squared:  0.4897,	Adjusted R-squared:  0.3196
F-statistic: 2.879 on 2 and 6 DF,  p-value: 0.1329

>
> #Phrases
> mEdComp_ASD.Phrases = lm(Edcompl.clean ~ FirstPhrases.months
+                #+ age
+                       + vabs_soc_ss
+                         , data = dASD.older22)
> summary(mEdComp_ASD.Phrases)

Call:
lm(formula = Edcompl.clean ~ FirstPhrases.months + vabs_soc_ss,
    data = dASD.older22)

Residuals:
       2        5        6       10       11       12       15
 0.06678  0.51453  0.22103 -1.81869  0.92911  1.10840 -1.02115

Coefficients:
                    Estimate Std. Error t value Pr(>|t|)
(Intercept)         7.760795   3.731213   2.080    0.106
FirstPhrases.months 0.107792   0.060743   1.775    0.151
vabs_soc_ss         0.001252   0.028003   0.045    0.966

Residual standard error: 1.3 on 4 degrees of freedom
  (9 observations deleted due to missingness)
Multiple R-squared:  0.4966,	Adjusted R-squared:  0.2449
F-statistic: 1.973 on 2 and 4 DF,  p-value: 0.2534

>
> mEdComp_NT.Phrases = lm(Edcompl.clean ~ FirstPhrases.months
+                 #+ age
+                       + vabs_soc_ss
+                         , data = dNT.older22)
> summary(mEdComp_NT.Phrases)

Call:
lm(formula = Edcompl.clean ~ FirstPhrases.months + vabs_soc_ss,
    data = dNT.older22)

Residuals:
    Min      1Q  Median      3Q     Max
-6.8108 -0.4414  0.3368  0.8806  2.8493

Coefficients:
                    Estimate Std. Error t value Pr(>|t|)
(Intercept)          8.58027    6.88129   1.247    0.232
FirstPhrases.months  0.04771    0.07255   0.658    0.521
vabs_soc_ss          0.02534    0.06517   0.389    0.703

Residual standard error: 2.082 on 15 degrees of freedom
  (1 observation deleted due to missingness)
Multiple R-squared:  0.04083,	Adjusted R-squared:  -0.08706
F-statistic: 0.3192 on 2 and 15 DF,  p-value: 0.7315

>
> mEdComp_LAD.Phrases = lm(Edcompl.clean ~ FirstPhrases.months
+                 #+ age
+                       + vabs_soc_ss
+                         , data = dLAD.older22)
> summary(mEdComp_LAD.Phrases)

Call:
lm(formula = Edcompl.clean ~ FirstPhrases.months + vabs_soc_ss,
    data = dLAD.older22)

Residuals:
     Min       1Q   Median       3Q      Max
-0.99515  0.00080  0.04992  0.24366  0.29984

Coefficients:
                    Estimate Std. Error t value Pr(>|t|)
(Intercept)          5.45255    2.42405   2.249  0.05925 .
FirstPhrases.months  0.07170    0.01521   4.715  0.00217 **
vabs_soc_ss          0.04506    0.02169   2.077  0.07637 .
---
Signif. codes:  0 ‘***’ 0.001 ‘**’ 0.01 ‘*’ 0.05 ‘.’ 0.1 ‘ ’ 1

Residual standard error: 0.43 on 7 degrees of freedom
  (7 observations deleted due to missingness)
Multiple R-squared:  0.7689,	Adjusted R-squared:  0.7029
F-statistic: 11.65 on 2 and 7 DF,  p-value: 0.005932


> ### RQ 2 C: Vocational Outcome ~ Group*Core Language (youth cohort)/Grammaticality Judgement (A’; all participants)/age of first words/phrases
> ######################
> ##### Have a job #####
> ######################
> ### ASD LAD ###
> mJobNotColl_ASDLAD.GJ = glm(edu_pro_4_par ~
+                          Group.NTref*aprime.log
+                 #+ age
+                       + vabs_soc_ss
+                        , family = binomial, data = dASDLAD.older18.nocoll)
> summary(mJobNotColl_ASDLAD.GJ)

Call:
glm(formula = edu_pro_4_par ~ Group.NTref * aprime.log + vabs_soc_ss,
    family = binomial, data = dASDLAD.older18.nocoll)

Deviance Residuals:
     Min        1Q    Median        3Q       Max
-1.61548  -0.00006   0.00000   0.00007   1.31255

Coefficients:
                        Estimate Std. Error z value Pr(>|z|)
(Intercept)            2.905e+02  1.035e+05   0.003    0.998
Group.NTref            5.786e+02  2.070e+05   0.003    0.998
aprime.log             1.821e+03  6.477e+05   0.003    0.998
vabs_soc_ss            1.165e-02  6.143e-02   0.190    0.850
Group.NTref:aprime.log 3.618e+03  1.295e+06   0.003    0.998

(Dispersion parameter for binomial family taken to be 1)

    Null deviance: 16.7515  on 13  degrees of freedom
Residual deviance:  5.8877  on  9  degrees of freedom
  (9 observations deleted due to missingness)
AIC: 15.888

Number of Fisher Scoring iterations: 23

>
> mJobNotColl_ASDLAD.Words = glm(edu_pro_4_par ~
+                          Group.NTref*FirstWords.months
+                 #+ age
+                       + vabs_soc_ss,
+                         family = binomial, data = dASDLAD.older18.nocoll)
> summary(mJobNotColl_ASDLAD.Words)

Call:
glm(formula = edu_pro_4_par ~ Group.NTref * FirstWords.months +
    vabs_soc_ss, family = binomial, data = dASDLAD.older18.nocoll)

Deviance Residuals:
     Min        1Q    Median        3Q       Max
-1.82905  -0.02535   0.07114   0.26887   1.18349

Coefficients:
                              Estimate Std. Error z value Pr(>|z|)
(Intercept)                   -31.4199    22.6402  -1.388    0.165
Group.NTref                    -9.5987    11.3652  -0.845    0.398
FirstWords.months               0.3974     0.4206   0.945    0.345
vabs_soc_ss                     0.2781     0.2190   1.270    0.204
Group.NTref:FirstWords.months   0.4948     0.8488   0.583    0.560

(Dispersion parameter for binomial family taken to be 1)

    Null deviance: 17.3975  on 14  degrees of freedom
Residual deviance:  5.9545  on 10  degrees of freedom
  (8 observations deleted due to missingness)
AIC: 15.955

Number of Fisher Scoring iterations: 9

>
> mJobNotColl_ASDLAD.Phrases = glm(edu_pro_4_par ~
+                          Group.NTref*FirstPhrases.months
+                 #+ age
+                       + vabs_soc_ss,
+                        family = binomial, data = dASDLAD.older18.nocoll)
> summary(mJobNotColl_ASDLAD.Phrases)

Call:
glm(formula = edu_pro_4_par ~ Group.NTref * FirstPhrases.months +
    vabs_soc_ss, family = binomial, data = dASDLAD.older18.nocoll)

Deviance Residuals:
       Min          1Q      Median          3Q         Max
-1.245e-05   0.000e+00   2.110e-08   9.290e-07   9.992e-06

Coefficients:
                                  Estimate Std. Error z value Pr(>|z|)
(Intercept)                       -448.181 965320.489       0        1
Group.NTref                        119.603 381458.926       0        1
FirstPhrases.months                  1.582   6482.599       0        1
vabs_soc_ss                          4.449   8992.074       0        1
Group.NTref:FirstPhrases.months     -6.216  15021.332       0        1

(Dispersion parameter for binomial family taken to be 1)

    Null deviance: 1.7397e+01  on 14  degrees of freedom
Residual deviance: 3.8309e-10  on 10  degrees of freedom
  (8 observations deleted due to missingness)
AIC: 10

Number of Fisher Scoring iterations: 25

>
>
>
>
>
> ### ASD NT ###
> mJobNotColl_ASDNT.GJ = glm(edu_pro_4_par ~
+                          Group.LADref*aprime.log
+                #+ age
+                       + vabs_soc_ss
+                        , family = binomial, data = dASDNT.older18.nocoll)
> summary(mJobNotColl_ASDNT.GJ)

Call:
glm(formula = edu_pro_4_par ~ Group.LADref * aprime.log + vabs_soc_ss,
    family = binomial, data = dASDNT.older18.nocoll)

Deviance Residuals:
    Min       1Q   Median       3Q      Max
-1.6580  -1.1407   0.6627   0.9699   1.2582

Coefficients:
                          Estimate Std. Error z value Pr(>|z|)
(Intercept)              1.5708428  5.7869757   0.271    0.786
Group.LADref            -1.2528896  2.5613823  -0.489    0.625
aprime.log               8.0619050  6.3387134   1.272    0.203
vabs_soc_ss              0.0006619  0.0583103   0.011    0.991
Group.LADref:aprime.log -6.6703006 12.7823121  -0.522    0.602

(Dispersion parameter for binomial family taken to be 1)

    Null deviance: 21.930  on 15  degrees of freedom
Residual deviance: 19.075  on 11  degrees of freedom
  (6 observations deleted due to missingness)
AIC: 29.075

Number of Fisher Scoring iterations: 4

>
> mJobNotColl_ASDNT.Words = glm(edu_pro_4_par ~
+                          Group.LADref*FirstWords.months
+                #+ age
+                       + vabs_soc_ss
+                        , family = binomial, data = dASDNT.older18.nocoll)
> summary(mJobNotColl_ASDNT.Words)

Call:
glm(formula = edu_pro_4_par ~ Group.LADref * FirstWords.months +
    vabs_soc_ss, family = binomial, data = dASDNT.older18.nocoll)

Deviance Residuals:
    Min       1Q   Median       3Q      Max
-1.9369  -0.4116   0.1725   0.6646   1.4086

Coefficients:
                               Estimate Std. Error z value Pr(>|z|)
(Intercept)                    -26.0886    16.9362  -1.540    0.123
Group.LADref                    -5.6964     6.4658  -0.881    0.378
FirstWords.months                0.1909     0.1576   1.211    0.226
vabs_soc_ss                      0.2423     0.1595   1.519    0.129
Group.LADref:FirstWords.months   0.1194     0.2684   0.445    0.657

(Dispersion parameter for binomial family taken to be 1)

    Null deviance: 20.190  on 14  degrees of freedom
Residual deviance: 12.046  on 10  degrees of freedom
  (7 observations deleted due to missingness)
AIC: 22.046

Number of Fisher Scoring iterations: 6

>
> mJobNotColl_ASDNT.Phrases = glm(edu_pro_4_par ~
+                          Group.LADref*FirstPhrases.months
+                #+ age
+                       + vabs_soc_ss
+                        , family = binomial, data = dASDNT.older18.nocoll)
> summary(mJobNotColl_ASDNT.Phrases)

Call:
glm(formula = edu_pro_4_par ~ Group.LADref * FirstPhrases.months +
    vabs_soc_ss, family = binomial, data = dASDNT.older18.nocoll)

Deviance Residuals:
    Min       1Q   Median       3Q      Max
-1.7590  -0.5587   0.1879   0.8307   1.4151

Coefficients:
                                 Estimate Std. Error z value Pr(>|z|)
(Intercept)                      -26.1959    16.6270  -1.576    0.115
Group.LADref                      -0.7175     6.4402  -0.111    0.911
FirstPhrases.months                0.2591     0.1575   1.644    0.100
vabs_soc_ss                        0.2105     0.1436   1.466    0.143
Group.LADref:FirstPhrases.months  -0.0414     0.2496  -0.166    0.868

(Dispersion parameter for binomial family taken to be 1)

    Null deviance: 23.035  on 16  degrees of freedom
Residual deviance: 15.245  on 12  degrees of freedom
  (5 observations deleted due to missingness)
AIC: 25.245

Number of Fisher Scoring iterations: 6

>
>
>
>
> ### LAD NT ###
> mJobNotColl_LADNT.GJ = glm(edu_pro_4_par ~
+                          Group.ASDref*aprime.log
+                #+ age
+                       + vabs_soc_ss
+                        , family = binomial, data = dLADNT.older18.nocoll)
> summary(mJobNotColl_LADNT.GJ)

Call:
glm(formula = edu_pro_4_par ~ Group.ASDref * aprime.log + vabs_soc_ss,
    family = binomial, data = dLADNT.older18.nocoll)

Deviance Residuals:
     Min        1Q    Median        3Q       Max
-1.71018  -0.00011   0.00000   0.72489   1.21037

Coefficients:
                          Estimate Std. Error z value Pr(>|z|)
(Intercept)              2.870e+02  6.158e+04   0.005    0.996
Group.ASDref            -5.427e+02  1.232e+05  -0.004    0.996
aprime.log               1.697e+03  3.854e+05   0.004    0.996
vabs_soc_ss             -1.392e-01  2.255e-01  -0.617    0.537
Group.ASDref:aprime.log -3.391e+03  7.708e+05  -0.004    0.996

(Dispersion parameter for binomial family taken to be 1)

    Null deviance: 21.270  on 17  degrees of freedom
Residual deviance: 12.749  on 13  degrees of freedom
  (3 observations deleted due to missingness)
AIC: 22.749

Number of Fisher Scoring iterations: 22

>
> mJobNotColl_LADNT.Words = glm(edu_pro_4_par ~
+                          Group.ASDref*FirstWords.months
+                #+ age
+                       + vabs_soc_ss
+                        , family = binomial, data = dLADNT.older18.nocoll)
> summary(mJobNotColl_LADNT.Words)

Call:
glm(formula = edu_pro_4_par ~ Group.ASDref * FirstWords.months +
    vabs_soc_ss, family = binomial, data = dLADNT.older18.nocoll)

Deviance Residuals:
     Min        1Q    Median        3Q       Max
-1.63732   0.00096   0.32199   0.69623   1.25348

Coefficients:
                               Estimate Std. Error z value Pr(>|z|)
(Intercept)                      7.8015    29.4685   0.265    0.791
Group.ASDref                     6.9862    11.1845   0.625    0.532
FirstWords.months                0.4321     0.4730   0.914    0.361
vabs_soc_ss                     -0.1099     0.2669  -0.412    0.680
Group.ASDref:FirstWords.months  -0.6865     0.9777  -0.702    0.483

(Dispersion parameter for binomial family taken to be 1)

    Null deviance: 19.069  on 17  degrees of freedom
Residual deviance: 14.723  on 13  degrees of freedom
  (3 observations deleted due to missingness)
AIC: 24.723

Number of Fisher Scoring iterations: 9

>
> mJobNotColl_LADNT.Phrases = glm(edu_pro_4_par ~
+                          Group.ASDref*FirstPhrases.months
+                #+ age
+                       + vabs_soc_ss
+                        , family = binomial, data = dLADNT.older18.nocoll)
> summary(mJobNotColl_LADNT.Phrases)

Call:
glm(formula = edu_pro_4_par ~ Group.ASDref * FirstPhrases.months +
    vabs_soc_ss, family = binomial, data = dLADNT.older18.nocoll)

Deviance Residuals:
    Min       1Q   Median       3Q      Max
-1.3199  -0.0001   0.0000   0.4515   1.4217

Coefficients:
                                   Estimate Std. Error z value Pr(>|z|)
(Intercept)                       3.897e+01  6.707e+03   0.006    0.995
Group.ASDref                     -6.852e+01  1.341e+04  -0.005    0.996
FirstPhrases.months              -4.309e-01  1.105e+02  -0.004    0.997
vabs_soc_ss                      -6.404e-02  2.255e-01  -0.284    0.776
Group.ASDref:FirstPhrases.months  1.157e+00  2.209e+02   0.005    0.996

(Dispersion parameter for binomial family taken to be 1)

    Null deviance: 21.270  on 17  degrees of freedom
Residual deviance: 11.913  on 13  degrees of freedom
  (3 observations deleted due to missingness)
AIC: 21.913

Number of Fisher Scoring iterations: 19


> ###Follow up
> mJobNotColl_ASD.GJ = glm(edu_pro_4_par ~ aprime.log
+                #+ age
+                       + vabs_soc_ss
+                      , family = binomial, data = dASD.older18.nocoll)
> summary(mJobNotColl_ASD.GJ)

Call:
glm(formula = edu_pro_4_par ~ aprime.log + vabs_soc_ss, family = binomial,
    data = dASD.older18.nocoll)

Deviance Residuals:
      1        2        7       10       11       12
-0.3625  -0.6700   0.8820   0.4438  -1.6155   1.3126

Coefficients:
            Estimate Std. Error z value Pr(>|z|)
(Intercept)  1.22036    5.79098   0.211    0.833
aprime.log  11.35783    9.37074   1.212    0.225
vabs_soc_ss  0.01165    0.06143   0.190    0.850

(Dispersion parameter for binomial family taken to be 1)

    Null deviance: 8.3178  on 5  degrees of freedom
Residual deviance: 5.8877  on 3  degrees of freedom
  (6 observations deleted due to missingness)
AIC: 11.888

Number of Fisher Scoring iterations: 4

>
> mJobNotColl_NT.GJ = glm(edu_pro_4_par ~ aprime.log
+                #+ age
+                       + vabs_soc_ss
+                        , family = binomial, data = dNT.older18.nocoll)
> summary(mJobNotColl_NT.GJ)

Call:
glm(formula = edu_pro_4_par ~ aprime.log + vabs_soc_ss, family = binomial,
    data = dNT.older18.nocoll)

Deviance Residuals:
    Min       1Q   Median       3Q      Max
-1.7102  -1.1360   0.6977   1.0456   1.2104

Coefficients:
            Estimate Std. Error z value Pr(>|z|)
(Intercept)  15.6174    23.7943   0.656    0.512
aprime.log    1.4822     9.8931   0.150    0.881
vabs_soc_ss  -0.1392     0.2255  -0.617    0.537

(Dispersion parameter for binomial family taken to be 1)

    Null deviance: 13.460  on 9  degrees of freedom
Residual deviance: 12.749  on 7  degrees of freedom
AIC: 18.749

Number of Fisher Scoring iterations: 4

>
> mJobNotColl_LAD.GJ = glm(edu_pro_4_par ~ aprime.log
+              #+ age
+                       + vabs_soc_ss
+                        , family = binomial, data = dLAD.older18.nocoll)
> summary(mJobNotColl_LAD.GJ)

Call:
glm(formula = edu_pro_4_par ~ aprime.log + vabs_soc_ss, family = binomial,
    data = dLAD.older18.nocoll)

Deviance Residuals:
         1           3           4           7           8           9          10          11
 8.152e-06   2.110e-08   9.375e-06   2.110e-08   2.166e-06   2.110e-08   2.110e-08  -9.140e-06

Coefficients:
              Estimate Std. Error z value Pr(>|z|)
(Intercept)    1555.05 4893632.70       0        1
aprime.log      804.18 2172732.35       0        1
vabs_soc_ss     -13.14   43794.34       0        1

(Dispersion parameter for binomial family taken to be 1)

    Null deviance: 6.0283e+00  on 7  degrees of freedom
Residual deviance: 2.4257e-10  on 5  degrees of freedom
  (3 observations deleted due to missingness)
AIC: 6

Number of Fisher Scoring iterations: 25

>
>
>
> #Words
> mJobNotColl_ASD.Words = glm(edu_pro_4_par ~ FirstWords.months
+                #+ age
+                       + vabs_soc_ss
+                        , family = binomial, data = dASD.older18.nocoll)
> summary(mJobNotColl_ASD.Words)

Call:
glm(formula = edu_pro_4_par ~ FirstWords.months + vabs_soc_ss,
    family = binomial, data = dASD.older18.nocoll)

Deviance Residuals:
         1           6           7           9          10          11
-1.101e-05   2.110e-08   2.110e-08  -2.110e-08   1.277e-05  -5.134e-06

Coefficients:
                    Estimate Std. Error z value Pr(>|z|)
(Intercept)         -263.023 623825.762       0        1
FirstWords.months      1.236   9131.098       0        1
vabs_soc_ss            2.851   5981.802       0        1

(Dispersion parameter for binomial family taken to be 1)

    Null deviance: 8.3178e+00  on 5  degrees of freedom
Residual deviance: 3.1076e-10  on 3  degrees of freedom
  (6 observations deleted due to missingness)
AIC: 6

Number of Fisher Scoring iterations: 24

>
> mJobNotColl_NT.Words = glm(edu_pro_4_par ~ FirstWords.months
+                #+ age
+                       + vabs_soc_ss
+                        , family = binomial, data = dNT.older18.nocoll)
> summary(mJobNotColl_NT.Words)

Call:
glm(formula = edu_pro_4_par ~ FirstWords.months + vabs_soc_ss,
    family = binomial, data = dNT.older18.nocoll)

Deviance Residuals:
    Min       1Q   Median       3Q      Max
-1.4952  -1.3135   0.5372   1.0096   1.3105

Coefficients:
                  Estimate Std. Error z value Pr(>|z|)
(Intercept)       -2.78720   30.25786  -0.092    0.927
FirstWords.months  0.14320    0.20107   0.712    0.476
vabs_soc_ss        0.01343    0.26439   0.051    0.959

(Dispersion parameter for binomial family taken to be 1)

    Null deviance: 11.457  on 8  degrees of freedom
Residual deviance: 10.602  on 6  degrees of freedom
  (1 observation deleted due to missingness)
AIC: 16.602

Number of Fisher Scoring iterations: 4

>
> mJobNotColl_LAD.Words = glm(edu_pro_4_par ~ FirstWords.months
+                #+ age
+                       + vabs_soc_ss
+                      , family = binomial, data = dLAD.older18.nocoll)
> summary(mJobNotColl_LAD.Words)

Call:
glm(formula = edu_pro_4_par ~ FirstWords.months + vabs_soc_ss,
    family = binomial, data = dLAD.older18.nocoll)

Deviance Residuals:
       Min          1Q      Median          3Q         Max
-3.017e-05   2.100e-08   2.100e-08   2.100e-08   3.672e-05

Coefficients:
                    Estimate Std. Error z value Pr(>|z|)
(Intercept)          2179.65 3284292.00   0.001    0.999
FirstWords.months      21.31   18398.14   0.001    0.999
vabs_soc_ss           -22.14   30726.63  -0.001    0.999

(Dispersion parameter for binomial family taken to be 1)

    Null deviance: 6.2790e+00  on 8  degrees of freedom
Residual deviance: 2.3712e-09  on 6  degrees of freedom
  (2 observations deleted due to missingness)
AIC: 6

Number of Fisher Scoring iterations: 25

>
>
>
> #Phrases
> mJobNotColl_ASD.Phrases = glm(edu_pro_4_par ~ FirstPhrases.months
+                #+ age
+                       + vabs_soc_ss
+                        , family = binomial, data = dASD.older18.nocoll)
> summary(mJobNotColl_ASD.Phrases)

Call:
glm(formula = edu_pro_4_par ~ FirstPhrases.months + vabs_soc_ss,
    family = binomial, data = dASD.older18.nocoll)

Deviance Residuals:
         1           6           7           9          10          11          12
-1.102e-05   2.110e-08   2.110e-08  -2.110e-08   8.942e-06  -2.110e-08   7.383e-06

Coefficients:
                      Estimate Std. Error z value Pr(>|z|)
(Intercept)         -5.130e+02  1.189e+06       0        1
FirstPhrases.months  4.736e+00  1.299e+04       0        1
vabs_soc_ss          4.493e+00  1.013e+04       0        1

(Dispersion parameter for binomial family taken to be 1)

    Null deviance: 9.5607e+00  on 6  degrees of freedom
Residual deviance: 2.5589e-10  on 4  degrees of freedom
  (5 observations deleted due to missingness)
AIC: 6

Number of Fisher Scoring iterations: 25

>
> mJobNotColl_NT.Phrases = glm(edu_pro_4_par ~ FirstPhrases.months
+                #+ age
+                       + vabs_soc_ss
+                        , family = binomial, data = dNT.older18.nocoll)
> summary(mJobNotColl_NT.Phrases)

Call:
glm(formula = edu_pro_4_par ~ FirstPhrases.months + vabs_soc_ss,
    family = binomial, data = dNT.older18.nocoll)

Deviance Residuals:
    Min       1Q   Median       3Q      Max
-1.3199  -1.1775   0.3849   1.0224   1.4217

Coefficients:
                    Estimate Std. Error z value Pr(>|z|)
(Intercept)          4.71226   25.42340   0.185    0.853
FirstPhrases.months  0.14782    0.18715   0.790    0.430
vabs_soc_ss         -0.06404    0.22550  -0.284    0.776

(Dispersion parameter for binomial family taken to be 1)

    Null deviance: 13.460  on 9  degrees of freedom
Residual deviance: 11.913  on 7  degrees of freedom
AIC: 17.913

Number of Fisher Scoring iterations: 5

>
> mJobNotColl_LAD.Phrases = glm(edu_pro_4_par ~ FirstPhrases.months
+                #+ age
+                       + vabs_soc_ss
+                        , family = binomial, data = dLAD.older18.nocoll)
> summary(mJobNotColl_LAD.Phrases)

Call:
glm(formula = edu_pro_4_par ~ FirstPhrases.months + vabs_soc_ss,
    family = binomial, data = dLAD.older18.nocoll)

Deviance Residuals:
         1           3           6           7           8           9          10          11
 2.701e-06   2.110e-08   2.110e-08   2.110e-08   2.110e-08   1.274e-05   6.230e-06  -8.545e-06

Coefficients:
                      Estimate Std. Error z value Pr(>|z|)
(Intercept)         -2.814e+02  2.190e+06       0        1
FirstPhrases.months -1.313e+00  3.342e+03       0        1
vabs_soc_ss          3.342e+00  2.114e+04       0        1

(Dispersion parameter for binomial family taken to be 1)

    Null deviance: 6.0283e+00  on 7  degrees of freedom
Residual deviance: 2.8156e-10  on 5  degrees of freedom
  (3 observations deleted due to missingness)
AIC: 6

Number of Fisher Scoring iterations: 24


> ########################################
> ##### Hours per week spent working #####
> ########################################
> ### ASD LAD ###
> mJobHoursNotColl_ASDLAD.GJ = lm(HoursWorking.log ~
+                           Group.NTref*aprime.log
+                  #+ age
+                       + vabs_soc_ss
+                         , data = dASDLAD.older18.nocoll)
> summary(mJobHoursNotColl_ASDLAD.GJ)

Call:
lm(formula = HoursWorking.log ~ Group.NTref * aprime.log + vabs_soc_ss,
    data = dASDLAD.older18.nocoll)

Residuals:
        4         6         7        13        15        16        18        19        20        23
 0.015117 -0.007072  0.043987 -0.026775 -0.004359  0.042327  0.006628  0.019193 -0.109192  0.020147

Coefficients:
                        Estimate Std. Error t value Pr(>|t|)
(Intercept)             5.308472   0.212768  24.950 1.93e-06 ***
Group.NTref             0.513903   0.081750   6.286   0.0015 **
aprime.log              0.734515   0.321528   2.284   0.0711 .
vabs_soc_ss            -0.017020   0.002035  -8.362   0.0004 ***
Group.NTref:aprime.log -0.557554   0.626243  -0.890   0.4141
---
Signif. codes:  0 ‘***’ 0.001 ‘**’ 0.01 ‘*’ 0.05 ‘.’ 0.1 ‘ ’ 1

Residual standard error: 0.05913 on 5 degrees of freedom
  (13 observations deleted due to missingness)
Multiple R-squared:  0.9597,	Adjusted R-squared:  0.9274
F-statistic: 29.76 on 4 and 5 DF,  p-value: 0.001109

>
> mJobHoursNotColl_ASDLAD.Words = lm(HoursWorking.log ~
+                           Group.NTref*FirstWords.months
+                  #+ age
+                       + vabs_soc_ss
+                         , data = dASDLAD.older18.nocoll)
> summary(mJobHoursNotColl_ASDLAD.Words)

Call:
lm(formula = HoursWorking.log ~ Group.NTref * FirstWords.months +
    vabs_soc_ss, data = dASDLAD.older18.nocoll)

Residuals:
      Min        1Q    Median        3Q       Max
-0.008587 -0.006009 -0.001073  0.003261  0.015409

Coefficients:
                                Estimate Std. Error t value Pr(>|t|)
(Intercept)                    2.9917303  0.0690534  43.325 1.01e-08 ***
Group.NTref                    0.8938916  0.0158343  56.453 2.08e-09 ***
FirstWords.months              0.0162210  0.0004319  37.558 2.38e-08 ***
vabs_soc_ss                    0.0023369  0.0006177   3.783  0.00914 **
Group.NTref:FirstWords.months -0.0325448  0.0008770 -37.109 2.56e-08 ***
---
Signif. codes:  0 ‘***’ 0.001 ‘**’ 0.01 ‘*’ 0.05 ‘.’ 0.1 ‘ ’ 1

Residual standard error: 0.009743 on 6 degrees of freedom
  (12 observations deleted due to missingness)
Multiple R-squared:  0.9987,	Adjusted R-squared:  0.9978
F-statistic:  1155 on 4 and 6 DF,  p-value: 8.727e-09

>
> mJobHoursNotColl_ASDLAD.Phrases = lm(HoursWorking.log ~
+                           Group.NTref*FirstPhrases.months
+                  #+ age
+                       + vabs_soc_ss
+                         , data = dASDLAD.older18.nocoll)
> summary(mJobHoursNotColl_ASDLAD.Phrases)

Call:
lm(formula = HoursWorking.log ~ Group.NTref * FirstPhrases.months +
    vabs_soc_ss, data = dASDLAD.older18.nocoll)

Residuals:
      Min        1Q    Median        3Q       Max
-0.127825 -0.067336  0.003518  0.060629  0.158947

Coefficients:
                                 Estimate Std. Error t value Pr(>|t|)
(Intercept)                      5.676296   0.598113   9.490  7.8e-05 ***
Group.NTref                     -0.254663   0.312077  -0.816  0.44567
FirstPhrases.months             -0.009499   0.004939  -1.923  0.10280
vabs_soc_ss                     -0.017714   0.004750  -3.729  0.00975 **
Group.NTref:FirstPhrases.months  0.021645   0.010329   2.096  0.08098 .
---
Signif. codes:  0 ‘***’ 0.001 ‘**’ 0.01 ‘*’ 0.05 ‘.’ 0.1 ‘ ’ 1

Residual standard error: 0.1164 on 6 degrees of freedom
  (12 observations deleted due to missingness)
Multiple R-squared:  0.8148,	Adjusted R-squared:  0.6914
F-statistic:   6.6 on 4 and 6 DF,  p-value: 0.02188

>
>
>
>
> ### ASD NT ###
> mJobHoursNotColl_ASDNT.GJ = lm(HoursWorking.log ~
+                           Group.LADref*aprime.log
+                 #+ age
+                       + vabs_soc_ss
+                         , data = dASDNT.older18.nocoll)
> summary(mJobHoursNotColl_ASDNT.GJ)

Call:
lm(formula = HoursWorking.log ~ Group.LADref * aprime.log + vabs_soc_ss,
    data = dASDNT.older18.nocoll)

Residuals:
         9         13         14         15         16         17         18         19         22
-0.0241346  0.0059740  0.1022944 -0.0088048  0.0028168 -0.0005449 -0.1665776  0.0708161  0.0181606

Coefficients:
                         Estimate Std. Error t value Pr(>|t|)
(Intercept)              5.220368   0.366270  14.253 0.000141 ***
Group.LADref             0.314782   0.134265   2.344 0.078990 .
aprime.log               0.129698   0.455449   0.285 0.789956
vabs_soc_ss             -0.017140   0.003613  -4.744 0.009013 **
Group.LADref:aprime.log -1.778734   0.943526  -1.885 0.132483
---
Signif. codes:  0 ‘***’ 0.001 ‘**’ 0.01 ‘*’ 0.05 ‘.’ 0.1 ‘ ’ 1

Residual standard error: 0.1052 on 4 degrees of freedom
  (13 observations deleted due to missingness)
Multiple R-squared:  0.8907,	Adjusted R-squared:  0.7813
F-statistic: 8.146 on 4 and 4 DF,  p-value: 0.03325

>
> mJobHoursNotColl_ASDNT.Words = lm(HoursWorking.log ~
+                           Group.LADref*FirstWords.months
+                 #+ age
+                       + vabs_soc_ss
+                         , data = dASDNT.older18.nocoll)
> summary(mJobHoursNotColl_ASDNT.Words)

Call:
lm(formula = HoursWorking.log ~ Group.LADref * FirstWords.months +
    vabs_soc_ss, data = dASDNT.older18.nocoll)

Residuals:
        7         9        13        14        15        16        17        18        19
 0.014887 -0.001654 -0.013233  0.057560 -0.121446  0.041073  0.077624 -0.086664  0.031852

Coefficients:
                                Estimate Std. Error t value Pr(>|t|)
(Intercept)                     3.038491   0.791701   3.838  0.01849 *
Group.LADref                    0.865334   0.165489   5.229  0.00639 **
FirstWords.months               0.015143   0.006176   2.452  0.07030 .
vabs_soc_ss                     0.001807   0.006825   0.265  0.80426
Group.LADref:FirstWords.months -0.033975   0.009122  -3.724  0.02039 *
---
Signif. codes:  0 ‘***’ 0.001 ‘**’ 0.01 ‘*’ 0.05 ‘.’ 0.1 ‘ ’ 1

Residual standard error: 0.09314 on 4 degrees of freedom
  (13 observations deleted due to missingness)
Multiple R-squared:   0.91,	Adjusted R-squared:   0.82
F-statistic: 10.11 on 4 and 4 DF,  p-value: 0.02283

>
> mJobHoursNotColl_ASDLAD.Phrases = lm(HoursWorking.log ~
+                           Group.NTref*FirstPhrases.months
+                  #+ age
+                       + vabs_soc_ss
+                         , data = dASDLAD.older18.nocoll)
> summary(mJobHoursNotColl_ASDLAD.Phrases)

Call:
lm(formula = HoursWorking.log ~ Group.NTref * FirstPhrases.months +
    vabs_soc_ss, data = dASDLAD.older18.nocoll)

Residuals:
      Min        1Q    Median        3Q       Max
-0.127825 -0.067336  0.003518  0.060629  0.158947

Coefficients:
                                 Estimate Std. Error t value Pr(>|t|)
(Intercept)                      5.676296   0.598113   9.490  7.8e-05 ***
Group.NTref                     -0.254663   0.312077  -0.816  0.44567
FirstPhrases.months             -0.009499   0.004939  -1.923  0.10280
vabs_soc_ss                     -0.017714   0.004750  -3.729  0.00975 **
Group.NTref:FirstPhrases.months  0.021645   0.010329   2.096  0.08098 .
---
Signif. codes:  0 ‘***’ 0.001 ‘**’ 0.01 ‘*’ 0.05 ‘.’ 0.1 ‘ ’ 1

Residual standard error: 0.1164 on 6 degrees of freedom
  (12 observations deleted due to missingness)
Multiple R-squared:  0.8148,	Adjusted R-squared:  0.6914
F-statistic:   6.6 on 4 and 6 DF,  p-value: 0.02188

>
>
>
>
>
> ### LAD NT ###
> mJobHoursNotColl_LADNT.GJ = lm(HoursWorking.log ~
+                           Group.ASDref*aprime.log
+                 #+ age
+                       + vabs_soc_ss
+                         , data = dLADNT.older18.nocoll)
> summary(mJobHoursNotColl_LADNT.GJ)

Call:
lm(formula = HoursWorking.log ~ Group.ASDref * aprime.log + vabs_soc_ss,
    data = dLADNT.older18.nocoll)

Residuals:
      Min        1Q    Median        3Q       Max
-0.109992 -0.000584  0.001585  0.005700  0.077086

Coefficients:
                         Estimate Std. Error t value Pr(>|t|)
(Intercept)              3.792326   0.620305   6.114 0.000285 ***
Group.ASDref            -0.103104   0.078860  -1.307 0.227387
aprime.log              -0.171485   0.309832  -0.553 0.595063
vabs_soc_ss             -0.001406   0.005780  -0.243 0.813954
Group.ASDref:aprime.log -0.418256   0.685702  -0.610 0.558807
---
Signif. codes:  0 ‘***’ 0.001 ‘**’ 0.01 ‘*’ 0.05 ‘.’ 0.1 ‘ ’ 1

Residual standard error: 0.06232 on 8 degrees of freedom
  (8 observations deleted due to missingness)
Multiple R-squared:  0.3048,	Adjusted R-squared:  -0.04283
F-statistic: 0.8768 on 4 and 8 DF,  p-value: 0.5184

>
> mJobHoursNotColl_LADNT.Words = lm(HoursWorking.log ~
+                           Group.ASDref*FirstWords.months
+                 #+ age
+                         , data = dLADNT.older18.nocoll)
> summary(mJobHoursNotColl_LADNT.Words)

Call:
lm(formula = HoursWorking.log ~ Group.ASDref * FirstWords.months,
    data = dLADNT.older18.nocoll)

Residuals:
     Min       1Q   Median       3Q      Max
-0.11965  0.00000  0.00000  0.01437  0.07396

Coefficients:
                                Estimate Std. Error t value Pr(>|t|)
(Intercept)                     3.684404   0.036075 102.132   <2e-16 ***
Group.ASDref                   -0.008950   0.072150  -0.124    0.904
FirstWords.months              -0.001413   0.001997  -0.708    0.494
Group.ASDref:FirstWords.months -0.002826   0.003994  -0.708    0.494
---
Signif. codes:  0 ‘***’ 0.001 ‘**’ 0.01 ‘*’ 0.05 ‘.’ 0.1 ‘ ’ 1

Residual standard error: 0.05509 on 11 degrees of freedom
  (6 observations deleted due to missingness)
Multiple R-squared:  0.2709,	Adjusted R-squared:  0.07206
F-statistic: 1.362 on 3 and 11 DF,  p-value: 0.3051

>
> mJobHoursNotColl_LADNT.Phrases = lm(HoursWorking.log ~
+                           Group.ASDref*FirstPhrases.months
+                 #+ age
+                       + vabs_soc_ss
+                         , data = dLADNT.older18.nocoll)
> summary(mJobHoursNotColl_LADNT.Phrases)

Call:
lm(formula = HoursWorking.log ~ Group.ASDref * FirstPhrases.months +
    vabs_soc_ss, data = dLADNT.older18.nocoll)

Residuals:
      Min        1Q    Median        3Q       Max
-0.107367 -0.011257  0.006443  0.026059  0.047669

Coefficients:
                                  Estimate Std. Error t value Pr(>|t|)
(Intercept)                       4.502569   0.566056   7.954 4.55e-05 ***
Group.ASDref                      0.126644   0.093922   1.348   0.2145
FirstPhrases.months              -0.003792   0.001757  -2.158   0.0630 .
vabs_soc_ss                      -0.007146   0.005146  -1.389   0.2024
Group.ASDref:FirstPhrases.months -0.008652   0.003907  -2.215   0.0577 .
---
Signif. codes:  0 ‘***’ 0.001 ‘**’ 0.01 ‘*’ 0.05 ‘.’ 0.1 ‘ ’ 1

Residual standard error: 0.04909 on 8 degrees of freedom
  (8 observations deleted due to missingness)
Multiple R-squared:  0.5687,	Adjusted R-squared:  0.353
F-statistic: 2.637 on 4 and 8 DF,  p-value: 0.1133


> ###Follow up
>
> #aprime.log
> mJobHoursNotColl_ASD.GJ = lm(HoursWorking.log ~ aprime.log
+                 #+ age
+                       + vabs_soc_ss
+                         , data = dASD.older18.nocoll)
> summary(mJobHoursNotColl_ASD.GJ)

Call:
lm(formula = HoursWorking.log ~ aprime.log + vabs_soc_ss, data = dASD.older18.nocoll)

Residuals:
ALL 3 residuals are 0: no residual degrees of freedom!

Coefficients:
            Estimate Std. Error t value Pr(>|t|)
(Intercept)  5.16769        NaN     NaN      NaN
aprime.log   1.07183        NaN     NaN      NaN
vabs_soc_ss -0.01824        NaN     NaN      NaN

Residual standard error: NaN on 0 degrees of freedom
  (9 observations deleted due to missingness)
Multiple R-squared:      1,	Adjusted R-squared:    NaN
F-statistic:   NaN on 2 and 0 DF,  p-value: NA

>
> mJobHoursNotColl_NT.GJ = lm(HoursWorking.log ~ aprime.log
+                 #+ age
+                       + vabs_soc_ss
+                         , data = dNT.older18.nocoll)
> summary(mJobHoursNotColl_NT.GJ)

Call:
lm(formula = HoursWorking.log ~ aprime.log + vabs_soc_ss, data = dNT.older18.nocoll)

Residuals:
        4         5         6         7         8         9
 0.079202 -0.075137  0.005458  0.062843 -0.114742  0.042376

Coefficients:
             Estimate Std. Error t value Pr(>|t|)
(Intercept)  3.878186   1.365017   2.841   0.0656 .
aprime.log  -0.412431   0.644770  -0.640   0.5679
vabs_soc_ss -0.002727   0.013099  -0.208   0.8484
---
Signif. codes:  0 ‘***’ 0.001 ‘**’ 0.01 ‘*’ 0.05 ‘.’ 0.1 ‘ ’ 1

Residual standard error: 0.1014 on 3 degrees of freedom
  (4 observations deleted due to missingness)
Multiple R-squared:  0.1237,	Adjusted R-squared:  -0.4604
F-statistic: 0.2118 on 2 and 3 DF,  p-value: 0.8203

>
> mJobHoursNotColl_LAD.GJ = lm(HoursWorking.log ~ aprime.log
+                 #+ age
+                       + vabs_soc_ss
+                         , data = dLADNT.older18.nocoll)
> summary(mJobHoursNotColl_LAD.GJ)

Call:
lm(formula = HoursWorking.log ~ aprime.log + vabs_soc_ss, data = dLADNT.older18.nocoll)

Residuals:
     Min       1Q   Median       3Q      Max
-0.13624  0.01268  0.02054  0.03131  0.04136

Coefficients:
             Estimate Std. Error t value Pr(>|t|)
(Intercept)  3.471311   0.580631   5.979 0.000136 ***
aprime.log  -0.124646   0.298005  -0.418 0.684593
vabs_soc_ss  0.001675   0.005429   0.309 0.763977
---
Signif. codes:  0 ‘***’ 0.001 ‘**’ 0.01 ‘*’ 0.05 ‘.’ 0.1 ‘ ’ 1

Residual standard error: 0.06587 on 10 degrees of freedom
  (8 observations deleted due to missingness)
Multiple R-squared:  0.02939,	Adjusted R-squared:  -0.1647
F-statistic: 0.1514 on 2 and 10 DF,  p-value: 0.8614

>
> #Words
> mJobHoursNotColl_ASD.Words = lm(HoursWorking.log ~ FirstWords.months
+                 #+ age
+                       + vabs_soc_ss
+                        , data = dASD.older18.nocoll)
> summary(mJobHoursNotColl_ASD.Words)

Call:
lm(formula = HoursWorking.log ~ FirstWords.months + vabs_soc_ss,
    data = dASD.older18.nocoll)

Residuals:
ALL 3 residuals are 0: no residual degrees of freedom!

Coefficients:
                  Estimate Std. Error t value Pr(>|t|)
(Intercept)       2.431940        NaN     NaN      NaN
FirstWords.months 0.033164        NaN     NaN      NaN
vabs_soc_ss       0.003316        NaN     NaN      NaN

Residual standard error: NaN on 0 degrees of freedom
  (9 observations deleted due to missingness)
Multiple R-squared:      1,	Adjusted R-squared:    NaN
F-statistic:   NaN on 2 and 0 DF,  p-value: NA

>
> mJobHoursNotColl_NT.Words = lm(HoursWorking.log ~ FirstWords.months
+                 #+ age
+                       + vabs_soc_ss
+                         , data = dNT.older18.nocoll)
> summary(mJobHoursNotColl_NT.Words)

Call:
lm(formula = HoursWorking.log ~ FirstWords.months + vabs_soc_ss,
    data = dNT.older18.nocoll)

Residuals:
       4        5        6        7        8        9
 0.08800 -0.09734  0.06518  0.02835 -0.07417 -0.01003

Coefficients:
                  Estimate Std. Error t value Pr(>|t|)
(Intercept)        6.27656    3.37496    1.86    0.160
FirstWords.months -0.01504    0.01709   -0.88    0.444
vabs_soc_ss       -0.02248    0.02920   -0.77    0.497

Residual standard error: 0.09639 on 3 degrees of freedom
  (4 observations deleted due to missingness)
Multiple R-squared:  0.2085,	Adjusted R-squared:  -0.3191
F-statistic: 0.3952 on 2 and 3 DF,  p-value: 0.7042

>
> mJobHoursNotColl_LAD.Words = lm(HoursWorking.log ~ FirstWords.months
+                 #+ age
+                       + vabs_soc_ss
+                         , data = dLADNT.older18.nocoll)
> summary(mJobHoursNotColl_LAD.Words)

Call:
lm(formula = HoursWorking.log ~ FirstWords.months + vabs_soc_ss,
    data = dLADNT.older18.nocoll)

Residuals:
     Min       1Q   Median       3Q      Max
-0.14283  0.01891  0.02005  0.02469  0.03240

Coefficients:
                   Estimate Std. Error t value Pr(>|t|)
(Intercept)       3.4921490  0.5713888   6.112 7.61e-05 ***
FirstWords.months 0.0001475  0.0017948   0.082    0.936
vabs_soc_ss       0.0015934  0.0052560   0.303    0.767
---
Signif. codes:  0 ‘***’ 0.001 ‘**’ 0.01 ‘*’ 0.05 ‘.’ 0.1 ‘ ’ 1

Residual standard error: 0.06389 on 11 degrees of freedom
  (7 observations deleted due to missingness)
Multiple R-squared:  0.008322,	Adjusted R-squared:  -0.172
F-statistic: 0.04615 on 2 and 11 DF,  p-value: 0.9551

>
> #Phrases
> mJobHoursNotColl_ASD.Phrases = lm(HoursWorking.log ~ FirstPhrases.months
+                 #+ age
+                       + vabs_soc_ss
+                         , data = dASD.older18.nocoll)
> summary(mJobHoursNotColl_ASD.Phrases)

Call:
lm(formula = HoursWorking.log ~ FirstPhrases.months + vabs_soc_ss,
    data = dASD.older18.nocoll)

Residuals:
       6        7       10       12
 0.16257 -0.09044 -0.14081  0.06869

Coefficients:
                    Estimate Std. Error t value Pr(>|t|)
(Intercept)          6.09660    1.54838   3.937    0.158
FirstPhrases.months -0.02314    0.01942  -1.191    0.445
vabs_soc_ss         -0.01980    0.01049  -1.887    0.310

Residual standard error: 0.2432 on 1 degrees of freedom
  (8 observations deleted due to missingness)
Multiple R-squared:  0.7842,	Adjusted R-squared:  0.3526
F-statistic: 1.817 on 2 and 1 DF,  p-value: 0.4646

>
> mJobHoursNotColl_NT.Phrases = lm(HoursWorking.log ~ FirstPhrases.months
+                 #+ age
+                       + vabs_soc_ss
+                         , data = dNT.older18.nocoll)
> summary(mJobHoursNotColl_NT.Phrases)

Call:
lm(formula = HoursWorking.log ~ FirstPhrases.months + vabs_soc_ss,
    data = dNT.older18.nocoll)

Residuals:
        4         5         6         7         8         9
 0.020487 -0.071972  0.048272 -0.020990  0.018207  0.005996

Coefficients:
                     Estimate Std. Error t value Pr(>|t|)
(Intercept)          6.420838   1.172075   5.478   0.0120 *
FirstPhrases.months -0.014092   0.004687  -3.007   0.0574 .
vabs_soc_ss         -0.023353   0.010227  -2.284   0.1066
---
Signif. codes:  0 ‘***’ 0.001 ‘**’ 0.01 ‘*’ 0.05 ‘.’ 0.1 ‘ ’ 1

Residual standard error: 0.05397 on 3 degrees of freedom
  (4 observations deleted due to missingness)
Multiple R-squared:  0.7519,	Adjusted R-squared:  0.5865
F-statistic: 4.545 on 2 and 3 DF,  p-value: 0.1236

>
> mJobHoursNotColl_LAD.Phrases = lm(HoursWorking.log ~ FirstPhrases.months
+                 #+ age
+                         , data = dLAD.older18.nocoll)
> summary(mJobHoursNotColl_LAD.Phrases)

Call:
lm(formula = HoursWorking.log ~ FirstPhrases.months, data = dLAD.older18.nocoll)

Residuals:
       Min         1Q     Median         3Q        Max
-2.724e-15 -2.233e-17  2.977e-16  2.977e-16  1.578e-15

Coefficients:
                      Estimate Std. Error    t value Pr(>|t|)
(Intercept)          3.689e+00  1.373e-15  2.686e+15   <2e-16 ***
FirstPhrases.months -5.334e-17  4.746e-17 -1.124e+00    0.304
---
Signif. codes:  0 ‘***’ 0.001 ‘**’ 0.01 ‘*’ 0.05 ‘.’ 0.1 ‘ ’ 1

Residual standard error: 1.308e-15 on 6 degrees of freedom
  (3 observations deleted due to missingness)
Multiple R-squared:  0.5183,	Adjusted R-squared:  0.438
F-statistic: 6.456 on 1 and 6 DF,  p-value: 0.04403
```

**NO SOCIALIZATION –**

FOR SMALL EDUCATION/VOCATION SAMPLES and FOLLOW-UP ANLAYSES

```
> #### Surveys
> ###############
> ##### ALL #####
> ###############
>
> #QoLLifeSatSocSupp.log
> ### ASD LAD ###
> mSurvey_ASDLAD.GJ = lm(QoLLifeSatSocSupp.log ~
+                           Group.NTref*aprime.log
+                  #+ age
+                       #+ vabs_soc_ss
+                         , data = dASDLAD.older18)
> summary(mSurvey_ASDLAD.GJ)

Call:
lm(formula = QoLLifeSatSocSupp.log ~ Group.NTref * aprime.log,
    data = dASDLAD.older18)

Residuals:
     Min       1Q   Median       3Q      Max
-0.45727 -0.10697  0.03096  0.10926  0.26634

Coefficients:
                       Estimate Std. Error t value Pr(>|t|)
(Intercept)             4.68241    0.05220  89.702   <2e-16 ***
Group.NTref             0.18759    0.10440   1.797   0.0807 .
aprime.log             -0.37832    0.28930  -1.308   0.1993
Group.NTref:aprime.log  0.08361    0.57860   0.145   0.8859
---
Signif. codes:  0 ‘***’ 0.001 ‘**’ 0.01 ‘*’ 0.05 ‘.’ 0.1 ‘ ’ 1

Residual standard error: 0.182 on 36 degrees of freedom
  (15 observations deleted due to missingness)
Multiple R-squared:  0.1799,	Adjusted R-squared:  0.1115
F-statistic: 2.632 on 3 and 36 DF,  p-value: 0.06479

>
> mSurvey_ASDLAD.Words = lm(QoLLifeSatSocSupp.log ~
+                           Group.NTref*FirstWords.months
+                  #+ age
+                       #+ vabs_soc_ss
+                         , data = dASDLAD.older18)
> summary(mSurvey_ASDLAD.Words)

Call:
lm(formula = QoLLifeSatSocSupp.log ~ Group.NTref * FirstWords.months,
    data = dASDLAD.older18)

Residuals:
     Min       1Q   Median       3Q      Max
-0.42350 -0.07507  0.00891  0.13508  0.34165

Coefficients:
                               Estimate Std. Error t value Pr(>|t|)
(Intercept)                    4.622103   0.076090  60.745   <2e-16 ***
Group.NTref                    0.204903   0.152181   1.346    0.190
FirstWords.months              0.006800   0.004306   1.579    0.127
Group.NTref:FirstWords.months -0.005959   0.008611  -0.692    0.495
---
Signif. codes:  0 ‘***’ 0.001 ‘**’ 0.01 ‘*’ 0.05 ‘.’ 0.1 ‘ ’ 1

Residual standard error: 0.1836 on 25 degrees of freedom
  (26 observations deleted due to missingness)
Multiple R-squared:  0.2128,	Adjusted R-squared:  0.1183
F-statistic: 2.253 on 3 and 25 DF,  p-value: 0.107

>
> mSurvey_ASDLAD.Phrases = lm(QoLLifeSatSocSupp.log ~
+                           Group.NTref*FirstPhrases.months
+                  #+ age
+                       #+ vabs_soc_ss
+                         , data = dASDLAD.older18)
> summary(mSurvey_ASDLAD.Phrases)

Call:
lm(formula = QoLLifeSatSocSupp.log ~ Group.NTref * FirstPhrases.months,
    data = dASDLAD.older18)

Residuals:
     Min       1Q   Median       3Q      Max
-0.47892 -0.08345  0.00926  0.14168  0.28530

Coefficients:
                                 Estimate Std. Error t value Pr(>|t|)
(Intercept)                      4.688710   0.104368  44.925   <2e-16 ***
Group.NTref                     -0.003708   0.208736  -0.018    0.986
FirstPhrases.months              0.001280   0.003144   0.407    0.687
Group.NTref:FirstPhrases.months  0.005665   0.006288   0.901    0.376
---
Signif. codes:  0 ‘***’ 0.001 ‘**’ 0.01 ‘*’ 0.05 ‘.’ 0.1 ‘ ’ 1

Residual standard error: 0.1877 on 25 degrees of freedom
  (26 observations deleted due to missingness)
Multiple R-squared:  0.233,	Adjusted R-squared:  0.1409
F-statistic: 2.531 on 3 and 25 DF,  p-value: 0.08

>
>
>
>
> ### ASD NT ###
> mSurvey_ASDNT.GJ = lm(QoLLifeSatSocSupp.log ~
+                           Group.LADref*aprime.log
+                 #+ age
+                       #+ vabs_soc_ss
+                         , data = dASDNT.older18)
> summary(mSurvey_ASDNT.GJ)

Call:
lm(formula = QoLLifeSatSocSupp.log ~ Group.LADref * aprime.log,
    data = dASDNT.older18)

Residuals:
     Min       1Q   Median       3Q      Max
-0.47771 -0.07667  0.05853  0.12510  0.26634

Coefficients:
                        Estimate Std. Error t value Pr(>|t|)
(Intercept)              4.70189    0.05576  84.322   <2e-16 ***
Group.LADref             0.22655    0.11152   2.031   0.0499 *
aprime.log              -0.15045    0.31300  -0.481   0.6337
Group.LADref:aprime.log  0.53936    0.62600   0.862   0.3948
---
Signif. codes:  0 ‘***’ 0.001 ‘**’ 0.01 ‘*’ 0.05 ‘.’ 0.1 ‘ ’ 1

Residual standard error: 0.1919 on 35 degrees of freedom
  (13 observations deleted due to missingness)
Multiple R-squared:  0.1397,	Adjusted R-squared:  0.06594
F-statistic: 1.894 on 3 and 35 DF,  p-value: 0.1486

>
> mSurvey_ASDNT.Words = lm(QoLLifeSatSocSupp.log ~
+                           Group.LADref*FirstWords.months
+                 #+ age
+                       #+ vabs_soc_ss
+                         , data = dASDNT.older18)
> summary(mSurvey_ASDNT.Words)

Call:
lm(formula = QoLLifeSatSocSupp.log ~ Group.LADref * FirstWords.months,
    data = dASDNT.older18)

Residuals:
     Min       1Q   Median       3Q      Max
-0.42350 -0.09347  0.03292  0.12621  0.34165

Coefficients:
                                Estimate Std. Error t value Pr(>|t|)
(Intercept)                     4.709447   0.090946  51.783   <2e-16 ***
Group.LADref                    0.379591   0.181893   2.087   0.0468 *
FirstWords.months               0.001629   0.006118   0.266   0.7921
Group.LADref:FirstWords.months -0.016302   0.012235  -1.332   0.1943
---
Signif. codes:  0 ‘***’ 0.001 ‘**’ 0.01 ‘*’ 0.05 ‘.’ 0.1 ‘ ’ 1

Residual standard error: 0.1958 on 26 degrees of freedom
  (22 observations deleted due to missingness)
Multiple R-squared:  0.1944,	Adjusted R-squared:  0.1015
F-statistic: 2.092 on 3 and 26 DF,  p-value: 0.1257

>
> mSurvey_ASDNT.Phrases = lm(QoLLifeSatSocSupp.log ~
+                           Group.LADref*FirstPhrases.months
+                 #+ age
+                       #+ vabs_soc_ss
+                         , data = dASDNT.older18)
> summary(mSurvey_ASDNT.Phrases)

Call:
lm(formula = QoLLifeSatSocSupp.log ~ Group.LADref * FirstPhrases.months,
    data = dASDNT.older18)

Residuals:
     Min       1Q   Median       3Q      Max
-0.47892 -0.08733  0.04683  0.13948  0.28530

Coefficients:
                                   Estimate Std. Error t value Pr(>|t|)
(Intercept)                       4.7616116  0.1179574  40.367   <2e-16 ***
Group.LADref                      0.1420957  0.2359149   0.602    0.552
FirstPhrases.months              -0.0016201  0.0042198  -0.384    0.704
Group.LADref:FirstPhrases.months -0.0001356  0.0084396  -0.016    0.987
---
Signif. codes:  0 ‘***’ 0.001 ‘**’ 0.01 ‘*’ 0.05 ‘.’ 0.1 ‘ ’ 1

Residual standard error: 0.2033 on 28 degrees of freedom
  (20 observations deleted due to missingness)
Multiple R-squared:  0.1352,	Adjusted R-squared:  0.04253
F-statistic: 1.459 on 3 and 28 DF,  p-value: 0.247

>
>
>
>
>
> ### LAD NT ###
> mSurvey_LADNT.GJ = lm(QoLLifeSatSocSupp.log ~
+                           Group.ASDref*aprime.log
+                 #+ age
+                       #+ vabs_soc_ss
+                         , data = dLADNT.older18)
> summary(mSurvey_LADNT.GJ)

Call:
lm(formula = QoLLifeSatSocSupp.log ~ Group.ASDref * aprime.log,
    data = dLADNT.older18)

Residuals:
     Min       1Q   Median       3Q      Max
-0.47771 -0.06932  0.02781  0.10197  0.25469

Coefficients:
                        Estimate Std. Error t value Pr(>|t|)
(Intercept)              4.79569    0.04341 110.479   <2e-16 ***
Group.ASDref             0.03896    0.08682   0.449    0.656
aprime.log              -0.10864    0.31337  -0.347    0.730
Group.ASDref:aprime.log  0.45575    0.62674   0.727    0.471
---
Signif. codes:  0 ‘***’ 0.001 ‘**’ 0.01 ‘*’ 0.05 ‘.’ 0.1 ‘ ’ 1

Residual standard error: 0.1606 on 45 degrees of freedom
  (8 observations deleted due to missingness)
Multiple R-squared:  0.01681,	Adjusted R-squared:  -0.04874
F-statistic: 0.2565 on 3 and 45 DF,  p-value: 0.8563

>
> mSurvey_LADNT.Words = lm(QoLLifeSatSocSupp.log ~
+                           Group.ASDref*FirstWords.months
+                 #+ age
+                       #+ vabs_soc_ss
+                         , data = dLADNT.older18)
> summary(mSurvey_LADNT.Words)

Call:
lm(formula = QoLLifeSatSocSupp.log ~ Group.ASDref * FirstWords.months,
    data = dLADNT.older18)

Residuals:
     Min       1Q   Median       3Q      Max
-0.41850 -0.07397  0.02216  0.09933  0.22681

Coefficients:
                                Estimate Std. Error t value Pr(>|t|)
(Intercept)                     4.811898   0.067778  70.995   <2e-16 ***
Group.ASDref                    0.174688   0.135557   1.289    0.206
FirstWords.months              -0.001351   0.004228  -0.320    0.751
Group.ASDref:FirstWords.months -0.010343   0.008455  -1.223    0.229
---
Signif. codes:  0 ‘***’ 0.001 ‘**’ 0.01 ‘*’ 0.05 ‘.’ 0.1 ‘ ’ 1

Residual standard error: 0.1636 on 35 degrees of freedom
  (18 observations deleted due to missingness)
Multiple R-squared:  0.05131,	Adjusted R-squared:  -0.03001
F-statistic: 0.631 on 3 and 35 DF,  p-value: 0.5999

>
> mSurvey_LADNT.Phrases = lm(QoLLifeSatSocSupp.log ~
+                           Group.ASDref*FirstPhrases.months
+                 #+ age
+                       #+ vabs_soc_ss
+                         , data = dLADNT.older18)
> summary(mSurvey_LADNT.Phrases)

Call:
lm(formula = QoLLifeSatSocSupp.log ~ Group.ASDref * FirstPhrases.months,
    data = dLADNT.older18)

Residuals:
     Min       1Q   Median       3Q      Max
-0.45973 -0.07161  0.01356  0.10025  0.25277

Coefficients:
                                  Estimate Std. Error t value Pr(>|t|)
(Intercept)                       4.759758   0.059173  80.438   <2e-16 ***
Group.ASDref                      0.145804   0.118346   1.232    0.226
FirstPhrases.months               0.001213   0.002534   0.478    0.635
Group.ASDref:FirstPhrases.months -0.005801   0.005069  -1.144    0.260
---
Signif. codes:  0 ‘***’ 0.001 ‘**’ 0.01 ‘*’ 0.05 ‘.’ 0.1 ‘ ’ 1

Residual standard error: 0.1615 on 37 degrees of freedom
  (16 observations deleted due to missingness)
Multiple R-squared:  0.08705,	Adjusted R-squared:  0.01303
F-statistic: 1.176 on 3 and 37 DF,  p-value: 0.332

>
>
>
>
> ###Follow up
> #aprime.log
> mSurvey_ASD.GJ = lm(QoLLifeSatSocSupp.log ~ aprime.log
+                 #+ age
+                       #+ vabs_soc_ss
+                         , data = dASD.older18)
> summary(mSurvey_ASD.GJ)

Call:
lm(formula = QoLLifeSatSocSupp.log ~ aprime.log, data = dASD.older18)

Residuals:
     Min       1Q   Median       3Q      Max
-0.45727 -0.18864  0.06534  0.16889  0.26634

Coefficients:
            Estimate Std. Error t value Pr(>|t|)
(Intercept)  4.58862    0.09754  47.045 6.59e-16 ***
aprime.log  -0.42013    0.37432  -1.122    0.282
---
Signif. codes:  0 ‘***’ 0.001 ‘**’ 0.01 ‘*’ 0.05 ‘.’ 0.1 ‘ ’ 1

Residual standard error: 0.2254 on 13 degrees of freedom
  (10 observations deleted due to missingness)
Multiple R-squared:  0.08834,	Adjusted R-squared:  0.01821
F-statistic:  1.26 on 1 and 13 DF,  p-value: 0.282

>
> mSurvey_NT.GJ = lm(QoLLifeSatSocSupp.log ~ aprime.log
+                 #+ age
+                       #+ vabs_soc_ss
+                         , data = dNT.older18)
> summary(mSurvey_NT.GJ)

Call:
lm(formula = QoLLifeSatSocSupp.log ~ aprime.log, data = dNT.older18)

Residuals:
     Min       1Q   Median       3Q      Max
-0.47771 -0.06497  0.05159  0.11172  0.20883

Coefficients:
            Estimate Std. Error t value Pr(>|t|)
(Intercept)  4.81516    0.06556  73.445   <2e-16 ***
aprime.log   0.11923    0.47458   0.251    0.804
---
Signif. codes:  0 ‘***’ 0.001 ‘**’ 0.01 ‘*’ 0.05 ‘.’ 0.1 ‘ ’ 1

Residual standard error: 0.1691 on 22 degrees of freedom
  (3 observations deleted due to missingness)
Multiple R-squared:  0.002861,	Adjusted R-squared:  -0.04246
F-statistic: 0.06312 on 1 and 22 DF,  p-value: 0.804

>
> mSurvey_LAD.GJ = lm(QoLLifeSatSocSupp.log ~ aprime.log
+                 #+ age
+                       #+ vabs_soc_ss
+                         , data = dLAD.older18)
> summary(mSurvey_LAD.GJ)

Call:
lm(formula = QoLLifeSatSocSupp.log ~ aprime.log, data = dLAD.older18)

Residuals:
     Min       1Q   Median       3Q      Max
-0.31743 -0.09218  0.02374  0.09896  0.25469

Coefficients:
            Estimate Std. Error t value Pr(>|t|)
(Intercept)  4.77621    0.05727  83.399   <2e-16 ***
aprime.log  -0.33652    0.41225  -0.816    0.423
---
Signif. codes:  0 ‘***’ 0.001 ‘**’ 0.01 ‘*’ 0.05 ‘.’ 0.1 ‘ ’ 1

Residual standard error: 0.1521 on 23 degrees of freedom
  (5 observations deleted due to missingness)
Multiple R-squared:  0.02816,	Adjusted R-squared:  -0.0141
F-statistic: 0.6663 on 1 and 23 DF,  p-value: 0.4227

>
> #Words
> mSurvey_ASD.Words = lm(QoLLifeSatSocSupp.log ~ FirstWords.months
+                 #+ age
+                       #+ vabs_soc_ss
+                         , data = dASD.older18)
> summary(mSurvey_ASD.Words)

Call:
lm(formula = QoLLifeSatSocSupp.log ~ FirstWords.months, data = dASD.older18)

Residuals:
     Min       1Q   Median       3Q      Max
-0.42350 -0.09408  0.02277  0.15625  0.34165

Coefficients:
                  Estimate Std. Error t value Pr(>|t|)
(Intercept)       4.519651   0.156077  28.958 2.19e-09 ***
FirstWords.months 0.009780   0.009842   0.994    0.349
---
Signif. codes:  0 ‘***’ 0.001 ‘**’ 0.01 ‘*’ 0.05 ‘.’ 0.1 ‘ ’ 1

Residual standard error: 0.2376 on 8 degrees of freedom
  (15 observations deleted due to missingness)
Multiple R-squared:  0.1099,	Adjusted R-squared:  -0.001392
F-statistic: 0.9875 on 1 and 8 DF,  p-value: 0.3495

>
> mSurvey_NT.Words = lm(QoLLifeSatSocSupp.log ~ FirstWords.months
+                 #+ age
+                       #+ vabs_soc_ss
+                         , data = dNT.older18)
> summary(mSurvey_NT.Words)

Call:
lm(formula = QoLLifeSatSocSupp.log ~ FirstWords.months, data = dNT.older18)

Residuals:
     Min       1Q   Median       3Q      Max
-0.41850 -0.08920  0.03789  0.09550  0.22211

Coefficients:
                   Estimate Std. Error t value Pr(>|t|)
(Intercept)        4.899242   0.114311  42.859   <2e-16 ***
FirstWords.months -0.006522   0.008142  -0.801    0.434
---
Signif. codes:  0 ‘***’ 0.001 ‘**’ 0.01 ‘*’ 0.05 ‘.’ 0.1 ‘ ’ 1

Residual standard error: 0.1741 on 18 degrees of freedom
  (7 observations deleted due to missingness)
Multiple R-squared:  0.03442,	Adjusted R-squared:  -0.01922
F-statistic: 0.6417 on 1 and 18 DF,  p-value: 0.4336

>
> mSurvey_LAD.Words = lm(QoLLifeSatSocSupp.log ~ FirstWords.months
+                 #+ age
+                       #+ vabs_soc_ss
+                         , data = dNT.older18)
> summary(mSurvey_LAD.Words)

Call:
lm(formula = QoLLifeSatSocSupp.log ~ FirstWords.months, data = dNT.older18)

Residuals:
     Min       1Q   Median       3Q      Max
-0.41850 -0.08920  0.03789  0.09550  0.22211

Coefficients:
                   Estimate Std. Error t value Pr(>|t|)
(Intercept)        4.899242   0.114311  42.859   <2e-16 ***
FirstWords.months -0.006522   0.008142  -0.801    0.434
---
Signif. codes:  0 ‘***’ 0.001 ‘**’ 0.01 ‘*’ 0.05 ‘.’ 0.1 ‘ ’ 1

Residual standard error: 0.1741 on 18 degrees of freedom
  (7 observations deleted due to missingness)
Multiple R-squared:  0.03442,	Adjusted R-squared:  -0.01922
F-statistic: 0.6417 on 1 and 18 DF,  p-value: 0.4336

>
> #Phrases
> mSurvey_ASD.Phrases = lm(QoLLifeSatSocSupp.log ~ FirstPhrases.months
+                 #+ age
+                       #+ vabs_soc_ss
+                         , data = dASD.older18)
> summary(mSurvey_ASD.Phrases)

Call:
lm(formula = QoLLifeSatSocSupp.log ~ FirstPhrases.months, data = dASD.older18)

Residuals:
     Min       1Q   Median       3Q      Max
-0.47892 -0.13347  0.07052  0.18037  0.28530

Coefficients:
                     Estimate Std. Error t value Pr(>|t|)
(Intercept)          4.690564   0.261943  17.907 9.69e-08 ***
FirstPhrases.months -0.001552   0.007899  -0.197    0.849
---
Signif. codes:  0 ‘***’ 0.001 ‘**’ 0.01 ‘*’ 0.05 ‘.’ 0.1 ‘ ’ 1

Residual standard error: 0.2589 on 8 degrees of freedom
  (15 observations deleted due to missingness)
Multiple R-squared:  0.004804,	Adjusted R-squared:  -0.1196
F-statistic: 0.03862 on 1 and 8 DF,  p-value: 0.8491

>
> mSurvey_NT.Phrases = lm(QoLLifeSatSocSupp.log ~ FirstPhrases.months
+                 #+ age
+                       #+ vabs_soc_ss
+                         , data = dNT.older18)
> summary(mSurvey_NT.Phrases)

Call:
lm(formula = QoLLifeSatSocSupp.log ~ FirstPhrases.months, data = dNT.older18)

Residuals:
     Min       1Q   Median       3Q      Max
-0.45973 -0.06241  0.04683  0.11544  0.21500

Coefficients:
                     Estimate Std. Error t value Pr(>|t|)
(Intercept)          4.832659   0.100211   48.23   <2e-16 ***
FirstPhrases.months -0.001688   0.004961   -0.34    0.737
---
Signif. codes:  0 ‘***’ 0.001 ‘**’ 0.01 ‘*’ 0.05 ‘.’ 0.1 ‘ ’ 1

Residual standard error: 0.1761 on 20 degrees of freedom
  (5 observations deleted due to missingness)
Multiple R-squared:  0.005754,	Adjusted R-squared:  -0.04396
F-statistic: 0.1158 on 1 and 20 DF,  p-value: 0.7372

>
> mSurvey_LAD.Phrases = lm(QoLLifeSatSocSupp.log ~ FirstPhrases.months
+                 #+ age
+                       #+ vabs_soc_ss
+                         , data = dLAD.older18)
> summary(mSurvey_LAD.Phrases)

Call:
lm(formula = QoLLifeSatSocSupp.log ~ FirstPhrases.months, data = dLAD.older18)

Residuals:
     Min       1Q   Median       3Q      Max
-0.25003 -0.07383  0.00926  0.06057  0.25277

Coefficients:
                    Estimate Std. Error t value Pr(>|t|)
(Intercept)         4.686856   0.065749  71.284   <2e-16 ***
FirstPhrases.months 0.004113   0.001971   2.086   0.0523 .
---
Signif. codes:  0 ‘***’ 0.001 ‘**’ 0.01 ‘*’ 0.05 ‘.’ 0.1 ‘ ’ 1

Residual standard error: 0.1423 on 17 degrees of freedom
  (11 observations deleted due to missingness)
Multiple R-squared:  0.2038,	Adjusted R-squared:  0.157
F-statistic: 4.353 on 1 and 17 DF,  p-value: 0.05233


> ###############
> ##### QoL #####
> ###############
>
> #qol.pos.log
>
> ### ASD LAD ###
> mQoL_ASDLAD.GJ = lm(qol.pos.log ~
+                           Group.NTref*aprime.log
+                  #+ age
+                       #+ vabs_soc_ss
+                         , data = dASDLAD.older18)
> summary(mQoL_ASDLAD.GJ)

Call:
lm(formula = qol.pos.log ~ Group.NTref * aprime.log, data = dASDLAD.older18)

Residuals:
     Min       1Q   Median       3Q      Max
-0.59570 -0.11433  0.07038  0.14180  0.31956

Coefficients:
                       Estimate Std. Error t value Pr(>|t|)
(Intercept)             3.70218    0.06459  57.314   <2e-16 ***
Group.NTref             0.23006    0.12919   1.781   0.0834 .
aprime.log             -0.45535    0.35800  -1.272   0.2115
Group.NTref:aprime.log  0.12560    0.71599   0.175   0.8617
---
Signif. codes:  0 ‘***’ 0.001 ‘**’ 0.01 ‘*’ 0.05 ‘.’ 0.1 ‘ ’ 1

Residual standard error: 0.2252 on 36 degrees of freedom
  (15 observations deleted due to missingness)
Multiple R-squared:  0.1733,	Adjusted R-squared:  0.1044
F-statistic: 2.515 on 3 and 36 DF,  p-value: 0.07374

>
> mQoL_ASDLAD.Words = lm(qol.pos.log ~
+                           Group.NTref*FirstWords.months
+                  #+ age
+                       #+ vabs_soc_ss
+                         , data = dASDLAD.older18)
> summary(mQoL_ASDLAD.Words)

Call:
lm(formula = qol.pos.log ~ Group.NTref * FirstWords.months, data = dASDLAD.older18)

Residuals:
    Min      1Q  Median      3Q     Max
-0.5036 -0.1235  0.0284  0.1688  0.4366

Coefficients:
                               Estimate Std. Error t value Pr(>|t|)
(Intercept)                    3.591283   0.094779  37.891   <2e-16 ***
Group.NTref                    0.353705   0.189558   1.866   0.0738 .
FirstWords.months              0.010195   0.005363   1.901   0.0689 .
Group.NTref:FirstWords.months -0.013021   0.010726  -1.214   0.2361
---
Signif. codes:  0 ‘***’ 0.001 ‘**’ 0.01 ‘*’ 0.05 ‘.’ 0.1 ‘ ’ 1

Residual standard error: 0.2287 on 25 degrees of freedom
  (26 observations deleted due to missingness)
Multiple R-squared:  0.2538,	Adjusted R-squared:  0.1643
F-statistic: 2.835 on 3 and 25 DF,  p-value: 0.05859

>
> mQoL_ASDLAD.Phrases = lm(qol.pos.log ~
+                           Group.NTref*FirstPhrases.months
+                  #+ age
+                       #+ vabs_soc_ss
+                         , data = dASDLAD.older18)
> summary(mQoL_ASDLAD.Phrases)

Call:
lm(formula = qol.pos.log ~ Group.NTref * FirstPhrases.months,
    data = dASDLAD.older18)

Residuals:
    Min      1Q  Median      3Q     Max
-0.6270 -0.1281  0.0174  0.1904  0.3144

Coefficients:
                                 Estimate Std. Error t value Pr(>|t|)
(Intercept)                      3.689470   0.128426  28.728   <2e-16 ***
Group.NTref                     -0.003246   0.256853  -0.013    0.990
FirstPhrases.months              0.002281   0.003869   0.590    0.561
Group.NTref:FirstPhrases.months  0.006197   0.007738   0.801    0.431
---
Signif. codes:  0 ‘***’ 0.001 ‘**’ 0.01 ‘*’ 0.05 ‘.’ 0.1 ‘ ’ 1

Residual standard error: 0.231 on 25 degrees of freedom
  (26 observations deleted due to missingness)
Multiple R-squared:  0.2124,	Adjusted R-squared:  0.1179
F-statistic: 2.247 on 3 and 25 DF,  p-value: 0.1076

>
>
>
>
> ### ASD NT ###
> mQoL_ASDNT.GJ = lm(qol.pos.log ~
+                           Group.LADref*aprime.log
+                 #+ age
+                       #+ vabs_soc_ss
+                         , data = dASDNT.older18)
> summary(mQoL_ASDNT.GJ)

Call:
lm(formula = qol.pos.log ~ Group.LADref * aprime.log, data = dASDNT.older18)

Residuals:
     Min       1Q   Median       3Q      Max
-0.59570 -0.08834  0.01985  0.14199  0.30467

Coefficients:
                        Estimate Std. Error t value Pr(>|t|)
(Intercept)              3.73694    0.06174  60.524   <2e-16 ***
Group.LADref             0.29960    0.12349   2.426   0.0206 *
aprime.log              -0.10578    0.34658  -0.305   0.7620
Group.LADref:aprime.log  0.82476    0.69316   1.190   0.2421
---
Signif. codes:  0 ‘***’ 0.001 ‘**’ 0.01 ‘*’ 0.05 ‘.’ 0.1 ‘ ’ 1

Residual standard error: 0.2125 on 35 degrees of freedom
  (13 observations deleted due to missingness)
Multiple R-squared:  0.1735,	Adjusted R-squared:  0.1026
F-statistic: 2.448 on 3 and 35 DF,  p-value: 0.07995

>
> mQoL_ASDNT.Words = lm(qol.pos.log ~
+                           Group.LADref*FirstWords.months
+                 #+ age
+                       #+ vabs_soc_ss
+                         , data = dASDNT.older18)
> summary(mQoL_ASDNT.Words)

Call:
lm(formula = qol.pos.log ~ Group.LADref * FirstWords.months,
    data = dASDNT.older18)

Residuals:
     Min       1Q   Median       3Q      Max
-0.50355 -0.10840  0.01539  0.10758  0.43658

Coefficients:
                                Estimate Std. Error t value Pr(>|t|)
(Intercept)                     3.650883   0.096871  37.688   <2e-16 ***
Group.LADref                    0.472906   0.193743   2.441   0.0218 *
FirstWords.months               0.007324   0.006516   1.124   0.2713
Group.LADref:FirstWords.months -0.018762   0.013032  -1.440   0.1619
---
Signif. codes:  0 ‘***’ 0.001 ‘**’ 0.01 ‘*’ 0.05 ‘.’ 0.1 ‘ ’ 1

Residual standard error: 0.2086 on 26 degrees of freedom
  (22 observations deleted due to missingness)
Multiple R-squared:  0.2929,	Adjusted R-squared:  0.2113
F-statistic:  3.59 on 3 and 26 DF,  p-value: 0.02704

>
> mQoL_ASDNT.Phrases = lm(qol.pos.log ~
+                           Group.LADref*FirstPhrases.months
+                 #+ age
+                       #+ vabs_soc_ss
+                         , data = dASDNT.older18)
> summary(mQoL_ASDNT.Phrases)

Call:
lm(formula = qol.pos.log ~ Group.LADref * FirstPhrases.months,
    data = dASDNT.older18)

Residuals:
     Min       1Q   Median       3Q      Max
-0.62696 -0.08768  0.04506  0.13774  0.28957

Coefficients:
                                  Estimate Std. Error t value Pr(>|t|)
(Intercept)                      3.763e+00  1.279e-01  29.415   <2e-16 ***
Group.LADref                     1.442e-01  2.559e-01   0.564    0.578
FirstPhrases.months              7.092e-06  4.577e-03   0.002    0.999
Group.LADref:FirstPhrases.months 1.648e-03  9.153e-03   0.180    0.858
---
Signif. codes:  0 ‘***’ 0.001 ‘**’ 0.01 ‘*’ 0.05 ‘.’ 0.1 ‘ ’ 1

Residual standard error: 0.2204 on 28 degrees of freedom
  (20 observations deleted due to missingness)
Multiple R-squared:  0.149,	Adjusted R-squared:  0.05779
F-statistic: 1.634 on 3 and 28 DF,  p-value: 0.204

>
>
>
>
>
> ### LAD NT ###
> mQoL_LADNT.GJ = lm(qol.pos.log ~
+                           Group.ASDref*aprime.log
+                 #+ age
+                       #+ vabs_soc_ss
+                         , data = dLADNT.older18)
> summary(mQoL_LADNT.GJ)

Call:
lm(formula = qol.pos.log ~ Group.ASDref * aprime.log, data = dLADNT.older18)

Residuals:
     Min       1Q   Median       3Q      Max
-0.44397 -0.07629  0.01698  0.12937  0.31956

Coefficients:
                        Estimate Std. Error t value Pr(>|t|)
(Intercept)              3.85197    0.04894  78.707   <2e-16 ***
Group.ASDref             0.06954    0.09788   0.710    0.481
aprime.log              -0.04297    0.35331  -0.122    0.904
Group.ASDref:aprime.log  0.69915    0.70662   0.989    0.328
---
Signif. codes:  0 ‘***’ 0.001 ‘**’ 0.01 ‘*’ 0.05 ‘.’ 0.1 ‘ ’ 1

Residual standard error: 0.1811 on 45 degrees of freedom
  (8 observations deleted due to missingness)
Multiple R-squared:  0.02309,	Adjusted R-squared:  -0.04204
F-statistic: 0.3545 on 3 and 45 DF,  p-value: 0.7861

>
> mQoL_LADNT.Words =  lm(qol.pos.log ~
+                           Group.ASDref*FirstWords.months
+                 #+ age
+                       #+ vabs_soc_ss
+                         , data = dLADNT.older18)
> summary(mQoL_LADNT.Words)

Call:
lm(formula = qol.pos.log ~ Group.ASDref * FirstWords.months,
    data = dLADNT.older18)

Residuals:
     Min       1Q   Median       3Q      Max
-0.40604 -0.10458  0.00238  0.16163  0.28936

Coefficients:
                                 Estimate Std. Error t value Pr(>|t|)
(Intercept)                     3.8277354  0.0786019  48.698   <2e-16 ***
Group.ASDref                    0.1192003  0.1572038   0.758    0.453
FirstWords.months               0.0008139  0.0049028   0.166    0.869
Group.ASDref:FirstWords.months -0.0057416  0.0098055  -0.586    0.562
---
Signif. codes:  0 ‘***’ 0.001 ‘**’ 0.01 ‘*’ 0.05 ‘.’ 0.1 ‘ ’ 1

Residual standard error: 0.1897 on 35 degrees of freedom
  (18 observations deleted due to missingness)
Multiple R-squared:  0.02529,	Adjusted R-squared:  -0.05826
F-statistic: 0.3026 on 3 and 35 DF,  p-value: 0.8233

>
> mQoL_LADNT.Phrases = lm(qol.pos.log ~
+                           Group.ASDref*FirstPhrases.months
+                 #+ age
+                       #+ vabs_soc_ss
+                         , data = dLADNT.older18)
> summary(mQoL_LADNT.Phrases)

Call:
lm(formula = qol.pos.log ~ Group.ASDref * FirstPhrases.months,
    data = dLADNT.older18)

Residuals:
     Min       1Q   Median       3Q      Max
-0.42209 -0.12031  0.00517  0.13428  0.31438

Coefficients:
                                  Estimate Std. Error t value Pr(>|t|)
(Intercept)                       3.761572   0.066486  56.577   <2e-16 ***
Group.ASDref                      0.147450   0.132972   1.109    0.275
FirstPhrases.months               0.003105   0.002848   1.091    0.283
Group.ASDref:FirstPhrases.months -0.004548   0.005695  -0.799    0.430
---
Signif. codes:  0 ‘***’ 0.001 ‘**’ 0.01 ‘*’ 0.05 ‘.’ 0.1 ‘ ’ 1

Residual standard error: 0.1814 on 37 degrees of freedom
  (16 observations deleted due to missingness)
Multiple R-squared:  0.1111,	Adjusted R-squared:  0.03903
F-statistic: 1.541 on 3 and 37 DF,  p-value: 0.22

>
>
>
>
> ###Follow up
> #aprime.log
> mQoL_ASD.GJ = lm(qol.pos.log ~ aprime.log
+                 #+ age
+                       #+ vabs_soc_ss
+                         , data = dASD.older18)
> summary(mQoL_ASD.GJ)

Call:
lm(formula = qol.pos.log ~ aprime.log, data = dASD.older18)

Residuals:
     Min       1Q   Median       3Q      Max
-0.59570 -0.15688  0.08087  0.15596  0.30467

Coefficients:
            Estimate Std. Error t value Pr(>|t|)
(Intercept)   3.5871     0.1179  30.417 1.82e-13 ***
aprime.log   -0.5182     0.4526  -1.145    0.273
---
Signif. codes:  0 ‘***’ 0.001 ‘**’ 0.01 ‘*’ 0.05 ‘.’ 0.1 ‘ ’ 1

Residual standard error: 0.2726 on 13 degrees of freedom
  (10 observations deleted due to missingness)
Multiple R-squared:  0.09159,	Adjusted R-squared:  0.02171
F-statistic: 1.311 on 1 and 13 DF,  p-value: 0.2729

>
> mQoL_NT.GJ = data = lm(qol.pos.log ~ aprime.log
+                 #+ age
+                       #+ vabs_soc_ss
+                         , dNT.older18)
> summary(mQoL_NT.GJ)

Call:
lm(formula = qol.pos.log ~ aprime.log, data = dNT.older18)

Residuals:
     Min       1Q   Median       3Q      Max
-0.43548 -0.05703  0.00159  0.11774  0.24608

Coefficients:
            Estimate Std. Error t value Pr(>|t|)
(Intercept)  3.88674    0.06485  59.938   <2e-16 ***
aprime.log   0.30660    0.46940   0.653     0.52
---
Signif. codes:  0 ‘***’ 0.001 ‘**’ 0.01 ‘*’ 0.05 ‘.’ 0.1 ‘ ’ 1

Residual standard error: 0.1672 on 22 degrees of freedom
  (3 observations deleted due to missingness)
Multiple R-squared:  0.01902,	Adjusted R-squared:  -0.02557
F-statistic: 0.4266 on 1 and 22 DF,  p-value: 0.5204

>
> mQoL_LAD.GJ = lm(qol.pos.log ~ aprime.log
+                 #+ age
+                       #+ vabs_soc_ss
+                         , data = dLAD.older18)
> summary(mQoL_LAD.GJ)

Call:
lm(formula = qol.pos.log ~ aprime.log, data = dLAD.older18)

Residuals:
     Min       1Q   Median       3Q      Max
-0.44397 -0.09765  0.05399  0.12937  0.31956

Coefficients:
            Estimate Std. Error t value Pr(>|t|)
(Intercept)  3.81721    0.07283  52.410   <2e-16 ***
aprime.log  -0.39255    0.52429  -0.749    0.462
---
Signif. codes:  0 ‘***’ 0.001 ‘**’ 0.01 ‘*’ 0.05 ‘.’ 0.1 ‘ ’ 1

Residual standard error: 0.1934 on 23 degrees of freedom
  (5 observations deleted due to missingness)
Multiple R-squared:  0.02379,	Adjusted R-squared:  -0.01865
F-statistic: 0.5606 on 1 and 23 DF,  p-value: 0.4616

>
> #Words
> mQoL_ASD.Words = lm(qol.pos.log ~ FirstWords.months
+                 #+ age
+                       #+ vabs_soc_ss
+                         , data = dASD.older18)
> summary(mQoL_ASD.Words)

Call:
lm(formula = qol.pos.log ~ FirstWords.months, data = dASD.older18)

Residuals:
     Min       1Q   Median       3Q      Max
-0.50355 -0.13286  0.05244  0.10343  0.43658

Coefficients:
                  Estimate Std. Error t value Pr(>|t|)
(Intercept)        3.41443    0.17833  19.146 5.74e-08 ***
FirstWords.months  0.01671    0.01125   1.486    0.176
---
Signif. codes:  0 ‘***’ 0.001 ‘**’ 0.01 ‘*’ 0.05 ‘.’ 0.1 ‘ ’ 1

Residual standard error: 0.2715 on 8 degrees of freedom
  (15 observations deleted due to missingness)
Multiple R-squared:  0.2162,	Adjusted R-squared:  0.1182
F-statistic: 2.207 on 1 and 8 DF,  p-value: 0.1757

>
> mQoL_NT.Words = lm(qol.pos.log ~ FirstWords.months
+                 #+ age
+                       #+ vabs_soc_ss
+                         , data = dNT.older18)
> summary(mQoL_NT.Words)

Call:
lm(formula = qol.pos.log ~ FirstWords.months, data = dNT.older18)

Residuals:
     Min       1Q   Median       3Q      Max
-0.40604 -0.07692 -0.00506  0.11935  0.24403

Coefficients:
                   Estimate Std. Error t value Pr(>|t|)
(Intercept)        3.887336   0.113911  34.126   <2e-16 ***
FirstWords.months -0.002057   0.008114  -0.254    0.803
---
Signif. codes:  0 ‘***’ 0.001 ‘**’ 0.01 ‘*’ 0.05 ‘.’ 0.1 ‘ ’ 1

Residual standard error: 0.1735 on 18 degrees of freedom
  (7 observations deleted due to missingness)
Multiple R-squared:  0.003558,	Adjusted R-squared:  -0.0518
F-statistic: 0.06427 on 1 and 18 DF,  p-value: 0.8027

>
> mQoL_LAD.Words = lm(qol.pos.log ~ FirstWords.months
+                 #+ age
+                       #+ vabs_soc_ss
+                         , data = dNT.older18)
> summary(mQoL_LAD.Words)

Call:
lm(formula = qol.pos.log ~ FirstWords.months, data = dNT.older18)

Residuals:
     Min       1Q   Median       3Q      Max
-0.40604 -0.07692 -0.00506  0.11935  0.24403

Coefficients:
                   Estimate Std. Error t value Pr(>|t|)
(Intercept)        3.887336   0.113911  34.126   <2e-16 ***
FirstWords.months -0.002057   0.008114  -0.254    0.803
---
Signif. codes:  0 ‘***’ 0.001 ‘**’ 0.01 ‘*’ 0.05 ‘.’ 0.1 ‘ ’ 1

Residual standard error: 0.1735 on 18 degrees of freedom
  (7 observations deleted due to missingness)
Multiple R-squared:  0.003558,	Adjusted R-squared:  -0.0518
F-statistic: 0.06427 on 1 and 18 DF,  p-value: 0.8027

>
> #Phrases
> mQoL_ASD.Phrases = lm(qol.pos.log ~ FirstPhrases.months
+                 #+ age
+                       #+ vabs_soc_ss
+                         , data = dASD.older18)
> summary(mQoL_ASD.Phrases)

Call:
lm(formula = qol.pos.log ~ FirstPhrases.months, data = dASD.older18)

Residuals:
     Min       1Q   Median       3Q      Max
-0.62696 -0.12688  0.08266  0.20028  0.28957

Coefficients:
                      Estimate Std. Error t value Pr(>|t|)
(Intercept)          3.6910927  0.3072933  12.012 2.13e-06 ***
FirstPhrases.months -0.0008171  0.0092665  -0.088    0.932
---
Signif. codes:  0 ‘***’ 0.001 ‘**’ 0.01 ‘*’ 0.05 ‘.’ 0.1 ‘ ’ 1

Residual standard error: 0.3038 on 8 degrees of freedom
  (15 observations deleted due to missingness)
Multiple R-squared:  0.0009711,	Adjusted R-squared:  -0.1239
F-statistic: 0.007776 on 1 and 8 DF,  p-value: 0.9319

>
> mQoL_NT.Phrases = lm(qol.pos.log ~ FirstPhrases.months
+                 #+ age
+                       #+ vabs_soc_ss
+                         , data = dNT.older18)
> summary(mQoL_NT.Phrases)

Call:
lm(formula = qol.pos.log ~ FirstPhrases.months, data = dNT.older18)

Residuals:
     Min       1Q   Median       3Q      Max
-0.42209 -0.07709  0.01623  0.11589  0.24408

Coefficients:
                     Estimate Std. Error t value Pr(>|t|)
(Intercept)         3.8352965  0.1003862  38.205   <2e-16 ***
FirstPhrases.months 0.0008313  0.0049699   0.167    0.869
---
Signif. codes:  0 ‘***’ 0.001 ‘**’ 0.01 ‘*’ 0.05 ‘.’ 0.1 ‘ ’ 1

Residual standard error: 0.1764 on 20 degrees of freedom
  (5 observations deleted due to missingness)
Multiple R-squared:  0.001397,	Adjusted R-squared:  -0.04853
F-statistic: 0.02798 on 1 and 20 DF,  p-value: 0.8688

>
> mQoL_LAD.Phrases = lm(qol.pos.log ~ FirstPhrases.months
+                 #+ age
+                       #+ vabs_soc_ss
+                         , data = dLAD.older18)
> summary(mQoL_LAD.Phrases)

Call:
lm(formula = qol.pos.log ~ FirstPhrases.months, data = dLAD.older18)

Residuals:
      Min        1Q    Median        3Q       Max
-0.307656 -0.126828  0.005173  0.162330  0.314375

Coefficients:
                    Estimate Std. Error t value Pr(>|t|)
(Intercept)         3.687847   0.086455  42.656   <2e-16 ***
FirstPhrases.months 0.005380   0.002592   2.075   0.0535 .
---
Signif. codes:  0 ‘***’ 0.001 ‘**’ 0.01 ‘*’ 0.05 ‘.’ 0.1 ‘ ’ 1

Residual standard error: 0.1871 on 17 degrees of freedom
  (11 observations deleted due to missingness)
Multiple R-squared:  0.2021,	Adjusted R-squared:  0.1552
F-statistic: 4.306 on 1 and 17 DF,  p-value: 0.05346


> #############################
> ##### Life satisfaction #####
> #############################
> #lifesat.log
>
> ### ASD LAD ###
> mLifeSat_ASDLAD.GJ = lm(lifesat.log ~
+                           Group.NTref*aprime.log
+                  #+ age
+                       #+ vabs_soc_ss
+                         , data = dASDLAD.older18)
> summary(mLifeSat_ASDLAD.GJ)

Call:
lm(formula = lifesat.log ~ Group.NTref * aprime.log, data = dASDLAD.older18)

Residuals:
     Min       1Q   Median       3Q      Max
-1.08346 -0.10778  0.03356  0.18627  0.55024

Coefficients:
                       Estimate Std. Error t value Pr(>|t|)
(Intercept)             2.93496    0.09058  32.400   <2e-16 ***
Group.NTref             0.27719    0.18117   1.530   0.1348
aprime.log             -0.86687    0.50203  -1.727   0.0928 .
Group.NTref:aprime.log -0.19431    1.00407  -0.194   0.8476
---
Signif. codes:  0 ‘***’ 0.001 ‘**’ 0.01 ‘*’ 0.05 ‘.’ 0.1 ‘ ’ 1

Residual standard error: 0.3158 on 36 degrees of freedom
  (15 observations deleted due to missingness)
Multiple R-squared:  0.1889,	Adjusted R-squared:  0.1213
F-statistic: 2.794 on 3 and 36 DF,  p-value: 0.05415

>
> mLifeSat_ASDLAD.Words = lm(lifesat.log ~
+                           Group.NTref*FirstWords.months
+                  #+ age
+                       #+ vabs_soc_ss
+                         , data = dASDLAD.older18)
> summary(mLifeSat_ASDLAD.Words)

Call:
lm(formula = lifesat.log ~ Group.NTref * FirstWords.months, data = dASDLAD.older18)

Residuals:
    Min      1Q  Median      3Q     Max
-1.0964 -0.1039  0.0440  0.1930  0.4552

Coefficients:
                               Estimate Std. Error t value Pr(>|t|)
(Intercept)                    2.930974   0.139753  20.973   <2e-16 ***
Group.NTref                    0.281162   0.279506   1.006    0.324
FirstWords.months              0.008847   0.007908   1.119    0.274
Group.NTref:FirstWords.months -0.006760   0.015816  -0.427    0.673
---
Signif. codes:  0 ‘***’ 0.001 ‘**’ 0.01 ‘*’ 0.05 ‘.’ 0.1 ‘ ’ 1

Residual standard error: 0.3372 on 25 degrees of freedom
  (26 observations deleted due to missingness)
Multiple R-squared:  0.1432,	Adjusted R-squared:  0.04036
F-statistic: 1.393 on 3 and 25 DF,  p-value: 0.2682

>
> mLifeSat_ASDLAD.Phrases = lm(lifesat.log ~
+                           Group.NTref*FirstPhrases.months
+                  #+ age
+                       #+ vabs_soc_ss
+                         , data = dASDLAD.older18)
> summary(mLifeSat_ASDLAD.Phrases)

Call:
lm(formula = lifesat.log ~ Group.NTref * FirstPhrases.months,
    data = dASDLAD.older18)

Residuals:
     Min       1Q   Median       3Q      Max
-1.13890 -0.14238  0.02909  0.22123  0.53508

Coefficients:
                                  Estimate Std. Error t value Pr(>|t|)
(Intercept)                      3.0232028  0.1935939  15.616  2.1e-14 ***
Group.NTref                     -0.0386692  0.3871877  -0.100    0.921
FirstPhrases.months              0.0006315  0.0058322   0.108    0.915
Group.NTref:FirstPhrases.months  0.0105864  0.0116644   0.908    0.373
---
Signif. codes:  0 ‘***’ 0.001 ‘**’ 0.01 ‘*’ 0.05 ‘.’ 0.1 ‘ ’ 1

Residual standard error: 0.3481 on 25 degrees of freedom
  (26 observations deleted due to missingness)
Multiple R-squared:  0.1917,	Adjusted R-squared:  0.09473
F-statistic: 1.977 on 3 and 25 DF,  p-value: 0.1432

>
>
>
>
>
> ### ASD NT ###
> mLifeSat_ASDNT.GJ = lm(lifesat.log ~
+                           Group.LADref*aprime.log
+                 #+ age
+                       #+ vabs_soc_ss
+                         , data = dASDNT.older18)
> summary(mLifeSat_ASDNT.GJ)

Call:
lm(formula = lifesat.log ~ Group.LADref * aprime.log, data = dASDNT.older18)

Residuals:
     Min       1Q   Median       3Q      Max
-1.08346 -0.16090  0.03815  0.20391  0.55024

Coefficients:
                        Estimate Std. Error t value Pr(>|t|)
(Intercept)              2.98933    0.09455  31.616   <2e-16 ***
Group.LADref             0.38592    0.18910   2.041   0.0489 *
aprime.log              -0.37994    0.53073  -0.716   0.4788
Group.LADref:aprime.log  0.77954    1.06146   0.734   0.4676
---
Signif. codes:  0 ‘***’ 0.001 ‘**’ 0.01 ‘*’ 0.05 ‘.’ 0.1 ‘ ’ 1

Residual standard error: 0.3255 on 35 degrees of freedom
  (13 observations deleted due to missingness)
Multiple R-squared:  0.1545,	Adjusted R-squared:  0.08201
F-statistic: 2.132 on 3 and 35 DF,  p-value: 0.1138

>
> mLifeSat_ASDNT.Words = lm(lifesat.log ~
+                           Group.LADref*FirstWords.months
+                 #+ age
+                       #+ vabs_soc_ss
+                         , data = dASDNT.older18)
> summary(mLifeSat_ASDNT.Words)

Call:
lm(formula = lifesat.log ~ Group.LADref * FirstWords.months,
    data = dASDNT.older18)

Residuals:
     Min       1Q   Median       3Q      Max
-1.09645 -0.08031  0.04861  0.19570  0.45525

Coefficients:
                                 Estimate Std. Error t value Pr(>|t|)
(Intercept)                     3.0710857  0.1555341  19.745   <2e-16 ***
Group.LADref                    0.5613849  0.3110682   1.805   0.0827 .
FirstWords.months               0.0009138  0.0104621   0.087   0.9311
Group.LADref:FirstWords.months -0.0226275  0.0209241  -1.081   0.2894
---
Signif. codes:  0 ‘***’ 0.001 ‘**’ 0.01 ‘*’ 0.05 ‘.’ 0.1 ‘ ’ 1

Residual standard error: 0.3349 on 26 degrees of freedom
  (22 observations deleted due to missingness)
Multiple R-squared:  0.1628,	Adjusted R-squared:  0.0662
F-statistic: 1.685 on 3 and 26 DF,  p-value: 0.1946

>
> mLifeSat_ASDNT.Phrases = lm(lifesat.log ~
+                           Group.NTref*FirstPhrases.months
+                  #+ age
+                       #+ vabs_soc_ss
+                         , data = dASDNT.older18)
> summary(mLifeSat_ASDNT.Phrases)

Call:
lm(formula = lifesat.log ~ Group.NTref * FirstPhrases.months,
    data = dASDNT.older18)

Residuals:
    Min      1Q  Median      3Q     Max
-1.1389 -0.1561  0.0767  0.2212  0.5351

Coefficients:
                                 Estimate Std. Error t value Pr(>|t|)
(Intercept)                      3.353223   0.197113  17.012  2.7e-16 ***
Group.NTref                      0.621371   0.804218   0.773    0.446
FirstPhrases.months             -0.008850   0.009759  -0.907    0.372
Group.NTref:FirstPhrases.months -0.008376   0.028770  -0.291    0.773
---
Signif. codes:  0 ‘***’ 0.001 ‘**’ 0.01 ‘*’ 0.05 ‘.’ 0.1 ‘ ’ 1

Residual standard error: 0.3464 on 28 degrees of freedom
  (20 observations deleted due to missingness)
Multiple R-squared:  0.1739,	Adjusted R-squared:  0.08539
F-statistic: 1.965 on 3 and 28 DF,  p-value: 0.1422

>
>
>
>
> ### LAD NT  ###
> mLifeSat_LADNT.GJ = lm(lifesat.log ~
+                           Group.ASDref*aprime.log
+                 #+ age
+                       #+ vabs_soc_ss
+                         , data = dLADNT.older18)
> summary(mLifeSat_LADNT.GJ)

Call:
lm(formula = lifesat.log ~ Group.ASDref * aprime.log, data = dLADNT.older18)

Residuals:
     Min       1Q   Median       3Q      Max
-0.61630 -0.17894  0.03709  0.18551  0.39850

Coefficients:
                        Estimate Std. Error t value Pr(>|t|)
(Intercept)               3.1279     0.0646  48.424   <2e-16 ***
Group.ASDref              0.1087     0.1292   0.842    0.404
aprime.log               -0.4771     0.4663  -1.023    0.312
Group.ASDref:aprime.log   0.9738     0.9326   1.044    0.302
---
Signif. codes:  0 ‘***’ 0.001 ‘**’ 0.01 ‘*’ 0.05 ‘.’ 0.1 ‘ ’ 1

Residual standard error: 0.239 on 45 degrees of freedom
  (8 observations deleted due to missingness)
Multiple R-squared:  0.04705,	Adjusted R-squared:  -0.01648
F-statistic: 0.7405 on 3 and 45 DF,  p-value: 0.5335

>
> mLifeSat_LADNT.Words = lm(lifesat.log ~
+                           Group.ASDref*FirstWords.months
+                 #+ age
+                       #+ vabs_soc_ss
+                         , data = dLADNT.older18)
> summary(mLifeSat_LADNT.Words)

Call:
lm(formula = lifesat.log ~ Group.ASDref * FirstWords.months,
    data = dLADNT.older18)

Residuals:
     Min       1Q   Median       3Q      Max
-0.65422 -0.13430  0.03427  0.16339  0.31813

Coefficients:
                                Estimate Std. Error t value Pr(>|t|)
(Intercept)                     3.211667   0.102711  31.269   <2e-16 ***
Group.ASDref                    0.280223   0.205423   1.364    0.181
FirstWords.months              -0.002466   0.006407  -0.385    0.703
Group.ASDref:FirstWords.months -0.015867   0.012813  -1.238    0.224
---
Signif. codes:  0 ‘***’ 0.001 ‘**’ 0.01 ‘*’ 0.05 ‘.’ 0.1 ‘ ’ 1

Residual standard error: 0.2479 on 35 degrees of freedom
  (18 observations deleted due to missingness)
Multiple R-squared:  0.05308,	Adjusted R-squared:  -0.02808
F-statistic: 0.654 on 3 and 35 DF,  p-value: 0.5858

>
> mLifeSat_LADNT.Phrases = lm(lifesat.log ~
+                           Group.ASDref*FirstPhrases.months
+                 #+ age
+                       #+ vabs_soc_ss
+                         , data = dLADNT.older18)
> summary(mLifeSat_LADNT.Phrases)

Call:
lm(formula = lifesat.log ~ Group.ASDref * FirstPhrases.months,
    data = dLADNT.older18)

Residuals:
     Min       1Q   Median       3Q      Max
-0.62898 -0.16607  0.03723  0.12762  0.35660

Coefficients:
                                  Estimate Std. Error t value Pr(>|t|)
(Intercept)                       3.178545   0.087706  36.241   <2e-16 ***
Group.ASDref                      0.349354   0.175412   1.992   0.0538 .
FirstPhrases.months              -0.001463   0.003756  -0.389   0.6993
Group.ASDref:FirstPhrases.months -0.014775   0.007513  -1.967   0.0568 .
---
Signif. codes:  0 ‘***’ 0.001 ‘**’ 0.01 ‘*’ 0.05 ‘.’ 0.1 ‘ ’ 1

Residual standard error: 0.2393 on 37 degrees of freedom
  (16 observations deleted due to missingness)
Multiple R-squared:  0.1178,	Adjusted R-squared:  0.04632
F-statistic: 1.648 on 3 and 37 DF,  p-value: 0.1951

>
>
>
> ###Follow up
> #aprime.log
> mLifeSat_ASD.GJ = lm(lifesat.log ~ aprime.log
+                 #+ age
+                       #+ vabs_soc_ss
+                         , data = dASD.older18)
> summary(mLifeSat_ASD.GJ)

Call:
lm(formula = lifesat.log ~ aprime.log, data = dASD.older18)

Residuals:
     Min       1Q   Median       3Q      Max
-1.08346 -0.10323  0.03179  0.24716  0.55024

Coefficients:
            Estimate Std. Error t value Pr(>|t|)
(Intercept)   2.7964     0.1845  15.156 1.22e-09 ***
aprime.log   -0.7697     0.7081  -1.087    0.297
---
Signif. codes:  0 ‘***’ 0.001 ‘**’ 0.01 ‘*’ 0.05 ‘.’ 0.1 ‘ ’ 1

Residual standard error: 0.4264 on 13 degrees of freedom
  (10 observations deleted due to missingness)
Multiple R-squared:  0.08331,	Adjusted R-squared:  0.0128
F-statistic: 1.182 on 1 and 13 DF,  p-value: 0.2968

>
> mLifeSat_NT.GJ = lm(lifesat.log ~ aprime.log
+                 #+ age
+                       #+ vabs_soc_ss
+                         , data = dNT.older18)
> summary(mLifeSat_NT.GJ)

Call:
lm(formula = lifesat.log ~ aprime.log, data = dNT.older18)

Residuals:
     Min       1Q   Median       3Q      Max
-0.61630 -0.18505  0.05776  0.18650  0.28399

Coefficients:
            Estimate Std. Error t value Pr(>|t|)
(Intercept) 3.182295   0.095823  33.210   <2e-16 ***
aprime.log  0.009828   0.693625   0.014    0.989
---
Signif. codes:  0 ‘***’ 0.001 ‘**’ 0.01 ‘*’ 0.05 ‘.’ 0.1 ‘ ’ 1

Residual standard error: 0.2471 on 22 degrees of freedom
  (3 observations deleted due to missingness)
Multiple R-squared:  9.125e-06,	Adjusted R-squared:  -0.04545
F-statistic: 0.0002008 on 1 and 22 DF,  p-value: 0.9888

>
> mLifeSat_LAD.GJ = lm(lifesat.log ~ aprime.log
+                 #+ age
+                       #+ vabs_soc_ss
+                         , data = dLAD.older18)
> summary(mLifeSat_LAD.GJ)

Call:
lm(formula = lifesat.log ~ aprime.log, data = dLAD.older18)

Residuals:
     Min       1Q   Median       3Q      Max
-0.55842 -0.10188  0.03534  0.17246  0.39850

Coefficients:
            Estimate Std. Error t value Pr(>|t|)
(Intercept)  3.07356    0.08698   35.34   <2e-16 ***
aprime.log  -0.96402    0.62614   -1.54    0.137
---
Signif. codes:  0 ‘***’ 0.001 ‘**’ 0.01 ‘*’ 0.05 ‘.’ 0.1 ‘ ’ 1

Residual standard error: 0.231 on 23 degrees of freedom
  (5 observations deleted due to missingness)
Multiple R-squared:  0.09343,	Adjusted R-squared:  0.05402
F-statistic:  2.37 on 1 and 23 DF,  p-value: 0.1373

>
> #Words
> mLifeSat_ASD.Words = lm(lifesat.log ~ FirstWords.months
+                 #+ age
+                       #+ vabs_soc_ss
+                         , data = dASD.older18)
> summary(mLifeSat_ASD.Words)

Call:
lm(formula = lifesat.log ~ FirstWords.months, data = dASD.older18)

Residuals:
     Min       1Q   Median       3Q      Max
-1.09645 -0.08031  0.10578  0.27790  0.45525

Coefficients:
                  Estimate Std. Error t value Pr(>|t|)
(Intercept)        2.79039    0.31195   8.945 1.94e-05 ***
FirstWords.months  0.01223    0.01967   0.622    0.551
---
Signif. codes:  0 ‘***’ 0.001 ‘**’ 0.01 ‘*’ 0.05 ‘.’ 0.1 ‘ ’ 1

Residual standard error: 0.4749 on 8 degrees of freedom
  (15 observations deleted due to missingness)
Multiple R-squared:  0.04607,	Adjusted R-squared:  -0.07317
F-statistic: 0.3864 on 1 and 8 DF,  p-value: 0.5515

>
> mLifeSat_NT.Words = lm(lifesat.log ~ FirstWords.months
+                 #+ age
+                       #+ vabs_soc_ss
+                         , data = dNT.older18)
> summary(mLifeSat_NT.Words)

Call:
lm(formula = lifesat.log ~ FirstWords.months, data = dNT.older18)

Residuals:
     Min       1Q   Median       3Q      Max
-0.64123 -0.06129  0.04375  0.15642  0.30116

Coefficients:
                  Estimate Std. Error t value Pr(>|t|)
(Intercept)        3.35178    0.16322  20.535  6.1e-14 ***
FirstWords.months -0.01040    0.01163  -0.895    0.383
---
Signif. codes:  0 ‘***’ 0.001 ‘**’ 0.01 ‘*’ 0.05 ‘.’ 0.1 ‘ ’ 1

Residual standard error: 0.2486 on 18 degrees of freedom
  (7 observations deleted due to missingness)
Multiple R-squared:  0.04256,	Adjusted R-squared:  -0.01063
F-statistic: 0.8002 on 1 and 18 DF,  p-value: 0.3828

>
> mLifeSat_LAD.Words = lm(lifesat.log ~ FirstWords.months
+                 #+ age
+                       #+ vabs_soc_ss
+                         , data = dLAD.older18)
> summary(mLifeSat_LAD.Words)

Call:
lm(formula = lifesat.log ~ FirstWords.months, data = dLAD.older18)

Residuals:
    Min      1Q  Median      3Q     Max
-0.6542 -0.1343  0.0280  0.1634  0.3181

Coefficients:
                  Estimate Std. Error t value Pr(>|t|)
(Intercept)       3.071555   0.124945  24.583    1e-14 ***
FirstWords.months 0.005467   0.005438   1.005    0.329
---
Signif. codes:  0 ‘***’ 0.001 ‘**’ 0.01 ‘*’ 0.05 ‘.’ 0.1 ‘ ’ 1

Residual standard error: 0.2472 on 17 degrees of freedom
  (11 observations deleted due to missingness)
Multiple R-squared:  0.05612,	Adjusted R-squared:  0.0005999
F-statistic: 1.011 on 1 and 17 DF,  p-value: 0.3288

>
> #Phrases
> mLifeSat_ASD.Phrases = lm(lifesat.log ~ FirstPhrases.months
+                 #+ age
+                       #+ vabs_soc_ss
+                         , data = dASD.older18)
> summary(mLifeSat_ASD.Phrases)

Call:
lm(formula = lifesat.log ~ FirstPhrases.months, data = dASD.older18)

Residuals:
    Min      1Q  Median      3Q     Max
-1.1389 -0.1289  0.1726  0.2872  0.5351

Coefficients:
                     Estimate Std. Error t value Pr(>|t|)
(Intercept)          3.042537   0.522699   5.821 0.000396 ***
FirstPhrases.months -0.004662   0.015762  -0.296 0.774949
---
Signif. codes:  0 ‘***’ 0.001 ‘**’ 0.01 ‘*’ 0.05 ‘.’ 0.1 ‘ ’ 1

Residual standard error: 0.5167 on 8 degrees of freedom
  (15 observations deleted due to missingness)
Multiple R-squared:  0.01082,	Adjusted R-squared:  -0.1128
F-statistic: 0.08747 on 1 and 8 DF,  p-value: 0.7749

>
> mLifeSat_NT.Phrases = lm(lifesat.log ~ FirstPhrases.months
+                 #+ age
+                       #+ vabs_soc_ss
+                         , data = dNT.older18)
> summary(mLifeSat_NT.Phrases)

Call:
lm(formula = lifesat.log ~ FirstPhrases.months, data = dNT.older18)

Residuals:
     Min       1Q   Median       3Q      Max
-0.62898 -0.16438  0.06534  0.14753  0.33267

Coefficients:
                     Estimate Std. Error t value Pr(>|t|)
(Intercept)          3.353223   0.140812  23.814 3.77e-16 ***
FirstPhrases.months -0.008850   0.006971  -1.269    0.219
---
Signif. codes:  0 ‘***’ 0.001 ‘**’ 0.01 ‘*’ 0.05 ‘.’ 0.1 ‘ ’ 1

Residual standard error: 0.2475 on 20 degrees of freedom
  (5 observations deleted due to missingness)
Multiple R-squared:  0.07457,	Adjusted R-squared:  0.0283
F-statistic: 1.612 on 1 and 20 DF,  p-value: 0.2188

>
> mLifeSat_LAD.Phrases = lm(lifesat.log ~ FirstPhrases.months
+                 #+ age
+                       #+ vabs_soc_ss
+                         , data = dLAD.older18)
> summary(mLifeSat_LAD.Phrases)

Call:
lm(formula = lifesat.log ~ FirstPhrases.months, data = dLAD.older18)

Residuals:
     Min       1Q   Median       3Q      Max
-0.54556 -0.15422  0.00758  0.11354  0.35660

Coefficients:
                    Estimate Std. Error t value Pr(>|t|)
(Intercept)         3.003868   0.105970  28.346 9.41e-16 ***
FirstPhrases.months 0.005925   0.003178   1.865   0.0796 .
---
Signif. codes:  0 ‘***’ 0.001 ‘**’ 0.01 ‘*’ 0.05 ‘.’ 0.1 ‘ ’ 1

Residual standard error: 0.2294 on 17 degrees of freedom
  (11 observations deleted due to missingness)
Multiple R-squared:  0.1698,	Adjusted R-squared:  0.1209
F-statistic: 3.477 on 1 and 17 DF,  p-value: 0.0796


> ##########################
> ##### Social support #####
> ##########################
> #support.log
>
> ### ASD LAD ###
> mSupp_ASDLAD.GJ = lm(support.log ~
+                           Group.NTref*aprime.log
+                  #+ age
+                       #+ vabs_soc_ss
+                         , data = dASDLAD.older18)
> summary(mSupp_ASDLAD.GJ)

Call:
lm(formula = support.log ~ Group.NTref * aprime.log, data = dASDLAD.older18)

Residuals:
     Min       1Q   Median       3Q      Max
-0.41597 -0.09554  0.00883  0.10470  0.28506

Coefficients:
                       Estimate Std. Error t value Pr(>|t|)
(Intercept)             3.86292    0.04363  88.529   <2e-16 ***
Group.NTref             0.15079    0.08727   1.728   0.0914 .
aprime.log             -0.06109    0.24459  -0.250   0.8040
Group.NTref:aprime.log  0.15893    0.48917   0.325   0.7469
---
Signif. codes:  0 ‘***’ 0.001 ‘**’ 0.01 ‘*’ 0.05 ‘.’ 0.1 ‘ ’ 1

Residual standard error: 0.1603 on 42 degrees of freedom
  (9 observations deleted due to missingness)
Multiple R-squared:  0.1304,	Adjusted R-squared:  0.06831
F-statistic:   2.1 on 3 and 42 DF,  p-value: 0.1147

>
> mSupp_ASDLAD.Words = lm(support.log ~
+                           Group.NTref*FirstWords.months
+                  #+ age
+                       #+ vabs_soc_ss
+                         , data = dASDLAD.older18)
> summary(mSupp_ASDLAD.Words)

Call:
lm(formula = support.log ~ Group.NTref * FirstWords.months, data = dASDLAD.older18)

Residuals:
     Min       1Q   Median       3Q      Max
-0.40452 -0.09162 -0.00282  0.07284  0.28109

Coefficients:
                              Estimate Std. Error t value Pr(>|t|)
(Intercept)                   3.837509   0.059943  64.019   <2e-16 ***
Group.NTref                   0.033439   0.119886   0.279    0.782
FirstWords.months             0.001067   0.003064   0.348    0.730
Group.NTref:FirstWords.months 0.004645   0.006129   0.758    0.455
---
Signif. codes:  0 ‘***’ 0.001 ‘**’ 0.01 ‘*’ 0.05 ‘.’ 0.1 ‘ ’ 1

Residual standard error: 0.1537 on 29 degrees of freedom
  (22 observations deleted due to missingness)
Multiple R-squared:  0.1805,	Adjusted R-squared:  0.0957
F-statistic: 2.129 on 3 and 29 DF,  p-value: 0.1182

>
> mSupp_ASDLAD.Phrases = lm(support.log ~
+                           Group.NTref*FirstPhrases.months
+                  #+ age
+                       #+ vabs_soc_ss
+                         , data = dASDLAD.older18)
> summary(mSupp_ASDLAD.Phrases)

Call:
lm(formula = support.log ~ Group.NTref * FirstPhrases.months,
    data = dASDLAD.older18)

Residuals:
     Min       1Q   Median       3Q      Max
-0.40780 -0.10761  0.00720  0.07273  0.32288

Coefficients:
                                  Estimate Std. Error t value Pr(>|t|)
(Intercept)                      3.8762741  0.0846009  45.818   <2e-16 ***
Group.NTref                     -0.0155786  0.1692017  -0.092    0.927
FirstPhrases.months             -0.0003833  0.0024462  -0.157    0.877
Group.NTref:FirstPhrases.months  0.0054888  0.0048923   1.122    0.271
---
Signif. codes:  0 ‘***’ 0.001 ‘**’ 0.01 ‘*’ 0.05 ‘.’ 0.1 ‘ ’ 1

Residual standard error: 0.1599 on 29 degrees of freedom
  (22 observations deleted due to missingness)
Multiple R-squared:  0.2388,	Adjusted R-squared:   0.16
F-statistic: 3.032 on 3 and 29 DF,  p-value: 0.04514

>
>
>
>
>
> ### ASD NT ###
> mSupp_ASDNT.GJ = lm(support.log ~
+                           Group.LADref*aprime.log
+                 #+ age
+                       #+ vabs_soc_ss
+                         , data = dASDNT.older18)
> summary(mSupp_ASDNT.GJ)

Call:
lm(formula = support.log ~ Group.LADref * aprime.log, data = dASDNT.older18)

Residuals:
     Min       1Q   Median       3Q      Max
-0.51617 -0.08873  0.03382  0.12568  0.28506

Coefficients:
                        Estimate Std. Error t value Pr(>|t|)
(Intercept)              3.85278    0.05190  74.237   <2e-16 ***
Group.LADref             0.13050    0.10380   1.257    0.216
aprime.log              -0.06443    0.29234  -0.220    0.827
Group.LADref:aprime.log  0.15225    0.58467   0.260    0.796
---
Signif. codes:  0 ‘***’ 0.001 ‘**’ 0.01 ‘*’ 0.05 ‘.’ 0.1 ‘ ’ 1

Residual standard error: 0.1911 on 41 degrees of freedom
  (7 observations deleted due to missingness)
Multiple R-squared:  0.07036,	Adjusted R-squared:  0.002333
F-statistic: 1.034 on 3 and 41 DF,  p-value: 0.3875

>
> mSupp_ASDNT.Words = lm(support.log ~
+                           Group.LADref*FirstWords.months
+                 #+ age
+                       #+ vabs_soc_ss
+                         , data = dASDNT.older18)
> summary(mSupp_ASDNT.Words)

Call:
lm(formula = support.log ~ Group.LADref * FirstWords.months,
    data = dASDNT.older18)

Residuals:
     Min       1Q   Median       3Q      Max
-0.43720 -0.04517  0.02102  0.13981  0.28109

Coefficients:
                                Estimate Std. Error t value Pr(>|t|)
(Intercept)                     3.920668   0.091948  42.640   <2e-16 ***
Group.LADref                    0.199757   0.183895   1.086    0.286
FirstWords.months              -0.004588   0.005958  -0.770    0.448
Group.LADref:FirstWords.months -0.006663   0.011915  -0.559    0.580
---
Signif. codes:  0 ‘***’ 0.001 ‘**’ 0.01 ‘*’ 0.05 ‘.’ 0.1 ‘ ’ 1

Residual standard error: 0.2051 on 29 degrees of freedom
  (19 observations deleted due to missingness)
Multiple R-squared:  0.09751,	Adjusted R-squared:  0.004154
F-statistic: 1.044 on 3 and 29 DF,  p-value: 0.3877

>
> mSupp_ASDNT.Phrases = lm(support.log ~
+                           Group.LADref*FirstPhrases.months
+                 #+ age
+                       #+ vabs_soc_ss
+                         , data = dASDNT.older18)
> summary(mSupp_ASDNT.Phrases)

Call:
lm(formula = support.log ~ Group.LADref * FirstPhrases.months,
    data = dASDNT.older18)

Residuals:
     Min       1Q   Median       3Q      Max
-0.50770 -0.09510  0.04252  0.14244  0.32288

Coefficients:
                                  Estimate Std. Error t value Pr(>|t|)
(Intercept)                       3.897016   0.115353  33.783   <2e-16 ***
Group.LADref                      0.025905   0.230706   0.112    0.911
FirstPhrases.months              -0.001585   0.004055  -0.391    0.698
Group.LADref:FirstPhrases.months  0.003085   0.008110   0.380    0.706
---
Signif. codes:  0 ‘***’ 0.001 ‘**’ 0.01 ‘*’ 0.05 ‘.’ 0.1 ‘ ’ 1

Residual standard error: 0.2074 on 31 degrees of freedom
  (17 observations deleted due to missingness)
Multiple R-squared:  0.0977,	Adjusted R-squared:  0.01039
F-statistic: 1.119 on 3 and 31 DF,  p-value: 0.3564

>
>
>
>
> ### LAD NT ###
> mSupp_LADNT.GJ = lm(support.log ~
+                           Group.ASDref*aprime.log
+                 #+ age
+                       #+ vabs_soc_ss
+                         , data = dLADNT.older18)
> summary(mSupp_LADNT.GJ)

Call:
lm(formula = support.log ~ Group.ASDref * aprime.log, data = dLADNT.older18)

Residuals:
     Min       1Q   Median       3Q      Max
-0.51617 -0.04558  0.03382  0.10793  0.17974

Coefficients:
                         Estimate Std. Error t value Pr(>|t|)
(Intercept)              3.928172   0.039285  99.993   <2e-16 ***
Group.ASDref            -0.020293   0.078569  -0.258    0.797
aprime.log               0.015035   0.293608   0.051    0.959
Group.ASDref:aprime.log -0.006684   0.587216  -0.011    0.991
---
Signif. codes:  0 ‘***’ 0.001 ‘**’ 0.01 ‘*’ 0.05 ‘.’ 0.1 ‘ ’ 1

Residual standard error: 0.1574 on 49 degrees of freedom
  (4 observations deleted due to missingness)
Multiple R-squared:  0.004194,	Adjusted R-squared:  -0.05677
F-statistic: 0.06878 on 3 and 49 DF,  p-value: 0.9763

>
> mSupp_LADNT.Words = lm(support.log ~
+                           Group.ASDref*FirstWords.months
+                 #+ age
+                       #+ vabs_soc_ss
+                         , data = dLADNT.older18)
> summary(mSupp_LADNT.Words)

Call:
lm(formula = support.log ~ Group.ASDref * FirstWords.months,
    data = dLADNT.older18)

Residuals:
     Min       1Q   Median       3Q      Max
-0.43720 -0.03723  0.00498  0.11172  0.21635

Coefficients:
                                Estimate Std. Error t value Pr(>|t|)
(Intercept)                     3.937387   0.062665  62.833   <2e-16 ***
Group.ASDref                    0.166318   0.125329   1.327    0.192
FirstWords.months              -0.002265   0.003936  -0.576    0.568
Group.ASDref:FirstWords.months -0.011308   0.007872  -1.437    0.159
---
Signif. codes:  0 ‘***’ 0.001 ‘**’ 0.01 ‘*’ 0.05 ‘.’ 0.1 ‘ ’ 1

Residual standard error: 0.155 on 38 degrees of freedom
  (15 observations deleted due to missingness)
Multiple R-squared:  0.06047,	Adjusted R-squared:  -0.01371
F-statistic: 0.8152 on 3 and 38 DF,  p-value: 0.4935

>
> mSupp_LADNT.Phrases = lm(support.log ~
+                           Group.ASDref*FirstPhrases.months
+                 #+ age
+                       #+ vabs_soc_ss
+                         , data = dLADNT.older18)
> summary(mSupp_LADNT.Phrases)

Call:
lm(formula = support.log ~ Group.ASDref * FirstPhrases.months,
    data = dLADNT.older18)

Residuals:
     Min       1Q   Median       3Q      Max
-0.50770 -0.04459  0.01094  0.11583  0.19280

Coefficients:
                                  Estimate Std. Error t value Pr(>|t|)
(Intercept)                       3.889227   0.057335  67.833   <2e-16 ***
Group.ASDref                      0.041484   0.114671   0.362    0.719
FirstPhrases.months               0.001159   0.002447   0.474    0.638
Group.ASDref:FirstPhrases.months -0.002404   0.004894  -0.491    0.626
---
Signif. codes:  0 ‘***’ 0.001 ‘**’ 0.01 ‘*’ 0.05 ‘.’ 0.1 ‘ ’ 1

Residual standard error: 0.1578 on 40 degrees of freedom
  (13 observations deleted due to missingness)
Multiple R-squared:  0.03775,	Adjusted R-squared:  -0.03442
F-statistic: 0.5231 on 3 and 40 DF,  p-value: 0.6689

>
>
>
>
>
>
> ###Follow up
> #aprime.log
> mSupp_ASD.GJ = lm(support.log ~ aprime.log
+                 #+ age
+                       #+ vabs_soc_ss
+                         , data = dASD.older18)
> summary(mSupp_ASD.GJ)

Call:
lm(formula = support.log ~ aprime.log, data = dASD.older18)

Residuals:
     Min       1Q   Median       3Q      Max
-0.41597 -0.13919  0.00474  0.18916  0.28506

Coefficients:
            Estimate Std. Error t value Pr(>|t|)
(Intercept)  3.78753    0.08282  45.734   <2e-16 ***
aprime.log  -0.14055    0.30852  -0.456    0.654
---
Signif. codes:  0 ‘***’ 0.001 ‘**’ 0.01 ‘*’ 0.05 ‘.’ 0.1 ‘ ’ 1

Residual standard error: 0.2003 on 17 degrees of freedom
  (6 observations deleted due to missingness)
Multiple R-squared:  0.01206,	Adjusted R-squared:  -0.04605
F-statistic: 0.2075 on 1 and 17 DF,  p-value: 0.6545

>
> mSupp_NT.GJ = lm(support.log ~ aprime.log
+                 #+ age
+                       #+ vabs_soc_ss
+                         , data = dNT.older18)
> summary(mSupp_NT.GJ)

Call:
lm(formula = support.log ~ aprime.log, data = dNT.older18)

Residuals:
     Min       1Q   Median       3Q      Max
-0.51617 -0.02485  0.05351  0.12180  0.17974

Coefficients:
            Estimate Std. Error t value Pr(>|t|)
(Intercept)  3.91803    0.06490  60.366   <2e-16 ***
aprime.log   0.01169    0.48731   0.024    0.981
---
Signif. codes:  0 ‘***’ 0.001 ‘**’ 0.01 ‘*’ 0.05 ‘.’ 0.1 ‘ ’ 1

Residual standard error: 0.1844 on 24 degrees of freedom
  (1 observation deleted due to missingness)
Multiple R-squared:  2.399e-05,	Adjusted R-squared:  -0.04164
F-statistic: 0.0005758 on 1 and 24 DF,  p-value: 0.9811

>
> mSupp_LAD.GJ = lm(support.log ~ aprime.log
+                 #+ age
+                       #+ vabs_soc_ss
+                         , data = dLAD.older18)
> summary(mSupp_LAD.GJ)

Call:
lm(formula = support.log ~ aprime.log, data = dLAD.older18)

Residuals:
     Min       1Q   Median       3Q      Max
-0.32552 -0.06472  0.01292  0.08995  0.15791

Coefficients:
            Estimate Std. Error t value Pr(>|t|)
(Intercept)  3.93832    0.04465  88.214   <2e-16 ***
aprime.log   0.01838    0.33215   0.055    0.956
---
Signif. codes:  0 ‘***’ 0.001 ‘**’ 0.01 ‘*’ 0.05 ‘.’ 0.1 ‘ ’ 1

Residual standard error: 0.1261 on 25 degrees of freedom
  (3 observations deleted due to missingness)
Multiple R-squared:  0.0001224,	Adjusted R-squared:  -0.03987
F-statistic: 0.003061 on 1 and 25 DF,  p-value: 0.9563

>
>
> #Words
> mSupp_ASD.Words = lm(support.log ~ FirstWords.months
+                 #+ age
+                       #+ vabs_soc_ss
+                         , data = dASD.older18)
> summary(mSupp_ASD.Words)

Call:
lm(formula = support.log ~ FirstWords.months, data = dASD.older18)

Residuals:
     Min       1Q   Median       3Q      Max
-0.40452 -0.11962 -0.00534  0.20612  0.28109

Coefficients:
                   Estimate Std. Error t value Pr(>|t|)
(Intercept)        3.820789   0.137160  27.856 8.25e-11 ***
FirstWords.months -0.001256   0.007693  -0.163    0.874
---
Signif. codes:  0 ‘***’ 0.001 ‘**’ 0.01 ‘*’ 0.05 ‘.’ 0.1 ‘ ’ 1

Residual standard error: 0.2227 on 10 degrees of freedom
  (13 observations deleted due to missingness)
Multiple R-squared:  0.002658,	Adjusted R-squared:  -0.09708
F-statistic: 0.02665 on 1 and 10 DF,  p-value: 0.8736

>
> mSupp_NT.Words = lm(support.log ~ FirstWords.months
+                 #+ age
+                       #+ vabs_soc_ss
+                         , data = dNT.older18)
> summary(mSupp_NT.Words)

Call:
lm(formula = support.log ~ FirstWords.months, data = dNT.older18)

Residuals:
     Min       1Q   Median       3Q      Max
-0.43720 -0.03352  0.04478  0.13493  0.21635

Coefficients:
                   Estimate Std. Error t value Pr(>|t|)
(Intercept)        4.020546   0.127190  31.611   <2e-16 ***
FirstWords.months -0.007919   0.009118  -0.869    0.396
---
Signif. codes:  0 ‘***’ 0.001 ‘**’ 0.01 ‘*’ 0.05 ‘.’ 0.1 ‘ ’ 1

Residual standard error: 0.1953 on 19 degrees of freedom
  (6 observations deleted due to missingness)
Multiple R-squared:  0.03819,	Adjusted R-squared:  -0.01244
F-statistic: 0.7543 on 1 and 19 DF,  p-value: 0.396

>
> mSupp_LAD.Words = lm(support.log ~ FirstWords.months
+                 #+ age
+                       #+ vabs_soc_ss
+                         , data = dLAD.older18)
> summary(mSupp_LAD.Words)

Call:
lm(formula = support.log ~ FirstWords.months, data = dLAD.older18)

Residuals:
      Min        1Q    Median        3Q       Max
-0.161545 -0.064366 -0.001269  0.017125  0.199447

Coefficients:
                  Estimate Std. Error t value Pr(>|t|)
(Intercept)       3.854228   0.047749  80.718   <2e-16 ***
FirstWords.months 0.003389   0.001988   1.705    0.104
---
Signif. codes:  0 ‘***’ 0.001 ‘**’ 0.01 ‘*’ 0.05 ‘.’ 0.1 ‘ ’ 1

Residual standard error: 0.09976 on 19 degrees of freedom
  (9 observations deleted due to missingness)
Multiple R-squared:  0.1327,	Adjusted R-squared:  0.08706
F-statistic: 2.907 on 1 and 19 DF,  p-value: 0.1045

>
> #Phrases
> mSupp_ASD.Phrases = lm(support.log ~ FirstPhrases.months
+                 #+ age
+                       #+ vabs_soc_ss
+                         , data = dASD.older18)
> summary(mSupp_ASD.Phrases)

Call:
lm(formula = support.log ~ FirstPhrases.months, data = dASD.older18)

Residuals:
     Min       1Q   Median       3Q      Max
-0.40780 -0.15566 -0.03147  0.19777  0.32288

Coefficients:
                     Estimate Std. Error t value Pr(>|t|)
(Intercept)          3.884063   0.222091  17.489 7.93e-09 ***
FirstPhrases.months -0.003128   0.006368  -0.491    0.634
---
Signif. codes:  0 ‘***’ 0.001 ‘**’ 0.01 ‘*’ 0.05 ‘.’ 0.1 ‘ ’ 1

Residual standard error: 0.2323 on 10 degrees of freedom
  (13 observations deleted due to missingness)
Multiple R-squared:  0.02356,	Adjusted R-squared:  -0.07409
F-statistic: 0.2413 on 1 and 10 DF,  p-value: 0.6339

>
> mSupp_NT.Phrases = lm(support.log ~ FirstPhrases.months
+                 #+ age
+                       #+ vabs_soc_ss
+                         , data = dNT.older18)
> summary(mSupp_NT.Phrases)

Call:
lm(formula = support.log ~ FirstPhrases.months, data = dNT.older18)

Residuals:
     Min       1Q   Median       3Q      Max
-0.50770 -0.02730  0.06084  0.13377  0.18541

Coefficients:
                      Estimate Std. Error t value Pr(>|t|)
(Intercept)          3.910e+00  1.105e-01  35.390   <2e-16 ***
FirstPhrases.months -4.298e-05  5.421e-03  -0.008    0.994
---
Signif. codes:  0 ‘***’ 0.001 ‘**’ 0.01 ‘*’ 0.05 ‘.’ 0.1 ‘ ’ 1

Residual standard error: 0.1945 on 21 degrees of freedom
  (4 observations deleted due to missingness)
Multiple R-squared:  2.993e-06,	Adjusted R-squared:  -0.04762
F-statistic: 6.285e-05 on 1 and 21 DF,  p-value: 0.9937

>
> mSupp_LAD.Phrases = lm(support.log ~ FirstPhrases.months
+                 #+ age
+                       #+ vabs_soc_ss
+                         , data = dLAD.older18)
> summary(mSupp_LAD.Phrases)

Call:
lm(formula = support.log ~ FirstPhrases.months, data = dLAD.older18)

Residuals:
      Min        1Q    Median        3Q       Max
-0.173315 -0.053950  0.007204  0.039682  0.192804

Coefficients:
                    Estimate Std. Error t value Pr(>|t|)
(Intercept)         3.868485   0.046720  82.801   <2e-16 ***
FirstPhrases.months 0.002361   0.001400   1.686    0.108
---
Signif. codes:  0 ‘***’ 0.001 ‘**’ 0.01 ‘*’ 0.05 ‘.’ 0.1 ‘ ’ 1

Residual standard error: 0.1032 on 19 degrees of freedom
  (9 observations deleted due to missingness)
Multiple R-squared:  0.1302,	Adjusted R-squared:  0.08439
F-statistic: 2.843 on 1 and 19 DF,  p-value: 0.1081


> ### ABCL/CBCL
> # Relationships: abcl_friends_t, abcl_spouse_t, cbcl_friends1 (1. About how many close friends does your child have? (Do not include brothers and sisters), cbcl_friends2 (2. About how many times a week does your child do things with any friends outside of regular school hours? (Do not include brothers and sisters), cbcl_social_t
> #USING COMBINED SCORES
> ###################
> ##### Friends #####
> ###################
> #combined friends and social
>
> ### ASD LAD ###
> mFriend.AB_ASDLAD.GJ = lm(friends ~
+                           Group.NTref*aprime.log
+                  #+ age
+                       #+ vabs_soc_ss
+                         , data = dASDLAD)
> summary(mFriend.AB_ASDLAD.GJ)

Call:
lm(formula = friends ~ Group.NTref * aprime.log, data = dASDLAD)

Residuals:
     Min       1Q   Median       3Q      Max
-20.4192  -6.6741  -0.4192   5.6712  18.5508

Coefficients:
                       Estimate Std. Error t value Pr(>|t|)
(Intercept)              46.296      2.287  20.247   <2e-16 ***
Group.NTref              10.599      4.573   2.318   0.0249 *
aprime.log                3.278     13.055   0.251   0.8029
Group.NTref:aprime.log  -38.422     26.110  -1.472   0.1478
---
Signif. codes:  0 ‘***’ 0.001 ‘**’ 0.01 ‘*’ 0.05 ‘.’ 0.1 ‘ ’ 1

Residual standard error: 9.224 on 47 degrees of freedom
  (20 observations deleted due to missingness)
Multiple R-squared:  0.4763,	Adjusted R-squared:  0.4429
F-statistic: 14.25 on 3 and 47 DF,  p-value: 9.818e-07

>
> mFriend.AB_ASDLAD.Words = lm(friends ~
+                           Group.NTref*FirstWords.months
+                  #+ age
+                       #+ vabs_soc_ss
+                         , data = dASDLAD)
> summary(mFriend.AB_ASDLAD.Words)

Call:
lm(formula = friends ~ Group.NTref * FirstWords.months, data = dASDLAD)

Residuals:
     Min       1Q   Median       3Q      Max
-21.1763  -4.2372   0.0345   5.4699  19.0116

Coefficients:
                              Estimate Std. Error t value Pr(>|t|)
(Intercept)                    50.0918     2.8806  17.389   <2e-16 ***
Group.NTref                    10.7042     5.7613   1.858   0.0704 .
FirstWords.months              -0.2219     0.1245  -1.783   0.0820 .
Group.NTref:FirstWords.months   0.3481     0.2489   1.399   0.1695
---
Signif. codes:  0 ‘***’ 0.001 ‘**’ 0.01 ‘*’ 0.05 ‘.’ 0.1 ‘ ’ 1

Residual standard error: 8.699 on 41 degrees of freedom
  (26 observations deleted due to missingness)
Multiple R-squared:  0.5469,	Adjusted R-squared:  0.5137
F-statistic:  16.5 on 3 and 41 DF,  p-value: 3.51e-07

>
> mFriend.AB_ASDLAD.Phrases = lm(friends ~
+                           Group.NTref*FirstPhrases.months
+                  #+ age
+                       #+ vabs_soc_ss
+                         , data = dASDLAD)
> summary(mFriend.AB_ASDLAD.Phrases)

Call:
lm(formula = friends ~ Group.NTref * FirstPhrases.months, data = dASDLAD)

Residuals:
     Min       1Q   Median       3Q      Max
-17.3211  -6.5922   0.8914   5.4584  20.4078

Coefficients:
                                Estimate Std. Error t value Pr(>|t|)
(Intercept)                     47.83834    3.62167  13.209 2.31e-16 ***
Group.NTref                      9.32796    7.24334   1.288    0.205
FirstPhrases.months             -0.07381    0.10787  -0.684    0.498
Group.NTref:FirstPhrases.months  0.23423    0.21573   1.086    0.284
---
Signif. codes:  0 ‘***’ 0.001 ‘**’ 0.01 ‘*’ 0.05 ‘.’ 0.1 ‘ ’ 1

Residual standard error: 9.112 on 41 degrees of freedom
  (26 observations deleted due to missingness)
Multiple R-squared:  0.4876,	Adjusted R-squared:  0.4501
F-statistic: 13.01 on 3 and 41 DF,  p-value: 4.139e-06

>
>
>
>
> ### ASD NT ###
> mFriend.AB_ASDNT.GJ = lm(friends ~
+                           Group.LADref*aprime.log
+                 #+ age
+                       #+ vabs_soc_ss
+                         , data = dASDNT)
> summary(mFriend.AB_ASDNT.GJ)

Call:
lm(formula = friends ~ Group.LADref * aprime.log, data = dASDNT)

Residuals:
     Min       1Q   Median       3Q      Max
-25.9655  -6.9067   0.8899   6.1327  18.5508

Coefficients:
                        Estimate Std. Error t value Pr(>|t|)
(Intercept)              48.1422     2.6576  18.115  < 2e-16 ***
Group.LADref             14.2901     5.3151   2.689  0.00972 **
aprime.log               22.9757    15.6849   1.465  0.14923
Group.LADref:aprime.log   0.9744    31.3699   0.031  0.97534
---
Signif. codes:  0 ‘***’ 0.001 ‘**’ 0.01 ‘*’ 0.05 ‘.’ 0.1 ‘ ’ 1

Residual standard error: 9.895 on 50 degrees of freedom
  (23 observations deleted due to missingness)
Multiple R-squared:  0.4032,	Adjusted R-squared:  0.3674
F-statistic: 11.26 on 3 and 50 DF,  p-value: 9.285e-06

>
> mFriend.AB_ASDNT.Words = lm(friends ~
+                           Group.LADref*FirstWords.months
+                 #+ age
+                       #+ vabs_soc_ss
+                         , data = dASDNT)
> summary(mFriend.AB_ASDNT.Words)

Call:
lm(formula = friends ~ Group.LADref * FirstWords.months, data = dASDNT)

Residuals:
    Min      1Q  Median      3Q     Max
-21.176  -5.362   1.061   6.663  19.012

Coefficients:
                               Estimate Std. Error t value Pr(>|t|)
(Intercept)                     53.6001     3.8657  13.865   <2e-16 ***
Group.LADref                    17.7208     7.7314   2.292   0.0267 *
FirstWords.months               -0.5968     0.2510  -2.378   0.0218 *
Group.LADref:FirstWords.months  -0.4016     0.5019  -0.800   0.4279
---
Signif. codes:  0 ‘***’ 0.001 ‘**’ 0.01 ‘*’ 0.05 ‘.’ 0.1 ‘ ’ 1

Residual standard error: 9.879 on 44 degrees of freedom
  (29 observations deleted due to missingness)
Multiple R-squared:  0.4358,	Adjusted R-squared:  0.3973
F-statistic: 11.33 on 3 and 44 DF,  p-value: 1.244e-05

>
> mFriend.AB_ASDNT.Phrases = lm(friends ~
+                           Group.LADref*FirstPhrases.months
+                 #+ age
+                       #+ vabs_soc_ss
+                         , data = dASDNT)
> summary(mFriend.AB_ASDNT.Phrases)

Call:
lm(formula = friends ~ Group.LADref * FirstPhrases.months, data = dASDNT)

Residuals:
    Min      1Q  Median      3Q     Max
-21.992  -6.829   2.356   6.653  20.408

Coefficients:
                                 Estimate Std. Error t value Pr(>|t|)
(Intercept)                       51.3191     4.5291  11.331    9e-15 ***
Group.LADref                      16.2895     9.0582   1.798   0.0788 .
FirstPhrases.months               -0.2849     0.1792  -1.590   0.1188
Group.LADref:FirstPhrases.months  -0.1879     0.3583  -0.524   0.6025
---
Signif. codes:  0 ‘***’ 0.001 ‘**’ 0.01 ‘*’ 0.05 ‘.’ 0.1 ‘ ’ 1

Residual standard error: 10.23 on 45 degrees of freedom
  (28 observations deleted due to missingness)
Multiple R-squared:  0.3894,	Adjusted R-squared:  0.3487
F-statistic: 9.565 on 3 and 45 DF,  p-value: 5.303e-05

>
>
>
>
> ### LAD NT ###
> mFriend.AB_LADNT.GJ = lm(friends ~
+                           Group.ASDref*aprime.log
+                 #+ age
+                       #+ vabs_soc_ss
+                         , data = dLADNT)
> summary(mFriend.AB_LADNT.GJ)

Call:
lm(formula = friends ~ Group.ASDref * aprime.log, data = dLADNT)

Residuals:
    Min      1Q  Median      3Q     Max
-25.965  -1.461   1.034   5.833  11.583

Coefficients:
                        Estimate Std. Error t value Pr(>|t|)
(Intercept)               53.441      2.028  26.350   <2e-16 ***
Group.ASDref               3.691      4.056   0.910    0.367
aprime.log                 3.765     13.454   0.280    0.781
Group.ASDref:aprime.log   39.396     26.908   1.464    0.150
---
Signif. codes:  0 ‘***’ 0.001 ‘**’ 0.01 ‘*’ 0.05 ‘.’ 0.1 ‘ ’ 1

Residual standard error: 7.96 on 49 degrees of freedom
  (17 observations deleted due to missingness)
Multiple R-squared:  0.04853,	Adjusted R-squared:  -0.009721
F-statistic: 0.8331 on 3 and 49 DF,  p-value: 0.4821

>
> mFriend.AB_LADNT.Words = lm(friends ~
+                           Group.ASDref*FirstWords.months
+                 #+ age
+                       #+ vabs_soc_ss
+                         , data = dLADNT)
> summary(mFriend.AB_LADNT.Words)

Call:
lm(formula = friends ~ Group.ASDref * FirstWords.months, data = dLADNT)

Residuals:
     Min       1Q   Median       3Q      Max
-17.8896  -3.0564   0.7522   4.8958  11.8958

Coefficients:
                               Estimate Std. Error t value Pr(>|t|)
(Intercept)                     58.9522     2.9066  20.282   <2e-16 ***
Group.ASDref                     7.0165     5.8132   1.207   0.2337
FirstWords.months               -0.4227     0.1829  -2.311   0.0254 *
Group.ASDref:FirstWords.months  -0.7497     0.3658  -2.050   0.0462 *
---
Signif. codes:  0 ‘***’ 0.001 ‘**’ 0.01 ‘*’ 0.05 ‘.’ 0.1 ‘ ’ 1

Residual standard error: 7.229 on 45 degrees of freedom
  (21 observations deleted due to missingness)
Multiple R-squared:  0.1359,	Adjusted R-squared:  0.0783
F-statistic: 2.359 on 3 and 45 DF,  p-value: 0.08413

>
> mFriend.AB_LADNT.Phrases = lm(friends ~
+                           Group.ASDref*FirstPhrases.months
+                 #+ age
+                       #+ vabs_soc_ss
+                         , data = dLADNT)
> summary(mFriend.AB_LADNT.Phrases)

Call:
lm(formula = friends ~ Group.ASDref * FirstPhrases.months, data = dLADNT)

Residuals:
    Min      1Q  Median      3Q     Max
-21.992  -2.477   1.112   5.369   9.629

Coefficients:
                                 Estimate Std. Error t value Pr(>|t|)
(Intercept)                       55.9831     2.7740  20.181   <2e-16 ***
Group.ASDref                       6.9615     5.5480   1.255   0.2159
FirstPhrases.months               -0.1678     0.1190  -1.409   0.1655
Group.ASDref:FirstPhrases.months  -0.4222     0.2381  -1.773   0.0828 .
---
Signif. codes:  0 ‘***’ 0.001 ‘**’ 0.01 ‘*’ 0.05 ‘.’ 0.1 ‘ ’ 1

Residual standard error: 7.39 on 46 degrees of freedom
  (20 observations deleted due to missingness)
Multiple R-squared:  0.0782,	Adjusted R-squared:  0.01809
F-statistic: 1.301 on 3 and 46 DF,  p-value: 0.2855

>
>
>
>
> ###Follow up
> #aprime.log
> mFriend.AB_ASD.GJ = lm(friends ~ aprime.log
+                 #+ age
+                       #+ vabs_soc_ss
+                         , data = dASD)
> summary(mFriend.AB_ASD.GJ)

Call:
lm(formula = friends ~ aprime.log, data = dASD)

Residuals:
     Min       1Q   Median       3Q      Max
-14.7303  -9.2281  -0.9441   7.9660  18.5508

Coefficients:
            Estimate Std. Error t value Pr(>|t|)
(Intercept)    41.00       4.07  10.073 4.27e-10 ***
aprime.log     22.49      20.01   1.124    0.272
---
Signif. codes:  0 ‘***’ 0.001 ‘**’ 0.01 ‘*’ 0.05 ‘.’ 0.1 ‘ ’ 1

Residual standard error: 10.98 on 24 degrees of freedom
  (13 observations deleted due to missingness)
Multiple R-squared:   0.05,	Adjusted R-squared:  0.01042
F-statistic: 1.263 on 1 and 24 DF,  p-value: 0.2722

>
> mFriend.AB_NT.GJ = lm(friends ~ aprime.log
+                 #+ age
+                       #+ vabs_soc_ss
+                         , data = dNT)
> summary(mFriend.AB_NT.GJ)

Call:
lm(formula = friends ~ aprime.log, data = dNT)

Residuals:
    Min      1Q  Median      3Q     Max
-25.965  -1.028   1.202   5.994  11.583

Coefficients:
            Estimate Std. Error t value Pr(>|t|)
(Intercept)   55.287      3.411  16.209 4.15e-15 ***
aprime.log    23.463     22.761   1.031    0.312
---
Signif. codes:  0 ‘***’ 0.001 ‘**’ 0.01 ‘*’ 0.05 ‘.’ 0.1 ‘ ’ 1

Residual standard error: 8.773 on 26 degrees of freedom
  (10 observations deleted due to missingness)
Multiple R-squared:  0.03927,	Adjusted R-squared:  0.002315
F-statistic: 1.063 on 1 and 26 DF,  p-value: 0.3121

>
> mFriend.AB_LAD.GJ = lm(friends ~ aprime.log
+                 #+ age
+                       #+ vabs_soc_ss
+                         , data = dLAD)
> summary(mFriend.AB_LAD.GJ)

Call:
lm(formula = friends ~ aprime.log, data = dLAD)

Residuals:
     Min       1Q   Median       3Q      Max
-20.4192  -2.2860   0.9383   4.8674   7.5808

Coefficients:
            Estimate Std. Error t value Pr(>|t|)
(Intercept)   51.596      2.282  22.611   <2e-16 ***
aprime.log   -15.933     15.011  -1.061      0.3
---
Signif. codes:  0 ‘***’ 0.001 ‘**’ 0.01 ‘*’ 0.05 ‘.’ 0.1 ‘ ’ 1

Residual standard error: 6.929 on 23 degrees of freedom
  (7 observations deleted due to missingness)
Multiple R-squared:  0.0467,	Adjusted R-squared:  0.005247
F-statistic: 1.127 on 1 and 23 DF,  p-value: 0.2995

>
> #Words
> mFriend.AB_ASD.Words = lm(friends ~ FirstWords.months
+                 #+ age
+                       #+ vabs_soc_ss
+                         , data = dASD)
> summary(mFriend.AB_ASD.Words)

Call:
lm(formula = friends ~ FirstWords.months, data = dASD)

Residuals:
    Min      1Q  Median      3Q     Max
-21.176  -7.836  -2.425   9.866  19.012

Coefficients:
                  Estimate Std. Error t value Pr(>|t|)
(Intercept)        44.7397     5.0502   8.859 2.33e-08 ***
FirstWords.months  -0.3959     0.2302  -1.720    0.101
---
Signif. codes:  0 ‘***’ 0.001 ‘**’ 0.01 ‘*’ 0.05 ‘.’ 0.1 ‘ ’ 1

Residual standard error: 11.23 on 20 degrees of freedom
  (17 observations deleted due to missingness)
Multiple R-squared:  0.1289,	Adjusted R-squared:  0.08533
F-statistic: 2.959 on 1 and 20 DF,  p-value: 0.1008

>
> mFriend.AB_NT.Words = lm(friends ~ FirstWords.months
+                 #+ age
+                       #+ vabs_soc_ss
+                         , data = dNT)
> summary(mFriend.AB_NT.Words)

Call:
lm(formula = friends ~ FirstWords.months, data = dNT)

Residuals:
    Min      1Q  Median      3Q     Max
-17.890  -2.750   2.212   5.863  11.896

Coefficients:
                  Estimate Std. Error t value Pr(>|t|)
(Intercept)        62.4605     5.5032  11.350 3.92e-11 ***
FirstWords.months  -0.7976     0.3994  -1.997   0.0573 .
---
Signif. codes:  0 ‘***’ 0.001 ‘**’ 0.01 ‘*’ 0.05 ‘.’ 0.1 ‘ ’ 1

Residual standard error: 8.592 on 24 degrees of freedom
  (12 observations deleted due to missingness)
Multiple R-squared:  0.1425,	Adjusted R-squared:  0.1067
F-statistic: 3.987 on 1 and 24 DF,  p-value: 0.05732

>
> mFriend.AB_LAD.Words = lm(friends ~ FirstWords.months
+                 #+ age
+                       #+ vabs_soc_ss
+                         , data = dLAD)
> summary(mFriend.AB_LAD.Words)

Call:
lm(formula = friends ~ FirstWords.months, data = dLAD)

Residuals:
     Min       1Q   Median       3Q      Max
-16.4344  -3.0564   0.1781   4.4173   6.8527

Coefficients:
                  Estimate Std. Error t value Pr(>|t|)
(Intercept)       55.44394    2.55497  21.700 7.29e-16 ***
FirstWords.months -0.04785    0.10492  -0.456    0.653
---
Signif. codes:  0 ‘***’ 0.001 ‘**’ 0.01 ‘*’ 0.05 ‘.’ 0.1 ‘ ’ 1

Residual standard error: 5.254 on 21 degrees of freedom
  (9 observations deleted due to missingness)
Multiple R-squared:  0.009807,	Adjusted R-squared:  -0.03735
F-statistic: 0.208 on 1 and 21 DF,  p-value: 0.653

>
> #Phrases
> mFriend.AB_ASD.Phrases = lm(friends ~ FirstPhrases.months
+                 #+ age
+                       #+ vabs_soc_ss
+                         , data = dASD)
> summary(mFriend.AB_ASD.Phrases)

Call:
lm(formula = friends ~ FirstPhrases.months, data = dASD)

Residuals:
    Min      1Q  Median      3Q     Max
-15.919 -10.647  -0.874  10.344  20.408

Coefficients:
                    Estimate Std. Error t value Pr(>|t|)
(Intercept)          43.1744     7.7253   5.589  1.8e-05 ***
FirstPhrases.months  -0.1909     0.2292  -0.833    0.415
---
Signif. codes:  0 ‘***’ 0.001 ‘**’ 0.01 ‘*’ 0.05 ‘.’ 0.1 ‘ ’ 1

Residual standard error: 11.83 on 20 degrees of freedom
  (17 observations deleted due to missingness)
Multiple R-squared:  0.03354,	Adjusted R-squared:  -0.01478
F-statistic: 0.6941 on 1 and 20 DF,  p-value: 0.4146

>
> mFriend.AB_NT.Phrases = lm(friends ~ FirstPhrases.months
+                 #+ age
+                       #+ vabs_soc_ss
+                         , data = dNT)
> summary(mFriend.AB_NT.Phrases)

Call:
lm(formula = friends ~ FirstPhrases.months, data = dNT)

Residuals:
    Min      1Q  Median      3Q     Max
-21.992  -2.660   2.629   6.166   9.629

Coefficients:
                    Estimate Std. Error t value Pr(>|t|)
(Intercept)          59.4638     5.2268  11.377 2.23e-11 ***
FirstPhrases.months  -0.3789     0.2550  -1.486     0.15
---
Signif. codes:  0 ‘***’ 0.001 ‘**’ 0.01 ‘*’ 0.05 ‘.’ 0.1 ‘ ’ 1

Residual standard error: 8.732 on 25 degrees of freedom
  (11 observations deleted due to missingness)
Multiple R-squared:  0.08112,	Adjusted R-squared:  0.04437
F-statistic: 2.207 on 1 and 25 DF,  p-value: 0.1499

>
> mFriend.AB_LAD.Phrases = lm(friends ~ FirstPhrases.months
+                 #+ age
+                       #+ vabs_soc_ss
+                         , data = dLAD)
> summary(mFriend.AB_LAD.Phrases)

Call:
lm(formula = friends ~ FirstPhrases.months, data = dLAD)

Residuals:
     Min       1Q   Median       3Q      Max
-17.3211  -2.4117   0.8914   3.6396   6.7182

Coefficients:
                    Estimate Std. Error t value Pr(>|t|)
(Intercept)         52.50232    2.43393  21.571 8.22e-16 ***
FirstPhrases.months  0.04330    0.07309   0.592     0.56
---
Signif. codes:  0 ‘***’ 0.001 ‘**’ 0.01 ‘*’ 0.05 ‘.’ 0.1 ‘ ’ 1

Residual standard error: 5.371 on 21 degrees of freedom
  (9 observations deleted due to missingness)
Multiple R-squared:  0.01644,	Adjusted R-squared:  -0.0304
F-statistic: 0.351 on 1 and 21 DF,  p-value: 0.5599


> ######################
> ##### Total Prob #####
> ######################
> #combined
>
> ### ASD LAD ###
> mProb.AB_ASDLAD.GJ = lm(TotalProb ~
+                           Group.NTref*aprime.log
+                  #+ age
+                       #+ vabs_soc_ss
+                         , data = dASDLAD)
> summary(mProb.AB_ASDLAD.GJ)

Call:
lm(formula = TotalProb ~ Group.NTref * aprime.log, data = dASDLAD)

Residuals:
    Min      1Q  Median      3Q     Max
-32.045  -9.669   1.482   9.333  21.050

Coefficients:
                       Estimate Std. Error t value Pr(>|t|)
(Intercept)              51.073      2.955  17.286   <2e-16 ***
Group.NTref             -15.019      5.909  -2.542   0.0143 *
aprime.log               -4.835     16.946  -0.285   0.7766
Group.NTref:aprime.log   -5.285     33.891  -0.156   0.8767
---
Signif. codes:  0 ‘***’ 0.001 ‘**’ 0.01 ‘*’ 0.05 ‘.’ 0.1 ‘ ’ 1

Residual standard error: 11.98 on 48 degrees of freedom
  (19 observations deleted due to missingness)
Multiple R-squared:  0.2851,	Adjusted R-squared:  0.2405
F-statistic: 6.382 on 3 and 48 DF,  p-value: 0.0009965

>
> mProb.AB_ASDLAD.Words = lm(TotalProb ~
+                           Group.NTref*FirstWords.months
+                  #+ age
+                       #+ vabs_soc_ss
+                         , data = dASDLAD)
> summary(mProb.AB_ASDLAD.Words)

Call:
lm(formula = TotalProb ~ Group.NTref * FirstWords.months, data = dASDLAD)

Residuals:
     Min       1Q   Median       3Q      Max
-30.2490  -9.5692   0.2997   9.2358  20.2760

Coefficients:
                              Estimate Std. Error t value Pr(>|t|)
(Intercept)                    48.3946     3.9594  12.223 2.97e-15 ***
Group.NTref                   -10.0286     7.9189  -1.266    0.213
FirstWords.months               0.1760     0.1708   1.030    0.309
Group.NTref:FirstWords.months  -0.2880     0.3417  -0.843    0.404
---
Signif. codes:  0 ‘***’ 0.001 ‘**’ 0.01 ‘*’ 0.05 ‘.’ 0.1 ‘ ’ 1

Residual standard error: 11.89 on 41 degrees of freedom
  (26 observations deleted due to missingness)
Multiple R-squared:  0.3373,	Adjusted R-squared:  0.2888
F-statistic: 6.955 on 3 and 41 DF,  p-value: 0.0006856

>
> mProb.AB_ASDLAD.Phrases = lm(TotalProb ~
+                           Group.NTref*FirstPhrases.months
+                  #+ age
+                       #+ vabs_soc_ss
+                         , data = dASDLAD)
> summary(mProb.AB_ASDLAD.Phrases)

Call:
lm(formula = TotalProb ~ Group.NTref * FirstPhrases.months, data = dASDLAD)

Residuals:
    Min      1Q  Median      3Q     Max
-30.925  -9.925   1.929   7.075  18.763

Coefficients:
                                 Estimate Std. Error t value Pr(>|t|)
(Intercept)                      48.53676    4.85215  10.003 1.45e-12 ***
Group.NTref                     -14.70865    9.70430  -1.516    0.137
FirstPhrases.months               0.10254    0.14432   0.711    0.481
Group.NTref:FirstPhrases.months   0.03562    0.28864   0.123    0.902
---
Signif. codes:  0 ‘***’ 0.001 ‘**’ 0.01 ‘*’ 0.05 ‘.’ 0.1 ‘ ’ 1

Residual standard error: 12.09 on 41 degrees of freedom
  (26 observations deleted due to missingness)
Multiple R-squared:  0.2717,	Adjusted R-squared:  0.2184
F-statistic: 5.099 on 3 and 41 DF,  p-value: 0.004318

>
>
>
>
> ### ASD NT ###
> mProb.AB_ASDNT.GJ = lm(TotalProb ~
+                           Group.LADref*aprime.log
+                 #+ age
+                       #+ vabs_soc_ss
+                         , data = dASDNT)
> summary(mProb.AB_ASDNT.GJ)

Call:
lm(formula = TotalProb ~ Group.LADref * aprime.log, data = dASDNT)

Residuals:
    Min      1Q  Median      3Q     Max
-32.045  -6.853   1.293   7.136  17.192

Coefficients:
                        Estimate Std. Error t value Pr(>|t|)
(Intercept)              52.0405     2.9895  17.408   <2e-16 ***
Group.LADref            -13.0853     5.9791  -2.189   0.0334 *
aprime.log                0.9872    17.8280   0.055   0.9561
Group.LADref:aprime.log   6.3586    35.6560   0.178   0.8592
---
Signif. codes:  0 ‘***’ 0.001 ‘**’ 0.01 ‘*’ 0.05 ‘.’ 0.1 ‘ ’ 1

Residual standard error: 11.12 on 49 degrees of freedom
  (24 observations deleted due to missingness)
Multiple R-squared:  0.3003,	Adjusted R-squared:  0.2575
F-statistic: 7.011 on 3 and 49 DF,  p-value: 0.000514

>
> mProb.AB_ASDNT.Words = lm(TotalProb ~
+                           Group.LADref*FirstWords.months
+                 #+ age
+                       #+ vabs_soc_ss
+                         , data = dASDNT)
> summary(mProb.AB_ASDNT.Words)

Call:
lm(formula = TotalProb ~ Group.LADref * FirstWords.months, data = dASDNT)

Residuals:
     Min       1Q   Median       3Q      Max
-30.2490  -7.0260  -0.1691   6.7726  20.0310

Coefficients:
                               Estimate Std. Error t value Pr(>|t|)
(Intercept)                     51.7351     4.2607  12.142 1.74e-15 ***
Group.LADref                    -3.3477     8.5214  -0.393    0.696
FirstWords.months               -0.0498     0.2806  -0.177    0.860
Group.LADref:FirstWords.months  -0.7396     0.5612  -1.318    0.195
---
Signif. codes:  0 ‘***’ 0.001 ‘**’ 0.01 ‘*’ 0.05 ‘.’ 0.1 ‘ ’ 1

Residual standard error: 10.79 on 43 degrees of freedom
  (30 observations deleted due to missingness)
Multiple R-squared:  0.3696,	Adjusted R-squared:  0.3257
F-statistic: 8.405 on 3 and 43 DF,  p-value: 0.0001648

>
> mProb.AB_ASDNT.Phrases = lm(TotalProb ~
+                           Group.LADref*FirstPhrases.months
+                 #+ age
+                       #+ vabs_soc_ss
+                         , data = dASDNT)
> summary(mProb.AB_ASDNT.Phrases)

Call:
lm(formula = TotalProb ~ Group.LADref * FirstPhrases.months,
    data = dASDNT)

Residuals:
    Min      1Q  Median      3Q     Max
-30.925  -7.154   1.813   7.804  18.075

Coefficients:
                                 Estimate Std. Error t value Pr(>|t|)
(Intercept)                      52.52394    4.99321  10.519 1.37e-13 ***
Group.LADref                     -6.73427    9.98641  -0.674    0.504
FirstPhrases.months              -0.06443    0.19718  -0.327    0.745
Group.LADref:FirstPhrases.months -0.29832    0.39436  -0.756    0.453
---
Signif. codes:  0 ‘***’ 0.001 ‘**’ 0.01 ‘*’ 0.05 ‘.’ 0.1 ‘ ’ 1

Residual standard error: 11.25 on 44 degrees of freedom
  (29 observations deleted due to missingness)
Multiple R-squared:  0.2901,	Adjusted R-squared:  0.2417
F-statistic: 5.993 on 3 and 44 DF,  p-value: 0.001624

>
>
>
>
>
> ### LAD NT ###
> mProb.AB_LADNT.GJ = lm(TotalProb ~
+                           Group.ASDref*aprime.log
+                 #+ age
+                       #+ vabs_soc_ss
+                         , data = dLADNT)
> summary(mProb.AB_LADNT.GJ)

Call:
lm(formula = TotalProb ~ Group.ASDref * aprime.log, data = dLADNT)

Residuals:
    Min      1Q  Median      3Q     Max
-18.330  -8.950  -0.721   9.589  21.050

Coefficients:
                        Estimate Std. Error t value Pr(>|t|)
(Intercept)               44.531      2.676  16.639   <2e-16 ***
Group.ASDref               1.934      5.352   0.361    0.719
aprime.log                -1.655     17.990  -0.092    0.927
Group.ASDref:aprime.log   11.644     35.979   0.324    0.748
---
Signif. codes:  0 ‘***’ 0.001 ‘**’ 0.01 ‘*’ 0.05 ‘.’ 0.1 ‘ ’ 1

Residual standard error: 10.54 on 49 degrees of freedom
  (17 observations deleted due to missingness)
Multiple R-squared:  0.003251,	Adjusted R-squared:  -0.05777
F-statistic: 0.05327 on 3 and 49 DF,  p-value: 0.9836

>
> mProb.AB_LADNT.Words = lm(TotalProb ~
+                           Group.ASDref*FirstWords.months
+                 #+ age
+                       #+ vabs_soc_ss
+                         , data = dLADNT)
> summary(mProb.AB_LADNT.Words)

Call:
lm(formula = TotalProb ~ Group.ASDref * FirstWords.months, data = dLADNT)

Residuals:
     Min       1Q   Median       3Q      Max
-17.7962  -9.9435  -0.1043   9.3154  20.2760

Coefficients:
                               Estimate Std. Error t value Pr(>|t|)
(Intercept)                     46.7208     4.3736  10.682 8.39e-14 ***
Group.ASDref                     6.6810     8.7473   0.764    0.449
FirstWords.months               -0.1938     0.2786  -0.696    0.490
Group.ASDref:FirstWords.months  -0.4516     0.5573  -0.810    0.422
---
Signif. codes:  0 ‘***’ 0.001 ‘**’ 0.01 ‘*’ 0.05 ‘.’ 0.1 ‘ ’ 1

Residual standard error: 10.74 on 44 degrees of freedom
  (22 observations deleted due to missingness)
Multiple R-squared:  0.01613,	Adjusted R-squared:  -0.05095
F-statistic: 0.2404 on 3 and 44 DF,  p-value: 0.8677

>
> mProb.AB_LADNT.Phrases = lm(TotalProb ~
+                           Group.ASDref*FirstPhrases.months
+                 #+ age
+                       #+ vabs_soc_ss
+                         , data = dLADNT)
> summary(mProb.AB_LADNT.Phrases)

Call:
lm(formula = TotalProb ~ Group.ASDref * FirstPhrases.months,
    data = dLADNT)

Residuals:
     Min       1Q   Median       3Q      Max
-18.0709  -7.9529  -0.0306   8.6199  18.7627

Coefficients:
                                 Estimate Std. Error t value Pr(>|t|)
(Intercept)                      45.16962    3.98826  11.326 9.14e-15 ***
Group.ASDref                      7.97438    7.97652   1.000    0.323
FirstPhrases.months              -0.04662    0.16968  -0.275    0.785
Group.ASDref:FirstPhrases.months -0.33395    0.33935  -0.984    0.330
---
Signif. codes:  0 ‘***’ 0.001 ‘**’ 0.01 ‘*’ 0.05 ‘.’ 0.1 ‘ ’ 1

Residual standard error: 10.48 on 45 degrees of freedom
  (21 observations deleted due to missingness)
Multiple R-squared:  0.02535,	Adjusted R-squared:  -0.03963
F-statistic: 0.3902 on 3 and 45 DF,  p-value: 0.7606

>
>
>
>
>
> ###Follow up
> #aprime.log
> mProb.AB_ASD.GJ = lm(TotalProb ~ aprime.log
+                 #+ age
+                       #+ vabs_soc_ss
+                         , data = dASD)
> summary(mProb.AB_ASD.GJ)

Call:
lm(formula = TotalProb ~ aprime.log, data = dASD)

Residuals:
    Min      1Q  Median      3Q     Max
-32.045  -5.694   2.091   6.361  17.192

Coefficients:
            Estimate Std. Error t value Pr(>|t|)
(Intercept)   58.583      4.633  12.646 4.19e-12 ***
aprime.log    -2.192     22.774  -0.096    0.924
---
Signif. codes:  0 ‘***’ 0.001 ‘**’ 0.01 ‘*’ 0.05 ‘.’ 0.1 ‘ ’ 1

Residual standard error: 12.5 on 24 degrees of freedom
  (13 observations deleted due to missingness)
Multiple R-squared:  0.0003859,	Adjusted R-squared:  -0.04126
F-statistic: 0.009265 on 1 and 24 DF,  p-value: 0.9241

>
> mProb.AB_NT.GJ = lm(TotalProb ~ aprime.log
+                 #+ age
+                       #+ vabs_soc_ss
+                         , data = dNT)
> summary(mProb.AB_NT.GJ)

Call:
lm(formula = TotalProb ~ aprime.log, data = dNT)

Residuals:
    Min      1Q  Median      3Q     Max
-17.278  -7.247  -1.263   8.362  14.944

Coefficients:
            Estimate Std. Error t value Pr(>|t|)
(Intercept)   45.498      3.743  12.154 5.46e-12 ***
aprime.log     4.167     25.353   0.164    0.871
---
Signif. codes:  0 ‘***’ 0.001 ‘**’ 0.01 ‘*’ 0.05 ‘.’ 0.1 ‘ ’ 1

Residual standard error: 9.605 on 25 degrees of freedom
  (11 observations deleted due to missingness)
Multiple R-squared:  0.001079,	Adjusted R-squared:  -0.03888
F-statistic: 0.02701 on 1 and 25 DF,  p-value: 0.8708

>
> mProb.AB_LAD.GJ = lm(TotalProb ~ aprime.log
+                 #+ age
+                       #+ vabs_soc_ss
+                         , data = dLAD)
> summary(mProb.AB_LAD.GJ)

Call:
lm(formula = TotalProb ~ aprime.log, data = dLAD)

Residuals:
    Min      1Q  Median      3Q     Max
-18.330  -9.880   0.961   9.912  21.050

Coefficients:
            Estimate Std. Error t value Pr(>|t|)
(Intercept)   43.564      3.722  11.703  2.1e-11 ***
aprime.log    -7.477     24.749  -0.302    0.765
---
Signif. codes:  0 ‘***’ 0.001 ‘**’ 0.01 ‘*’ 0.05 ‘.’ 0.1 ‘ ’ 1

Residual standard error: 11.43 on 24 degrees of freedom
  (6 observations deleted due to missingness)
Multiple R-squared:  0.003789,	Adjusted R-squared:  -0.03772
F-statistic: 0.09128 on 1 and 24 DF,  p-value: 0.7652

>
> #Words
> mProb.AB_ASD.Words = lm(TotalProb ~ FirstWords.months
+                 #+ age
+                       #+ vabs_soc_ss
+                         , data = dASD)
> summary(mProb.AB_ASD.Words)

Call:
lm(formula = TotalProb ~ FirstWords.months, data = dASD)

Residuals:
    Min      1Q  Median      3Q     Max
-30.249  -4.939   1.291   6.701  20.031

Coefficients:
                  Estimate Std. Error t value Pr(>|t|)
(Intercept)        53.4090     5.3787   9.930 3.56e-09 ***
FirstWords.months   0.3200     0.2451   1.305    0.207
---
Signif. codes:  0 ‘***’ 0.001 ‘**’ 0.01 ‘*’ 0.05 ‘.’ 0.1 ‘ ’ 1

Residual standard error: 11.96 on 20 degrees of freedom
  (17 observations deleted due to missingness)
Multiple R-squared:  0.07851,	Adjusted R-squared:  0.03244
F-statistic: 1.704 on 1 and 20 DF,  p-value: 0.2066

>
> mProb.AB_NT.Words = lm(TotalProb ~ FirstWords.months
+                 #+ age
+                       #+ vabs_soc_ss
+                         , data = dNT)
> summary(mProb.AB_NT.Words)

Call:
lm(formula = TotalProb ~ FirstWords.months, data = dNT)

Residuals:
     Min       1Q   Median       3Q      Max
-15.5084  -7.0260  -0.5084   6.5544  16.5896

Coefficients:
                  Estimate Std. Error t value Pr(>|t|)
(Intercept)        50.0613     6.2703   7.984 4.44e-08 ***
FirstWords.months  -0.4196     0.4617  -0.909    0.373
---
Signif. codes:  0 ‘***’ 0.001 ‘**’ 0.01 ‘*’ 0.05 ‘.’ 0.1 ‘ ’ 1

Residual standard error: 9.661 on 23 degrees of freedom
  (13 observations deleted due to missingness)
Multiple R-squared:  0.03467,	Adjusted R-squared:  -0.007305
F-statistic: 0.826 on 1 and 23 DF,  p-value: 0.3729

>
> mProb.AB_LAD.Words = lm(TotalProb ~ FirstWords.months
+                 #+ age
+                       #+ vabs_soc_ss
+                         , data = dLAD)
> summary(mProb.AB_LAD.Words)

Call:
lm(formula = TotalProb ~ FirstWords.months, data = dLAD)

Residuals:
     Min       1Q   Median       3Q      Max
-17.7962 -10.7761   0.2997  10.2518  20.2760

Coefficients:
                  Estimate Std. Error t value Pr(>|t|)
(Intercept)       43.38032    5.80773   7.469 2.43e-07 ***
FirstWords.months  0.03199    0.23810   0.134    0.894
---
Signif. codes:  0 ‘***’ 0.001 ‘**’ 0.01 ‘*’ 0.05 ‘.’ 0.1 ‘ ’ 1

Residual standard error: 11.82 on 21 degrees of freedom
  (9 observations deleted due to missingness)
Multiple R-squared:  0.000859,	Adjusted R-squared:  -0.04672
F-statistic: 0.01805 on 1 and 21 DF,  p-value: 0.8944

>
> #Phrases
> mProb.AB_ASD.Phrases = lm(TotalProb ~ FirstPhrases.months
+                 #+ age
+                       #+ vabs_soc_ss
+                         , data = dASD)
> summary(mProb.AB_ASD.Phrases)

Call:
lm(formula = TotalProb ~ FirstPhrases.months, data = dASD)

Residuals:
    Min      1Q  Median      3Q     Max
-30.925  -4.675   2.059   6.991  18.075

Coefficients:
                    Estimate Std. Error t value Pr(>|t|)
(Intercept)         55.89108    8.40163   6.652 1.78e-06 ***
FirstPhrases.months  0.08473    0.24923   0.340    0.737
---
Signif. codes:  0 ‘***’ 0.001 ‘**’ 0.01 ‘*’ 0.05 ‘.’ 0.1 ‘ ’ 1

Residual standard error: 12.87 on 20 degrees of freedom
  (17 observations deleted due to missingness)
Multiple R-squared:  0.005746,	Adjusted R-squared:  -0.04397
F-statistic: 0.1156 on 1 and 20 DF,  p-value: 0.7374

>
> mProb.AB_NT.Phrases = lm(TotalProb ~ FirstPhrases.months
+                 #+ age
+                       #+ vabs_soc_ss
+                         , data = dNT)
> summary(mProb.AB_NT.Phrases)

Call:
lm(formula = TotalProb ~ FirstPhrases.months, data = dNT)

Residuals:
     Min       1Q   Median       3Q      Max
-17.0306  -7.4020  -0.5306   8.3480  14.9694

Coefficients:
                    Estimate Std. Error t value Pr(>|t|)
(Intercept)          49.1568     5.8320   8.429 1.24e-08 ***
FirstPhrases.months  -0.2136     0.2833  -0.754    0.458
---
Signif. codes:  0 ‘***’ 0.001 ‘**’ 0.01 ‘*’ 0.05 ‘.’ 0.1 ‘ ’ 1

Residual standard error: 9.692 on 24 degrees of freedom
  (12 observations deleted due to missingness)
Multiple R-squared:  0.02314,	Adjusted R-squared:  -0.01757
F-statistic: 0.5684 on 1 and 24 DF,  p-value: 0.4582

>
> mProb.AB_LAD.Phrases = lm(TotalProb ~ FirstPhrases.months
+                 #+ age
+                       #+ vabs_soc_ss
+                         , data = dLAD)
> summary(mProb.AB_LAD.Phrases)

Call:
lm(formula = TotalProb ~ FirstPhrases.months, data = dLAD)

Residuals:
     Min       1Q   Median       3Q      Max
-18.0709 -10.6081   0.9291   8.3919  18.7627

Coefficients:
                    Estimate Std. Error t value Pr(>|t|)
(Intercept)          41.1824     5.2733   7.810 1.21e-07 ***
FirstPhrases.months   0.1204     0.1577   0.763    0.454
---
Signif. codes:  0 ‘***’ 0.001 ‘**’ 0.01 ‘*’ 0.05 ‘.’ 0.1 ‘ ’ 1

Residual standard error: 11.31 on 21 degrees of freedom
  (9 observations deleted due to missingness)
Multiple R-squared:  0.027,	Adjusted R-squared:  -0.01933
F-statistic: 0.5827 on 1 and 21 DF,  p-value: 0.4537

> ###################
> ##### AnxDep ######
> ###################
> #combined
>
> ### ASD LAD ###
> mAnxDep.AB_ASDLAD.GJ = lm(AnxDep ~
+                           Group.NTref*aprime.log
+                  #+ age
+                       #+ vabs_soc_ss
+                         , data = dASDLAD)
> summary(mAnxDep.AB_ASDLAD.GJ)

Call:
lm(formula = AnxDep ~ Group.NTref * aprime.log, data = dASDLAD)

Residuals:
    Min      1Q  Median      3Q     Max
-16.302  -4.229  -2.473   6.276  17.775

Coefficients:
                       Estimate Std. Error t value Pr(>|t|)
(Intercept)              58.800      2.223  26.452   <2e-16 ***
Group.NTref              -8.770      4.446  -1.973   0.0544 .
aprime.log               -5.659     12.922  -0.438   0.6634
Group.NTref:aprime.log   18.239     25.844   0.706   0.4839
---
Signif. codes:  0 ‘***’ 0.001 ‘**’ 0.01 ‘*’ 0.05 ‘.’ 0.1 ‘ ’ 1

Residual standard error: 8.948 on 47 degrees of freedom
  (20 observations deleted due to missingness)
Multiple R-squared:  0.3222,	Adjusted R-squared:  0.279
F-statistic: 7.448 on 3 and 47 DF,  p-value: 0.0003525

>
> mAnxDep.AB_ASDLAD.Words = lm(AnxDep ~
+                           Group.NTref*FirstWords.months
+                  #+ age
+                       #+ vabs_soc_ss
+                         , data = dASDLAD)
> summary(mAnxDep.AB_ASDLAD.Words)

Call:
lm(formula = AnxDep ~ Group.NTref * FirstWords.months, data = dASDLAD)

Residuals:
    Min      1Q  Median      3Q     Max
-16.202  -4.164  -2.491   4.051  17.428

Coefficients:
                              Estimate Std. Error t value Pr(>|t|)
(Intercept)                   56.39399    2.97506  18.956   <2e-16 ***
Group.NTref                   -9.84947    5.95012  -1.655    0.105
FirstWords.months              0.13704    0.12836   1.068    0.292
Group.NTref:FirstWords.months -0.08822    0.25672  -0.344    0.733
---
Signif. codes:  0 ‘***’ 0.001 ‘**’ 0.01 ‘*’ 0.05 ‘.’ 0.1 ‘ ’ 1

Residual standard error: 8.933 on 41 degrees of freedom
  (26 observations deleted due to missingness)
Multiple R-squared:  0.3193,	Adjusted R-squared:  0.2694
F-statistic: 6.409 on 3 and 41 DF,  p-value: 0.00116

>
> mAnxDep.AB_ASDLAD.Phrases = lm(AnxDep ~
+                           Group.NTref*FirstPhrases.months
+                  #+ age
+                       #+ vabs_soc_ss
+                         , data = dASDLAD)
> summary(mAnxDep.AB_ASDLAD.Phrases)

Call:
lm(formula = AnxDep ~ Group.NTref * FirstPhrases.months, data = dASDLAD)

Residuals:
    Min      1Q  Median      3Q     Max
-15.329  -3.797  -2.658   3.918  19.326

Coefficients:
                                Estimate Std. Error t value Pr(>|t|)
(Intercept)                     57.52407    3.70183  15.539   <2e-16 ***
Group.NTref                     -6.62219    7.40367  -0.894    0.376
FirstPhrases.months              0.03526    0.11011   0.320    0.750
Group.NTref:FirstPhrases.months -0.11673    0.22021  -0.530    0.599
---
Signif. codes:  0 ‘***’ 0.001 ‘**’ 0.01 ‘*’ 0.05 ‘.’ 0.1 ‘ ’ 1

Residual standard error: 9.227 on 41 degrees of freedom
  (26 observations deleted due to missingness)
Multiple R-squared:  0.2587,	Adjusted R-squared:  0.2044
F-statistic: 4.769 on 3 and 41 DF,  p-value: 0.006082

>
>
>
>
>
> ### ASD NT ###
> mAnxDep.AB_ASDNT.GJ = lm(AnxDep ~
+                           Group.LADref*aprime.log
+                 #+ age
+                       #+ vabs_soc_ss
+                         , data = dASDNT)
> summary(mAnxDep.AB_ASDNT.GJ)

Call:
lm(formula = AnxDep ~ Group.LADref * aprime.log, data = dASDNT)

Residuals:
    Min      1Q  Median      3Q     Max
-16.302  -3.942  -1.785   6.247  22.058

Coefficients:
                        Estimate Std. Error t value Pr(>|t|)
(Intercept)             59.35460    2.41101  24.618   <2e-16 ***
Group.LADref            -7.66154    4.82202  -1.589    0.119
aprime.log               0.06431   14.37930   0.004    0.996
Group.LADref:aprime.log 29.68621   28.75860   1.032    0.307
---
Signif. codes:  0 ‘***’ 0.001 ‘**’ 0.01 ‘*’ 0.05 ‘.’ 0.1 ‘ ’ 1

Residual standard error: 8.924 on 49 degrees of freedom
  (24 observations deleted due to missingness)
Multiple R-squared:  0.3407,	Adjusted R-squared:  0.3004
F-statistic: 8.441 on 3 and 49 DF,  p-value: 0.0001267

>
> mAnxDep.AB_ASDNT.Words = lm(AnxDep ~
+                           Group.LADref*FirstWords.months
+                 #+ age
+                       #+ vabs_soc_ss
+                         , data = dASDNT)
> summary(mAnxDep.AB_ASDNT.Words)

Call:
lm(formula = AnxDep ~ Group.LADref * FirstWords.months, data = dASDNT)

Residuals:
    Min      1Q  Median      3Q     Max
-16.202  -4.098  -2.124   5.152  20.258

Coefficients:
                               Estimate Std. Error t value Pr(>|t|)
(Intercept)                     59.6819     3.5198  16.956   <2e-16 ***
Group.LADref                    -3.2736     7.0397  -0.465    0.644
FirstWords.months               -0.0739     0.2285  -0.323    0.748
Group.LADref:FirstWords.months  -0.5101     0.4570  -1.116    0.270
---
Signif. codes:  0 ‘***’ 0.001 ‘**’ 0.01 ‘*’ 0.05 ‘.’ 0.1 ‘ ’ 1

Residual standard error: 8.995 on 44 degrees of freedom
  (29 observations deleted due to missingness)
Multiple R-squared:  0.3091,	Adjusted R-squared:  0.262
F-statistic: 6.563 on 3 and 44 DF,  p-value: 0.0009169

>
> mAnxDep.AB_ASDNT.Phrases = lm(AnxDep ~
+                           Group.LADref*FirstPhrases.months
+                 #+ age
+                       #+ vabs_soc_ss
+                         , data = dASDNT)
> summary(mAnxDep.AB_ASDNT.Phrases)

Call:
lm(formula = AnxDep ~ Group.LADref * FirstPhrases.months, data = dASDNT)

Residuals:
    Min      1Q  Median      3Q     Max
-15.329  -4.351  -2.067   3.918  20.227

Coefficients:
                                 Estimate Std. Error t value Pr(>|t|)
(Intercept)                      60.00956    4.02673  14.903   <2e-16 ***
Group.LADref                     -1.65120    8.05345  -0.205    0.838
FirstPhrases.months              -0.09534    0.15930  -0.598    0.553
Group.LADref:FirstPhrases.months -0.37791    0.31860  -1.186    0.242
---
Signif. codes:  0 ‘***’ 0.001 ‘**’ 0.01 ‘*’ 0.05 ‘.’ 0.1 ‘ ’ 1

Residual standard error: 9.091 on 45 degrees of freedom
  (28 observations deleted due to missingness)
Multiple R-squared:  0.2688,	Adjusted R-squared:  0.2201
F-statistic: 5.515 on 3 and 45 DF,  p-value: 0.002595

>
>
>
>
>
> ### LAD NT ###
> mAnxDep.AB_LADNT.GJ = lm(AnxDep ~
+                           Group.ASDref*aprime.log
+                 #+ age
+                       #+ vabs_soc_ss
+                         , data = dLADNT)
> summary(mAnxDep.AB_LADNT.GJ)

Call:
lm(formula = AnxDep ~ Group.ASDref * aprime.log, data = dLADNT)

Residuals:
   Min     1Q Median     3Q    Max
-5.524 -3.927 -2.304  2.732 22.058

Coefficients:
                        Estimate Std. Error t value Pr(>|t|)
(Intercept)               54.970      1.463  37.563   <2e-16 ***
Group.ASDref               1.108      2.927   0.379    0.707
aprime.log                 9.184      9.751   0.942    0.351
Group.ASDref:aprime.log   11.448     19.501   0.587    0.560
---
Signif. codes:  0 ‘***’ 0.001 ‘**’ 0.01 ‘*’ 0.05 ‘.’ 0.1 ‘ ’ 1

Residual standard error: 5.771 on 50 degrees of freedom
  (16 observations deleted due to missingness)
Multiple R-squared:  0.02233,	Adjusted R-squared:  -0.03632
F-statistic: 0.3808 on 3 and 50 DF,  p-value: 0.7673

>
> mAnxDep.AB_LADNT.Words = lm(AnxDep ~
+                           Group.ASDref*FirstWords.months
+                 #+ age
+                       #+ vabs_soc_ss
+                         , data = dLADNT)
> summary(mAnxDep.AB_LADNT.Words)

Call:
lm(formula = AnxDep ~ Group.ASDref * FirstWords.months, data = dLADNT)

Residuals:
   Min     1Q Median     3Q    Max
-5.930 -3.440 -2.584  2.743 20.258

Coefficients:
                               Estimate Std. Error t value Pr(>|t|)
(Intercept)                     54.7572     2.3004  23.803   <2e-16 ***
Group.ASDref                     6.5759     4.6008   1.429    0.160
FirstWords.months               -0.1180     0.1444  -0.817    0.418
Group.ASDref:FirstWords.months  -0.4219     0.2888  -1.461    0.151
---
Signif. codes:  0 ‘***’ 0.001 ‘**’ 0.01 ‘*’ 0.05 ‘.’ 0.1 ‘ ’ 1

Residual standard error: 5.699 on 45 degrees of freedom
  (21 observations deleted due to missingness)
Multiple R-squared:  0.04687,	Adjusted R-squared:  -0.01667
F-statistic: 0.7377 on 3 and 45 DF,  p-value: 0.5351

>
> mAnxDep.AB_LADNT.Phrases = lm(AnxDep ~
+                           Group.ASDref*FirstPhrases.months
+                 #+ age
+                       #+ vabs_soc_ss
+                         , data = dLADNT)
> summary(mAnxDep.AB_LADNT.Phrases)

Call:
lm(formula = AnxDep ~ Group.ASDref * FirstPhrases.months, data = dLADNT)

Residuals:
   Min     1Q Median     3Q    Max
-5.772 -3.658 -2.361  2.227 20.227

Coefficients:
                                 Estimate Std. Error t value Pr(>|t|)
(Intercept)                      56.69847    2.12105  26.731   <2e-16 ***
Group.ASDref                      4.97099    4.24211   1.172   0.2473
FirstPhrases.months              -0.15370    0.09047  -1.699   0.0961 .
Group.ASDref:FirstPhrases.months -0.26118    0.18094  -1.443   0.1557
---
Signif. codes:  0 ‘***’ 0.001 ‘**’ 0.01 ‘*’ 0.05 ‘.’ 0.1 ‘ ’ 1

Residual standard error: 5.592 on 46 degrees of freedom
  (20 observations deleted due to missingness)
Multiple R-squared:  0.06365,	Adjusted R-squared:  0.002588
F-statistic: 1.042 on 3 and 46 DF,  p-value: 0.3828

>
>
>
>
> ###Follow up
> #aprime.log
> mAnxDep.AB_ASD.GJ = lm(AnxDep ~ aprime.log
+                 #+ age
+                       #+ vabs_soc_ss
+                         , data = dASD)
> summary(mAnxDep.AB_ASD.GJ)

Call:
lm(formula = AnxDep ~ aprime.log, data = dASD)

Residuals:
     Min       1Q   Median       3Q      Max
-16.3019  -9.1854  -0.3019   9.2469  17.7749

Coefficients:
            Estimate Std. Error t value Pr(>|t|)
(Intercept)   63.185      4.286  14.743 3.28e-13 ***
aprime.log   -14.779     21.834  -0.677    0.505
---
Signif. codes:  0 ‘***’ 0.001 ‘**’ 0.01 ‘*’ 0.05 ‘.’ 0.1 ‘ ’ 1

Residual standard error: 11.42 on 23 degrees of freedom
  (14 observations deleted due to missingness)
Multiple R-squared:  0.01953,	Adjusted R-squared:  -0.0231
F-statistic: 0.4582 on 1 and 23 DF,  p-value: 0.5052

>
> mAnxDep.AB_NT.GJ = lm(AnxDep ~ aprime.log
+                 #+ age
+                       #+ vabs_soc_ss
+                         , data = dNT)
> summary(mAnxDep.AB_NT.GJ)

Call:
lm(formula = AnxDep ~ aprime.log, data = dNT)

Residuals:
    Min      1Q  Median      3Q     Max
-5.5238 -3.3335 -1.9696  0.3281 22.0576

Coefficients:
            Estimate Std. Error t value Pr(>|t|)
(Intercept)    55.52       2.29  24.247   <2e-16 ***
aprime.log     14.91      15.28   0.976    0.338
---
Signif. codes:  0 ‘***’ 0.001 ‘**’ 0.01 ‘*’ 0.05 ‘.’ 0.1 ‘ ’ 1

Residual standard error: 5.89 on 26 degrees of freedom
  (10 observations deleted due to missingness)
Multiple R-squared:  0.03531,	Adjusted R-squared:  -0.001789
F-statistic: 0.9518 on 1 and 26 DF,  p-value: 0.3383

>
> mAnxDep.AB_LAD.GJ = lm(AnxDep ~ aprime.log
+                 #+ age
+                       #+ vabs_soc_ss
+                         , data = dLAD)
> summary(mAnxDep.AB_LAD.GJ)

Call:
lm(formula = AnxDep ~ aprime.log, data = dLAD)

Residuals:
   Min     1Q Median     3Q    Max
-4.415 -4.048 -3.244  3.672 13.763

Coefficients:
            Estimate Std. Error t value Pr(>|t|)
(Intercept)   54.415      1.836  29.630   <2e-16 ***
aprime.log     3.460     12.210   0.283    0.779
---
Signif. codes:  0 ‘***’ 0.001 ‘**’ 0.01 ‘*’ 0.05 ‘.’ 0.1 ‘ ’ 1

Residual standard error: 5.64 on 24 degrees of freedom
  (6 observations deleted due to missingness)
Multiple R-squared:  0.003334,	Adjusted R-squared:  -0.03819
F-statistic: 0.08029 on 1 and 24 DF,  p-value: 0.7793

>
> #Words
> mAnxDep.AB_ASD.Words = lm(AnxDep ~ FirstWords.months
+                 #+ age
+                       #+ vabs_soc_ss
+                         , data = dASD)
> summary(mAnxDep.AB_ASD.Words)

Call:
lm(formula = AnxDep ~ FirstWords.months, data = dASD)

Residuals:
    Min      1Q  Median      3Q     Max
-16.202 -10.308  -0.036   9.143  17.428

Coefficients:
                  Estimate Std. Error t value Pr(>|t|)
(Intercept)        61.3187     5.2104  11.769 1.92e-10 ***
FirstWords.months   0.1811     0.2375   0.763    0.454
---
Signif. codes:  0 ‘***’ 0.001 ‘**’ 0.01 ‘*’ 0.05 ‘.’ 0.1 ‘ ’ 1

Residual standard error: 11.59 on 20 degrees of freedom
  (17 observations deleted due to missingness)
Multiple R-squared:  0.02827,	Adjusted R-squared:  -0.02031
F-statistic: 0.5819 on 1 and 20 DF,  p-value: 0.4545

>
> mAnxDep.AB_NT.Words = lm(AnxDep ~ FirstWords.months
+                 #+ age
+                       #+ vabs_soc_ss
+                         , data = dNT)
> summary(mAnxDep.AB_NT.Words)

Call:
lm(formula = AnxDep ~ FirstWords.months, data = dNT)

Residuals:
   Min     1Q Median     3Q    Max
-5.413 -3.687 -2.275  2.119 20.258

Coefficients:
                  Estimate Std. Error t value Pr(>|t|)
(Intercept)        58.0451     3.8676  15.008 1.07e-13 ***
FirstWords.months  -0.3290     0.2807  -1.172    0.253
---
Signif. codes:  0 ‘***’ 0.001 ‘**’ 0.01 ‘*’ 0.05 ‘.’ 0.1 ‘ ’ 1

Residual standard error: 6.038 on 24 degrees of freedom
  (12 observations deleted due to missingness)
Multiple R-squared:  0.05412,	Adjusted R-squared:  0.01471
F-statistic: 1.373 on 1 and 24 DF,  p-value: 0.2528

>
> mAnxDep.AB_LAD.Words = lm(AnxDep ~ FirstWords.months
+                 #+ age
+                       #+ vabs_soc_ss
+                         , data = dLAD)
> summary(mAnxDep.AB_LAD.Words)

Call:
lm(formula = AnxDep ~ FirstWords.months, data = dLAD)

Residuals:
   Min     1Q Median     3Q    Max
-5.930 -3.142 -2.584  2.289 12.628

Coefficients:
                  Estimate Std. Error t value Pr(>|t|)
(Intercept)       51.46926    2.59686  19.820 4.49e-15 ***
FirstWords.months  0.09293    0.10646   0.873    0.393
---
Signif. codes:  0 ‘***’ 0.001 ‘**’ 0.01 ‘*’ 0.05 ‘.’ 0.1 ‘ ’ 1

Residual standard error: 5.285 on 21 degrees of freedom
  (9 observations deleted due to missingness)
Multiple R-squared:  0.03501,	Adjusted R-squared:  -0.01094
F-statistic: 0.7619 on 1 and 21 DF,  p-value: 0.3926

>
> #Phrases
> mAnxDep.AB_ASD.Phrases = lm(AnxDep ~ FirstPhrases.months
+                 #+ age
+                       #+ vabs_soc_ss
+                         , data = dASD)
> summary(mAnxDep.AB_ASD.Phrases)

Call:
lm(formula = AnxDep ~ FirstPhrases.months, data = dASD)

Residuals:
     Min       1Q   Median       3Q      Max
-15.3288 -10.5820   0.5754   9.1531  19.3265

Coefficients:
                    Estimate Std. Error t value Pr(>|t|)
(Intercept)         60.83516    7.84475   7.755 1.88e-07 ***
FirstPhrases.months  0.09362    0.23271   0.402    0.692
---
Signif. codes:  0 ‘***’ 0.001 ‘**’ 0.01 ‘*’ 0.05 ‘.’ 0.1 ‘ ’ 1

Residual standard error: 12.01 on 20 degrees of freedom
  (17 observations deleted due to missingness)
Multiple R-squared:  0.008027,	Adjusted R-squared:  -0.04157
F-statistic: 0.1618 on 1 and 20 DF,  p-value: 0.6917

>
> mAnxDep.AB_NT.Phrases = lm(AnxDep ~ FirstPhrases.months
+                 #+ age
+                       #+ vabs_soc_ss
+                         , data = dNT)
> summary(mAnxDep.AB_NT.Phrases)

Call:
lm(formula = AnxDep ~ FirstPhrases.months, data = dNT)

Residuals:
   Min     1Q Median     3Q    Max
-5.772 -3.635 -2.361  1.050 20.227

Coefficients:
                    Estimate Std. Error t value Pr(>|t|)
(Intercept)          59.1840     3.4556  17.127 2.53e-15 ***
FirstPhrases.months  -0.2843     0.1686  -1.686    0.104
---
Signif. codes:  0 ‘***’ 0.001 ‘**’ 0.01 ‘*’ 0.05 ‘.’ 0.1 ‘ ’ 1

Residual standard error: 5.773 on 25 degrees of freedom
  (11 observations deleted due to missingness)
Multiple R-squared:  0.1021,	Adjusted R-squared:  0.0662
F-statistic: 2.843 on 1 and 25 DF,  p-value: 0.1042

>
> mAnxDep.AB_LAD.Phrases = lm(AnxDep ~ FirstPhrases.months
+                 #+ age
+                       #+ vabs_soc_ss
+                         , data = dLAD)
> summary(mAnxDep.AB_LAD.Phrases)

Call:
lm(formula = AnxDep ~ FirstPhrases.months, data = dLAD)

Residuals:
   Min     1Q Median     3Q    Max
-3.889 -3.658 -2.797  2.980 14.758

Coefficients:
                    Estimate Std. Error t value Pr(>|t|)
(Intercept)         54.21297    2.50203  21.668 7.52e-16 ***
FirstPhrases.months -0.02311    0.07481  -0.309     0.76
---
Signif. codes:  0 ‘***’ 0.001 ‘**’ 0.01 ‘*’ 0.05 ‘.’ 0.1 ‘ ’ 1

Residual standard error: 5.367 on 21 degrees of freedom
  (9 observations deleted due to missingness)
Multiple R-squared:  0.004523,	Adjusted R-squared:  -0.04288
F-statistic: 0.09542 on 1 and 21 DF,  p-value: 0.7604


> ### RQ 2 B: Educational Outcome ~ Group*Core Language (youth cohort)/Grammaticality Judgement (A’; all participants)/age of first words/phrases
> ###############################
> ##### Full time anything ######
> ###############################
> # dASD.older18
> # dASDLAD.older18
>
>
> ### ASD LAD ###
> mFullTime_ASDLAD.GJ = glm(NotvsFullTime ~
+                           Group.NTref*aprime.log
+                  #+ age
+                       #+ vabs_soc_ss
+                         , family = binomial, data = dASDLAD.older18)
> summary(mFullTime_ASDLAD.GJ)

Call:
glm(formula = NotvsFullTime ~ Group.NTref * aprime.log, family = binomial,
    data = dASDLAD.older18)

Deviance Residuals:
    Min       1Q   Median       3Q      Max
-2.2607   0.4000   0.4090   0.4211   1.2944

Coefficients:
                       Estimate Std. Error z value Pr(>|z|)
(Intercept)              2.0876     0.9028   2.312   0.0208 *
Group.NTref              0.5811     1.8056   0.322   0.7476
aprime.log               3.1835     5.6574   0.563   0.5736
Group.NTref:aprime.log  -7.5316    11.3149  -0.666   0.5056
---
Signif. codes:  0 ‘***’ 0.001 ‘**’ 0.01 ‘*’ 0.05 ‘.’ 0.1 ‘ ’ 1

(Dispersion parameter for binomial family taken to be 1)

    Null deviance: 40.032  on 39  degrees of freedom
Residual deviance: 32.152  on 36  degrees of freedom
  (15 observations deleted due to missingness)
AIC: 40.152

Number of Fisher Scoring iterations: 5

>
> mFullTime_ASDLAD.Words = glm(NotvsFullTime ~
+                           Group.NTref*FirstWords.months
+                  #+ age
+                       #+ vabs_soc_ss
+                         , family = binomial, data = dASDLAD.older18)
> summary(mFullTime_ASDLAD.Words)

Call:
glm(formula = NotvsFullTime ~ Group.NTref * FirstWords.months,
    family = binomial, data = dASDLAD.older18)

Deviance Residuals:
    Min       1Q   Median       3Q      Max
-2.2573   0.3454   0.4475   0.5534   1.0614

Coefficients:
                               Estimate Std. Error z value Pr(>|z|)
(Intercept)                    0.968257   1.019009   0.950    0.342
Group.NTref                    1.702591   2.038017   0.835    0.403
FirstWords.months              0.025500   0.049219   0.518    0.604
Group.NTref:FirstWords.months -0.003096   0.098438  -0.031    0.975

(Dispersion parameter for binomial family taken to be 1)

    Null deviance: 35.467  on 35  degrees of freedom
Residual deviance: 31.339  on 32  degrees of freedom
  (19 observations deleted due to missingness)
AIC: 39.339

Number of Fisher Scoring iterations: 5

>
> mFullTime_ASDLAD.Phrases = glm(NotvsFullTime ~
+                           Group.NTref*FirstPhrases.months
+                  #+ age
+                       #+ vabs_soc_ss
+                        , family = binomial, data = dASDLAD.older18)
> summary(mFullTime_ASDLAD.Phrases)

Call:
glm(formula = NotvsFullTime ~ Group.NTref * FirstPhrases.months,
    family = binomial, data = dASDLAD.older18)

Deviance Residuals:
    Min       1Q   Median       3Q      Max
-2.6411   0.2418   0.3011   0.6262   1.0928

Coefficients:
                                 Estimate Std. Error z value Pr(>|z|)
(Intercept)                      1.897025   1.439185   1.318   0.1875
Group.NTref                      5.431453   2.878370   1.887   0.0592 .
FirstPhrases.months             -0.003755   0.041233  -0.091   0.9274
Group.NTref:FirstPhrases.months -0.120957   0.082465  -1.467   0.1424
---
Signif. codes:  0 ‘***’ 0.001 ‘**’ 0.01 ‘*’ 0.05 ‘.’ 0.1 ‘ ’ 1

(Dispersion parameter for binomial family taken to be 1)

    Null deviance: 32.440  on 35  degrees of freedom
Residual deviance: 26.499  on 32  degrees of freedom
  (19 observations deleted due to missingness)
AIC: 34.499

Number of Fisher Scoring iterations: 5

>
>
>
>
> ### ASD NT ###
> mFullTime_ASDNT.GJ = glm(NotvsFullTime ~
+                           Group.LADref*aprime.log
+                 #+ age
+                       #+ vabs_soc_ss
+                         , family = binomial, data = dASDNT.older18)
> summary(mFullTime_ASDNT.GJ)

Call:
glm(formula = NotvsFullTime ~ Group.LADref * aprime.log, family = binomial,
    data = dASDNT.older18)

Deviance Residuals:
    Min       1Q   Median       3Q      Max
-1.9478  -0.4298   0.5731   0.6454   1.2944

Coefficients:
                        Estimate Std. Error z value Pr(>|z|)
(Intercept)              1.82824    0.82934   2.204   0.0275 *
Group.LADref             0.06235    1.65867   0.038   0.9700
aprime.log               4.68375    4.56980   1.025   0.3054
Group.LADref:aprime.log -4.53104    9.13960  -0.496   0.6201
---
Signif. codes:  0 ‘***’ 0.001 ‘**’ 0.01 ‘*’ 0.05 ‘.’ 0.1 ‘ ’ 1

(Dispersion parameter for binomial family taken to be 1)

    Null deviance: 43.801  on 37  degrees of freedom
Residual deviance: 39.366  on 34  degrees of freedom
  (14 observations deleted due to missingness)
AIC: 47.366

Number of Fisher Scoring iterations: 4

>
> mFullTime_ASDNT.Words = glm(NotvsFullTime ~
+                           Group.LADref*FirstWords.months
+                 #+ age
+                       #+ vabs_soc_ss
+                         , family = binomial, data = dASDNT.older18)
> summary(mFullTime_ASDNT.Words)

Call:
glm(formula = NotvsFullTime ~ Group.LADref * FirstWords.months,
    family = binomial, data = dASDNT.older18)

Deviance Residuals:
    Min       1Q   Median       3Q      Max
-2.1166   0.3306   0.4745   0.8367   1.0614

Coefficients:
                               Estimate Std. Error z value Pr(>|z|)
(Intercept)                     2.07333    1.18039   1.756   0.0790 .
Group.LADref                    3.91275    2.36078   1.657   0.0974 .
FirstWords.months              -0.06574    0.06887  -0.955   0.3398
Group.LADref:FirstWords.months -0.18557    0.13773  -1.347   0.1779
---
Signif. codes:  0 ‘***’ 0.001 ‘**’ 0.01 ‘*’ 0.05 ‘.’ 0.1 ‘ ’ 1

(Dispersion parameter for binomial family taken to be 1)

    Null deviance: 37.100  on 33  degrees of freedom
Residual deviance: 33.269  on 30  degrees of freedom
  (18 observations deleted due to missingness)
AIC: 41.269

Number of Fisher Scoring iterations: 5

>
> mFullTime_ASDNT.Phrases = glm(NotvsFullTime ~
+                           Group.LADref*FirstPhrases.months
+                 #+ age
+                       #+ vabs_soc_ss
+                         , family = binomial, data = dASDNT.older18)
> summary(mFullTime_ASDNT.Phrases)

Call:
glm(formula = NotvsFullTime ~ Group.LADref * FirstPhrases.months,
    family = binomial, data = dASDNT.older18)

Deviance Residuals:
    Min       1Q   Median       3Q      Max
-1.9909   0.4962   0.5702   0.7022   1.0928

Coefficients:
                                  Estimate Std. Error z value Pr(>|z|)
(Intercept)                       0.830430   1.342015   0.619    0.536
Group.LADref                      3.298263   2.684030   1.229    0.219
FirstPhrases.months               0.003516   0.049781   0.071    0.944
Group.LADref:FirstPhrases.months -0.106415   0.099562  -1.069    0.285

(Dispersion parameter for binomial family taken to be 1)

    Null deviance: 38.139  on 35  degrees of freedom
Residual deviance: 36.417  on 32  degrees of freedom
  (16 observations deleted due to missingness)
AIC: 44.417

Number of Fisher Scoring iterations: 4

>
>
>
>
> ### LAD NT ###
> mFullTime_LADNT.GJ = glm(NotvsFullTime ~
+                           Group.ASDref*aprime.log
+                 #+ age
+                       #+ vabs_soc_ss
+                         , family = binomial, data = dLADNT.older18)
> summary(mFullTime_LADNT.GJ)

Call:
glm(formula = NotvsFullTime ~ Group.ASDref * aprime.log, family = binomial,
    data = dLADNT.older18)

Deviance Residuals:
    Min       1Q   Median       3Q      Max
-2.2607   0.4033   0.4151   0.5731   0.7413

Coefficients:
                        Estimate Std. Error z value Pr(>|z|)
(Intercept)               2.1188     0.8556   2.476   0.0133 *
Group.ASDref             -0.5188     1.7112  -0.303   0.7618
aprime.log                0.9180     6.2044   0.148   0.8824
Group.ASDref:aprime.log   3.0005    12.4087   0.242   0.8089
---
Signif. codes:  0 ‘***’ 0.001 ‘**’ 0.01 ‘*’ 0.05 ‘.’ 0.1 ‘ ’ 1

(Dispersion parameter for binomial family taken to be 1)

    Null deviance: 36.170  on 47  degrees of freedom
Residual deviance: 35.085  on 44  degrees of freedom
  (9 observations deleted due to missingness)
AIC: 43.085

Number of Fisher Scoring iterations: 5

>
> mFullTime_LADNT.Words = glm(NotvsFullTime ~
+                           Group.ASDref*FirstWords.months
+                 #+ age
+                       #+ vabs_soc_ss
+                         , family = binomial, data = dLADNT.older18)
> summary(mFullTime_LADNT.Words)

Call:
glm(formula = NotvsFullTime ~ Group.ASDref * FirstWords.months,
    family = binomial, data = dLADNT.older18)

Deviance Residuals:
    Min       1Q   Median       3Q      Max
-2.2573   0.3571   0.4475   0.4779   1.0194

Coefficients:
                               Estimate Std. Error z value Pr(>|z|)
(Intercept)                     2.92463    1.32634   2.205   0.0275 *
Group.ASDref                    2.21016    2.65268   0.833   0.4047
FirstWords.months              -0.06728    0.07340  -0.917   0.3593
Group.ASDref:FirstWords.months -0.18247    0.14680  -1.243   0.2139
---
Signif. codes:  0 ‘***’ 0.001 ‘**’ 0.01 ‘*’ 0.05 ‘.’ 0.1 ‘ ’ 1

(Dispersion parameter for binomial family taken to be 1)

    Null deviance: 30.662  on 41  degrees of freedom
Residual deviance: 28.533  on 38  degrees of freedom
  (15 observations deleted due to missingness)
AIC: 36.533

Number of Fisher Scoring iterations: 5

>
> mFullTime_LADNT.Phrases = glm(NotvsFullTime ~
+                           Group.ASDref*FirstPhrases.months
+                 #+ age
+                       #+ vabs_soc_ss
+                         , family = binomial, data = dLADNT.older18)
> summary(mFullTime_LADNT.Phrases)

Call:
glm(formula = NotvsFullTime ~ Group.ASDref * FirstPhrases.months,
    family = binomial, data = dLADNT.older18)

Deviance Residuals:
    Min       1Q   Median       3Q      Max
-2.6411   0.2743   0.4508   0.5769   0.9014

Coefficients:
                                 Estimate Std. Error z value Pr(>|z|)
(Intercept)                       3.54616    1.19033   2.979  0.00289 **
Group.ASDref                     -2.13319    2.38065  -0.896  0.37023
FirstPhrases.months              -0.05696    0.03968  -1.435  0.15116
Group.ASDref:FirstPhrases.months  0.01454    0.07937   0.183  0.85462
---
Signif. codes:  0 ‘***’ 0.001 ‘**’ 0.01 ‘*’ 0.05 ‘.’ 0.1 ‘ ’ 1

(Dispersion parameter for binomial family taken to be 1)

    Null deviance: 35.051  on 43  degrees of freedom
Residual deviance: 30.783  on 40  degrees of freedom
  (13 observations deleted due to missingness)
AIC: 38.783

Number of Fisher Scoring iterations: 5

>
>
>
> ###Follow up
> #aprime.log
> mFullTime_ASD.GJ = glm(NotvsFullTime ~ aprime.log
+                 #+ age
+                       #+ vabs_soc_ss
+                         , family = binomial, data = dASD.older18)
> summary(mFullTime_ASD.GJ)

Call:
glm(formula = NotvsFullTime ~ aprime.log, family = binomial,
    data = dASD.older18)

Deviance Residuals:
    Min       1Q   Median       3Q      Max
-1.6591  -0.9512   0.5539   1.0210   1.2944

Coefficients:
            Estimate Std. Error z value Pr(>|z|)
(Intercept)    1.797      1.242   1.447    0.148
aprime.log     6.949      5.366   1.295    0.195

(Dispersion parameter for binomial family taken to be 1)

    Null deviance: 20.190  on 14  degrees of freedom
Residual deviance: 18.217  on 13  degrees of freedom
  (10 observations deleted due to missingness)
AIC: 22.217

Number of Fisher Scoring iterations: 4

>
> mFullTime_NT.GJ = glm(NotvsFullTime ~ aprime.log
+                 #+ age
+                       #+ vabs_soc_ss
+                         , family = binomial, data = dNT.older18)
> summary(mFullTime_NT.GJ)

Call:
glm(formula = NotvsFullTime ~ aprime.log, family = binomial,
    data = dNT.older18)

Deviance Residuals:
    Min       1Q   Median       3Q      Max
-1.9478   0.5702   0.5731   0.6390   0.7413

Coefficients:
            Estimate Std. Error z value Pr(>|z|)
(Intercept)    1.859      1.100   1.691   0.0909 .
aprime.log     2.418      7.399   0.327   0.7438
---
Signif. codes:  0 ‘***’ 0.001 ‘**’ 0.01 ‘*’ 0.05 ‘.’ 0.1 ‘ ’ 1

(Dispersion parameter for binomial family taken to be 1)

    Null deviance: 21.254  on 22  degrees of freedom
Residual deviance: 21.150  on 21  degrees of freedom
  (4 observations deleted due to missingness)
AIC: 25.15

Number of Fisher Scoring iterations: 4

>
> mFullTime_LAD.GJ = glm(NotvsFullTime ~ aprime.log
+                 #+ age
+                       #+ vabs_soc_ss
+                         , family = binomial, data = dLAD.older18)
> summary(mFullTime_LAD.GJ)

Call:
glm(formula = NotvsFullTime ~ aprime.log, family = binomial,
    data = dLAD.older18)

Deviance Residuals:
    Min       1Q   Median       3Q      Max
-2.2607   0.4033   0.4088   0.4151   0.4211

Coefficients:
            Estimate Std. Error z value Pr(>|z|)
(Intercept)   2.3782     1.3110   1.814   0.0697 .
aprime.log   -0.5823     9.9617  -0.058   0.9534
---
Signif. codes:  0 ‘***’ 0.001 ‘**’ 0.01 ‘*’ 0.05 ‘.’ 0.1 ‘ ’ 1

(Dispersion parameter for binomial family taken to be 1)

    Null deviance: 13.938  on 24  degrees of freedom
Residual deviance: 13.935  on 23  degrees of freedom
  (5 observations deleted due to missingness)
AIC: 17.935

Number of Fisher Scoring iterations: 5

>
> #Words
> mFullTime_ASD.Words = glm(NotvsFullTime ~ FirstWords.months
+                 #+ age
+                       #+ vabs_soc_ss
+                         , family = binomial, data = dASD.older18)
> summary(mFullTime_ASD.Words)

Call:
glm(formula = NotvsFullTime ~ FirstWords.months, family = binomial,
    data = dASD.older18)

Deviance Residuals:
    Min       1Q   Median       3Q      Max
-1.5883  -1.3129   0.8497   0.9809   1.0614

Coefficients:
                  Estimate Std. Error z value Pr(>|z|)
(Intercept)        0.11696    1.15976   0.101     0.92
FirstWords.months  0.02705    0.05962   0.454     0.65

(Dispersion parameter for binomial family taken to be 1)

    Null deviance: 18.249  on 13  degrees of freedom
Residual deviance: 18.037  on 12  degrees of freedom
  (11 observations deleted due to missingness)
AIC: 22.037

Number of Fisher Scoring iterations: 4

>
> mFullTime_NT.Words = glm(NotvsFullTime ~ FirstWords.months
+                 #+ age
+                       #+ vabs_soc_ss
+                         , family = binomial, data = dNT.older18)
> summary(mFullTime_NT.Words)

Call:
glm(formula = NotvsFullTime ~ FirstWords.months, family = binomial,
    data = dNT.older18)

Deviance Residuals:
    Min       1Q   Median       3Q      Max
-2.1166   0.3501   0.4573   0.4837   1.0194

Coefficients:
                  Estimate Std. Error z value Pr(>|z|)
(Intercept)         4.0297     2.0563   1.960    0.050 .
FirstWords.months  -0.1585     0.1242  -1.277    0.202
---
Signif. codes:  0 ‘***’ 0.001 ‘**’ 0.01 ‘*’ 0.05 ‘.’ 0.1 ‘ ’ 1

(Dispersion parameter for binomial family taken to be 1)

    Null deviance: 16.908  on 19  degrees of freedom
Residual deviance: 15.232  on 18  degrees of freedom
  (7 observations deleted due to missingness)
AIC: 19.232

Number of Fisher Scoring iterations: 5

>
> mFullTime_LAD.Words = glm(NotvsFullTime ~ FirstWords.months
+                 #+ age
+                       #+ vabs_soc_ss
+                         , family = binomial, data = dLAD.older18)
> summary(mFullTime_LAD.Words)

Call:
glm(formula = NotvsFullTime ~ FirstWords.months, family = binomial,
    data = dLAD.older18)

Deviance Residuals:
    Min       1Q   Median       3Q      Max
-2.2573   0.3945   0.4475   0.4777   0.4900

Coefficients:
                  Estimate Std. Error z value Pr(>|z|)
(Intercept)        1.81955    1.67585   1.086    0.278
FirstWords.months  0.02395    0.07833   0.306    0.760

(Dispersion parameter for binomial family taken to be 1)

    Null deviance: 13.404  on 21  degrees of freedom
Residual deviance: 13.302  on 20  degrees of freedom
  (8 observations deleted due to missingness)
AIC: 17.302

Number of Fisher Scoring iterations: 5

>
> #Phrases
> mFullTime_ASD.Phrases = glm(NotvsFullTime ~ FirstPhrases.months
+                 #+ age
+                       #+ vabs_soc_ss
+                         , family = binomial, data = dASD.older18)
> summary(mFullTime_ASD.Phrases)

Call:
glm(formula = NotvsFullTime ~ FirstPhrases.months, family = binomial,
    data = dASD.older18)

Deviance Residuals:
    Min       1Q   Median       3Q      Max
-1.7212  -0.9294   0.6578   0.9361   1.0928

Coefficients:
                    Estimate Std. Error z value Pr(>|z|)
(Intercept)         -0.81870    2.21602  -0.369    0.712
FirstPhrases.months  0.05672    0.07216   0.786    0.432

(Dispersion parameter for binomial family taken to be 1)

    Null deviance: 16.752  on 13  degrees of freedom
Residual deviance: 16.067  on 12  degrees of freedom
  (11 observations deleted due to missingness)
AIC: 20.067

Number of Fisher Scoring iterations: 4

>
> mFullTime_NT.Phrases = glm(NotvsFullTime ~ FirstPhrases.months
+                 #+ age
+                       #+ vabs_soc_ss
+                         , family = binomial, data = dNT.older18)
> summary(mFullTime_NT.Phrases)

Call:
glm(formula = NotvsFullTime ~ FirstPhrases.months, family = binomial,
    data = dNT.older18)

Deviance Residuals:
    Min       1Q   Median       3Q      Max
-1.9909   0.5052   0.5637   0.6764   0.9014

Coefficients:
                    Estimate Std. Error z value Pr(>|z|)
(Intercept)          2.47956    1.51435   1.637    0.102
FirstPhrases.months -0.04969    0.06860  -0.724    0.469

(Dispersion parameter for binomial family taken to be 1)

    Null deviance: 20.862  on 21  degrees of freedom
Residual deviance: 20.350  on 20  degrees of freedom
  (5 observations deleted due to missingness)
AIC: 24.35

Number of Fisher Scoring iterations: 4

>
> mFullTime_LAD.Phrases = glm(NotvsFullTime ~ FirstPhrases.months
+                 #+ age
+                       #+ vabs_soc_ss
+                         , family = binomial, data = dLAD.older18)
> summary(mFullTime_LAD.Phrases)

Call:
glm(formula = NotvsFullTime ~ FirstPhrases.months, family = binomial,
    data = dLAD.older18)

Deviance Residuals:
    Min       1Q   Median       3Q      Max
-2.6411   0.2492   0.3011   0.3751   0.6262

Coefficients:
                    Estimate Std. Error z value Pr(>|z|)
(Intercept)          4.61275    1.83691   2.511    0.012 *
FirstPhrases.months -0.06423    0.03992  -1.609    0.108
---
Signif. codes:  0 ‘***’ 0.001 ‘**’ 0.01 ‘*’ 0.05 ‘.’ 0.1 ‘ ’ 1

(Dispersion parameter for binomial family taken to be 1)

    Null deviance: 13.404  on 21  degrees of freedom
Residual deviance: 10.432  on 20  degrees of freedom
  (8 observations deleted due to missingness)
AIC: 14.432

Number of Fisher Scoring iterations: 5


> ##################################################
> ##### Education completed 22 years and older #####
> ##################################################
> # dASD.older22
> # dASDLAD.older22
>
> ### ASD LAD ###
> mEdComp_ASDLAD.GJ = lm(Edcompl.clean ~
+                           Group.NTref*aprime.log
+                  #+ age
+                       #+ vabs_soc_ss
+                         , data = dASDLAD.older22)
> summary(mEdComp_ASDLAD.GJ)

Call:
lm(formula = Edcompl.clean ~ Group.NTref * aprime.log, data = dASDLAD.older22)

Residuals:
    Min      1Q  Median      3Q     Max
-4.6189 -0.3486 -0.0801  1.0036  1.6514

Coefficients:
                       Estimate Std. Error t value Pr(>|t|)
(Intercept)             11.7416     0.6993  16.791 5.11e-12 ***
Group.NTref              1.4904     1.3986   1.066    0.301
aprime.log               2.5561     5.3102   0.481    0.636
Group.NTref:aprime.log   0.2340    10.6203   0.022    0.983
---
Signif. codes:  0 ‘***’ 0.001 ‘**’ 0.01 ‘*’ 0.05 ‘.’ 0.1 ‘ ’ 1

Residual standard error: 1.599 on 17 degrees of freedom
  (12 observations deleted due to missingness)
Multiple R-squared:  0.2373,	Adjusted R-squared:  0.1027
F-statistic: 1.763 on 3 and 17 DF,  p-value: 0.1924

>
> mEdComp_ASDLAD.Words = lm(Edcompl.clean ~
+                           Group.NTref*FirstWords.months
+                  #+ age
+                       #+ vabs_soc_ss
+                         , data = dASDLAD.older22)
> summary(mEdComp_ASDLAD.Words)

Call:
lm(formula = Edcompl.clean ~ Group.NTref * FirstWords.months,
    data = dASDLAD.older22)

Residuals:
    Min      1Q  Median      3Q     Max
-4.6929 -0.3071  0.0120  1.0124  1.7055

Coefficients:
                               Estimate Std. Error t value Pr(>|t|)
(Intercept)                   10.828889   0.887788  12.198 3.46e-09 ***
Group.NTref                    1.467141   1.775575   0.826    0.422
FirstWords.months              0.034330   0.038832   0.884    0.391
Group.NTref:FirstWords.months  0.002258   0.077665   0.029    0.977
---
Signif. codes:  0 ‘***’ 0.001 ‘**’ 0.01 ‘*’ 0.05 ‘.’ 0.1 ‘ ’ 1

Residual standard error: 1.659 on 15 degrees of freedom
  (14 observations deleted due to missingness)
Multiple R-squared:  0.2439,	Adjusted R-squared:  0.09263
F-statistic: 1.613 on 3 and 15 DF,  p-value: 0.2284

>
> mEdComp_ASDLAD.Phrases = lm(Edcompl.clean ~
+                           Group.NTref*FirstPhrases.months
+                  #+ age
+                       #+ vabs_soc_ss
+                         , data = dASDLAD.older22)
> summary(mEdComp_ASDLAD.Phrases)

Call:
lm(formula = Edcompl.clean ~ Group.NTref * FirstPhrases.months,
    data = dASDLAD.older22)

Residuals:
    Min      1Q  Median      3Q     Max
-3.9811 -0.1813  0.1488  0.4658  2.0189

Coefficients:
                                Estimate Std. Error t value Pr(>|t|)
(Intercept)                      8.71641    0.98290   8.868 8.73e-08 ***
Group.NTref                      3.68280    1.96579   1.873  0.07830 .
FirstPhrases.months              0.09489    0.03211   2.955  0.00886 **
Group.NTref:FirstPhrases.months -0.06907    0.06421  -1.076  0.29712
---
Signif. codes:  0 ‘***’ 0.001 ‘**’ 0.01 ‘*’ 0.05 ‘.’ 0.1 ‘ ’ 1

Residual standard error: 1.327 on 17 degrees of freedom
  (12 observations deleted due to missingness)
Multiple R-squared:  0.4572,	Adjusted R-squared:  0.3614
F-statistic: 4.773 on 3 and 17 DF,  p-value: 0.01365

>
>
>
> ### ASD NT ###
> mEdComp_ASDNT = lm(Edcompl.clean ~
+                           Group.LADref
+                 #+ age
+                       #+ vabs_soc_ss
+                         , data = dASDNT.older22)
> summary(mEdComp_ASDNT)

Call:
lm(formula = Edcompl.clean ~ Group.LADref, data = dASDNT.older22)

Residuals:
    Min      1Q  Median      3Q     Max
-7.1111 -0.1111  0.0909  1.0909  2.8889

Coefficients:
             Estimate Std. Error t value Pr(>|t|)
(Intercept)   11.5101     0.3839  29.980   <2e-16 ***
Group.LADref   1.2020     0.7678   1.565    0.129
---
Signif. codes:  0 ‘***’ 0.001 ‘**’ 0.01 ‘*’ 0.05 ‘.’ 0.1 ‘ ’ 1

Residual standard error: 2.006 on 27 degrees of freedom
  (6 observations deleted due to missingness)
Multiple R-squared:  0.08321,	Adjusted R-squared:  0.04926
F-statistic: 2.451 on 1 and 27 DF,  p-value: 0.1291

>
> mEdComp_ASDNT.GJ = lm(Edcompl.clean ~
+                           Group.LADref*aprime.log
+                 #+ age
+                       #+ vabs_soc_ss
+                         , data = dASDNT.older22)
> summary(mEdComp_ASDNT.GJ)

Call:
lm(formula = Edcompl.clean ~ Group.LADref * aprime.log, data = dASDNT.older22)

Residuals:
    Min      1Q  Median      3Q     Max
-7.0079 -0.1638  0.3423  0.9990  2.9852

Coefficients:
                        Estimate Std. Error t value Pr(>|t|)
(Intercept)              11.4640     0.8307  13.801 2.59e-12 ***
Group.LADref              0.9353     1.6614   0.563    0.579
aprime.log                0.4825     4.9422   0.098    0.923
Group.LADref:aprime.log  -3.9133     9.8843  -0.396    0.696
---
Signif. codes:  0 ‘***’ 0.001 ‘**’ 0.01 ‘*’ 0.05 ‘.’ 0.1 ‘ ’ 1

Residual standard error: 2.184 on 22 degrees of freedom
  (9 observations deleted due to missingness)
Multiple R-squared:  0.1102,	Adjusted R-squared:  -0.01111
F-statistic: 0.9085 on 3 and 22 DF,  p-value: 0.4529

>
> mEdComp_ASDNT.Words = lm(Edcompl.clean ~
+                           Group.LADref*FirstWords.months
+                 #+ age
+                       #+ vabs_soc_ss
+                         , data = dASDNT.older22)
> summary(mEdComp_ASDNT.Words)

Call:
lm(formula = Edcompl.clean ~ Group.LADref * FirstWords.months,
    data = dASDNT.older22)

Residuals:
    Min      1Q  Median      3Q     Max
-7.0115 -0.0115  0.2913  1.2407  2.5344

Coefficients:
                               Estimate Std. Error t value Pr(>|t|)
(Intercept)                    10.59926    1.20537   8.793 1.75e-08 ***
Group.LADref                    1.00789    2.41075   0.418    0.680
FirstWords.months               0.05444    0.06742   0.808    0.428
Group.LADref:FirstWords.months  0.04249    0.13483   0.315    0.756
---
Signif. codes:  0 ‘***’ 0.001 ‘**’ 0.01 ‘*’ 0.05 ‘.’ 0.1 ‘ ’ 1

Residual standard error: 2.225 on 21 degrees of freedom
  (10 observations deleted due to missingness)
Multiple R-squared:  0.1168,	Adjusted R-squared:  -0.009388
F-statistic: 0.9256 on 3 and 21 DF,  p-value: 0.4457

>
> mEdComp_ASDNT.Phrases = lm(Edcompl.clean ~
+                           Group.LADref*FirstPhrases.months
+                 #+ age
+                       #+ vabs_soc_ss
+                         , data = dASDNT.older22)
> summary(mEdComp_ASDNT.Phrases)

Call:
lm(formula = Edcompl.clean ~ Group.LADref * FirstPhrases.months,
    data = dASDNT.older22)

Residuals:
    Min      1Q  Median      3Q     Max
-6.8563 -0.4738  0.2006  1.0807  2.8917

Coefficients:
                                 Estimate Std. Error t value Pr(>|t|)
(Intercept)                       9.03796    1.31339   6.881 4.07e-07 ***
Group.LADref                      4.32589    2.62677   1.647    0.113
FirstPhrases.months               0.08992    0.04902   1.834    0.079 .
Group.LADref:FirstPhrases.months -0.07901    0.09804  -0.806    0.428
---
Signif. codes:  0 ‘***’ 0.001 ‘**’ 0.01 ‘*’ 0.05 ‘.’ 0.1 ‘ ’ 1

Residual standard error: 1.972 on 24 degrees of freedom
  (7 observations deleted due to missingness)
Multiple R-squared:  0.2096,	Adjusted R-squared:  0.1108
F-statistic: 2.121 on 3 and 24 DF,  p-value: 0.124

>
>
>
>
> ### LAD NT ###
> mEdComp_LADNT.GJ = lm(Edcompl.clean ~
+                           Group.ASDref*aprime.log
+                 #+ age
+                       #+ vabs_soc_ss
+                         , data = dLADNT.older22)
> summary(mEdComp_LADNT.GJ)

Call:
lm(formula = Edcompl.clean ~ Group.ASDref * aprime.log, data = dLADNT.older22)

Residuals:
    Min      1Q  Median      3Q     Max
-7.0079 -0.3262 -0.0881  0.8057  2.9852

Coefficients:
                        Estimate Std. Error t value Pr(>|t|)
(Intercept)              12.2092     0.6228  19.603   <2e-16 ***
Group.ASDref             -0.5550     1.2456  -0.446    0.659
aprime.log                0.5995     5.3491   0.112    0.912
Group.ASDref:aprime.log  -4.1473    10.6982  -0.388    0.701
---
Signif. codes:  0 ‘***’ 0.001 ‘**’ 0.01 ‘*’ 0.05 ‘.’ 0.1 ‘ ’ 1

Residual standard error: 1.651 on 27 degrees of freedom
  (5 observations deleted due to missingness)
Multiple R-squared:  0.00759,	Adjusted R-squared:  -0.1027
F-statistic: 0.06883 on 3 and 27 DF,  p-value: 0.976

>
> mEdComp_LADNT.Words = lm(Edcompl.clean ~
+                           Group.ASDref*FirstWords.months
+                 #+ age
+                       #+ vabs_soc_ss
+                         , data = dLADNT.older22)
> summary(mEdComp_LADNT.Words)

Call:
lm(formula = Edcompl.clean ~ Group.ASDref * FirstWords.months,
    data = dLADNT.older22)

Residuals:
    Min      1Q  Median      3Q     Max
-7.0115 -0.3603  0.0381  0.9253  2.5344

Coefficients:
                               Estimate Std. Error t value Pr(>|t|)
(Intercept)                    11.33284    0.84759  13.371 4.85e-12 ***
Group.ASDref                   -0.45925    1.69518  -0.271    0.789
FirstWords.months               0.05557    0.04893   1.136    0.268
Group.ASDref:FirstWords.months  0.04023    0.09786   0.411    0.685
---
Signif. codes:  0 ‘***’ 0.001 ‘**’ 0.01 ‘*’ 0.05 ‘.’ 0.1 ‘ ’ 1

Residual standard error: 1.756 on 22 degrees of freedom
  (10 observations deleted due to missingness)
Multiple R-squared:  0.05875,	Adjusted R-squared:  -0.0696
F-statistic: 0.4577 on 3 and 22 DF,  p-value: 0.7146

>
> mEdComp_LADNT.Phrases = lm(Edcompl.clean ~
+                           Group.ASDref*FirstPhrases.months
+                 #+ age
+                       #+ vabs_soc_ss
+                         , data = dLADNT.older22)
> summary(mEdComp_LADNT.Phrases)

Call:
lm(formula = Edcompl.clean ~ Group.ASDref * FirstPhrases.months,
    data = dLADNT.older22)

Residuals:
    Min      1Q  Median      3Q     Max
-6.8563 -0.1083  0.0933  0.5388  2.8917

Coefficients:
                                 Estimate Std. Error t value Pr(>|t|)
(Intercept)                      10.87936    0.94559  11.505 1.76e-11 ***
Group.ASDref                      0.64310    1.89119   0.340    0.737
FirstPhrases.months               0.05538    0.03933   1.408    0.171
Group.ASDref:FirstPhrases.months -0.00994    0.07866  -0.126    0.900
---
Signif. codes:  0 ‘***’ 0.001 ‘**’ 0.01 ‘*’ 0.05 ‘.’ 0.1 ‘ ’ 1

Residual standard error: 1.648 on 25 degrees of freedom
  (7 observations deleted due to missingness)
Multiple R-squared:  0.07538,	Adjusted R-squared:  -0.03558
F-statistic: 0.6794 on 3 and 25 DF,  p-value: 0.5729

>
>
> ###Follow up
> #aprime.log
> mEdComp_ASD.GJ = lm(Edcompl.clean ~ aprime.log
+                 #+ age
+                       #+ vabs_soc_ss
+                        , data = dASD.older22)
> summary(mEdComp_ASD.GJ)

Call:
lm(formula = Edcompl.clean ~ aprime.log, data = dASD.older22)

Residuals:
    Min      1Q  Median      3Q     Max
-4.6189 -0.0256  1.0036  1.4122  1.5053

Coefficients:
            Estimate Std. Error t value Pr(>|t|)
(Intercept)   10.996      1.554   7.078 0.000399 ***
aprime.log     2.439      8.399   0.290 0.781275
---
Signif. codes:  0 ‘***’ 0.001 ‘**’ 0.01 ‘*’ 0.05 ‘.’ 0.1 ‘ ’ 1

Residual standard error: 2.495 on 6 degrees of freedom
  (8 observations deleted due to missingness)
Multiple R-squared:  0.01386,	Adjusted R-squared:  -0.1505
F-statistic: 0.08434 on 1 and 6 DF,  p-value: 0.7813

>
> mEdComp_NT.GJ = lm(Edcompl.clean ~ aprime.log
+                 #+ age
+                       #+ vabs_soc_ss
+                         , data = dNT.older22)
> summary(mEdComp_NT.GJ)

Call:
lm(formula = Edcompl.clean ~ aprime.log, data = dNT.older22)

Residuals:
    Min      1Q  Median      3Q     Max
-7.0079 -0.1638 -0.0079  0.8900  2.9852

Coefficients:
            Estimate Std. Error t value Pr(>|t|)
(Intercept)  11.9317     0.8984  13.281 4.67e-10 ***
aprime.log   -1.4742     6.2183  -0.237    0.816
---
Signif. codes:  0 ‘***’ 0.001 ‘**’ 0.01 ‘*’ 0.05 ‘.’ 0.1 ‘ ’ 1

Residual standard error: 2.055 on 16 degrees of freedom
  (1 observation deleted due to missingness)
Multiple R-squared:  0.0035,	Adjusted R-squared:  -0.05878
F-statistic: 0.0562 on 1 and 16 DF,  p-value: 0.8156

>
> mEdComp_LAD.GJ = lm(Edcompl.clean ~ aprime.log
+                 #+ age
+                       #+ vabs_soc_ss
+                         , data = dLAD.older22)
> summary(mEdComp_LAD.GJ)

Call:
lm(formula = Edcompl.clean ~ aprime.log, data = dLAD.older22)

Residuals:
     Min       1Q   Median       3Q      Max
-1.07302 -0.34860 -0.20317 -0.07302  1.65140

Coefficients:
            Estimate Std. Error t value Pr(>|t|)
(Intercept)  12.4867     0.4573  27.304 1.85e-11 ***
aprime.log    2.6731     4.2619   0.627    0.543
---
Signif. codes:  0 ‘***’ 0.001 ‘**’ 0.01 ‘*’ 0.05 ‘.’ 0.1 ‘ ’ 1

Residual standard error: 0.7441 on 11 degrees of freedom
  (4 observations deleted due to missingness)
Multiple R-squared:  0.03453,	Adjusted R-squared:  -0.05324
F-statistic: 0.3934 on 1 and 11 DF,  p-value: 0.5433

>
> #Words
> mEdComp_ASD.Words = lm(Edcompl.clean ~ FirstWords.months
+                 #+ age
+                       #+ vabs_soc_ss
+                         , data = dASD.older22)
> summary(mEdComp_ASD.Words)

Call:
lm(formula = Edcompl.clean ~ FirstWords.months, data = dASD.older22)

Residuals:
    Min      1Q  Median      3Q     Max
-4.6929  0.1743  0.9419  1.2407  1.7055

Coefficients:
                  Estimate Std. Error t value Pr(>|t|)
(Intercept)       10.09532    1.95867   5.154  0.00132 **
FirstWords.months  0.03320    0.08731   0.380  0.71504
---
Signif. codes:  0 ‘***’ 0.001 ‘**’ 0.01 ‘*’ 0.05 ‘.’ 0.1 ‘ ’ 1

Residual standard error: 2.353 on 7 degrees of freedom
  (7 observations deleted due to missingness)
Multiple R-squared:  0.02024,	Adjusted R-squared:  -0.1197
F-statistic: 0.1446 on 1 and 7 DF,  p-value: 0.715

>
> mEdComp_NT.Words = lm(Edcompl.clean ~ FirstWords.months
+                 #+ age
+                       #+ vabs_soc_ss
+                         , data = dNT.older22)
> summary(mEdComp_NT.Words)

Call:
lm(formula = Edcompl.clean ~ FirstWords.months, data = dNT.older22)

Residuals:
    Min      1Q  Median      3Q     Max
-7.0115 -0.1250  0.1858  1.0642  2.5344

Coefficients:
                  Estimate Std. Error t value Pr(>|t|)
(Intercept)       11.10321    1.49652   7.419 3.25e-06 ***
FirstWords.months  0.07569    0.10339   0.732    0.476
---
Signif. codes:  0 ‘***’ 0.001 ‘**’ 0.01 ‘*’ 0.05 ‘.’ 0.1 ‘ ’ 1

Residual standard error: 2.159 on 14 degrees of freedom
  (3 observations deleted due to missingness)
Multiple R-squared:  0.03687,	Adjusted R-squared:  -0.03193
F-statistic: 0.5359 on 1 and 14 DF,  p-value: 0.4762

>
> mEdComp_LAD.Words = lm(Edcompl.clean ~ FirstWords.months
+                 #+ age
+                       #+ vabs_soc_ss
+                         , data = dLAD.older22)
> summary(mEdComp_LAD.Words)

Call:
lm(formula = Edcompl.clean ~ FirstWords.months, data = dLAD.older22)

Residuals:
     Min       1Q   Median       3Q      Max
-0.59076 -0.36028 -0.05888  0.01204  1.08295

Coefficients:
                  Estimate Std. Error t value Pr(>|t|)
(Intercept)       11.56246    0.37981  30.443 1.47e-09 ***
FirstWords.months  0.03546    0.01611   2.201   0.0589 .
---
Signif. codes:  0 ‘***’ 0.001 ‘**’ 0.01 ‘*’ 0.05 ‘.’ 0.1 ‘ ’ 1

Residual standard error: 0.565 on 8 degrees of freedom
  (7 observations deleted due to missingness)
Multiple R-squared:  0.3771,	Adjusted R-squared:  0.2992
F-statistic: 4.843 on 1 and 8 DF,  p-value: 0.05894

>
> #Phrases
> mEdComp_ASD.Phrases = lm(Edcompl.clean ~ FirstPhrases.months
+                #+ age
+                       #+ vabs_soc_ss
+                         , data = dASD.older22)
> summary(mEdComp_ASD.Phrases)

Call:
lm(formula = Edcompl.clean ~ FirstPhrases.months, data = dASD.older22)

Residuals:
    Min      1Q  Median      3Q     Max
-3.9811 -0.8607  0.3364  1.1865  2.0189

Coefficients:
                    Estimate Std. Error t value Pr(>|t|)
(Intercept)          6.87502    2.14221   3.209   0.0124 *
FirstPhrases.months  0.12942    0.06623   1.954   0.0865 .
---
Signif. codes:  0 ‘***’ 0.001 ‘**’ 0.01 ‘*’ 0.05 ‘.’ 0.1 ‘ ’ 1

Residual standard error: 1.86 on 8 degrees of freedom
  (6 observations deleted due to missingness)
Multiple R-squared:  0.3231,	Adjusted R-squared:  0.2385
F-statistic: 3.818 on 1 and 8 DF,  p-value: 0.08645

>
> mEdComp_NT.Phrases = lm(Edcompl.clean ~ FirstPhrases.months
+                 #+ age
+                       #+ vabs_soc_ss
+                         , data = dNT.older22)
> summary(mEdComp_NT.Phrases)

Call:
lm(formula = Edcompl.clean ~ FirstPhrases.months, data = dNT.older22)

Residuals:
    Min      1Q  Median      3Q     Max
-6.8563 -0.3352  0.1437  1.0051  2.8917

Coefficients:
                    Estimate Std. Error t value Pr(>|t|)
(Intercept)         11.20091    1.35579   8.262 3.64e-07 ***
FirstPhrases.months  0.05041    0.07028   0.717    0.484
---
Signif. codes:  0 ‘***’ 0.001 ‘**’ 0.01 ‘*’ 0.05 ‘.’ 0.1 ‘ ’ 1

Residual standard error: 2.026 on 16 degrees of freedom
  (1 observation deleted due to missingness)
Multiple R-squared:  0.03116,	Adjusted R-squared:  -0.0294
F-statistic: 0.5145 on 1 and 16 DF,  p-value: 0.4835

>
> mEdComp_LAD.Phrases = lm(Edcompl.clean ~ FirstPhrases.months
+                 #+ age
+                       #+ vabs_soc_ss
+                         , data = dLAD.older22)
> summary(mEdComp_LAD.Phrases)

Call:
lm(formula = Edcompl.clean ~ FirstPhrases.months, data = dLAD.older22)

Residuals:
     Min       1Q   Median       3Q      Max
-1.00625 -0.00625 -0.00625  0.35586  0.54531

Coefficients:
                    Estimate Std. Error t value Pr(>|t|)
(Intercept)         10.55781    0.46559  22.676 2.99e-09 ***
FirstPhrases.months  0.06035    0.01637   3.686  0.00503 **
---
Signif. codes:  0 ‘***’ 0.001 ‘**’ 0.01 ‘*’ 0.05 ‘.’ 0.1 ‘ ’ 1

Residual standard error: 0.4995 on 9 degrees of freedom
  (6 observations deleted due to missingness)
Multiple R-squared:  0.6016,	Adjusted R-squared:  0.5573
F-statistic: 13.59 on 1 and 9 DF,  p-value: 0.005026


> ### RQ 2 C: Vocational Outcome ~ Group*Core Language (youth cohort)/Grammaticality Judgement (A’; all participants)/age of first words/phrases
> ######################
> ##### Have a job #####
> ######################
> # == 0 -0.5 No, 1 0.5 Yes
>
> ### ASD LAD ###
> mJobNotColl_ASDLAD.GJ = glm(edu_pro_4_par ~
+                          Group.NTref*aprime.log
+                 #+ age
+                       #+ vabs_soc_ss
+                        , family = binomial, data = dASDLAD.older18.nocoll)
> summary(mJobNotColl_ASDLAD.GJ)

Call:
glm(formula = edu_pro_4_par ~ Group.NTref * aprime.log, family = binomial,
    data = dASDLAD.older18.nocoll)

Deviance Residuals:
    Min       1Q   Median       3Q      Max
-1.4023  -0.2182   0.0000   0.2421   1.2284

Coefficients:
                       Estimate Std. Error z value Pr(>|z|)
(Intercept)               278.3    57809.2   0.005    0.996
Group.NTref               555.6   115618.5   0.005    0.996
aprime.log               1737.7   362849.2   0.005    0.996
Group.NTref:aprime.log   3469.2   725698.4   0.005    0.996

(Dispersion parameter for binomial family taken to be 1)

    Null deviance: 24.435  on 19  degrees of freedom
Residual deviance: 13.504  on 16  degrees of freedom
  (3 observations deleted due to missingness)
AIC: 21.504

Number of Fisher Scoring iterations: 22

>
> mJobNotColl_ASDLAD.Words = glm(edu_pro_4_par ~
+                          Group.NTref*FirstWords.months
+                 #+ age
+                       #+ vabs_soc_ss,
+                         family = binomial, data = dASDLAD.older18.nocoll)
Error: unexpected symbol in:
"                      #+ vabs_soc_ss,
                        family"
> summary(mJobNotColl_ASDLAD.Words)
Error in h(simpleError(msg, call)) :
  error in evaluating the argument 'object' in selecting a method for function 'summary': object 'mJobNotColl_ASDLAD.Words' not found
>
> mJobNotColl_ASDLAD.Phrases = glm(edu_pro_4_par ~
+                          Group.NTref*FirstPhrases.months
+                 #+ age
+                       #+ vabs_soc_ss,
+                        family = binomial, data = dASDLAD.older18.nocoll)
Error: unexpected symbol in:
"                      #+ vabs_soc_ss,
                       family"
> summary(mJobNotColl_ASDLAD.Phrases)
Error in h(simpleError(msg, call)) :
  error in evaluating the argument 'object' in selecting a method for function 'summary': object 'mJobNotColl_ASDLAD.Phrases' not found
>
>
>
>
>
> ### ASD NT ###
> mJobNotColl_ASDNT.GJ = glm(edu_pro_4_par ~
+                          Group.LADref*aprime.log
+                #+ age
+                       #+ vabs_soc_ss
+                        , family = binomial, data = dASDNT.older18.nocoll)
> summary(mJobNotColl_ASDNT.GJ)

Call:
glm(formula = edu_pro_4_par ~ Group.LADref * aprime.log, family = binomial,
    data = dASDNT.older18.nocoll)

Deviance Residuals:
    Min       1Q   Median       3Q      Max
-1.5170  -1.2630   0.8803   1.1001   1.2580

Coefficients:
                        Estimate Std. Error z value Pr(>|z|)
(Intercept)               0.7641     0.8443   0.905    0.365
Group.LADref              0.4988     1.6886   0.295    0.768
aprime.log                3.8956     5.0166   0.777    0.437
Group.LADref:aprime.log   1.6291    10.0332   0.162    0.871

(Dispersion parameter for binomial family taken to be 1)

    Null deviance: 27.526  on 19  degrees of freedom
Residual deviance: 26.655  on 16  degrees of freedom
  (2 observations deleted due to missingness)
AIC: 34.655

Number of Fisher Scoring iterations: 4

>
> mJobNotColl_ASDNT.Words = glm(edu_pro_4_par ~
+                          Group.LADref*FirstWords.months
+                #+ age
+                       #+ vabs_soc_ss
+                        , family = binomial, data = dASDNT.older18.nocoll)
> summary(mJobNotColl_ASDNT.Words)

Call:
glm(formula = edu_pro_4_par ~ Group.LADref * FirstWords.months,
    family = binomial, data = dASDNT.older18.nocoll)

Deviance Residuals:
    Min       1Q   Median       3Q      Max
-1.5666  -1.1176   0.5278   1.0163   1.5175

Coefficients:
                               Estimate Std. Error z value Pr(>|z|)
(Intercept)                    -1.21946    1.37540  -0.887    0.375
Group.LADref                   -0.07080    2.75079  -0.026    0.979
FirstWords.months               0.10300    0.09002   1.144    0.253
Group.LADref:FirstWords.months  0.06840    0.18005   0.380    0.704

(Dispersion parameter for binomial family taken to be 1)

    Null deviance: 25.864  on 18  degrees of freedom
Residual deviance: 23.594  on 15  degrees of freedom
  (3 observations deleted due to missingness)
AIC: 31.594

Number of Fisher Scoring iterations: 4

>
> mJobNotColl_ASDNT.Phrases = glm(edu_pro_4_par ~
+                          Group.LADref*FirstPhrases.months
+                #+ age
+                       #+ vabs_soc_ss
+                        , family = binomial, data = dASDNT.older18.nocoll)
> summary(mJobNotColl_ASDNT.Phrases)

Call:
glm(formula = edu_pro_4_par ~ Group.LADref * FirstPhrases.months,
    family = binomial, data = dASDNT.older18.nocoll)

Deviance Residuals:
    Min       1Q   Median       3Q      Max
-1.5634  -1.1110   0.5635   1.0230   1.4079

Coefficients:
                                 Estimate Std. Error z value Pr(>|z|)
(Intercept)                      -2.41045    2.10985  -1.142    0.253
Group.LADref                     -0.13773    4.21969  -0.033    0.974
FirstPhrases.months               0.12598    0.09941   1.267    0.205
Group.LADref:FirstPhrases.months  0.07338    0.19883   0.369    0.712

(Dispersion parameter for binomial family taken to be 1)

    Null deviance: 26.920  on 19  degrees of freedom
Residual deviance: 24.448  on 16  degrees of freedom
  (2 observations deleted due to missingness)
AIC: 32.448

Number of Fisher Scoring iterations: 5

>
>
>
>
> ### LAD NT ###
> mJobNotColl_LADNT.GJ = glm(edu_pro_4_par ~
+                          Group.ASDref*aprime.log
+                #+ age
+                       #+ vabs_soc_ss
+                        , family = binomial, data = dLADNT.older18.nocoll)
> summary(mJobNotColl_LADNT.GJ)

Call:
glm(formula = edu_pro_4_par ~ Group.ASDref * aprime.log, family = binomial,
    data = dLADNT.older18.nocoll)

Deviance Residuals:
     Min        1Q    Median        3Q       Max
-1.51696  -0.00004   0.00000   0.88029   1.25796

Coefficients:
                        Estimate Std. Error z value Pr(>|z|)
(Intercept)                278.6    57809.2   0.005    0.996
Group.ASDref              -555.1   115618.5  -0.005    0.996
aprime.log                1738.5   362849.2   0.005    0.996
Group.ASDref:aprime.log  -3467.6   725698.4  -0.005    0.996

(Dispersion parameter for binomial family taken to be 1)

    Null deviance: 22.493  on 19  degrees of freedom
Residual deviance: 13.151  on 16  degrees of freedom
  (1 observation deleted due to missingness)
AIC: 21.151

Number of Fisher Scoring iterations: 22

>
> mJobNotColl_LADNT.Words = glm(edu_pro_4_par ~
+                          Group.ASDref*FirstWords.months
+                #+ age
+                       #+ vabs_soc_ss
+                        , family = binomial, data = dLADNT.older18.nocoll)
> summary(mJobNotColl_LADNT.Words)

Call:
glm(formula = edu_pro_4_par ~ Group.ASDref * FirstWords.months,
    family = binomial, data = dLADNT.older18.nocoll)

Deviance Residuals:
     Min        1Q    Median        3Q       Max
-1.67177   0.00185   0.34832   0.62437   1.30474

Coefficients:
                               Estimate Std. Error z value Pr(>|z|)
(Intercept)                     -4.6317     5.9208  -0.782    0.434
Group.ASDref                     6.7536    11.8416   0.570    0.568
FirstWords.months                0.4832     0.5220   0.926    0.355
Group.ASDref:FirstWords.months  -0.6921     1.0440  -0.663    0.507

(Dispersion parameter for binomial family taken to be 1)

    Null deviance: 19.557  on 18  degrees of freedom
Residual deviance: 15.206  on 15  degrees of freedom
  (2 observations deleted due to missingness)
AIC: 23.206

Number of Fisher Scoring iterations: 9

>
> mJobNotColl_LADNT.Phrases = glm(edu_pro_4_par ~
+                          Group.ASDref*FirstPhrases.months
+                #+ age
+                       #+ vabs_soc_ss
+                        , family = binomial, data = dLADNT.older18.nocoll)
> summary(mJobNotColl_LADNT.Phrases)

Call:
glm(formula = edu_pro_4_par ~ Group.ASDref * FirstPhrases.months,
    family = binomial, data = dLADNT.older18.nocoll)

Deviance Residuals:
     Min        1Q    Median        3Q       Max
-1.37295  -0.00007   0.00000   0.43456   1.40793

Coefficients:
                                   Estimate Std. Error z value Pr(>|z|)
(Intercept)                         31.5505  6168.3279   0.005    0.996
Group.ASDref                       -68.0595 12336.6559  -0.006    0.996
FirstPhrases.months                 -0.4188   101.4397  -0.004    0.997
Group.ASDref:FirstPhrases.months     1.1630   202.8793   0.006    0.995

(Dispersion parameter for binomial family taken to be 1)

    Null deviance: 21.901  on 18  degrees of freedom
Residual deviance: 11.994  on 15  degrees of freedom
  (2 observations deleted due to missingness)
AIC: 19.994

Number of Fisher Scoring iterations: 19

>
>
>
>
>
> ###Follow up
> #aprime.log
> mJobNotColl_ASD.GJ = glm(edu_pro_4_par ~ aprime.log
+                #+ age
+                       #+ vabs_soc_ss
+                      , family = binomial, data = dASD.older18.nocoll)
> summary(mJobNotColl_ASD.GJ)

Call:
glm(formula = edu_pro_4_par ~ aprime.log, family = binomial,
    data = dASD.older18.nocoll)

Deviance Residuals:
     Min        1Q    Median        3Q       Max
-1.40232  -1.18886   0.04789   1.16138   1.22835

Coefficients:
            Estimate Std. Error z value Pr(>|z|)
(Intercept)   0.5147     1.0834   0.475    0.635
aprime.log    3.0810     5.2625   0.585    0.558

(Dispersion parameter for binomial family taken to be 1)

    Null deviance: 13.863  on 9  degrees of freedom
Residual deviance: 13.504  on 8  degrees of freedom
  (2 observations deleted due to missingness)
AIC: 17.504

Number of Fisher Scoring iterations: 4

>
> mJobNotColl_NT.GJ = glm(edu_pro_4_par ~ aprime.log
+                #+ age
+                       #+ vabs_soc_ss
+                        , family = binomial, data = dNT.older18.nocoll)
> summary(mJobNotColl_NT.GJ)

Call:
glm(formula = edu_pro_4_par ~ aprime.log, family = binomial,
    data = dNT.older18.nocoll)

Deviance Residuals:
    Min       1Q   Median       3Q      Max
-1.5170  -1.2472   0.8803   0.9464   1.2580

Coefficients:
            Estimate Std. Error z value Pr(>|z|)
(Intercept)    1.014      1.295   0.782    0.434
aprime.log     4.710      8.542   0.551    0.581

(Dispersion parameter for binomial family taken to be 1)

    Null deviance: 13.460  on 9  degrees of freedom
Residual deviance: 13.151  on 8  degrees of freedom
AIC: 17.151

Number of Fisher Scoring iterations: 4

>
> mJobNotColl_LAD.GJ = glm(edu_pro_4_par ~ aprime.log
+              #+ age
+                       #+ vabs_soc_ss
+                        , family = binomial, data = dLAD.older18.nocoll)
> summary(mJobNotColl_LAD.GJ)

Call:
glm(formula = edu_pro_4_par ~ aprime.log, family = binomial,
    data = dLAD.older18.nocoll)

Deviance Residuals:
       Min          1Q      Median          3Q         Max
-3.235e-05   2.100e-08   2.100e-08   2.100e-08   2.701e-05

Coefficients:
            Estimate Std. Error z value Pr(>|z|)
(Intercept)      646     518113   0.001    0.999
aprime.log      4034    3252018   0.001    0.999

(Dispersion parameter for binomial family taken to be 1)

    Null deviance: 6.5017e+00  on 9  degrees of freedom
Residual deviance: 2.5053e-09  on 8  degrees of freedom
  (1 observation deleted due to missingness)
AIC: 4

Number of Fisher Scoring iterations: 25

>
>
>
> #Words
> mJobNotColl_ASD.Words = glm(edu_pro_4_par ~ FirstWords.months
+                #+ age
+                       #+ vabs_soc_ss
+                        , family = binomial, data = dASD.older18.nocoll)
> summary(mJobNotColl_ASD.Words)

Call:
glm(formula = edu_pro_4_par ~ FirstWords.months, family = binomial,
    data = dASD.older18.nocoll)

Deviance Residuals:
     Min        1Q    Median        3Q       Max
-1.56655  -0.96214   0.00514   1.02737   1.51746

Coefficients:
                  Estimate Std. Error z value Pr(>|z|)
(Intercept)       -1.18406    1.47051  -0.805    0.421
FirstWords.months  0.06880    0.07623   0.903    0.367

(Dispersion parameter for binomial family taken to be 1)

    Null deviance: 13.863  on 9  degrees of freedom
Residual deviance: 12.989  on 8  degrees of freedom
  (2 observations deleted due to missingness)
AIC: 16.989

Number of Fisher Scoring iterations: 4

>
> mJobNotColl_NT.Words = glm(edu_pro_4_par ~ FirstWords.months
+                #+ age
+                       #+ vabs_soc_ss
+                        , family = binomial, data = dNT.older18.nocoll)
> summary(mJobNotColl_NT.Words)

Call:
glm(formula = edu_pro_4_par ~ FirstWords.months, family = binomial,
    data = dNT.older18.nocoll)

Deviance Residuals:
    Min       1Q   Median       3Q      Max
-1.4701  -1.3476   0.5278   1.0163   1.3047

Coefficients:
                  Estimate Std. Error z value Pr(>|z|)
(Intercept)        -1.2549     2.3247  -0.540    0.589
FirstWords.months   0.1372     0.1631   0.841    0.400

(Dispersion parameter for binomial family taken to be 1)

    Null deviance: 11.457  on 8  degrees of freedom
Residual deviance: 10.604  on 7  degrees of freedom
  (1 observation deleted due to missingness)
AIC: 14.604

Number of Fisher Scoring iterations: 4

>
> mJobNotColl_LAD.Words = glm(edu_pro_4_par ~ FirstWords.months
+                #+ age
+                       #+ vabs_soc_ss
+                      , family = binomial, data = dLAD.older18.nocoll)
> summary(mJobNotColl_LAD.Words)

Call:
glm(formula = edu_pro_4_par ~ FirstWords.months, family = binomial,
    data = dLAD.older18.nocoll)

Deviance Residuals:
     Min        1Q    Median        3Q       Max
-1.67177   0.01389   0.09918   0.47526   1.05937

Coefficients:
                  Estimate Std. Error z value Pr(>|z|)
(Intercept)        -8.0084    11.6112  -0.690    0.490
FirstWords.months   0.8293     1.0311   0.804    0.421

(Dispersion parameter for binomial family taken to be 1)

    Null deviance: 6.5017  on 9  degrees of freedom
Residual deviance: 4.6018  on 8  degrees of freedom
  (1 observation deleted due to missingness)
AIC: 8.6018

Number of Fisher Scoring iterations: 9

>
>
>
> #Phrases
> mJobNotColl_ASD.Phrases = glm(edu_pro_4_par ~ FirstPhrases.months
+                #+ age
+                       #+ vabs_soc_ss
+                        , family = binomial, data = dASD.older18.nocoll)
> summary(mJobNotColl_ASD.Phrases)

Call:
glm(formula = edu_pro_4_par ~ FirstPhrases.months, family = binomial,
    data = dASD.older18.nocoll)

Deviance Residuals:
    Min       1Q   Median       3Q      Max
-1.5634  -1.0944   0.6027   1.0420   1.2628

Coefficients:
                    Estimate Std. Error z value Pr(>|z|)
(Intercept)         -2.34158    2.96945  -0.789    0.430
FirstPhrases.months  0.08930    0.09657   0.925    0.355

(Dispersion parameter for binomial family taken to be 1)

    Null deviance: 13.460  on 9  degrees of freedom
Residual deviance: 12.454  on 8  degrees of freedom
  (2 observations deleted due to missingness)
AIC: 16.454

Number of Fisher Scoring iterations: 3

>
> mJobNotColl_NT.Phrases = glm(edu_pro_4_par ~ FirstPhrases.months
+                #+ age
+                       #+ vabs_soc_ss
+                        , family = binomial, data = dNT.older18.nocoll)
> summary(mJobNotColl_NT.Phrases)

Call:
glm(formula = edu_pro_4_par ~ FirstPhrases.months, family = binomial,
    data = dNT.older18.nocoll)

Deviance Residuals:
    Min       1Q   Median       3Q      Max
-1.3729  -1.1274   0.4346   0.9937   1.4079

Coefficients:
                    Estimate Std. Error z value Pr(>|z|)
(Intercept)          -2.4793     2.9979  -0.827    0.408
FirstPhrases.months   0.1627     0.1738   0.936    0.349

(Dispersion parameter for binomial family taken to be 1)

    Null deviance: 13.460  on 9  degrees of freedom
Residual deviance: 11.994  on 8  degrees of freedom
AIC: 15.994

Number of Fisher Scoring iterations: 5

>
> mJobNotColl_LAD.Phrases = glm(edu_pro_4_par ~ FirstPhrases.months
+                #+ age
+                       #+ vabs_soc_ss
+                        , family = binomial, data = dLAD.older18.nocoll)
> summary(mJobNotColl_LAD.Phrases)

Call:
glm(formula = edu_pro_4_par ~ FirstPhrases.months, family = binomial,
    data = dLAD.older18.nocoll)

Deviance Residuals:
       Min          1Q      Median          3Q         Max
-1.147e-05   2.110e-08   2.110e-08   2.110e-08   1.780e-05

Coefficients:
                      Estimate Std. Error z value Pr(>|z|)
(Intercept)             83.913 150292.673   0.001        1
FirstPhrases.months     -1.278   2471.576  -0.001        1

(Dispersion parameter for binomial family taken to be 1)

    Null deviance: 6.2790e+00  on 8  degrees of freedom
Residual deviance: 4.4839e-10  on 7  degrees of freedom
  (2 observations deleted due to missingness)
AIC: 4

Number of Fisher Scoring iterations: 24


> ########################################
> ##### Hours per week spent working #####
> ########################################
> #HoursWorking.log
>
> ### ASD LAD ###
> mJobHoursNotColl_ASDLAD.GJ = lm(HoursWorking.log ~
+                           Group.NTref*aprime.log
+                  #+ age
+                       #+ vabs_soc_ss
+                         , data = dASDLAD.older18.nocoll)
> summary(mJobHoursNotColl_ASDLAD.GJ)

Call:
lm(formula = HoursWorking.log ~ Group.NTref * aprime.log, data = dASDLAD.older18.nocoll)

Residuals:
     Min       1Q   Median       3Q      Max
-0.49484  0.00000  0.00000  0.02684  0.17271

Coefficients:
                       Estimate Std. Error t value Pr(>|t|)
(Intercept)              3.5508     0.1084  32.768 1.65e-11 ***
Group.NTref              0.2761     0.2167   1.274    0.231
aprime.log              -0.2514     0.8237  -0.305    0.766
Group.NTref:aprime.log   0.5029     1.6474   0.305    0.766
---
Signif. codes:  0 ‘***’ 0.001 ‘**’ 0.01 ‘*’ 0.05 ‘.’ 0.1 ‘ ’ 1

Residual standard error: 0.1785 on 10 degrees of freedom
  (9 observations deleted due to missingness)
Multiple R-squared:  0.306,	Adjusted R-squared:  0.09778
F-statistic:  1.47 on 3 and 10 DF,  p-value: 0.2813

>
> mJobHoursNotColl_ASDLAD.Words = lm(HoursWorking.log ~
+                           Group.NTref*FirstWords.months
+                  #+ age
+                       #+ vabs_soc_ss
+                         , data = dASDLAD.older18.nocoll)
> summary(mJobHoursNotColl_ASDLAD.Words)

Call:
lm(formula = HoursWorking.log ~ Group.NTref * FirstWords.months,
    data = dASDLAD.older18.nocoll)

Residuals:
     Min       1Q   Median       3Q      Max
-0.05246  0.00000  0.00000  0.00000  0.12639

Coefficients:
                               Estimate Std. Error t value Pr(>|t|)
(Intercept)                    3.280594   0.032943  99.585 2.55e-16 ***
Group.NTref                    0.816571   0.065885  12.394 2.16e-07 ***
FirstWords.months              0.014657   0.001539   9.521 2.49e-06 ***
Group.NTref:FirstWords.months -0.029315   0.003079  -9.521 2.49e-06 ***
---
Signif. codes:  0 ‘***’ 0.001 ‘**’ 0.01 ‘*’ 0.05 ‘.’ 0.1 ‘ ’ 1

Residual standard error: 0.04731 on 10 degrees of freedom
  (9 observations deleted due to missingness)
Multiple R-squared:  0.9513,	Adjusted R-squared:  0.9368
F-statistic: 65.18 on 3 and 10 DF,  p-value: 7.229e-07

>
> mJobHoursNotColl_ASDLAD.Phrases = lm(HoursWorking.log ~
+                           Group.NTref*FirstPhrases.months
+                  #+ age
+                       #+ vabs_soc_ss
+                         , data = dASDLAD.older18.nocoll)
> summary(mJobHoursNotColl_ASDLAD.Phrases)

Call:
lm(formula = HoursWorking.log ~ Group.NTref * FirstPhrases.months,
    data = dASDLAD.older18.nocoll)

Residuals:
    Min      1Q  Median      3Q     Max
-0.5080  0.0000  0.0000  0.0738  0.1619

Coefficients:
                                 Estimate Std. Error t value Pr(>|t|)
(Intercept)                      3.512580   0.168654  20.827 1.44e-09 ***
Group.NTref                      0.352599   0.337308   1.045    0.320
FirstPhrases.months              0.002326   0.005178   0.449    0.663
Group.NTref:FirstPhrases.months -0.004651   0.010357  -0.449    0.663
---
Signif. codes:  0 ‘***’ 0.001 ‘**’ 0.01 ‘*’ 0.05 ‘.’ 0.1 ‘ ’ 1

Residual standard error: 0.178 on 10 degrees of freedom
  (9 observations deleted due to missingness)
Multiple R-squared:  0.3113,	Adjusted R-squared:  0.1047
F-statistic: 1.507 on 3 and 10 DF,  p-value: 0.2722

>
>
>
>
> ### ASD NT ###
> mJobHoursNotColl_ASDNT.GJ = lm(HoursWorking.log ~
+                           Group.LADref*aprime.log
+                 #+ age
+                       #+ vabs_soc_ss
+                         , data = dASDNT.older18.nocoll)
> summary(mJobHoursNotColl_ASDNT.GJ)

Call:
lm(formula = HoursWorking.log ~ Group.LADref * aprime.log, data = dASDNT.older18.nocoll)

Residuals:
     Min       1Q   Median       3Q      Max
-0.49484 -0.04086  0.03700  0.09423  0.17271

Coefficients:
                        Estimate Std. Error t value Pr(>|t|)
(Intercept)               3.5036     0.1373  25.526 3.62e-08 ***
Group.LADref              0.1818     0.2745   0.662    0.529
aprime.log               -0.4248     0.9064  -0.469    0.654
Group.LADref:aprime.log   0.1562     1.8129   0.086    0.934
---
Signif. codes:  0 ‘***’ 0.001 ‘**’ 0.01 ‘*’ 0.05 ‘.’ 0.1 ‘ ’ 1

Residual standard error: 0.2236 on 7 degrees of freedom
  (11 observations deleted due to missingness)
Multiple R-squared:  0.1707,	Adjusted R-squared:  -0.1847
F-statistic: 0.4804 on 3 and 7 DF,  p-value: 0.7061

>
> mJobHoursNotColl_ASDNT.Words = lm(HoursWorking.log ~
+                           Group.LADref*FirstWords.months
+                 #+ age
+                       #+ vabs_soc_ss
+                         , data = dASDNT.older18.nocoll)
> summary(mJobHoursNotColl_ASDNT.Words)

Call:
lm(formula = HoursWorking.log ~ Group.LADref * FirstWords.months,
    data = dASDNT.older18.nocoll)

Residuals:
     Min       1Q   Median       3Q      Max
-0.11965 -0.05098  0.00913  0.05135  0.12639

Coefficients:
                                Estimate Std. Error t value Pr(>|t|)
(Intercept)                     3.276119   0.074449  44.005 8.17e-10 ***
Group.LADref                    0.807620   0.148899   5.424 0.000983 ***
FirstWords.months               0.013244   0.003909   3.388 0.011629 *
Group.LADref:FirstWords.months -0.032141   0.007818  -4.111 0.004509 **
---
Signif. codes:  0 ‘***’ 0.001 ‘**’ 0.01 ‘*’ 0.05 ‘.’ 0.1 ‘ ’ 1

Residual standard error: 0.08925 on 7 degrees of freedom
  (11 observations deleted due to missingness)
Multiple R-squared:  0.8628,	Adjusted R-squared:  0.804
F-statistic: 14.67 on 3 and 7 DF,  p-value: 0.002106

>
> mJobHoursNotColl_ASDLAD.Phrases = lm(HoursWorking.log ~
+                           Group.NTref*FirstPhrases.months
+                  #+ age
+                       #+ vabs_soc_ss
+                         , data = dASDLAD.older18.nocoll)
> summary(mJobHoursNotColl_ASDLAD.Phrases)

Call:
lm(formula = HoursWorking.log ~ Group.NTref * FirstPhrases.months,
    data = dASDLAD.older18.nocoll)

Residuals:
    Min      1Q  Median      3Q     Max
-0.5080  0.0000  0.0000  0.0738  0.1619

Coefficients:
                                 Estimate Std. Error t value Pr(>|t|)
(Intercept)                      3.512580   0.168654  20.827 1.44e-09 ***
Group.NTref                      0.352599   0.337308   1.045    0.320
FirstPhrases.months              0.002326   0.005178   0.449    0.663
Group.NTref:FirstPhrases.months -0.004651   0.010357  -0.449    0.663
---
Signif. codes:  0 ‘***’ 0.001 ‘**’ 0.01 ‘*’ 0.05 ‘.’ 0.1 ‘ ’ 1

Residual standard error: 0.178 on 10 degrees of freedom
  (9 observations deleted due to missingness)
Multiple R-squared:  0.3113,	Adjusted R-squared:  0.1047
F-statistic: 1.507 on 3 and 10 DF,  p-value: 0.2722

>
>
>
>
>
> ### LAD NT ###
> mJobHoursNotColl_LADNT.GJ = lm(HoursWorking.log ~
+                           Group.ASDref*aprime.log
+                 #+ age
+                       #+ vabs_soc_ss
+                         , data = dLADNT.older18.nocoll)
> summary(mJobHoursNotColl_LADNT.GJ)

Call:
lm(formula = HoursWorking.log ~ Group.ASDref * aprime.log, data = dLADNT.older18.nocoll)

Residuals:
      Min        1Q    Median        3Q       Max
-0.104936  0.000000  0.000000  0.002979  0.074834

Coefficients:
                        Estimate Std. Error t value Pr(>|t|)
(Intercept)              3.64170    0.02830 128.669   <2e-16 ***
Group.ASDref            -0.09437    0.05661  -1.667    0.124
aprime.log              -0.17337    0.23991  -0.723    0.485
Group.ASDref:aprime.log -0.34674    0.47982  -0.723    0.485
---
Signif. codes:  0 ‘***’ 0.001 ‘**’ 0.01 ‘*’ 0.05 ‘.’ 0.1 ‘ ’ 1

Residual standard error: 0.05335 on 11 degrees of freedom
  (6 observations deleted due to missingness)
Multiple R-squared:  0.3162,	Adjusted R-squared:  0.1297
F-statistic: 1.696 on 3 and 11 DF,  p-value: 0.2253

>
> mJobHoursNotColl_LADNT.Words = lm(HoursWorking.log ~
+                           Group.ASDref*FirstWords.months
+                 #+ age
+                         , data = dLADNT.older18.nocoll)
> summary(mJobHoursNotColl_LADNT.Words)

Call:
lm(formula = HoursWorking.log ~ Group.ASDref * FirstWords.months,
    data = dLADNT.older18.nocoll)

Residuals:
     Min       1Q   Median       3Q      Max
-0.11965  0.00000  0.00000  0.01437  0.07396

Coefficients:
                                Estimate Std. Error t value Pr(>|t|)
(Intercept)                     3.684404   0.036075 102.132   <2e-16 ***
Group.ASDref                   -0.008950   0.072150  -0.124    0.904
FirstWords.months              -0.001413   0.001997  -0.708    0.494
Group.ASDref:FirstWords.months -0.002826   0.003994  -0.708    0.494
---
Signif. codes:  0 ‘***’ 0.001 ‘**’ 0.01 ‘*’ 0.05 ‘.’ 0.1 ‘ ’ 1

Residual standard error: 0.05509 on 11 degrees of freedom
  (6 observations deleted due to missingness)
Multiple R-squared:  0.2709,	Adjusted R-squared:  0.07206
F-statistic: 1.362 on 3 and 11 DF,  p-value: 0.3051

>
> mJobHoursNotColl_LADNT.Phrases = lm(HoursWorking.log ~
+                           Group.ASDref*FirstPhrases.months
+                 #+ age
+                       #+ vabs_soc_ss
+                         , data = dLADNT.older18.nocoll)
> summary(mJobHoursNotColl_LADNT.Phrases)

Call:
lm(formula = HoursWorking.log ~ Group.ASDref * FirstPhrases.months,
    data = dLADNT.older18.nocoll)

Residuals:
      Min        1Q    Median        3Q       Max
-0.122972  0.000000  0.000000  0.004979  0.077940

Coefficients:
                                  Estimate Std. Error t value Pr(>|t|)
(Intercept)                       3.718468   0.037946  97.992    3e-16 ***
Group.ASDref                      0.059177   0.075893   0.780    0.454
FirstPhrases.months              -0.002742   0.001543  -1.777    0.106
Group.ASDref:FirstPhrases.months -0.005485   0.003086  -1.777    0.106
---
Signif. codes:  0 ‘***’ 0.001 ‘**’ 0.01 ‘*’ 0.05 ‘.’ 0.1 ‘ ’ 1

Residual standard error: 0.04891 on 10 degrees of freedom
  (7 observations deleted due to missingness)
Multiple R-squared:  0.4716,	Adjusted R-squared:  0.3131
F-statistic: 2.975 on 3 and 10 DF,  p-value: 0.08326

>
>
> ###Follow up
> #aprime.log
> mJobHoursNotColl_ASD.GJ = lm(HoursWorking.log ~ aprime.log
+                 #+ age
+                       #+ vabs_soc_ss
+                         , data = dASD.older18.nocoll)
> summary(mJobHoursNotColl_ASD.GJ)

Call:
lm(formula = HoursWorking.log ~ aprime.log, data = dASD.older18.nocoll)

Residuals:
       3        5        7       10       12
 0.03579  0.17271 -0.49484  0.11363  0.17271

Coefficients:
            Estimate Std. Error t value Pr(>|t|)
(Intercept)   3.4127     0.3139  10.872  0.00167 **
aprime.log   -0.5029     1.9281  -0.261  0.81110
---
Signif. codes:  0 ‘***’ 0.001 ‘**’ 0.01 ‘*’ 0.05 ‘.’ 0.1 ‘ ’ 1

Residual standard error: 0.3259 on 3 degrees of freedom
  (7 observations deleted due to missingness)
Multiple R-squared:  0.02217,	Adjusted R-squared:  -0.3038
F-statistic: 0.06803 on 1 and 3 DF,  p-value: 0.8111

>
> mJobHoursNotColl_NT.GJ = lm(HoursWorking.log ~ aprime.log
+                 #+ age
+                       #+ vabs_soc_ss
+                         , data = dNT.older18.nocoll)
> summary(mJobHoursNotColl_NT.GJ)

Call:
lm(formula = HoursWorking.log ~ aprime.log, data = dNT.older18.nocoll)

Residuals:
        4         5         6         7         8         9
 0.074834 -0.087685  0.005958  0.074834 -0.104936  0.036996

Coefficients:
            Estimate Std. Error t value Pr(>|t|)
(Intercept)  3.59451    0.06735  53.369 7.38e-07 ***
aprime.log  -0.34674    0.49044  -0.707    0.519
---
Signif. codes:  0 ‘***’ 0.001 ‘**’ 0.01 ‘*’ 0.05 ‘.’ 0.1 ‘ ’ 1

Residual standard error: 0.08847 on 4 degrees of freedom
  (4 observations deleted due to missingness)
Multiple R-squared:  0.1111,	Adjusted R-squared:  -0.1111
F-statistic: 0.4999 on 1 and 4 DF,  p-value: 0.5186

>
> mJobHoursNotColl_LAD.GJ = lm(HoursWorking.log ~ aprime.log
+                 #+ age
+                       #+ vabs_soc_ss
+                         , data = dLADNT.older18.nocoll)
> summary(mJobHoursNotColl_LAD.GJ)

Call:
lm(formula = HoursWorking.log ~ aprime.log, data = dLADNT.older18.nocoll)

Residuals:
     Min       1Q   Median       3Q      Max
-0.14133  0.01439  0.02119  0.02745  0.03454

Coefficients:
            Estimate Std. Error t value Pr(>|t|)
(Intercept)  3.65434    0.02992 122.138   <2e-16 ***
aprime.log  -0.12586    0.25217  -0.499    0.626
---
Signif. codes:  0 ‘***’ 0.001 ‘**’ 0.01 ‘*’ 0.05 ‘.’ 0.1 ‘ ’ 1

Residual standard error: 0.05878 on 13 degrees of freedom
  (6 observations deleted due to missingness)
Multiple R-squared:  0.0188,	Adjusted R-squared:  -0.05667
F-statistic: 0.2491 on 1 and 13 DF,  p-value: 0.626

>
> #Words
> mJobHoursNotColl_ASD.Words = lm(HoursWorking.log ~ FirstWords.months
+                 #+ age
+                       #+ vabs_soc_ss
+                        , data = dASD.older18.nocoll)
> summary(mJobHoursNotColl_ASD.Words)

Call:
lm(formula = HoursWorking.log ~ FirstWords.months, data = dASD.older18.nocoll)

Residuals:
       3        5        6        7       10
 0.12639 -0.03355  0.00913 -0.05246 -0.04950

Coefficients:
                  Estimate Std. Error t value Pr(>|t|)
(Intercept)       2.872309   0.105914  27.119  0.00011 ***
FirstWords.months 0.029315   0.004981   5.886  0.00979 **
---
Signif. codes:  0 ‘***’ 0.001 ‘**’ 0.01 ‘*’ 0.05 ‘.’ 0.1 ‘ ’ 1

Residual standard error: 0.08638 on 3 degrees of freedom
  (7 observations deleted due to missingness)
Multiple R-squared:  0.9203,	Adjusted R-squared:  0.8937
F-statistic: 34.64 on 1 and 3 DF,  p-value: 0.009788

>
> mJobHoursNotColl_NT.Words = lm(HoursWorking.log ~ FirstWords.months
+                 #+ age
+                       #+ vabs_soc_ss
+                         , data = dNT.older18.nocoll)
> summary(mJobHoursNotColl_NT.Words)

Call:
lm(formula = HoursWorking.log ~ FirstWords.months, data = dNT.older18.nocoll)

Residuals:
       4        5        6        7        8        9
 0.05983 -0.11965  0.04287  0.07396 -0.08573  0.02874

Coefficients:
                   Estimate Std. Error t value Pr(>|t|)
(Intercept)        3.679929   0.103339  35.610 3.71e-06 ***
FirstWords.months -0.002826   0.006023  -0.469    0.663
---
Signif. codes:  0 ‘***’ 0.001 ‘**’ 0.01 ‘*’ 0.05 ‘.’ 0.1 ‘ ’ 1

Residual standard error: 0.09135 on 4 degrees of freedom
  (4 observations deleted due to missingness)
Multiple R-squared:  0.05217,	Adjusted R-squared:  -0.1848
F-statistic: 0.2202 on 1 and 4 DF,  p-value: 0.6633

>
> mJobHoursNotColl_LAD.Words = lm(HoursWorking.log ~ FirstWords.months
+                 #+ age
+                       #+ vabs_soc_ss
+                         , data = dLADNT.older18.nocoll)
> summary(mJobHoursNotColl_LAD.Words)

Call:
lm(formula = HoursWorking.log ~ FirstWords.months, data = dLADNT.older18.nocoll)

Residuals:
     Min       1Q   Median       3Q      Max
-0.14122  0.02124  0.02137  0.02169  0.02360

Coefficients:
                    Estimate Std. Error t value Pr(>|t|)
(Intercept)        3.668e+00  3.240e-02  113.23   <2e-16 ***
FirstWords.months -6.378e-05  1.610e-03   -0.04    0.969
---
Signif. codes:  0 ‘***’ 0.001 ‘**’ 0.01 ‘*’ 0.05 ‘.’ 0.1 ‘ ’ 1

Residual standard error: 0.05934 on 13 degrees of freedom
  (6 observations deleted due to missingness)
Multiple R-squared:  0.0001208,	Adjusted R-squared:  -0.07679
F-statistic: 0.00157 on 1 and 13 DF,  p-value: 0.969

>
> #Phrases
> mJobHoursNotColl_ASD.Phrases = lm(HoursWorking.log ~ FirstPhrases.months
+                 #+ age
+                       #+ vabs_soc_ss
+                         , data = dASD.older18.nocoll)
> summary(mJobHoursNotColl_ASD.Phrases)

Call:
lm(formula = HoursWorking.log ~ FirstPhrases.months, data = dASD.older18.nocoll)

Residuals:
       3        5        6        7       10       12
 0.07845  0.16190  0.07845 -0.50799  0.05985  0.12934

Coefficients:
                    Estimate Std. Error t value Pr(>|t|)
(Intercept)         3.336281   0.443942   7.515  0.00168 **
FirstPhrases.months 0.004651   0.012800   0.363  0.73472
---
Signif. codes:  0 ‘***’ 0.001 ‘**’ 0.01 ‘*’ 0.05 ‘.’ 0.1 ‘ ’ 1

Residual standard error: 0.2815 on 4 degrees of freedom
  (6 observations deleted due to missingness)
Multiple R-squared:  0.03195,	Adjusted R-squared:  -0.2101
F-statistic: 0.132 on 1 and 4 DF,  p-value: 0.7347

>
> mJobHoursNotColl_NT.Phrases = lm(HoursWorking.log ~ FirstPhrases.months
+                 #+ age
+                       #+ vabs_soc_ss
+                         , data = dNT.older18.nocoll)
> summary(mJobHoursNotColl_NT.Phrases)

Call:
lm(formula = HoursWorking.log ~ FirstPhrases.months, data = dNT.older18.nocoll)

Residuals:
        4         5         6         7         8         9
 0.039547 -0.122972  0.023093  0.077940 -0.024248  0.006639

Coefficients:
                     Estimate Std. Error t value Pr(>|t|)
(Intercept)          3.748056   0.088338  42.429 1.84e-06 ***
FirstPhrases.months -0.005485   0.003992  -1.374    0.241
---
Signif. codes:  0 ‘***’ 0.001 ‘**’ 0.01 ‘*’ 0.05 ‘.’ 0.1 ‘ ’ 1

Residual standard error: 0.07734 on 4 degrees of freedom
  (4 observations deleted due to missingness)
Multiple R-squared:  0.3206,	Adjusted R-squared:  0.1508
F-statistic: 1.888 on 1 and 4 DF,  p-value: 0.2414

>
> mJobHoursNotColl_LAD.Phrases = lm(HoursWorking.log ~ FirstPhrases.months
+                 #+ age
+                         , data = dLAD.older18.nocoll)
> summary(mJobHoursNotColl_LAD.Phrases)

Call:
lm(formula = HoursWorking.log ~ FirstPhrases.months, data = dLAD.older18.nocoll)

Residuals:
       Min         1Q     Median         3Q        Max
-2.724e-15 -2.233e-17  2.977e-16  2.977e-16  1.578e-15

Coefficients:
                      Estimate Std. Error    t value Pr(>|t|)
(Intercept)          3.689e+00  1.373e-15  2.686e+15   <2e-16 ***
FirstPhrases.months -5.334e-17  4.746e-17 -1.124e+00    0.304
---
Signif. codes:  0 ‘***’ 0.001 ‘**’ 0.01 ‘*’ 0.05 ‘.’ 0.1 ‘ ’ 1

Residual standard error: 1.308e-15 on 6 degrees of freedom
  (3 observations deleted due to missingness)
Multiple R-squared:  0.5183,	Adjusted R-squared:  0.438
F-statistic: 6.456 on 1 and 6 DF,  p-value: 0.04403

>
```
